# Whole genome sequencing association analysis of quantitative red blood cell phenotypes: the NHLBI TOPMed program

**DOI:** 10.1101/2020.12.09.20246736

**Authors:** Yao Hu, Adrienne M. Stilp, Caitlin P. McHugh, Shuquan Rao, Deepti Jain, Xiuwen Zheng, John Lane, Sébastian Méric de Bellefon, Laura M. Raffield, Ming-Huei Chen, Lisa R. Yanek, Marsha Wheeler, Yao Yao, Chunyan Ren, Jai Broome, Jee-Young Moon, Paul S. de Vries, Brian D. Hobbs, Quan Sun, Praveen Surendran, Jennifer A. Brody, Thomas W. Blackwell, Hélène Choquet, Kathleen Ryan, Ravindranath Duggirala, Nancy Heard-Costa, Zhe Wang, Nathalie Chami, Michael H. Preuss, Nancy Min, Lynette Ekunwe, Leslie A. Lange, Mary Cushman, Nauder Faraday, Joanne E. Curran, Laura Almasy, Kousik Kundu, Albert V. Smith, Stacey Gabriel, Jerome I. Rotter, Myriam Fornage, Donald M. Lloyd-Jones, Ramachandran S. Vasan, Nicholas L. Smith, Kari E. North, Eric Boerwinkle, Lewis C. Becker, Joshua P. Lewis, Goncalo R. Abecasis, Lifang Hou, Jeffrey R. O’Connell, Alanna C. Morrison, Terri H. Beaty, Robert Kaplan, Adolfo Correa, John Blangero, Eric Jorgenson, Bruce M. Psaty, Charles Kooperberg, Russell T. Walton, Benjamin P. Kleinstiver, Hua Tang, Ruth J.F. Loos, Nicole Soranzo, Adam S. Butterworth, Debbie Nickerson, Stephen S. Rich, Braxton D. Mitchell, Andrew D. Johnson, Paul L. Auer, Yun Li, Rasika A. Mathias, Guillaume Lettre, Nathan Pankratz, Cathy C. Laurie, Cecelia A. Laurie, Daniel E. Bauer, Matthew P. Conomos, Alexander P. Reiner, the NHLBI Trans-Omics for Precision Medicine (TOPMed) Consortium

## Abstract

Whole genome sequencing (WGS), a powerful tool for detecting novel coding and non-coding disease-causing variants, has largely been applied to clinical diagnosis of inherited disorders. Here we leveraged WGS data in up to 62,653 ethnically diverse participants from the NHLBI Trans-Omics for Precision Medicine (TOPMed) program and assessed statistical association of variants with seven red blood cell (RBC) quantitative traits. We discovered 14 single variant-RBC trait associations at 12 genomic loci. Several of the RBC trait-variant associations (*RPN1, ELL2, MIDN, HBB, HBA1, PIEZO1, G6PD*) were replicated in independent GWAS datasets imputed to the TOPMed reference panel. Most of these newly discovered variants are rare/low frequency, and several are observed disproportionately among non-European Ancestry (African, Hispanic/Latino, or East Asian) populations. We identified a 3bp indel p.Lys2169del (common only in the Ashkenazi Jewish population) of *PIEZO1*, a gene responsible for the Mendelian red cell disorder hereditary xerocytosis [OMIM 194380], associated with higher MCHC. In stepwise conditional analysis and in gene-based rare variant aggregated association analysis, we identified several of the variants in *HBB, HBA1, TMPRSS6*, and *G6PD* that represent the carrier state for known coding, promoter, or splice site loss-of-function variants that cause inherited RBC disorders. Finally, we applied base and nuclease editing to demonstrate that the sentinel variant rs112097551 (nearest gene *RPN1*) acts through a cis-regulatory element that exerts long-range control of the gene *RUVBL1* which is essential for hematopoiesis. Together, these results demonstrate the utility of WGS in ethnically-diverse population-based samples and gene editing for expanding knowledge of the genetic architecture of quantitative hematologic traits and suggest a continuum between complex trait and Mendelian red cell disorders.

## Introduction

Red blood cells (RBCs) or erythrocytes contain hemoglobin, an iron-rich tetramer composed of two alpha-globin and two beta-globin chains. RBCs play an essential role in oxygen transport and also serve important secondary functions in nitric oxide production, regulation of vascular tone, and immune response to pathogens ^1^. RBC indices, including hemoglobin (HGB), hematocrit (HCT), mean corpuscular hemoglobin (MCH), mean corpuscular hemoglobin concentration (MCHC), mean corpuscular volume (MCV), RBC count, and red blood cell width (RDW), are primary indicators of RBC development, size, and hemoglobin content^2^. These routinely measured clinical laboratory assays may be altered in Mendelian genetic conditions (e.g., hemoglobinopathies such as sickle cell disease or thalassemia, red cell cytoskeletal defects, or G6PD deficiency) ^3^ as well as by non-genetic or nutritional factors (e.g., vitamin B and iron deficiency).

RBC indices have estimated family-based heritability values ranging between 40% to 90% ^4,5^ and have been extensively studied as complex quantitative traits in genome-wide association studies (GWAS). Early GWAS identified common genetic variants with relatively large effects associated with RBC indices ^6,7,8^. With improved imputation, increased sample sizes and deeper interrogation of coding regions of the genome, additional common variants associated with RBC indices with progressively smaller effect sizes and coding variants of larger effect with lower minor allele frequency (MAF) have been identified ^9–19^. However, the full allelic spectrum (e.g., lower frequency non-coding variants, indels, structural variants) that explain the genetic architecture of complex traits remains incomplete ^9^. In addition, non-European populations (including admixed U.S. minority populations such as African Americans and Hispanics/Latinos) have been under-represented in these studies. Since RBCs play a key role in pathogen invasion and defense, associated quantitative trait loci may be relatively isolated to a particular ancestral population due to local evolutionary selective pressures and population history. Emerging studies with greater inclusion of East Asian, African, and Hispanic ancestry populations have identified ancestry-specific variants associated with RBC quantitative traits ^15–17,20,21^. These may account, at least in part, for inter-population differences in RBC indices as well as ethnic disparities in rates of hematologic and other related chronic diseases ^18,22^.

Whole genome sequencing (WGS) data have been generated through the NHLBI Trans-Omics for Precision Medicine (TOPMed) program in very large and ethnically-diverse population samples with existing hematologic laboratory measures. These TOPMed WGS data provide novel opportunities to assess rare and common single nucleotide and indel variants across the genome, including variants more common in African, East Asian or Native American ancestry individuals that are not captured by existing GWAS arrays or imputation reference panels. We thereby aimed to identify novel genetic variants and genes associated with the seven RBC indices, and to dissect association signals at previously reported regions through conditional analysis and fine-mapping.

## Subjects and Methods

### TOPMed study population

The analyses reported here included 62,653 participants from 13 TOPMed studies: Genetics of Cardiometabolic Health in the Amish (Amish, n=1,102), Atherosclerosis Risk in Communities Study VTE cohort (ARIC, n=8,118), Mount Sinai BioMe Biobank (BioMe, n=10,993), Coronary Artery Risk Development in Young Adults (CARDIA, n=3,042), Cardiovascular Health Study (CHS, n=3,490), Genetic Epidemiology of COPD Study (COPDGene, n=5,794), Framingham Heart Study (FHS, n=3,141), Genetic Studies of Atherosclerosis Risk (GeneSTAR, n=1,713), Hispanic Community Health Study - Study of Latinos (HCHS_SOL, n=7,655), Jackson Heart Study (JHS, n=3,033), Multi-Ethnic Study of Atherosclerosis (MESA, n=2,499), Whole Genome Sequencing to Identify Causal Genetic Variants Influencing CVD Risk - San Antonio Family Studies (SAFS, n=1,153) and Women’s Health Initiative (WHI, n=10,920). We analyzed each of seven red blood cell traits separately; the total counts of participants, mean age, and the count of male participants, from each study stratified by trait are shown in **Table 1**. Further descriptions of the design of the participating TOPMed cohorts and the sampling of individuals within each cohort for TOPMed WGS are provided in the section “Participating studies” in the **Supplemental Methods**. All studies were approved by the appropriate institutional review boards (IRBs), and informed consent was obtained from all participants.

**Table 1.** Characteristics of the TOPMed samples by study ^1^.

| Study | N (male) | Age | HCT | HGB | MCH | MCHC | MCV | RBC | RDW |
| --- | --- | --- | --- | --- | --- | --- | --- | --- | --- |
| Amish | 1,102 (557) | 50.6±16.9 | 40.6±3.5 | 13.8±1.2 | 30.9±1.3 | 34.1±0.8 | 90.7±3.4 | 4.5±0.4 | - |
| ARIC | 8,113 (3,577) | 54.8±5.8 | 41.6±4.0 | 13.9±1.4 | 30.5±2.1 | 33.3±1.0 | 89.6±5.1 | 4.5±0.5 | 14.1±1.1 |
| BioMe | 10,990 (4,559) | 52.1±13.5 | 39.5±5.2 | 13.1±1.7 | 30.3±2.8 | 33.7±1.0 | 89.0±7.2 | 4.4±0.6 | 14.2±1.8 |
| CARDIA | 3,042 (1,319) | 25.0±3.6 | 42.1±4.4 | 14.2±1.5 | 29.8±2.1 | 33.8±1.0 | 88.1±5.4 | 4.8±0.5 | - |
| CHS | 3,490 (1,459) | 72.6±5.4 | 41.8±3.9 | 14.0±1.3 | - | 33.5±1.0 | - | - | - |
| COPDGene | 5,794 (2,913) | 64.8±8.8 | 42.0±4.1 | 13.9±1.5 | 30.3±2.3 | 33.2±1.1 | 91.4±5.8 | 4.6±0.5 | - |
| FHS | 3,140 (1,514) | 58.4±15.0 | 41.6±4.0 | 14.1±1.3 | 31.1±1.8 | 33.9±1.0 | 91.9±4.9 | 4.5±0.5 | 13.1±1.0 |
| GeneSTAR | 1,713 (699) | 43.7±12.9 | 40.9±3.9 | 13.5±1.4 | 29.6±2.1 | 33.0±0.8 | 89.5±5.3 | 4.6±0.4 | - |
| HCHS/SOL | 7,655 (3,186) | 46.6±14.0 | 42.1±4.1 | 13.8±1.5 | 29.1±2.2 | 32.7±1.4 | 89.2±6.0 | 4.7±0.4 | 13.8±1.3 |
| JHS | 2,905 (1,089) | 53.5±12.8 | 39.4±4.3 | 13.1±1.5 | 28.9±2.5 | 33.2±0.9 | 86.9±6.3 | 4.5±0.5 | 13.7±1.4 |
| MESA | 2,499 (1,211) | 69.4±9.2 | 40.1±4.0 | 13.4±1.4 | 30.1±2.3 | 33.4±1.1 | 89.9±6.0 | 4.5±0.5 | - |
| SAFS | 1,152 (492) | 40.6±15.9 | 40.3±4.5 | 13.1±1.5 | 29.0±2.3 | 32.6±1.4 | 88.9±5.4 | 4.5±0.5 | - |
| WHI | 10,913 (0) | 66.7±6.8 | 40.2±2.9 | 13.5±1.0 | 29.9±2.1 | 32.9±1.1 | 90.9±5.8 | 4.4±0.4 | 14.2±1.3 |

HCT, hematocrit; HGB, hemoglobin; MCH, mean corpuscular hemoglobin; MCHC, mean corpuscular hemoglobin concentration; MCV, mean corpuscular volume; RBC, red blood cell count; RDW, red blood cell width.
<sup>1</sup> Values are shown as mean±SD.

### RBC trait measurements and exclusion criteria in TOPMed

The seven RBC traits considered for analyses were measured from freshly collected whole blood samples at local clinical laboratories using automated hematology analyzers calibrated to manufacturer recommendations according to clinical laboratory standards. Each trait was defined as follows. HCT is the percentage of volume of blood that is composed of red blood cells. HGB is the mass per volume (grams per deciliter) of hemoglobin in the blood. MCH is the average mass in picograms of hemoglobin per red blood cell. MCHC is the average mass concentration (grams per deciliter) of hemoglobin per red blood cell. MCV is the average volume of red blood cells, measured in femtoliters. RBC count is the count of red blood cells in the blood, by number concentration in millions per microliter. RDW is the measurement of the ratio of variation in width to the mean width of the red blood cell volume distribution curve taken at +/- one CV. In studies where multiple blood cell measurements per participant were available, we selected a single measurement for each trait and each participant as described further in **Supplemental Methods**. Each trait was analyzed to identify extreme values that may have been measurement or recording errors and such observations were removed from the analysis (see **Supplemental Methods**). **Table 1** displays the mean and standard deviation among participants analyzed after exclusions by study.

### WGS data and quality control in TOPMed

WGS was performed as part of the NHLBI TOPMed program. The WGS was performed at an average depth of 38X by six sequencing centers (Broad Genomics, Northwest Genome Institute, Illumina, New York Genome Center, Baylor, and McDonnell Genome Institute) using Illumina X10 technology and DNA from blood. Here we report analyses from ‘Freeze 8,’ for which reads were aligned to human-genome build GRCh38 using a common pipeline across all centers. To perform variant quality control (QC), a support vector machine (SVM) classifier was trained on known variant sites (positive labels) and Mendelian inconsistent variants (negative labels). Further variant filtering was done for variants with excess heterozygosity and Mendelian discordance. Sample QC measures included: concordance between annotated and inferred genetic sex, concordance between prior array genotype data and TOPMed WGS data, and pedigree checks. Details regarding the genotype ‘freezes,’ laboratory methods, data processing, and quality control are described on the TOPMed website and in a common document accompanying each study’s dbGaP accession (https://www.nhlbiwgs.org/topmed-whole-genome-sequencing-methods-freeze-8) ^23^.

### Single variant association analysis

Single variant association tests were performed for each of the seven RBC traits separately using linear mixed models (LMMs). In each case, a model assuming no association between the outcome and any genetic variant was first fit; we refer to this as the ‘null model’. In the null model, covariates modelled as fixed effects were sex; age at trait measurement; a variable indicating TOPMed study and phase of genotyping (study_phase); indicators of whether the participant is known to have had a stroke, chronic obstructive pulmonary disease (COPD), or a venous thromboembolism (VTE) event; and the first 11 PC-AiR ^24^ principal components (PCs) of genetic ancestry. A 4th degree sparse empirical kinship matrix (KM) computed with PC-Relate ^25^ was included to account for genetic relatedness among participants. Additional details on the computation of the ancestry PCs and the sparse KM are provided in the **Supplemental Methods**. Finally, we allowed for heterogeneous residual variances by study and ancestry group (e.g., ARIC_White), as this has been shown previously to control inflation ^26^. The details on how we estimated the ancestry group for this adjustment are in the **Supplemental Methods**. The numbers of individuals per ancestry group per study and the respective mean and standard deviation for each trait are shown in **Supplemental Table 1**.

To improve power and control of false positives when phenotypes have a non-Normal distribution, we implemented a fully-adjusted two-stage procedure for rank-Normalization when fitting the null model, for each of the 7 RBC traits in turn ^27^ :

1. Fit a LMM, with the fixed effect covariates, sparse KM, and heterogeneous residual variance model as described above. Perform a rank-based inverse-Normal transformation of the marginal residuals, and subsequently rescale by their variance prior to transformation. This rescaling allows for clearer interpretation of estimated genotype effect sizes from the subsequent association tests.
2. Fit a second LMM using the rank-Normalized and re-scaled residuals as the outcome, with the same fixed effect covariates, sparse KM, and heterogeneous residual variance model as in Stage 1.

The output of the Stage 2 null model was then used to perform genome-wide score tests of genetic association for all individual variants with minor allele count (MAC) ≥ 5 that passed the TOPMed variant quality filters and had less than 10% of samples freeze-wide with sequencing read depth < 10 at that particular variant. We tested up to 102,674,666 SNVs and 7,722,116 indels (**Supplemental Table 2**). Genome-wide significance was determined at the *P* < 5E-9 level ^28^. For each locus, we defined the top variant as the most significant variant within a 2Mb window. All association analyses were performed using the GENESIS software ^29^.

### Conditional analysis

Because of the very large number of variants and genomic loci that have recently been associated with quantitative RBC traits, following the single variant association analyses, we systematically performed a series of conditional association analyses for each trait to determine which genome-wide significant associations were independent of previously reported RBC variants. We gathered the variants known to be associated with each phenotype from previous publications (**Supplemental Table 3**) and matched these to TOPMed variants using position and alleles. Then, genome-wide conditional association analyses were performed by including known variants as fixed effects covariates in the null model using the same fully-adjusted two stage LMM association testing procedure described above. We performed three types of conditional analysis, namely the trait-specific, the trait-agnostic, and the iterative, step-wise conditional analysis (**Supplemental Methods**).

### Single variant association analysis of chromosome 16

The alpha-globin gene region on chromosome 16p13.3 contains a large, 3.7kb structural variant common among African ancestry individuals known to be highly significantly associated with all RBC traits ^15,18^. This large copy number variant is not well-tagged by SNVs in the region. Therefore, we performed genotype calling for the alpha-globin 3.7kb CNV in 52,772 available TOPMed whole genomes using MosDepth ^30^. Since the chromosome 16 alpha globin CNV calls were only available for a subset of the samples in the primary analyses, to assess the effect of conditioning on the alpha globin CNV, the same set of analyses described above were run for chromosome 16 restricted to the sample set with alpha globin CNV calls. The most probable alpha globin copy number was included as a categorical variable to allow for potential non-linear effects on the phenotype.

### Proportion of variance explained

For each trait, we estimated the proportion of variance explained (PVE) by the set of LD-pruned known associated variants, by the final set of conditionally-independent variants we identified following the iterative stepwise conditional analysis, and by both sets together. These cumulative PVE values were estimated jointly from the null model using approximations from multi-parameter score tests, thus accounting for covariance between the variant genotypes. The estimates were calculated using the full sample set and did not include the alpha globin CNV as a known variant but did include the set of conditionally-independent novel SNVs and indels identified on chromosome 16 after conditioning on the alpha globin CNV.

### Replication studies for single variant association findings

We sought replication of the lead variants at genome-wide significant loci identified in the trait-specific conditional analysis in independent studies including the INTERVAL study (https://www.intervalstudy.org.uk/), the Kaiser-Permanente Genetic Epidemiology Research on Aging (GERA) cohort (https://www.ncbi.nlm.nih.gov/projects/gap/cgi-bin/study.cgi?study_id=phs000674.v3.p3/), samples from the Women’s Health Initiative - SNP Health Association Resource (WHI-SHARe) ^31^ not included in TOPMed, European ancestry samples from phase 1 of the UK BioBank (UKBB) ^9^ and African and East Asian ancestry samples from phase 2 of UKBB ^21^. WGS data was used in INTERVAL while genotyping on various arrays and imputation to TOPMed WGS data or 1000 Genome Phase 3 reference panels were performed in Kaiser, WHI-SHARe, and UKBB. Residuals were obtained by regressing the harmonized RBC traits on age, sex, the first 10 PCs in each study stratified by ancestry, followed by association analyses testing each genetic variant with the inverse-normalized residual values. Summary statistics from each study were combined through fixed-effect inverse-weighting meta-analysis using METAL ^32^.

### Aggregate variant association analysis of rare variants within each gene

Association tests aggregating rare variants by gene were performed for each RBC trait in order to assess the cumulative effect of rare variants within each gene and associated regulatory regions. We applied five strategies for grouping and filtering variants. Three of them aggregated coding variants and two of them aggregated coding and non-coding regulatory variants. For each aggregation strategy we filtered variants using one or more deleterious prediction scores creating relatively relaxed or stringent sets of variants (see details in **Supplemental Methods**). The five strategies are referred as C1-S,C1-R,C2-R,C2-R+NC-S and C2-R +NC-R by abbreviating, coding to “C”, Non-coding to “NC”, Stringent to “S” and Relaxed to “R”. For all aggregate units, only variants with MAF < 0.01 that passed the quality filters and had less than 10% of samples with sequencing read depth < 10 were considered. The aggregate association tests were performed using the Efficient Variant-Set Mixed Model Association Test (SMMAT) ^33^. The SMMAT test used the same fully-adjusted two-stage null model as was fit for the single variant association tests. For each aggregation unit, SMMAT efficiently combines a burden test *P* value with an asymptotically independent adjusted “SKAT-type” test *P* value using Fisher’s method. This testing approach is more powerful than either a burden or SKAT ^34^ test alone, and is computationally more efficient than the SKAT-O test ^35^. Wu weights ^34^ based on the variant MAF were used to upweight rarer variants in the aggregation units. Significance was determined using a Bonferroni threshold, adjusting for the number of gene-based aggregation units tested genome-wide with cumulative MAC >= 5. Two types of conditional analysis were run (“trait-specific” and “trait-agnostic), conditioning previously reported RBC trait-associated variants as well as those discovered in the TOPMed single variant tests (**Supplemental Table 3**). In addition, any previously reported RBC trait-associated variants and the set of conditionally-independent novel variants identified in our single variant analyses were excluded from the gene-based aggregation units.

### Predicted loss-of-function variants and predicted gene knockouts and their association with RBC traits

Our analyses of predicted loss-of-function (pLoF) variants in TOPMed freeze 8 focused on variants annotated by ENSEMBL’s Variant Effect Predictor (VEP) as nonsense, essential splice site and frameshift insertion-deletion (indel) variants. From this list, we excluded variants that map to predicted transcripts ^36^ and also variants located in the first and last 5% of the gene as these variants are more likely to give rise to transcripts that escape nonsense-mediated mRNA decay ^37^. We used a method previously described to identify predicted gene knockouts (pKO) ^38^. Briefly, we considered individuals that were homozygotes for LoF variants, but also individuals who inherited two different LoF variants in *trans* using available phased information (compound heterozygotes).

We analyzed each study-ethnic group separately, adjusting for sex, age and smoking status. We then normalized the residuals with each group using inverse normal transformation. We performed association testing per ethnic group with EPACTS. We adjusted all analyses using the first ten PCs and a kinship matrix (EMMAX) calculated using 150,000 common variants in LD. For pLoF, we tested an additive genetic model. For pKO, we coded individuals as “0” if they were not a pKO, and as “1” if they were a pKO. We meta-analyzed association results using METAL ^32^. We excluded variants located in the alpha-globin region in self-reported African-ancestry individuals. The genome-wide significant threshold for each ancestral group was defined as *P*<0.05/number of variants.

### Lentivirus packaging

HEK293T cells (ATCC, cat# CRL-3216) were cultured with DMEM with 10% fetal bovine serum and 1% Penicillin-Streptomycin solution (10,000 U/mL stock). To produce lentivirus, HEK293T cells were transfected at 70-80% confluence with 13.3 μg psPAX2, 6.7 μg VSV-G and 20 μg of the lentiviral construct plasmid of interest using 180 μg of linear polyethylenimine in 15 cm tissue culture dishes. Lentiviral supernatant was collected at both 48 h and 72 h post-transfection and concentrated by ultracentrifugation at 24,000 rpm for 4 h at 4 °C with a Beckman Coulter SW 32 Ti rotor.

### HUDEP-2 cell and human CD34^+^ hematopoietic stem and progenitor cells (HSPCs) culture

HUDEP-2 cells ^39^ were generously shared by Ryo Kurita (Japanese Red Cross) and Yukio Nakamura (RIKEN BioResource Research Center, University of Tsukuba, Japan) and cultured as previously described ^40^. Expansion phase medium for HUDEP-2 cells consists of SFEM (Stemcell Technologies #09650) base medium supplemented with 50 ng/ml recombinant human SCF (R&D systems #255-SC), 1 µg/ml doxycycline (Sigma Aldrich #D9891), 0.4 µg/ml dexamethasone (Sigma Aldrich #D4902), 3 IU/ml EPO (Epoetin Alfa, Epogen, Amgen) and 1% Penicillin-Streptomycin solution (10,000 U/mL stock). Human CD34^+^ HSPCs from mobilized peripheral blood of deidentified healthy donors were obtained from Fred Hutchinson Cancer Research Center, Seattle, Washington. CD34^+^ cells were maintained in SFEM supplemented with 1x StemSpan CD34^+^ expansion supplement (Cat# 02691, STEMCELL Technology).

### Generation of AncBE4max-SpRY-expressing stable HUDEP-2 cell lines ^41^

The lentiviral plasmid for AncBE4max-SpRY was generated by subcloning the coding sequence of nSpRY(D10A) into the AgeI and XcmI restriction sites of pRDA_257 (pLenti-BPNLS-AncBE4-gsXTENgs-nSpCas9-gs-UGI-gs-BPNLS-P2A-Puro), generously provided by John Doench (Broad Institute). Lentivirus was produced as described above. HUDEP-2 cells were transduced with lentivirus, and 1 μg/ml puromycin was added into culture medium 2 days after lentiviral transduction. After 2-week positive selection, AncBE4max-SpRY editing efficiency was tested using multiple sgRNAs with variable PAM sequence.

### C-to-T base editing at the rs112097551 locus in HUDEP-2 cells

The sequence of single guide RNA targeting rs112097551 is summarized in **Supplemental Table 4**. Oligos (from GENEWIZ company) were annealed and ligated into LentiGuide-Puro (Addgene plasmid 52963). Following lentiviral production and transduction into cell lines with stable SpCas9 expression, 1 μg/ml puromycin were added to select for sgRNA integrants in HUDEP-2 cells expressing AncBE4max-SpRY. C-to-T editing efficiency was determined in bulk cells 10 days after lentiviral delivery into AncBE4max-SpRY-expressing HUDEP-2 cells (**Supplemental Fig. 1**). Briefly, genomic DNA was extracted using the Qiagen Blood and Tissue kit. Genomic region surrounding the sgRNA targeting site was amplified using HotStarTaq DNA polymerase (QIAGEN, Cat# 203203) for other PCR reactions strictly following the manufactory instructions with variable annealing temperature. PCR products were subject to Sanger sequencing and then EditR analysis to estimate the editing efficiency based on sequencing chromatograms ^42^. Single HUDEP-2 cells were plated to obtain highly edited clones. Primers for PCR were summarized in **Supplemental Table 5**.

**Figure 1.**
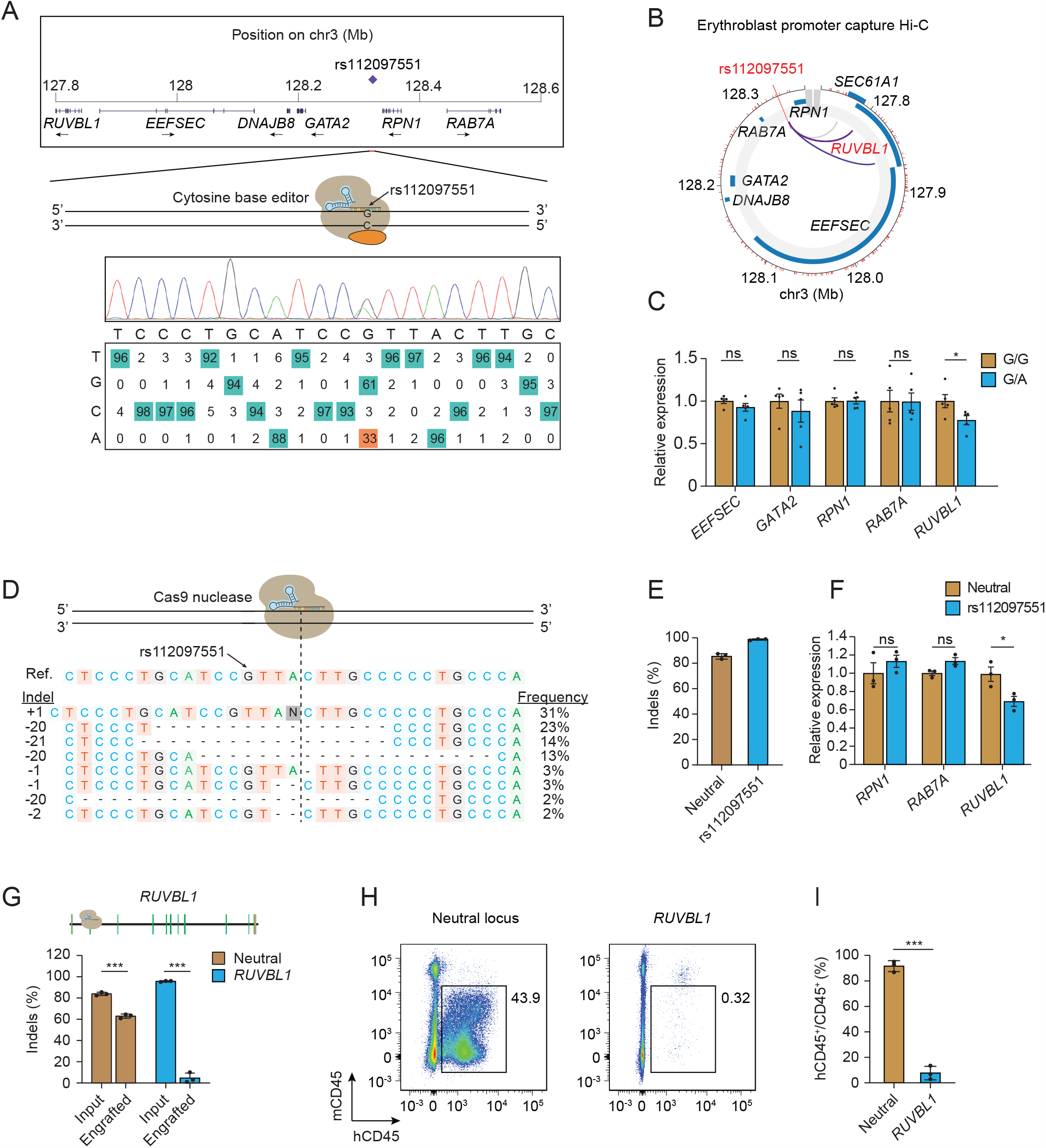
Gene editing implicates *RUVBL1* in rs112097551 association. (A) The MCV/MCH associated variant rs112097551 was targeted by cytosine base editing in HUDEP-2 cells expressing AncBE4max-SpRY and sgRNA to convert G-to-A. Sequencing chromatogram and heatmap of bulk edited HUDEP-2 cells generated by EditR analysis. (B) Promoter capture Hi-C from ChiCP analysis ^105^ of erythroblasts ^106^. (C) Gene expression measured by RT-qPCR in rs112097551-G/G (n=5) and -G/A (n=5) HUDEP-2 base edited clones. Expression normalized to mean of G/G clones for each gene. (D) Representative allele table demonstrating type and frequency of indels following nuclease editing in CD34+ HSPCs following 3xNLS-SpCas9:sgRNA electroporation. Indels analyzed by TIDE analysis ^45^. (E) Indel frequency measured by Sanger sequencing with TIDE analysis in CD34+ HSPCs 4 days following 3xNLS-SpCas9:sgRNA electroporation with indicated sgRNA (n = 3 biological replicates). (F) Gene expression measured by RT-qPCR in CD34+ HSPCs 4 days following 3xNLS-SpCas9:sgRNA targeting adjacent to rs112097551 compared to neutral locus. Expression of *EEFSEC* and *GATA2* was undetectable in HSPCs. (G) Indel frequency following 3xNLS-SpCas9:sgRNA targeting *RUVBL1* coding sequence or neutral control locus in input cell 4 days after RNP electroporation or engrafted bone marrow samples 16 weeks after infusion to NBSGW mice. (H) Representative flow cytometry of human and mouse CD45+ cells from NBSGW bone marrow 16 weeks after cell infusion (representative of 3 mice). (I) Mean human hematopoietic chimerism determined by hCD45+/total CD45+ cells from NBSGW bone marrow 16 weeks after cell infusion (n = 3 mice per group). Student’s t test (two-tailed test). *** P < 0.001; ** P < 0.01; * P < 0.05; ns, not significant.

### CRISPR/Cas genome editing in CD34^+^ HSPCs

CD34+ cells were thawed and maintained in SFEM supplemented with 1x StemSpan CD34^+^ expansion supplement (Cat# 02691, STEMCELL Technology) for 24 hours before electroporation. 100,000 cells per condition were electroporated using the Lonza 4D nucleofector with 100 pmol 3xNLS-SpCas9 ^43^ protein and 300 pmol modified sgRNA targeting the locus of interest. In addition to mock treated cells, “safe-targeting” RNPs were used as experimental controls as indicated in each figure legend. After electroporation, cells were differentiated to erythroblasts as described previously ^44^. 4 days after electroporation, genomic DNA was isolated from an aliquot of cells, the sgRNA targeted locus was amplified by PCR. PCR products were subject to Sanger sequencing and then TIDE analysis to quantify indel mutations ^45^. Meanwhile, total RNA was extracted from bulk cells and expression of genes of interest was determined by real time RT-qPCR as described below.

### Determination of target gene expression

Total RNA was extracted from cell cultures 4 days after electroporation using the RNeasy Plus Mini Kit (QIAGEN), and reverse transcribed using the iScript cDNA synthesis kit (Biorad) according to the manufacturer’s instructions. Expression of target genes was quantified using real-time RT-qPCR with GAPDH as an internal control. All gene expression data represent the mean of at least three biological replicates. Primers for PCR were summarized in **Supplemental Table 5**.

### Immunophenotyping of human CD34^+^ HSPCs xenograft from NBSGW mice

NOD.Cg-KitW-41J Tyr + Prkdcscid Il2rgtm1Wjl (NBSGW) mice were obtained from Jackson Laboratory (Stock 026622). CD34^+^ HSPCs were maintained and edited as described above. After electroporation, cells were allowed to recover for 24-48 hours in SFEM medium with 1x StemSpan CD34^+^ expansion supplement (Cat# 02691, STEMCELL Technology). Cells were then washed twice by PBS, resuspended in 200 ul DPBS per million cells, and then infused by retro-orbital injection into non-irradiated NBSGW female mice. 16 weeks post transplantation, mice were euthanized, and bone marrow was collected and analyzed as previously described ^45^. Analysis of bone marrow subpopulations was performed by flow cytometry. Antibodies for flow cytometry included Human TruStainFcX (422302, BioLegend), TruStainfcX (anti-mouse CD16/32, 101320, BioLegend), anti-mouse CD45 (30-F11), anti-human CD45 (HI30), and Fixable Viability Dye eFluor 780 for live/dead staining (65-0865-14, Thermo Fisher). Percentage human engraftment was calculated as hCD45^+^ cells/(hCD45^+^ + mCD45^+^ cells). Cell sorting was performed on a FACSAria II machine (BD Biosciences).

## Results

### Single variant association analysis

In the single variant association analyses, the genomic inflation factors ranged from 1.015 to 1.038, indicating adequate control of population stratification and relatedness (**Supplemental Table 6**). A total of 69 loci reached genome-wide significance for any of the seven RBC traits (*P*<5E-9, **Supplemental Fig. 2** and **Supplemental Table 7**). Of the 69 loci, nine (*HBB, HBA1, RPN1, ELL2, EIF5-MARK3, MIDN, PIEZO1, TMPRSS6*, and *G6PD*) remained significant in the conditional analysis after accounting for RBC trait-specific known loci. In addition, three more loci reached genome-wide significance following RBC trait-specific conditional analysis (*19q12, 10q26*, and *SHANK2, P*<5E-9, **Supplemental Fig. 3**). Therefore, a total of 12 loci showed genome-wide significance for association with at least one of the seven RBC traits in the trait-specific conditional analysis, indicating signals independent of previously reported variants (*P*<5E-9) (**Supplemental Fig. 4, Table 2**).

**Table 2.**
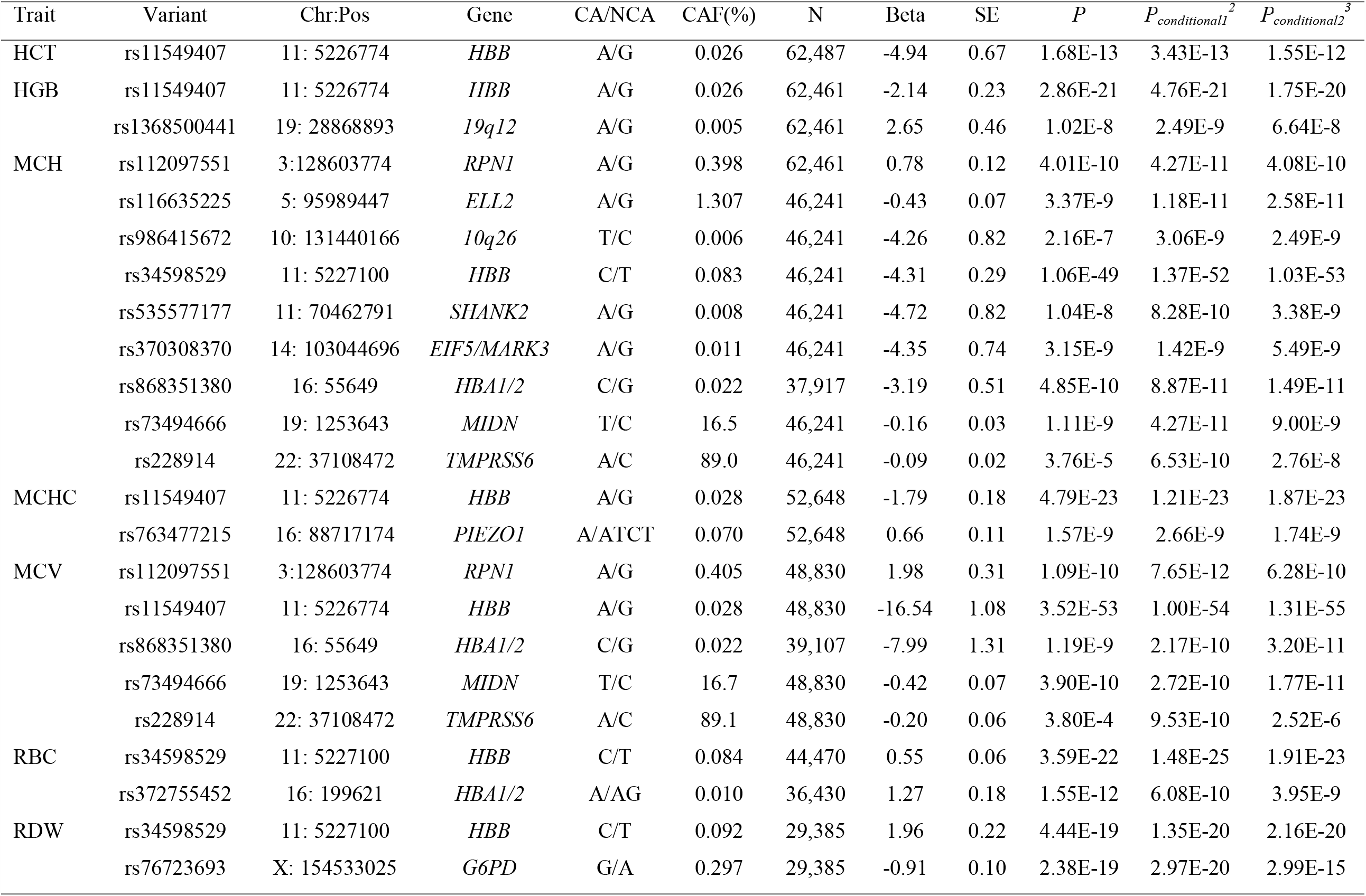
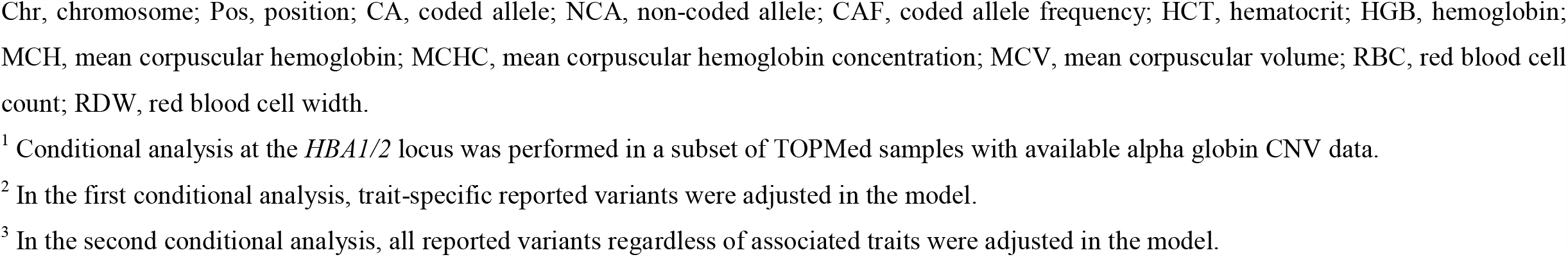
Genome-wide significant loci identified in the trait-specific conditional analysis in TOPMed ^1^.

At the 12 significant novel loci identified in the trait-specific conditional analyses, the number of genome-wide significant variants ranged from one to 162 (**Supplemental Fig. 4** and **Supplemental Table 8**). Six loci harbored more than one genome-wide significant variants (*HBB, HBA1, ELL2, MIDN, TMPRSS6*, and *G6PD*). The lead variants for each trait at each of these 12 loci (including 14 distinct variants across the 7 traits -- 12 SNVs and two small indels) are shown in **Table 2**. Notably, only two lead variants (*MIDN*-rs73494666 and *TMPRSS6-*rs228914) had MAF>5% in TOPMed. Most of these 14 lead variants were located within non-coding regions of the genome and most were low frequency (n=3 between MAF 0.1% and MAF 2%) or rare (n=9 with MAF<0.1%). The latter category included three loci (*SHANK2, 10q26*, and *19q12*) in which the lead variant was extremely rare with MAF<0.01%. Several of the lead variants showed large allele frequency differences between race/ethnicity groups as assessed from the genome aggregation database or gnomAD (https://gnomad.broadinstitute.org; **Supplemental Table 9**). The *RPN1, HBB* -rs34598529, *G6PD, MIDN*, and *ELL2* variants are found disproportionately among individuals of African ancestry. The *EIF5-MARK3* and chromosome 16p13.3 alpha-globin locus rs372755452 variants are found only among East Asians. The alpha-globin locus variant rs868351380 and *PIEZO1* variant are more common among Hispanics/Latinos and Europeans, respectively.

### Replication of single variant discoveries

We sought replication for each of the 14 newly discovered variants in INTERVAL, the Kaiser Permanente GERA Study, the WHI-SHARe study, and UKBB phase 1 European and phase 2 African, and East Asian samples (**Supplemental Table 10**). Several of the rare variants (*SHANK2* rs535577177, *10q26* rs986415672, *19q12* rs1368500441, *EIF5/MARK3* rs370308370, and *HBB* rs11549407), were not available for testing in any of the replication studies due to low frequency, population specificity, and/or poor imputation quality. For eight of the nine lead variants with available genotype data for testing, we successfully replicated each of the trait-specific associations for *HBB-*rs34598529, *HBA1*-rs868351380, *HBA1*-rs372755452, *RPN1, ELL2, PIEZO1, G6PD*, and *MIDN* (meta-analysis *P*<5.6E-3, 0.05/9 loci, with consistent directions of effect). The replication *P* value for the lead variant at *TMPRSS6* did not reach the predetermined significance threshold, but the association was directionally consistent. We further note that several of our newly identified TOPMed single variant-RBC trait associations (*RPN1, HBB-* rs11549407 and rs34598529, and *MIDN*) reached genome-wide significance in recently published very large European ancestry or multi-ethnic imputed GWAS ^19,21,46^.

### Relationship of single variants discovered in TOPMed to previously known RBC genetic loci

Several of the newly discovered variants (particularly those replicated in independent samples) in **Table 2** are located within genomic regions known to harbor common variants associated with RBC quantitative traits and/or variants responsible for Mendelian blood cell disorders, such as hemoglobinopathies (*HBB, HBA1/HBA2*) and various hemolytic or non-hemolytic anemias (*G6PD, PIEZO1, TMPRSS6*, and *GATA2-RPN1*). At the *HBB* locus, the lead variant associated with lower HCT, HGB, MCHC, and MCV is a LoF variant (rs11549407 encoding p.Gln40Ter, MAF=0.026%) while the lead variant associated with lower MCH and higher RBC, and higher RDW is a variant located within the *HBB* promoter region (rs34598529, MAF=0.083%). At the *HBA1/HBA2* locus, the lead variant for MCH and MCV, rs868351380 (MAF=0.022%), is located ∼125kb upstream of *HBA1/HBA2* in an intron of the *SNRNP25* gene, and the lead variant for RBC, rs372755452 (MAF=0.010%), is located ∼30kb downstream of *HBA1/HBA2* in an intron of the *LUC7L* gene. The *GATA2-RPN1* locus, which contains variants previously reported for association with MCH and RDW in a European-only analysis (rs2977562 and rs147412900) ^13^, was associated with MCH and MCV in TOPMed (lead variant rs112097551, *P*=4.27E-11). The MAF of the lead variant at the *GATA2-RPN1* locus in all TOPMed samples is 0.4% but is 5.9 times more common among African than non-African samples according to gnomAD. At the *G6PD* locus, the lead variant associated with lower RDW was a missense variant rs76723693, which encodes p.Leu323Pro. At the *PIEZO1* locus, the most significant variant was an in-frame 3bp deletion rs763477215 (p.Lys2169del) associated with higher MCHC. While the index SNP rs228914 at *TMPRSS6* has not been previously associated with RBC parameters, rs228914 is a cis-eQTL for *TMPRSS6* and an LD surrogate rs228916 has been previously associated with serum iron levels ^47^. The remaining genetic loci (*SHANK2, ELL2, 19q12, 10q26, EIF5/MARK3*, and *MIDN*) have less clear functional relationships to RBC phenotypes. Moreover, the lead variants at *EIF5/MARK3*, and *MIDN* for MCH and the lead variant at *TMPRSS6* for MCH and MCV were partially attenuated in the trait-agnostic conditional analysis.

### Iterative conditional analysis identifies extensive allelic heterogeneity at HBB locus

We next performed stepwise conditional analysis to dissect association signals within each of the six loci harboring more than one genome-wide significant variants in the RBC trait-specific conditional analysis. One of the six regions (*HBB*) was found to have multiple, genome-wide significant variants independent of previously reported loci. The largest number of independent signals were observed for association with MCH (11 signals, **Supplemental Table 11**). All independent variants at the *HBB* locus had MAF <1%. No secondary independent signals were discovered in other regions (*HBA1/2, ELL2, MIDN, TMPRSS6*, and *G6PD*). For each RBC trait, we estimated the PVE by the set of LD-pruned known variants, by the novel conditionally-independent variants identified in stepwise conditional analysis, and by both sets together (**Supplemental Table 12**). In total, the PVE ranged from 3.4% (HCT) to 21.3% (MCH). The newly identified set of genetic variants explained up to 3% of phenotypic variance (for MCH and MCV).

### Rare variant aggregated association analysis

We next examined rare variants with MAF <1% in TOPMed, aggregated based on protein-coding and non-coding gene units from GENCODE. To enrich for likely causal variants in the aggregation units we used five different variant grouping and filtering strategies based on coding sequence and regulatory (gene promoter/enhancer) functional annotations (see **Supplemental Methods**). After accounting for all previously reported RBC trait-specific single variants, a total of five loci were significantly associated with one or more RBC traits using various aggregation strategies (**Table 3** and **Supplemental Table 13**). These include genes encoding *HBA1/HBA2, TMPRSS6, G6PD*, and *CD36*, as well as several genes and non-coding RNAs within the beta-globin locus on chromosome 11p15 (*HBB, HBG1, CTD-264317*.*6, OR52H1, RF60021*, and *OR52R1*). Some of the gene units in the chromosome 11p15 beta-globin region (*HBG1, OR52R1*, and *RF00621*) became non-significant after further adjustment for all known RBC variants in the trait-agnostic conditional analysis (**Table 3**). After additionally accounting for all 11 independent single variant signals identified in TOPMed at the *HBB* locus in step-wise conditional analysis (**Supplemental Table 11**), as well as all trait-specific known variants, five coding genes remained significant (*HBA1/HBA2, HBB, TMPRSS6, G6PD*, and *CD36*, **Supplemental Table 14**) and two additional genes (*TFRC* and *SLC12A7*) reached significance threshold (**Supplemental Table 14**). *AC104389*.*6* a non-coding gene 2bp downstream of *HBB* was also found significant in the aggregation approach where we included upstream regulatory variants, however the variants including in this unit are predominately the same ones tested in the *HBB* gene unit and hence we have not reported this gene unit as a distinct signal.

**Table 3.**
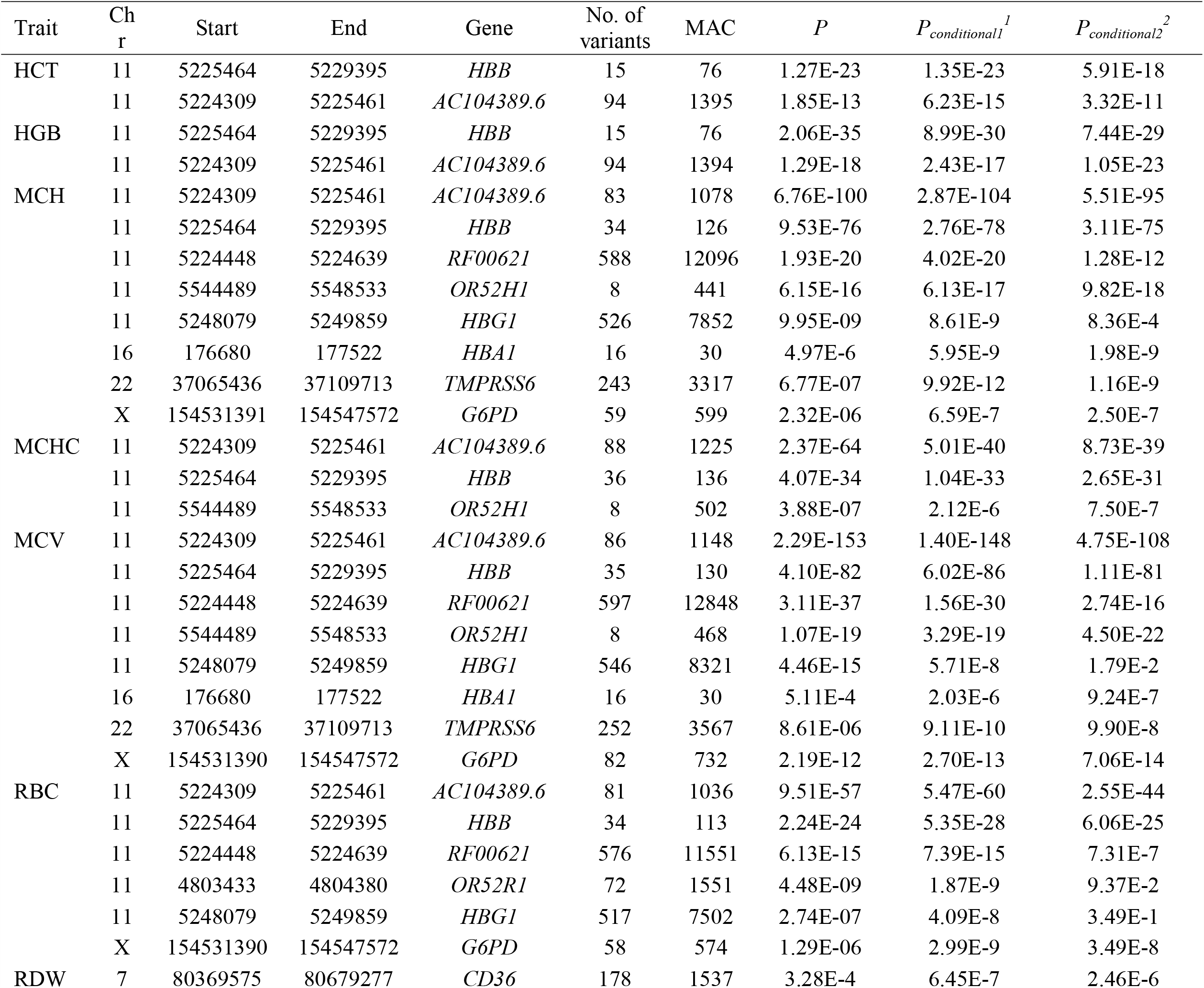
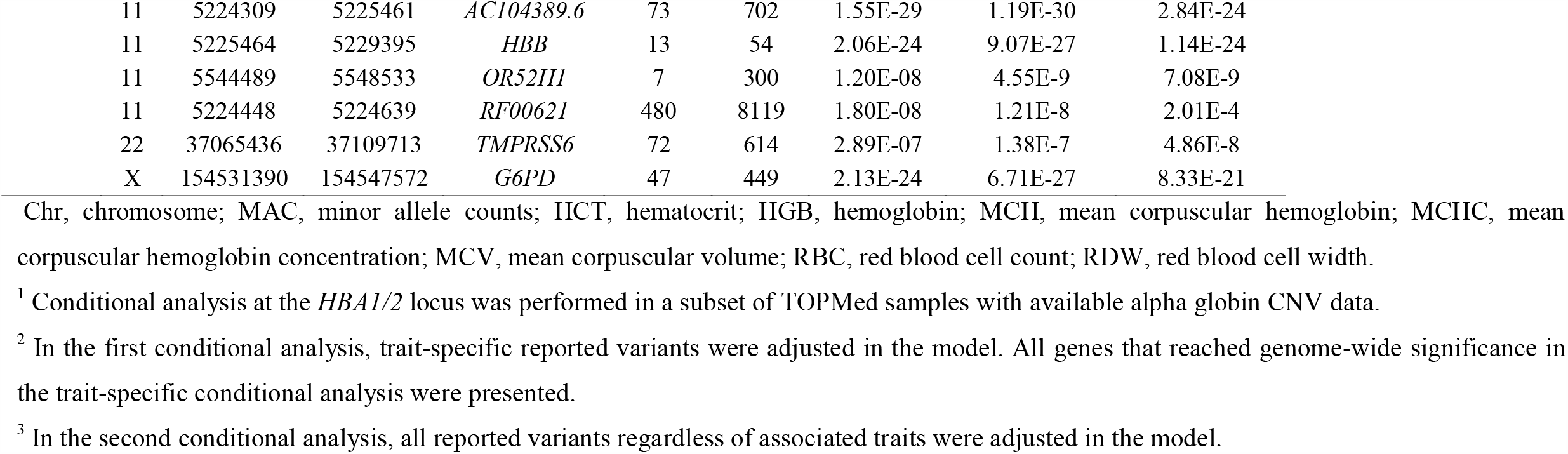
Genome-wide significant genes in the aggregated association analysis in TOPMed ^1^.

Notably, each of the seven genes (*HBA1/HBA2, HBB, TMPRSS6, G6PD, CD36, TFRC* and *SLC12A7*) identified in rare variant aggregate analyses are known to harbor common non-coding or coding variants previously associated with RBC traits or disorders. We further explored the overall patterns of association, individual rare variants driving the associations and their annotations (**Supplemental Fig. 5** and **Supplemental Table 15**). Several observations are noteworthy. (1) In general, for each gene, there are multiple rare missense and small indel (frameshift or stop-gain) variants contributing to the aggregate association signals, rather than a single strongly associated variant. (2) The patterns of phenotypic association are generally uni-directional, and consistent with the biologic contribution of these genes to inherited RBC disorders: *HBA1* and *HBB* variants are associated with lower MCV/MCH, with *HBB* variants additionally associated with lower HCT/HGB and higher RBC/RDW, consistent with ineffective erythropoiesis and shortened red cell survival in alpha and beta thalassemia; *TMPRSS6* variants associated with lower MCH/MCV (**Supplemental Fig. 5C16-19 and 5E13-14**) and higher RDW (**Supplemental Fig. 5G14**), consistent with iron-refractory iron deficiency anemia. On the other hand, for *G6PD* rare variants, a bi-directional pattern of phenotypic association was observed for MCH, MCV, RBC, and RDW. (3) Several of the variants contributing to the *HBA1, HBB, TMPRSS6*, and *G6PD* signals are known to be pathogenic for inherited RBC disorders. Other variants that appear to contribute to the gene-based phenotypic effect are classified in ClinVar as variants of uncertain significance (VUS) or have conflicting evidence to support their pathogenicity. (4) Three of the genes (*CD36, TFRC* and *SLC12A7*) are located within regions of the genome containing common variants previously associated with RBC traits but have less clear relation to RBC biology. The presence of rare coding or LoF variants within these genes provides additional fine-mapping evidence that these three genes are causally responsible for RBC phenotypic variation.

### pLoF and pKO variants associated with RBC traits

Predicted loss-of-function (pLoF) and predicted gene knockout (pKO) variants were examined in European, African, Hispanic, and Asian ancestry populations in TOPMed. The European ancestry population subset had the largest sample size and the largest number of both pLoF and pKO variants (**Supplemental Table 16**). Two pLoF variants reached genome-wide significance, namely *CD36*-rs3211938 for RDW in African participants and *HBB*-rs11549407 for multiple RBC traits in Hispanic and European participants (**Supplemental Table 17**), which have been reported in previously published studies. No pKO variant reached genome-wide significance in any of the ancestral groups (**Supplemental Table 18**). All pLoF and pKO variants with *P*<1E-4 are presented in **Supplemental Table 17 and 18**.

### Gene editing in human erythroid precursors and xenotransplantation of edited primary HSPCs identifies RUVBL1 as likely target gene of RPN1-rs112097551

In silico functional annotation of the *RPN1*-rs112097551 variant revealed a CADD-PHRED score of 20.4 and that the variant lies in a putative enhancer element bound by erythroid transcription factors GATA1 and TAL1. We therefore undertook additional experiments to investigate the causal gene underlying the association signal. First, we used cytosine base editing to modify the rs112097551 reference G to alternative A allele in HUDEP-2 erythroid precursor cells. Since there was no appropriately positioned NGG PAM motif, we utilized the recently described near-PAMless SpCas9 variant cytosine base editor AncBE4max-SpRY ^41^, achieving 33% G-to-A conversion efficiency (**Fig. 1A**). Analysis of erythroblast promoter capture Hi-C datasets showed that the SNP interacts with the gene *RUVBL1* which is 500 kb upstream but not with intervening genes which include *RPN1* and the hematopoietic transcription factor *GATA2* (**Fig. 1B**). In five G/A heterozygous HUDEP-2 clones compared to G/G clones, we observed significantly reduced expression of *RUVBL1* without significant change in expression of 4 more proximal genes *EEFSEC, GATA2, RPN1* and *RAB7A* (**Fig. 1C**). Next, we performed SpCas9 nuclease editing to produce indels adjacent to rs112097551 in CD34+ hematopoietic stem/progenitor cell (HSPC) derived primary erythroid precursors (**Fig. 1D and 1E**). Cells bearing these short insertions and deletions centered 3 bp from the rs112097551 position demonstrated significantly reduced *RUVBL1* expression compared to control cells, while *RPN1* and *RAB7A* expression was unchanged (**Fig. 1F**). Together, these base and nuclease editing results suggest that rs112097551-G contributes to a regulatory element that exerts long-range control of *RUVBL1* expression. Prior work has shown the mouse homolog of *RUVBL1* is required for murine hematopoiesis ^48^. To test the role of *RUVBL1* in human hematopoiesis, we performed gene editing studies in CD34+ HSPCs in which we targeted indels to coding sequences at *RUVBL1*. We observed 96.1% indels at *RUVBL1* compared to 84.2% indels in control cells targeted at a neutral locus. We infused edited HSPCs to immunodeficient NBSGW mice and analyzed bone marrow after 16 weeks for engrafting human hematopoietic chimerism and gene editing. Compared to CD34+ HSPCs edited at a neutral locus which showed 91.6% mean human chimerism, human CD34+ HSPCs edited at *RUVBL1* demonstrated only 7.7% mean chimerism (**Fig. 1G-I**). Engrafting human cells were marked by frequent gene edits (60.1%) when targeted at the neutral locus but only 4.8% gene edits after *RUVBL1* editing, indicating that *RUVBL1* edited cells inefficiently engrafted. Together these results suggest rs112097551-G contributes to long-range enhancement of *RUVBL1* expression, which in turn supports human hematopoiesis.

## Discussion

We report here the first WGS-based association analysis of RBC traits in an ethnically diverse sample of 62,653 participants from TOPMed. We identified 14 association signals across 12 genomic regions conditionally independent of previously reported RBC trait loci and replicated eight of these (*RPN1, ELL2, PIEZO1, G6PD, MIDN, HBB*-rs34598529, *HBA1-*rs868351380 and -rs372755452) in independent samples with available imputed genome-wide genotype data. The replicated association signals are described further below. Stepwise, iterative conditional analysis of the beta-globin gene regions on chromosomes 11 additionally identified 12 independent association signals at the *HBB* locus. Further investigation of aggregated rare variants identified seven genes (*HBA1/HBA2, HBB, TMPRSS6, G6PD, CD36, TFRC* and *SLC12A7*) containing significant rare variant association signals independent of previously reported and newly discovered RBC trait-associated single variants. For the *RPN1* locus, we used base and nuclease editing to demonstrate that the sentinel variant rs112097551 acts through a cis-regulatory element that exerts long-range control of the gene *RUVBL1* which is essential for hematopoiesis.

Our study highlights the benefits of increasing participant ethnic diversity and coverage of the genome in genetic association studies of complex polygenic traits. Among the 24 unique novel and independent variants we identified in the single variant association analyses, 21 showed MAF <1% in all TOPMed samples and 18 were monomorphic in at least one of the four major contributing ancestral populations in our analysis (European, African, East Asian, and Hispanic). These low-frequency or ancestry-specific variants were most likely missed by previous GWAS analysis using imputed genotype data or focusing on one ancestral population (**Supplemental Table 11**).

### GATA2-RPN1

Here we report and replicate a distinct low-frequency variant [MAF=0.4% overall but considerably higher frequency among African (0.94%) than European (0.07%) ancestry individuals) associated with higher MCH and MCV in TOPMed (rs112097551). The region between *GATA2* and *RPN1* on chromosome 3q21 contains several common variants previously associated with various WBC-related traits in European, Asian, and Hispanic ancestry individuals and two variants previously associated with MCH and RDW in Europeans (rs2977562 and rs147412900) ^13^. GATA2 is a hematopoietic transcription factor and heterozygous coding or enhancer mutations of *GATA2* are responsible for autosomal dominant hereditary mononuclear cytopenia, immunodeficiency and myelodysplastic syndromes, as well as lymphatic dysfunction ^49,50^ (MIM 137295). There was no evidence of association of the TOPMed MCH/MCV-associated rs112097551 variant with WBC-related traits in TOPMed (data not shown), though the variant was associated with higher monocyte count and percentage in Astle et al ^9^, but was not conditionally independent of other variants in the region. The MCV/MCH-associated rs112097551 variant lies in a putative enhancer element bound by erythroid transcription factors GATA-1 and TAL-1 and demonstrates physical interaction in erythroblasts with gene *RUVBL1* 500kb away. Our results from gene editing of *RUVBL1* in primary human HPSC and xenotransplantation suggest that *RUVBL1* plays a role in human hematopoiesis, consistent with data from mouse models suggesting that *RUVBL1* (which encodes the protein product pontin) to be essential for murine hematopoietic stem cell survival ^48^. This finding also highlights the complexity and importance of experimentally validating the causal gene(s) underlying GWAS signals for complex traits, which are often assigned according to physical proximity (*RPN1*) or assumed on the basis of biologic function (*GATA2*).

### ELL2

The chromosome *5q15* non-coding variant rs116635225 associated with lower MCH also has a low frequency in TOPMed (1.3%) and is considerably more common among African ancestry individuals (3.9%). The rs116635225 variant is located ∼27kb upstream of *ELL2*, a gene responsible for immunoglobulin mRNA production and transcriptional regulation in plasma cells. Coding and regulatory variants of *ELL2* have been associated with risk of multiple myeloma in European and African ancestry individuals as well as reduced levels of immunoglobulin A and G in healthy subjects ^51–53^. Another set of genetic variants located ∼200kb away in the promoter region of *GLRX* or glutaredoxin-1 (rs10067881, rs17462893, rs57675369) have been associated with higher reticulocyte count in UKBB Europeans ^9^. Glutaredoxin-1 is a cytoplasmic enzyme that catalyzes the reversible reduction of glutathione-protein mixed disulfides and contributes to the antioxidant defense system. Congenital deficiencies of other members of the glutaredoxin enzyme family (*GLRX5*) have been reported in patients with sideroblastic anemia ^54–56^. Notably, our *ELL2* rs116635225 MCH-associated variant remained genome-wide significant after conditioning on the myeloma or reticulocyte-related variants. Therefore, the precise genetic regulatory mechanisms of the red cell trait associations in this region remain to be determined.

### MIDN

The chromosome *19p13* African variant rs73494666 associated with lower MCV/MCH is located in an open chromatin region of an intron of *MIDN*, which encodes the midbrain nucleolar protein midnolin. The gene-rich region on chromosome *19p13* also includes *SBNO2, STK11, CBARP, ATP5F1D, CIRBP, EFNA2*, and *GPX4*. However, none of these genes have clear relationships to hematopoiesis or red structure/function. Other variants in the region have been associated with MCH and RBC count (rs757293) ^13^ or reticulocytes (rs35971149) ^9^. The *MIDN*-rs73494666 variant overlaps ENCODE cis-regulatory elements for CD34 stem cells and other blood cell progenitors.

### PIEZO1

Mutations in the mechanosensitive ion channel *PIEZO1* on chromosome *16q24* have been reported in patients with autosomal dominant hereditary xerocytosis (OMIM 194380), a congenital hemolytic anemia associated with increased calcium influx, red cell dehydration, and potassium efflux along with various red cell laboratory abnormalities including increased MCHC, MHC, and reticulocytosis ^57,58^. Most reported hereditary xerocytosis *PIEZO1* missense mutations are associated with at least partial gain-of-function and are located within the highly conserved C-terminal region near the pore of the ion channel. In some individuals carrying *PIEZO1* missense mutations, mild red cell laboratory parameter alterations without frank hemolytic anemia have been reported ^59^. The *PIEZO1* 3bp short tandem repeat (STR) rs763477215 in-frame coding variant (p.Lys2169del) associated with higher MCHC in TOPMed is extremely rare in all populations except for the Ashkenazi Jewish population (frequency of 1.5% in gnomAD), has not been previously associated with hereditary xerocytosis, and therefore has been reported as ‘benign’ in ClinVar. The p.Lys2169del variant is located in a highly basic -K-K-K-K-motif near the C-terminus of the 36 transmembrane domain protein within a 14-residue linker region between the central ion channel pore and the peripheral propeller-like mechanosensitive domains important for modulating PIEZO1 channel function ^60,61^. Interestingly, another 3bp in-frame deletion of *PIEZO1* (p.Glu657del) reported to be highly enriched in prevalence among African populations was recently associated with dehydrated red blood cells and reduced susceptibility to malaria ^62,63^. In TOPMed, however, we were unable to confirm any association between the rs59446030 (p.Glu657del) putative malaria susceptibility allele variant and phenotypic variation in MCHC (*P*-value for trait-specific conditional analysis=0.21).

### TMPRSS6

*TMPRSS6* on chromosome *22q12* encodes matriptase-2, a transmembrane serine protease that down-regulates the production of hepcidin in the liver and therefore plays an essential role in iron homeostasis ^64^. Rare mutations of *TMPRSS6* are associated with iron-refractory iron deficiency anemia ^65^ characterized by microcytic hypochromic anemia and low transferrin saturation. Several common *TMPRSS6* variants have been associated with multiple RBC traits through prior GWAS. The common *TMPRSS6* intronic variant associated with *TMPRSS6* expression and lower MCH/MCV in TOPMed (rs228914/rs228916) was previously reported to be associated with lower iron levels ^47^, and therefore likely contributes to lower MCH and MCV via iron deficiency. In rare variant aggregated association testing, we were able to identify several additional rare coding missense, stop-gain, or splice variants that appear to drive the gene-based association of *TMPRSS6* with lower MCH/MCV and higher RDW. At least one of these variants at exon 13 rs387907018 (p.Glu5323Lys, loss-of-function mutation) has been reported in a compound heterozygous iron-refractory iron deficiency anemia (IRIDA) patient ^66^, suggesting that inheritance of this or similar LoF variants in the heterozygote state may contribute to mild reductions in MCV/MCH or increased RDW ^65^.

### G6PD

X-linked *G6PD* mutations (glucose-6-phosphate dehydrogenase) are the most common cause worldwide of acute and chronic hemolytic anemia. The *G6PD-*rs76723693 (c.968T>C) low-frequency missense variant (p.Leu323Pro, referred to as *G6PD* Nefza^67^) associated is common in persons of African ancestry and is associated with lower RDW in TOPMed. In persons of African ancestry, the p.Leu323Pro variant is often co-inherited with another *G6PD* missense variant, p.Asn126Asp, encoded by rs1050829 (c.A376>G). The 968C/376G haplotype in African ancestry individuals constitutes one of several forms of the *G6PD* variant A-^68–71^. Functional studies of the p.Leu323Pro, p.Asn126Asp, and the double mutant suggest the p.Leu323Pro variant is the primary contributor to reduced catalytic activity^72^. In the US, another African ancestry *G6PD* A-variant is due to the haplotypic combination of rs1050829 (p.Asn126Asp0) and rs1050828 (p.Val68Met), which has an allele frequency of ∼12%. Our finding that rs76723693 is significantly associated with lower RDW after conditioning on rs1050828 is consistent with the independence of effects of the *G6PD* Nefza and A-variants on red cell physiology and morphology. Importantly, both rs76723693 and rs1050828 *G6PD* variants were recently reported to have the effect of lowering hemoglobin A1c (HbA1c) values and therefore should be considered when screening African Americans for type-2 diabetes ^73^

In gene-based analyses, several additional *G6PD* missense variants contributed to the aggregated rare variant association signals for MCH, MCV, RBC, and RDW, including the class II Southeast Asian Mahidol variant p.Gly163Ser ^74^ and the class II Union variant (p.Arg454Cys) ^75^. For a third previously reported variant associated with G6PD deficiency, the East Asian class II Gahoe variant (p.His32Arg) ^76^, there is conflicting evidence of pathogenicity in ClinVar. Of the two female rs137852340 (p.His32Arg) variant allele carriers in TOPMed, one has a normal RDW and one has an elevated RDW. These findings add to the further genotypic-phenotypic complexity and clinical spectrum of G6PD deficiency, which is influenced its sex-linkage and zygosity, residual *G6PD* variant enzyme activity and stability, genetic background, and environmental exposures ^77^.

### HBB

Heterozygosity for the common African *HBB-*rs334 hemoglobin S (p.Glu7Val) or rs33930165 hemoglobin C (p.Glu7Gln) beta-globin structural variants have recently been associated with alterations in various red cell laboratory parameters including lower hemoglobin, MCV, MCH, and RDW, along with higher MCHC, RDW, and HbA1c ^17,18,20,78–80^. In TOPMed, we were able to identify at least 10 additional low-frequency or rare variants within the *HBB* locus independently associated with HGB, RBC, MCV, MCH, MCHC, and/or RDW. Notably, six of the 10 variants correspond to *HBB* 5’ UTR and promoter regions (rs34598529 or -29 A>G ^81^; rs33944208 or -88C>T ^82–84^; splice site (rs33915217 or IVS1-5G>C) ^85 82^; rs33945777 or IVS2-1G>A ^82,85^; rs35004220 or IVS-I-110 G->A ^86,87^, and nonsense mutations (rs11549407 or p.Gln40Ter) ^88,89^ previously identified in patients with beta-thalassemia. These findings confirm the very mild phenotype and clinically “silent” nature of the heterozygote carrier state of these beta-globin gene variants ^90^. Several of these mutations occur more commonly in populations of South Asian (rs33915217), African (rs34598529, rs33944208), or Mediterranean (rs11549407) ancestry. Four additional association signals in the region rs73404549 (*HBG2*), rs77333754, rs1189661759, and rs539384429 are all rare non-coding variants without obvious functional consequences. In addition to the *HBB* protein-coding variants identified in single variant analyses, several of the rare variants driving the aggregate *HBB* gene-based association with lower HGB/HCT and MCH/MCV/MCHC and higher RBC/RDW are similarly previously reported missense, frameshift or nonsense mutations previously identified in beta-thalassemia patients and categorized as pathogenic in ClinVar (**Supplemental Fig. 5** and **Supplemental Table 15**).

### HBA1/HBA2

Several common DNA polymorphisms located in the α-globin gene cluster on chromosome 16p13.3 have been associated with red cell traits in large GWAS ^7,8,91^, including heterozygosity for the common African ancestral 3.7kb deletion which contributes to quantitative RBC phenotypes among African Americans and Hispanics/Latinos. In TOPMed, we identified two low-frequency variants in single variant testing associated with MCH, MCV, and/or RBC count, independently of the 3.7kb deletion. The rs868351380 variant is found primarily among Hispanics/Latinos while the rs372755452 variant is found primarily among East Asians. Neither of these two non-coding variants is located in any known alpha-globin regulatory region, and therefore requires further mechanistic confirmation. By contrast, in gene-based rare variant analysis, we identified several known alpha-globin variants associated in aggregate with lower MCH and MCV including the South Asian variant Hb Q India (*HBA1*, p.Asp64His) ^92–94^ and the African variant Hb Groene Hart (*HBA1*, p.Pro120Ser) ^95–97^. In homozygous or compound heterozygous forms, these latter variants have been reported in probands with alpha-thalassemia, whereas heterozygotes generally have mild microcytic phenotype. Several additional variants contributing to the *HBA1* gene-based rare variant MCH/MCV signal (e.g., a 1 bp indel causing frameshift p.Asn79Ter) may represent previously undetected alpha-thalassemia mutations.

### CD36, TFRC and SLC12A7

The presence of rare coding or LoF variants within *CD36, TFRC* and *SLC12A7* provides evidence that these genes are causally responsible for RBC phenotypic variation. A common African ancestral null variant of CD36 (rs3211938 or p.Tyr325Ter) has been previously associated with higher RDW and with lower CD36 expression in erythroblasts ^98^. In TOPMed, additional *CD36* rare coding variants were associated in aggregate with higher RDW independent of rs3211938, including several nonsense and frameshift or splice acceptor mutations, which have been previously classified as VUS. Further characterization of the genetic complexity of the CD36□null phenotype (common in African and Asian populations) may provide information relevant to the tissue-specific expression of this receptor on red cells, platelets, monocytes, and endothelial cells and its role in malaria infection and disease severity^99^. *TFRC* encodes the transferrin receptors (TfR1), which is required for iron uptake and erythropoiesis ^100^. While common non-coding variants of *TFRC* have been associated with MCV and RDW, the only known *TFRC*-related Mendelian disorder is a homozygous p.Tyr20His substitution reported to cause combined immunodeficiency affecting leukocytes and platelets but not red cells ^101^. Common variants of *SLC12A7* encoding the potassium ion channel KCC4 have been associated with RDW and other RBC phenotypes. While KCC4 is expressed in erythroblasts ^102^, its role in red blood cell function is not well-described ^103^. Further characterization of KCC4 LoF variants may illuminate the role of this ion transporter in red cell dehydration with potential implications for treatment of patients with sickle cell disease ^104^.

In summary, we illustrate that expanding coverage of the genome using WGS as applied to large, population-based multi-ethnic samples can lead to discovery of novel variants associated with quantitative RBC traits. Most of the newly discovered variants were of low frequency and/or disproportionately observed in non-Europeans. We also report extensive allelic heterogeneity at the chromosome 11 beta-globin locus, including associations with several known beta-thalassemia carrier variants. The gene-based association of rare variants within *HBA1/HBA2, HBB, TMPRSS6, G6PD, CD36, TFRC* and *SLC12A7* independent of known single variants in the same genes further suggest that rare functional variants in genes responsible for Mendelian RBC disorders contribute to the genetic architecture of RBC phenotypic variation among the population at large. Together these results demonstrate the utility of WGS in ethnically-diverse population-based samples for expanding our understanding of the genetic architecture of quantitative hematologic traits and suggest a continuum between complex traits and Mendelian red cell disorders.

## Data Availability

Data from TOPMed are available via dbGaP.

## Acknowledgements

Molecular data for the Trans-Omics in Precision Medicine (TOPMed) program was supported by the National Heart, Lung and Blood Institute (NHLBI). The table below presents study specific omics support information. Core support including centralized genomic read mapping and genotype calling, along with variant quality metrics and filtering were provided by the TOPMed Informatics Research Center (3R01HL-117626-02S1; contract HHSN268201800002I). Core support including phenotype harmonization, data management, sample-identity QC, and general program coordination were provided by the TOPMed Data Coordinating Center (R01HL-120393; U01HL-120393; contract HHSN268201800001I). We gratefully acknowledge the studies and participants who provided biological samples and data for TOPMed.

Paul S. de Vries was supported by American Heart Association grant number 18CDA34110116. H.C. and E.J. were supported by the National Eye Institute (NEI) grant R01 EY027004, the National Institute of Diabetes and Digestive and Kidney Diseases (NIDDK) R01 DK116738 and by the National Cancer Institute (NCI) R01CA2416323. M.P.C and D.J were supported by NHLBI grant U01HL137162. D.E.B. was supported by NHLBI P01HL032262, DP2HL137300, R01HL130733. B.P.K. was supported by NCI R00 CA218870 and NHLBI P01HL142494.

---

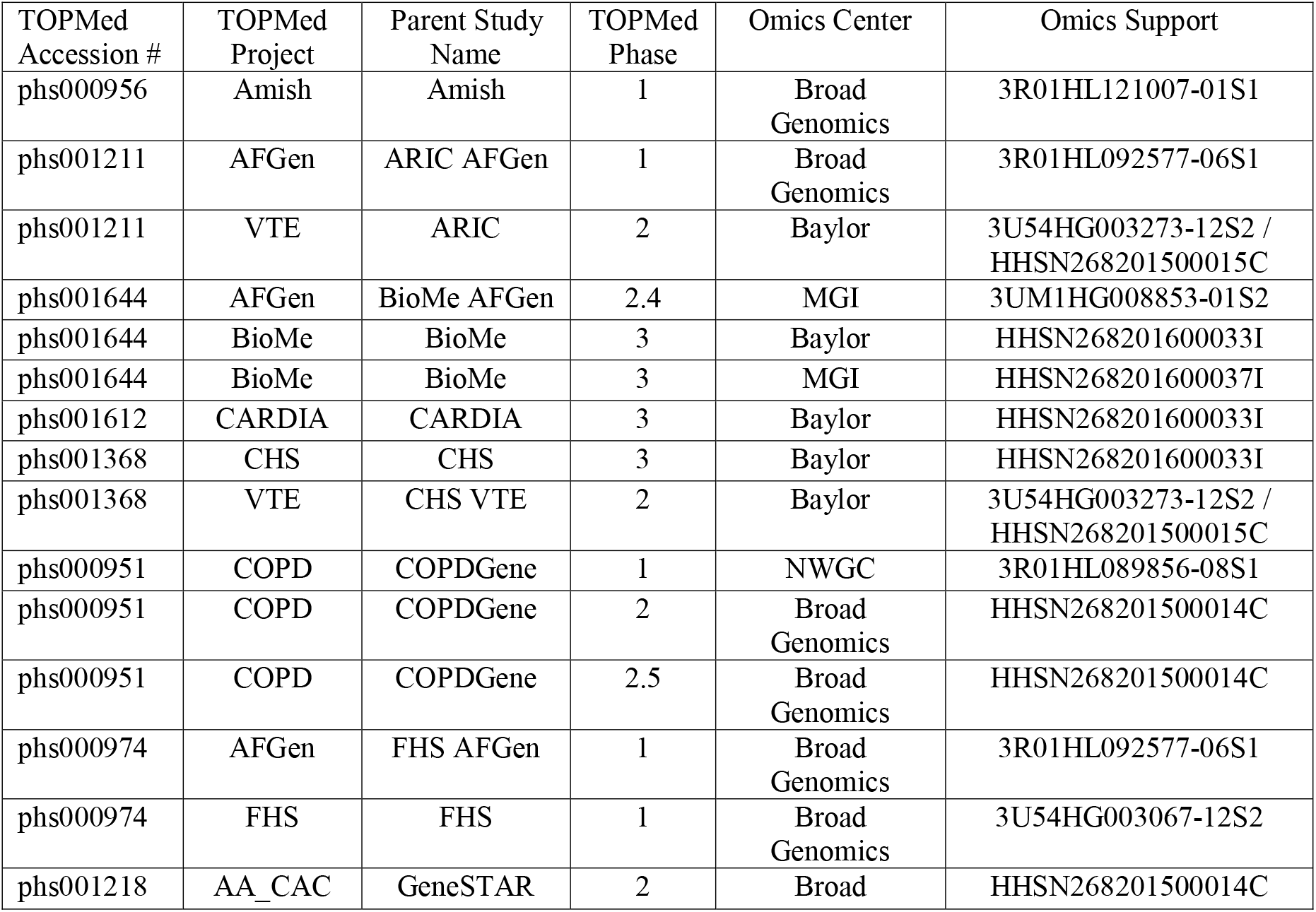
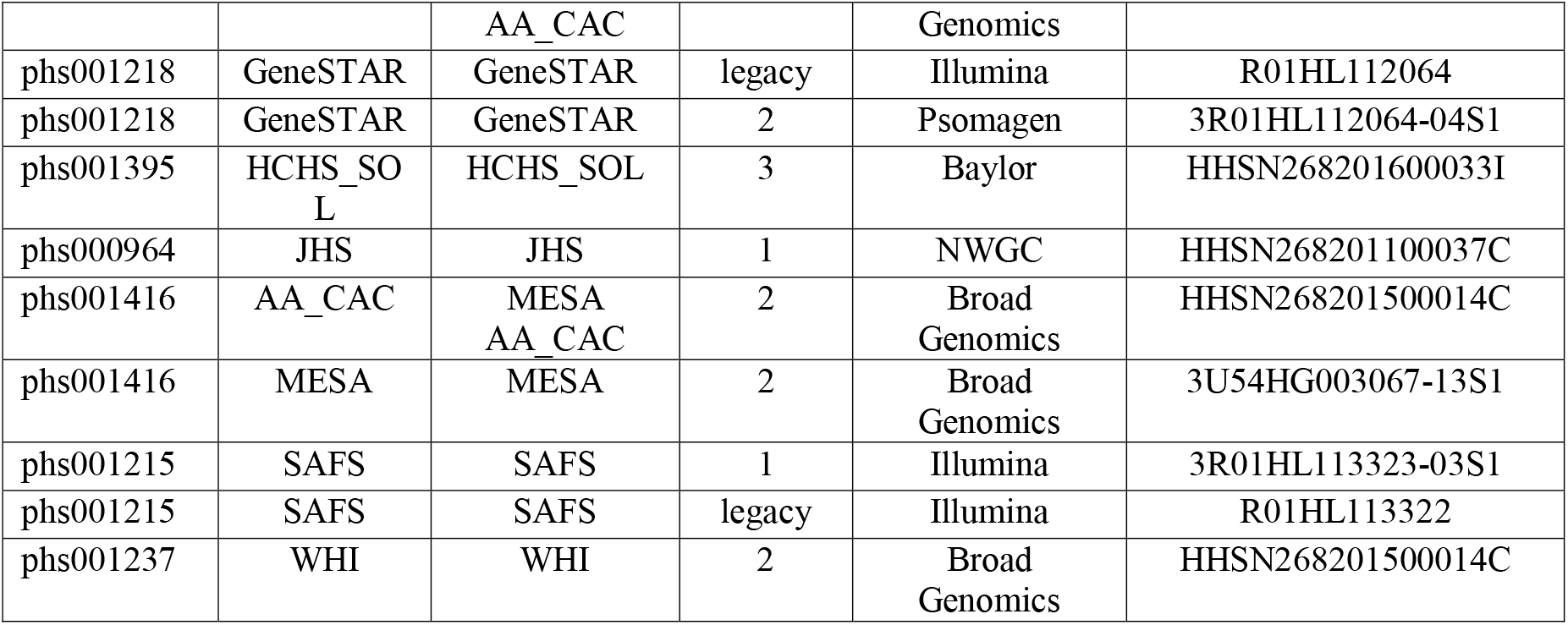

Amish: The TOPMed component of the Amish Research Program was supported by NIH grants R01 HL121007, U01 HL072515, and R01 AG18728. Email Rhea Cosentino for additional input.

ARIC: The Atherosclerosis Risk in Communities study has been funded in whole or in part with Federal funds from the National Heart, Lung, and Blood Institute, National Institutes of Health, Department of Health and Human Services (contract numbers HHSN268201700001I, HHSN268201700002I, HHSN268201700003I, HHSN268201700004I and HHSN268201700005I). The authors thank the staff and participants of the ARIC study for their important contributions.

BioMe: The Mount Sinai BioMe Biobank has been supported by The Andrea and Charles Bronfman Philanthropies and in part by Federal funds from the NHLBI and NHGRI (U01HG00638001; U01HG007417; X01HL134588). We thank all participants in the Mount Sinai Biobank. We also thank all our recruiters who have assisted and continue to assist in data collection and management and are grateful for the computational resources and staff expertise provided by Scientific Computing at the Icahn School of Medicine at Mount Sinai.

CARDIA: The Coronary Artery Risk Development in Young Adults Study (CARDIA) is conducted and supported by the National Heart, Lung, and Blood Institute (NHLBI) in collaboration with the University of Alabama at Birmingham (HHSN268201800005I & HHSN268201800007I), Northwestern University (HHSN268201800003I), University of Minnesota (HHSN268201800006I), and Kaiser Foundation Research Institute (HHSN268201800004I). CARDIA was also partially supported by the Intramural Research Program of the National Institute on Aging (NIA) and an intra□agency agreement between NIA and NHLBI (AG0005).

CHS: Cardiovascular Health Study: This research was supported by contracts HHSN268201200036C, HHSN268200800007C, HHSN268201800001C, N01HC55222, N01HC85079, N01HC85080,

N01HC85081, N01HC85082, N01HC85083, N01HC85086, and grants U01HL080295 and U01HL130114 from the National Heart, Lung, and Blood Institute (NHLBI), with additional contribution from the National Institute of Neurological Disorders and Stroke (NINDS). Additional support was provided by R01AG023629 from the National Institute on Aging (NIA). A full list of principal CHS investigators and institutions can be found at CHS-NHLBI.org. The content is solely the responsibility of the authors and does not necessarily represent the official views of the National Institutes of Health.

COPDGene: The COPDGene project described was supported by Award Number U01 HL089897 and Award Number U01 HL089856 from the National Heart, Lung, and Blood Institute. The content is solely the responsibility of the authors and does not necessarily represent the official views of the National Heart, Lung, and Blood Institute or the National Institutes of Health. The COPDGene project is also supported by the COPD Foundation through contributions made to an Industry Advisory Board comprised of AstraZeneca, Boehringer Ingelheim, GlaxoSmithKline, Novartis, Pfizer, Siemens and Sunovion. A full listing of COPDGene investigators can be found at: http://www.copdgene.org/directory FHS: The Framingham Heart Study (FHS) acknowledges the support of contracts NO1-HC-25195 and HHSN268201500001I from the National Heart, Lung and Blood Institute and grant supplement R01 HL092577-06S1 for this research. We also acknowledge the dedication of the FHS study participants without whom this research would not be possible.

GeneSTAR: GeneSTAR was supported by the National Institutes of Health/National Heart, Lung, and Blood Institute (U01 HL72518, HL087698, HL112064, HL11006, HL118356) and by a grant from the National Institutes of Health/National Center for Research Resources (M01-RR000052) to the Johns Hopkins General Clinical Research Center. We would like to thank our participants and staff for their valuable contributions.

HCHS/SOL: The Hispanic Community Health Study/Study of Latinos is a collaborative study supported by contracts from the National Heart, Lung, and Blood Institute (NHLBI) to the University of North Carolina (HHSN268201300001I / N01-HC-65233), University of Miami (HHSN268201300004I / N01-HC-65234), Albert Einstein College of Medicine (HHSN268201300002I / N01-HC-65235), University of Illinois at Chicago – HHSN268201300003I / N01-HC-65236 Northwestern Univ), and San Diego State University (HHSN268201300005I / N01-HC-65237). The following Institutes/Centers/Offices have contributed to the HCHS/SOL through a transfer of funds to the NHLBI: National Institute on Minority Health and Health Disparities, National Institute on Deafness and Other Communication Disorders, National Institute of Dental and Craniofacial Research, National Institute of Diabetes and Digestive and Kidney Diseases, National Institute of Neurological Disorders and Stroke, NIH Institution-Office of Dietary Supplements.

JHS: The Jackson Heart Study (JHS) is supported and conducted in collaboration with Jackson State University (HHSN268201300049C and HHSN268201300050C), Tougaloo College (HHSN268201300048C), and the University of Mississippi Medical Center (HHSN268201300046C and HHSN268201300047C) contracts from the National Heart, Lung, and Blood Institute (NHLBI) and the National Institute for Minority Health and Health Disparities (NIMHD). The authors also wish to thank the staffs and participants of the JHS.

MESA: MESA and the MESA SHARe project are conducted and supported by the National Heart, Lung, and Blood Institute (NHLBI) in collaboration with MESA investigators. Support for MESA is provided by contracts 75N92020D00001, HHSN268201500003I, N01-HC-95159, 75N92020D00005, N01-HC-95160, 75N92020D00002, N01-HC-95161, 75N92020D00003, N01-HC-95162, 75N92020D00006, N01-HC-95163, 75N92020D00004, N01-HC-95164, 75N92020D00007, N01-HC-95165, N01-HC-95166, N01-HC-95167, N01-HC-95168, N01-HC-95169, UL1-TR-000040, UL1-TR-001079, UL1-TR-001420.

MESA Family is conducted and supported by the National Heart, Lung, and Blood Institute (NHLBI) in collaboration with MESA investigators. Support is provided by grants and contracts R01HL071051, R01HL071205, R01HL071250, R01HL071251, R01HL071258, R01HL071259, by the National Center for Research Resources, Grant UL1RR033176. The provision of genotyping data was supported in part by the National Center for Advancing Translational Sciences, CTSI grant UL1TR001881, and the National Institute of Diabetes and Digestive and Kidney Disease Diabetes Research Center (DRC) grant DK063491 to the Southern California Diabetes Endocrinology Research Center.

SAFS: Collection of the San Antonio Family Study data was supported in part by National Institutes of Health (NIH) grants R01 HL045522, MH078143, MH078111 and MH083824; and whole genome sequencing of SAFS subjects was supported by U01 DK085524 and R01 HL113323. We are very grateful to the participants of the San Antonio Family Study for their continued involvement in our research programs.

WHI: The WHI program is funded by the National Heart, Lung, and Blood Institute, National Institutes of Health, U.S. Department of Health and Human Services through contracts HHSN268201600018C, HHSN268201600001C, HHSN268201600002C, HHSN268201600003C, and HHSN268201600004C.

