## Supplemental Figures for "Whole genome sequencing association analysis of quantitative red blood cell phenotypes: the NHLBI TOPMed program"

**Figure S1. Rs112097551 C-to-T base editing and single cell cloning in HUDEP-2 cells.** (A) Scheme of rs112097551 C-to-T base editing and FACS-based single cell separation. (B) Efficiency of rs112097551 C-to-T (G-to-A on opposing strand) base editing efficiency in all five clones. Since base editor and sgRNA are constitutively expressed, the frequency of C-to-T conversion may exceed 50% in heterozygous clones due to base editing after single cell cloning.

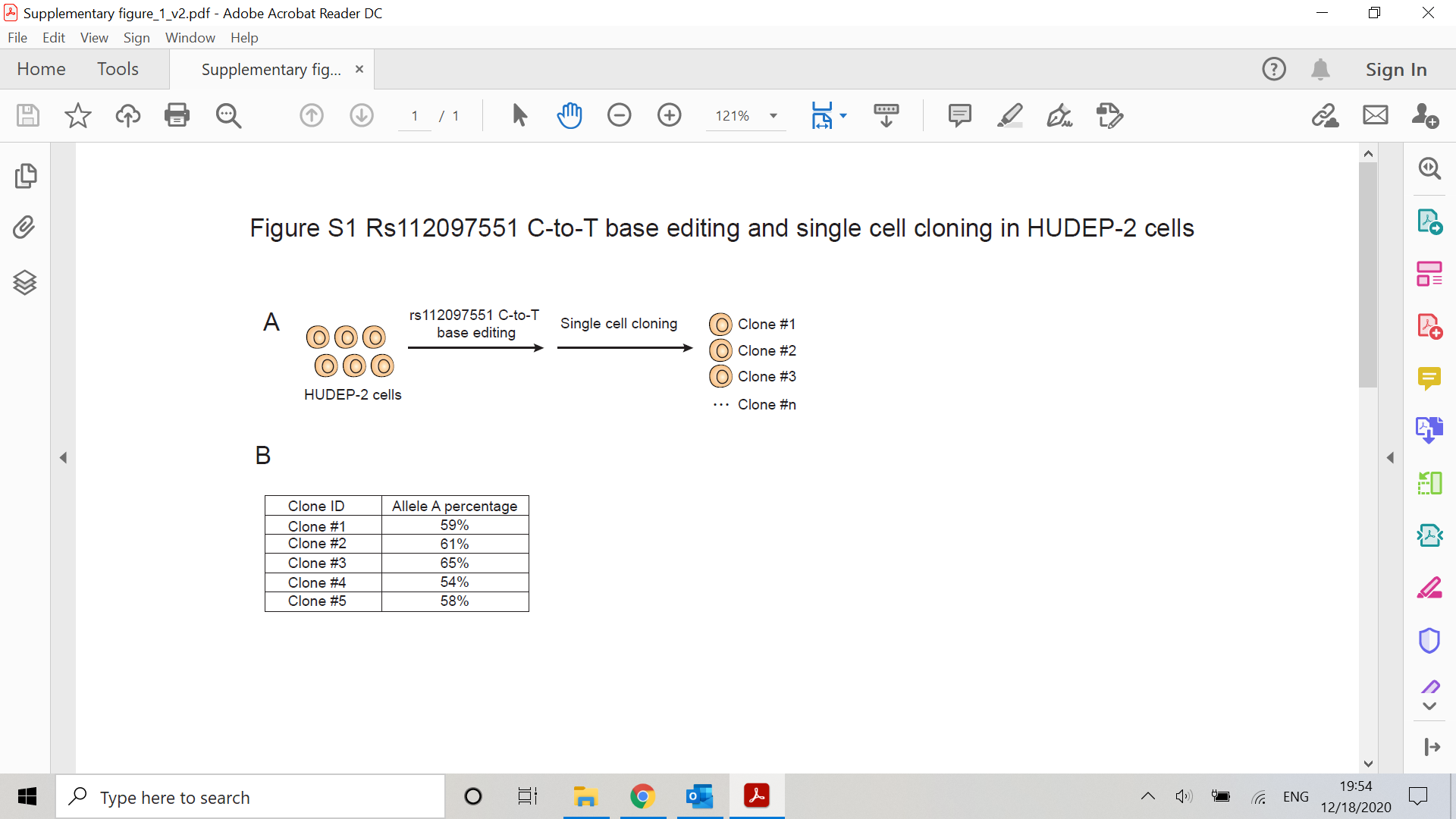

**Figure S2. Manhattan plots of the marginal single-variant analyses in TOPMed. (A) HCT; (B) HGB; (C) MCH; (D) MCHC; (E) MCV; (F) RBC; (G) RDW.**

(A)

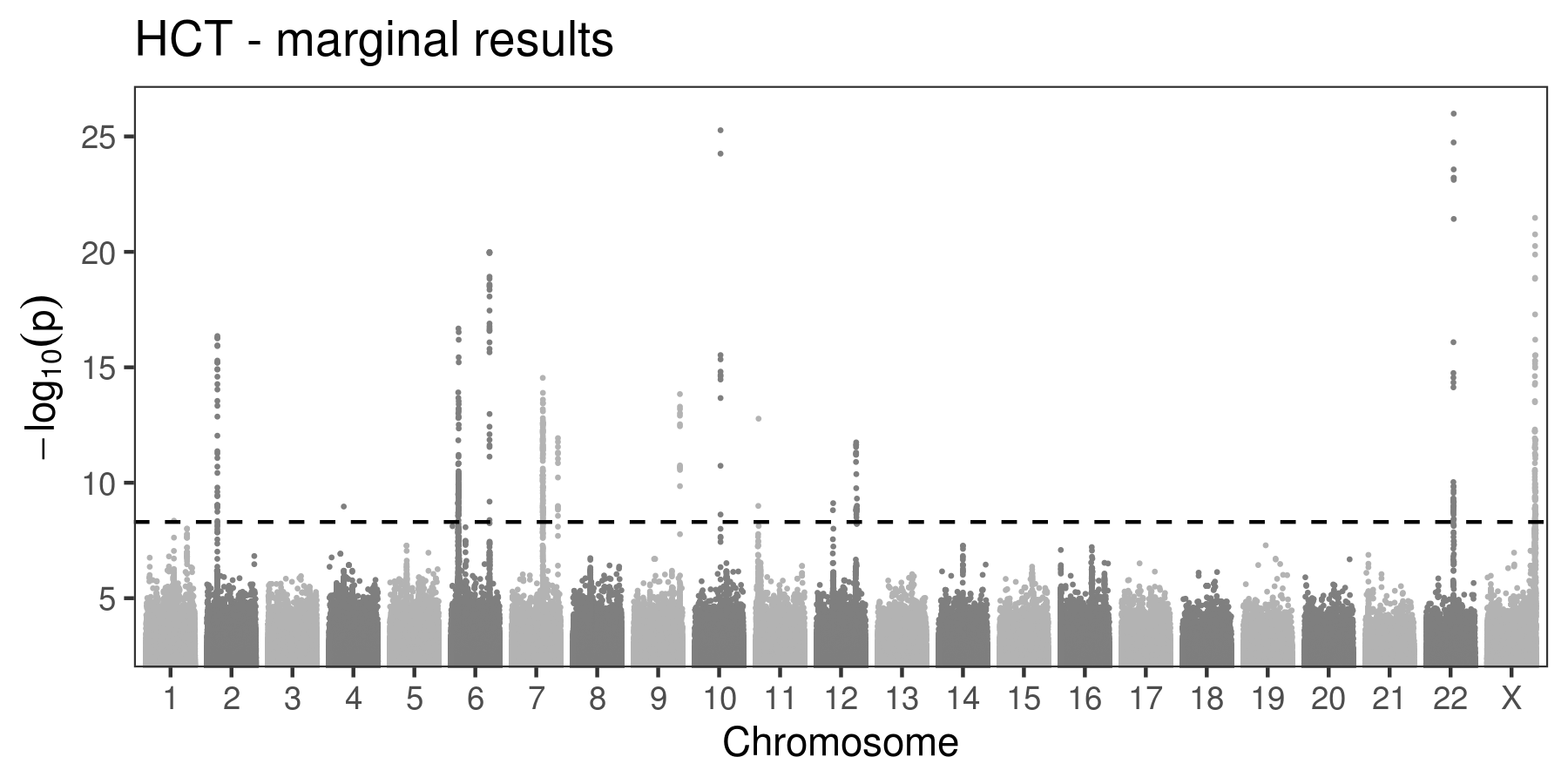

(B)

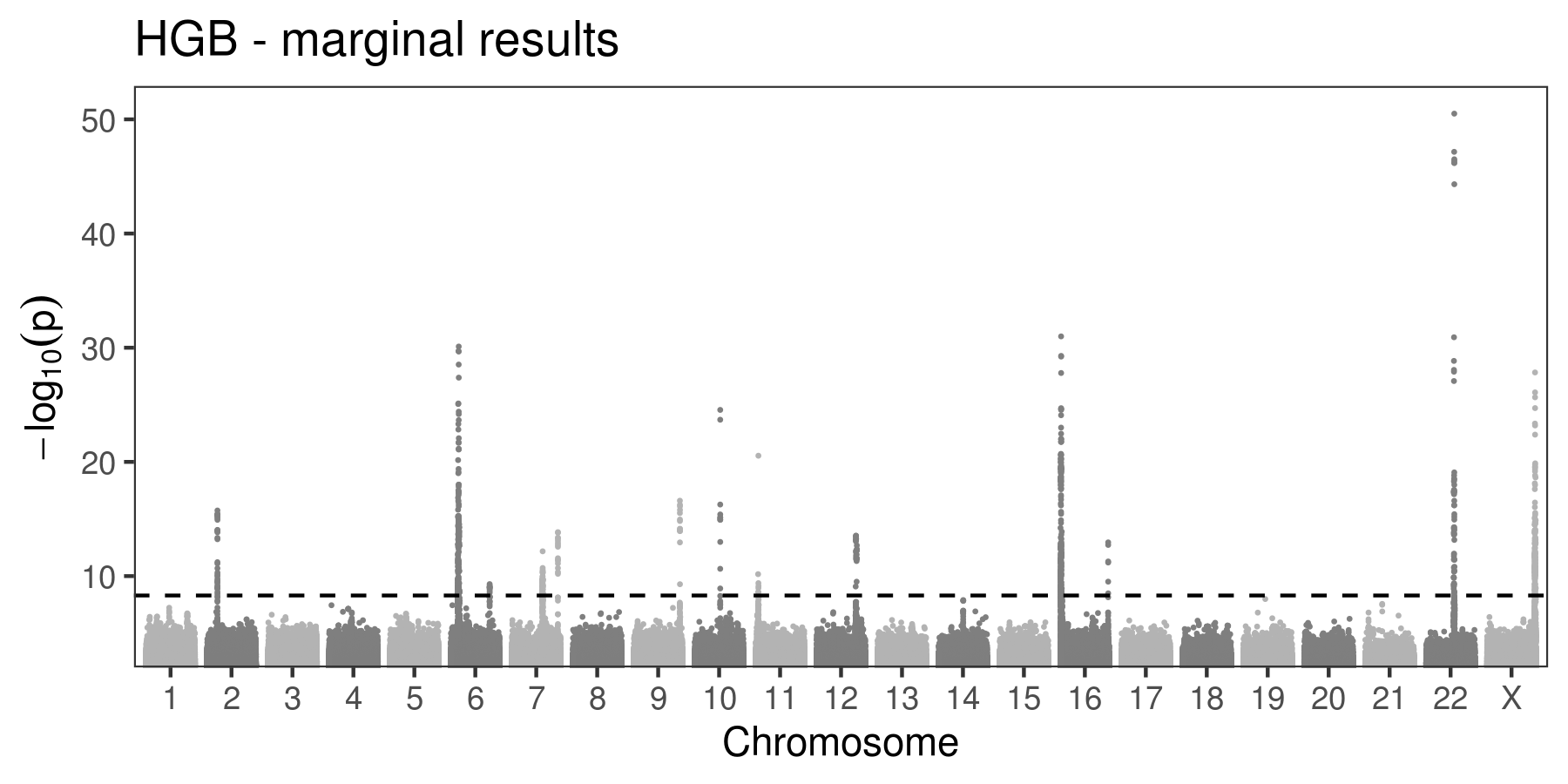

(C)

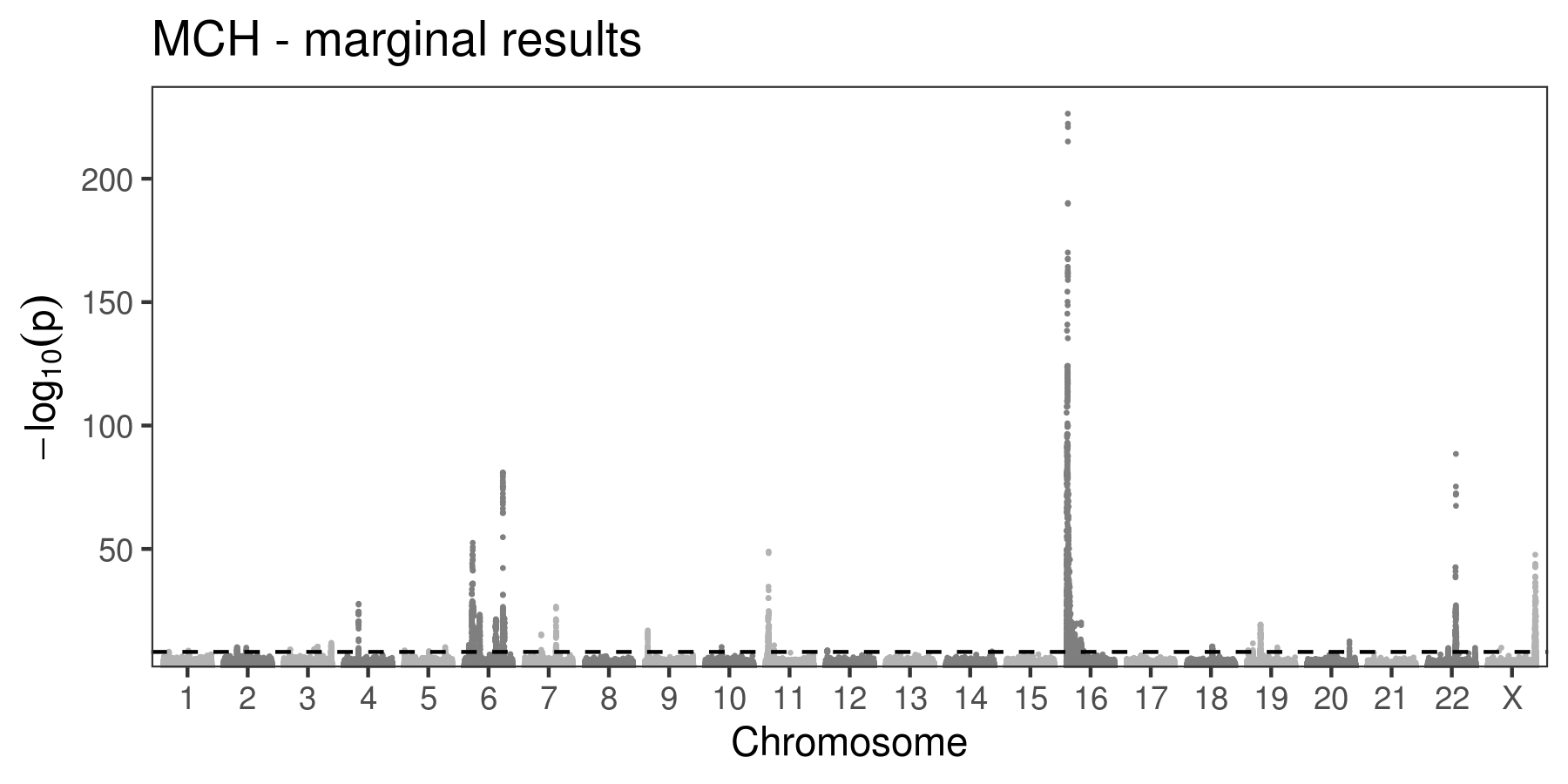

(D)

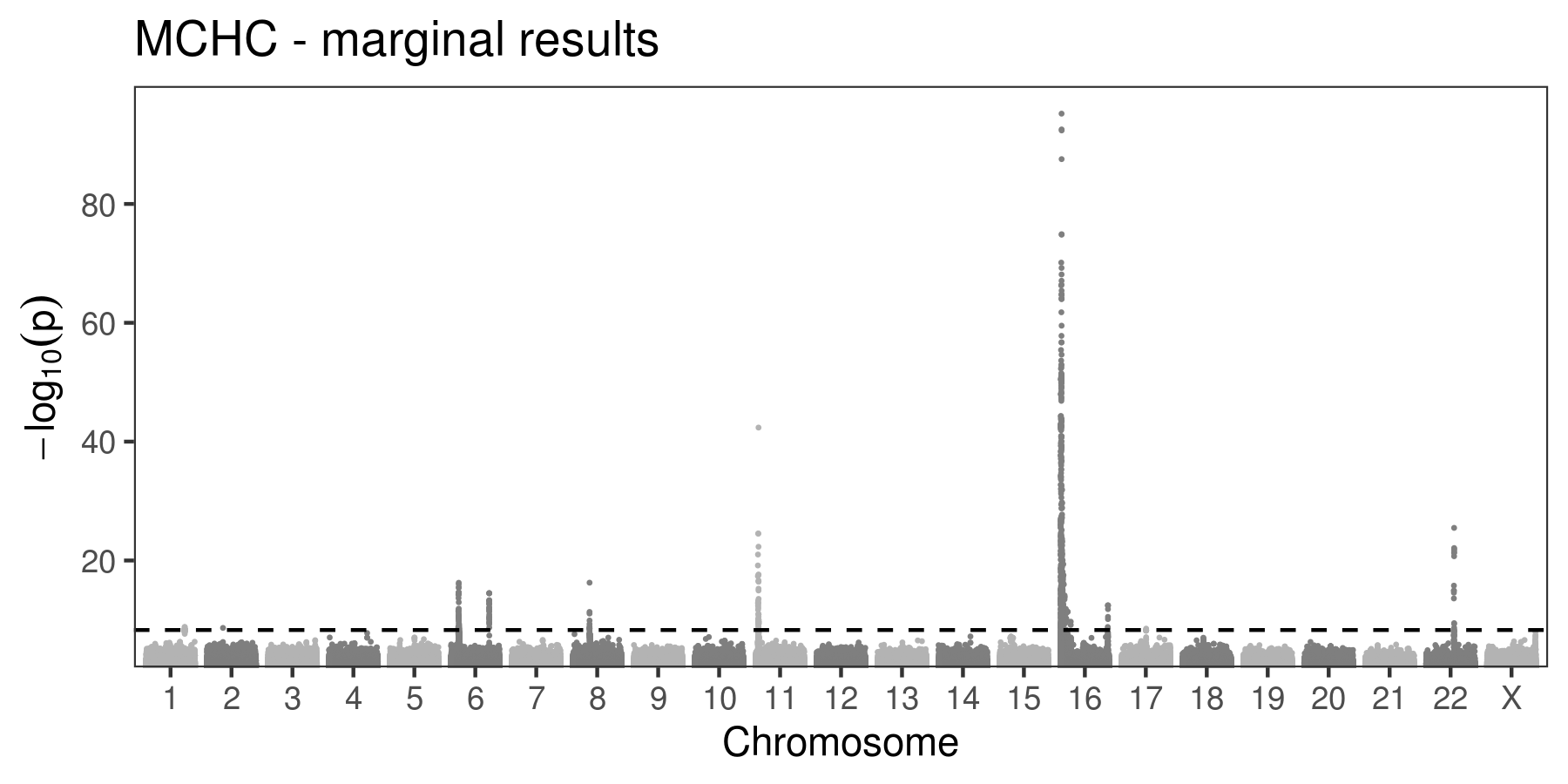

(E)

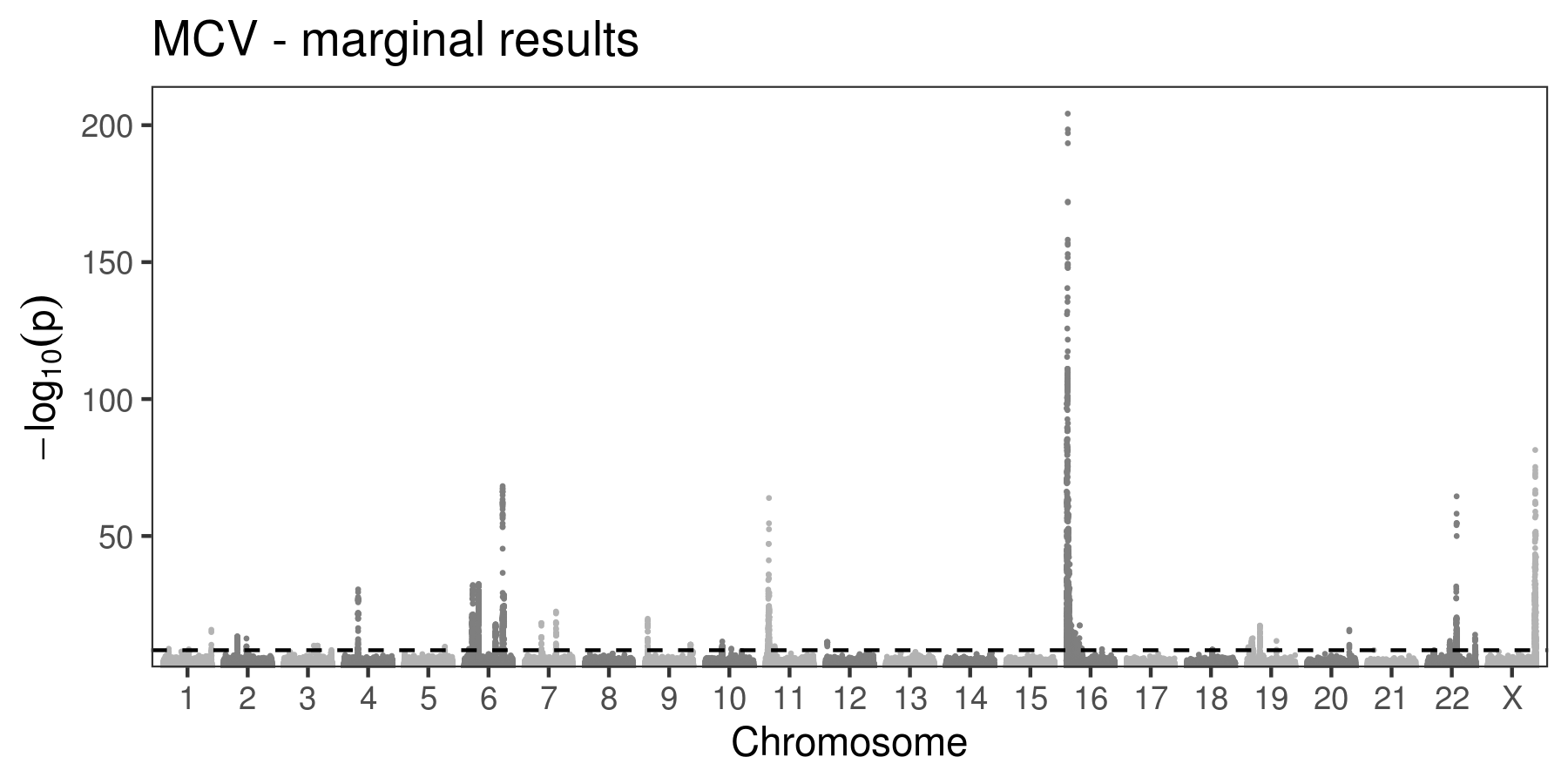

(F)

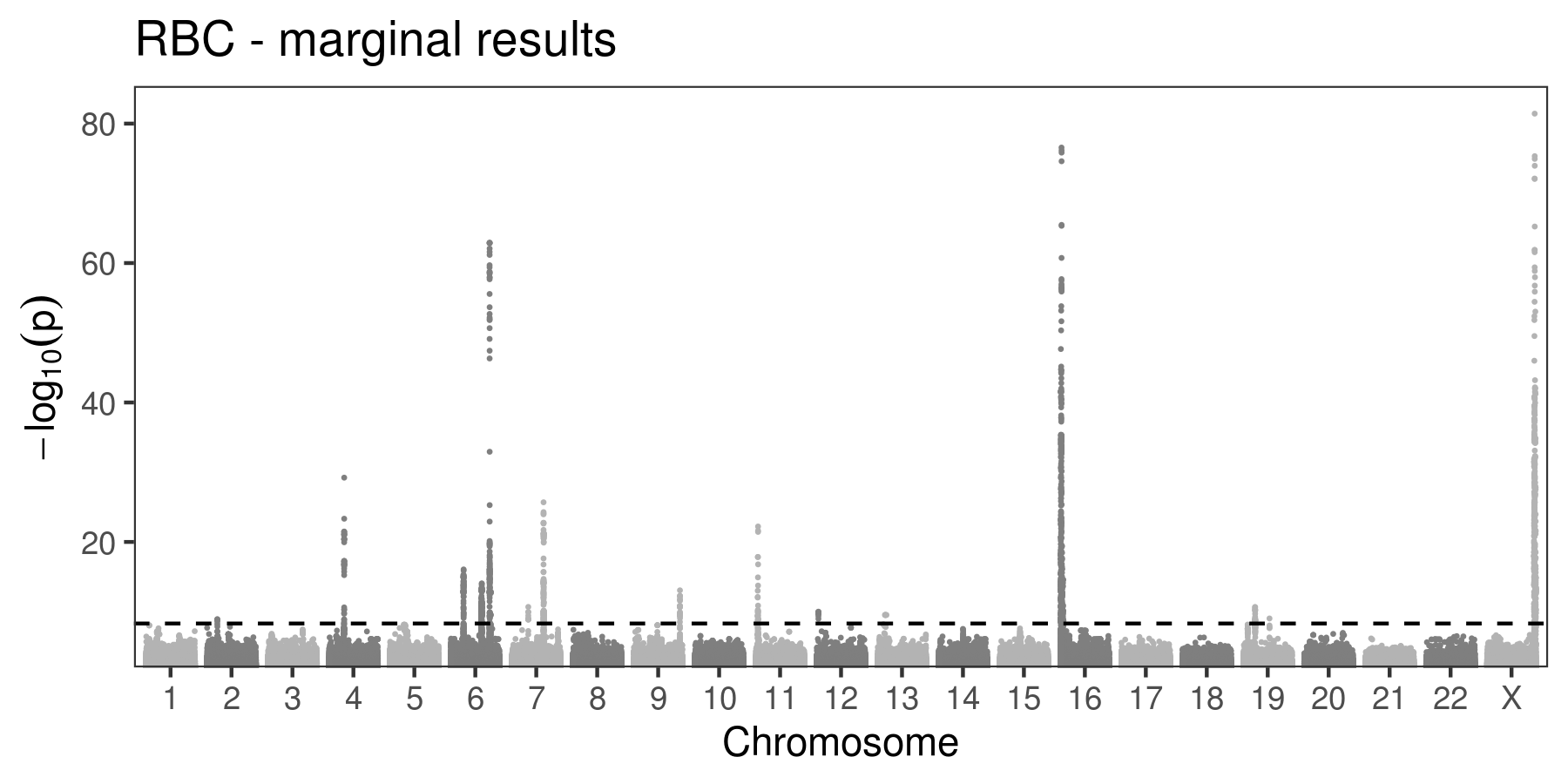

(G)

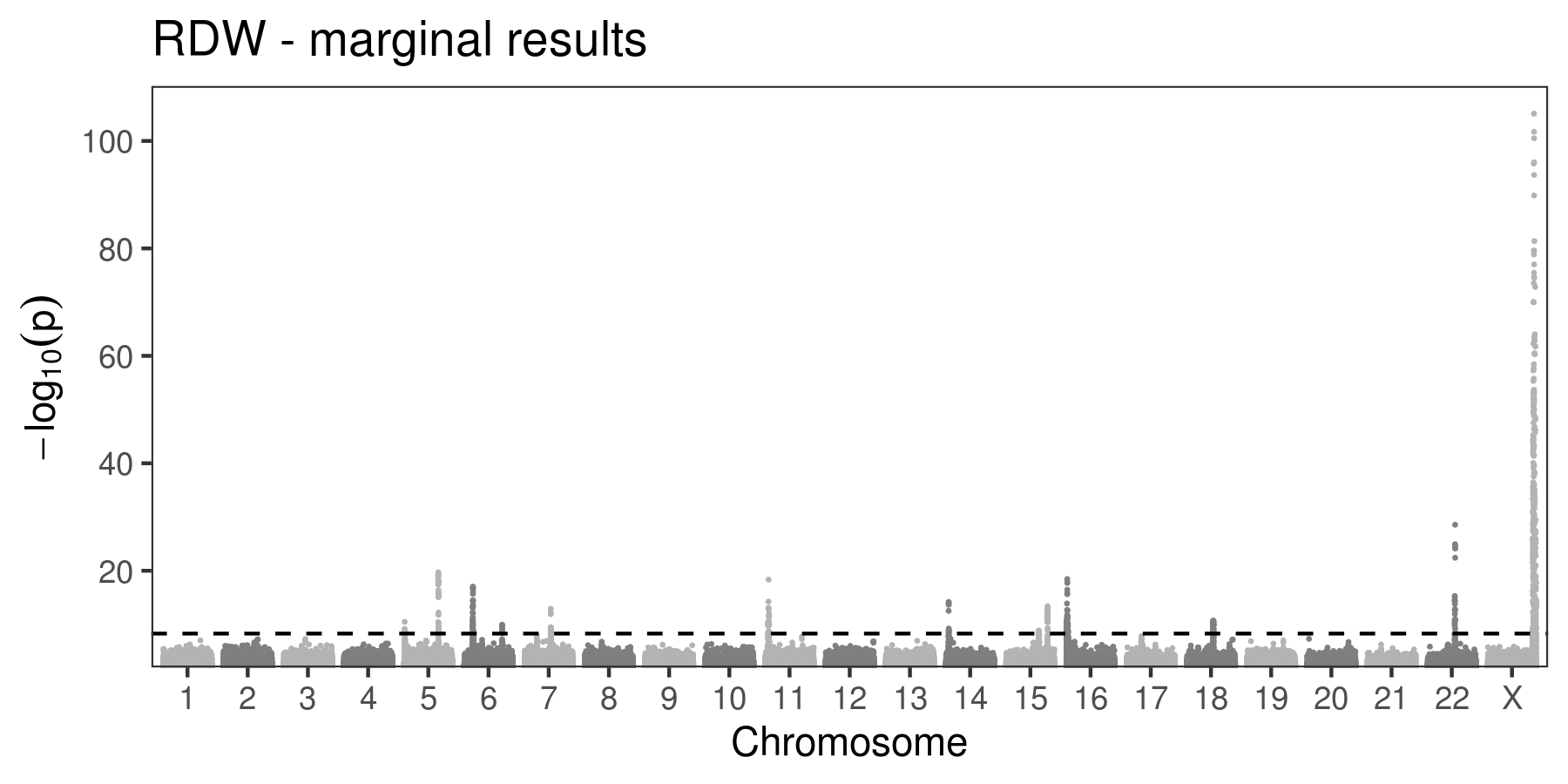

**Figure S3. Manhattan plots of the trait-specific conditional single-variant analyses in TOPMed. (A) HCT; (B) HGB; (C) MCH; (D) MCHC; (E) MCV; (F) RBC; (G) RDW.**

(A)

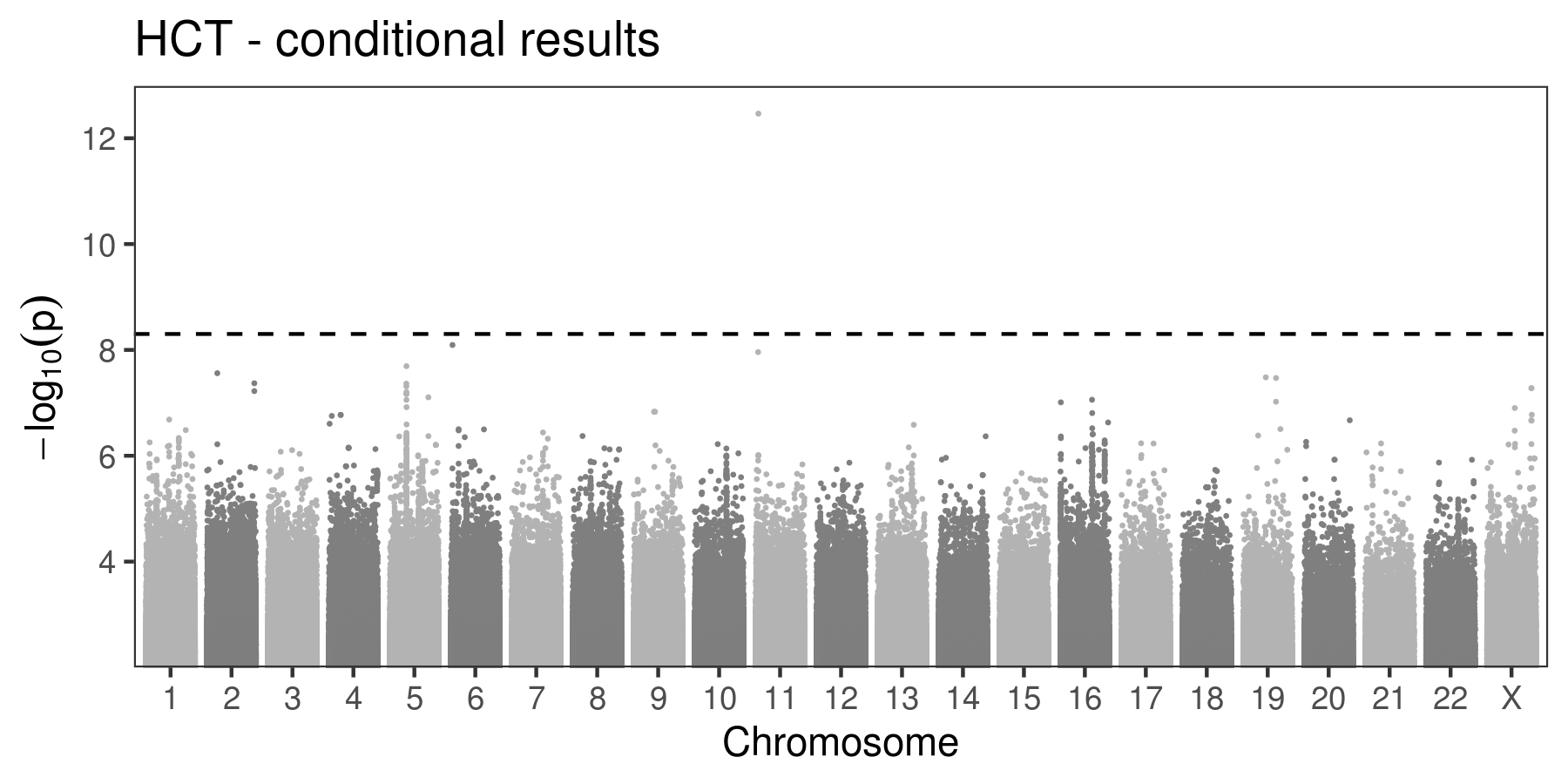

(B)

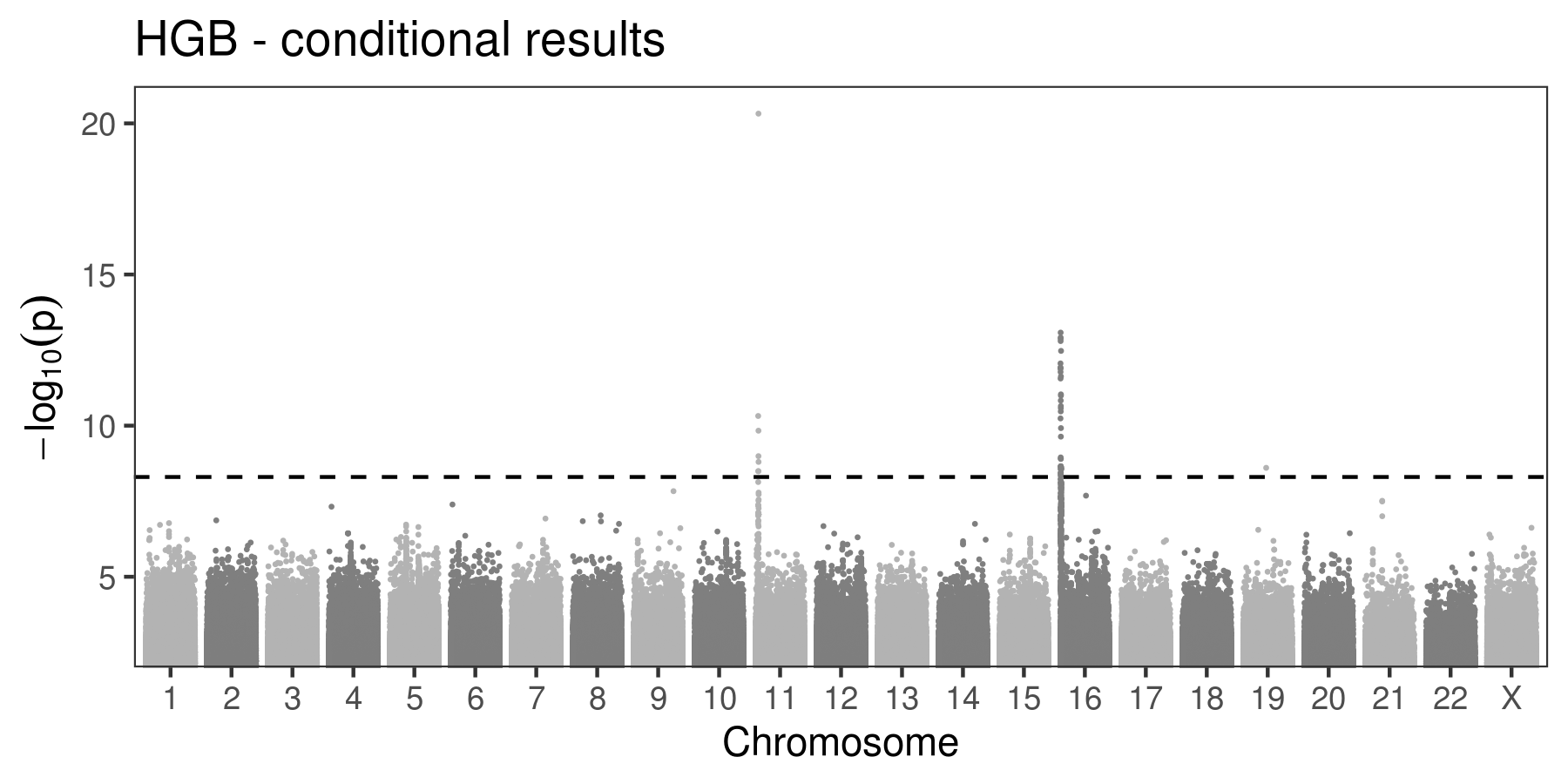

(C)

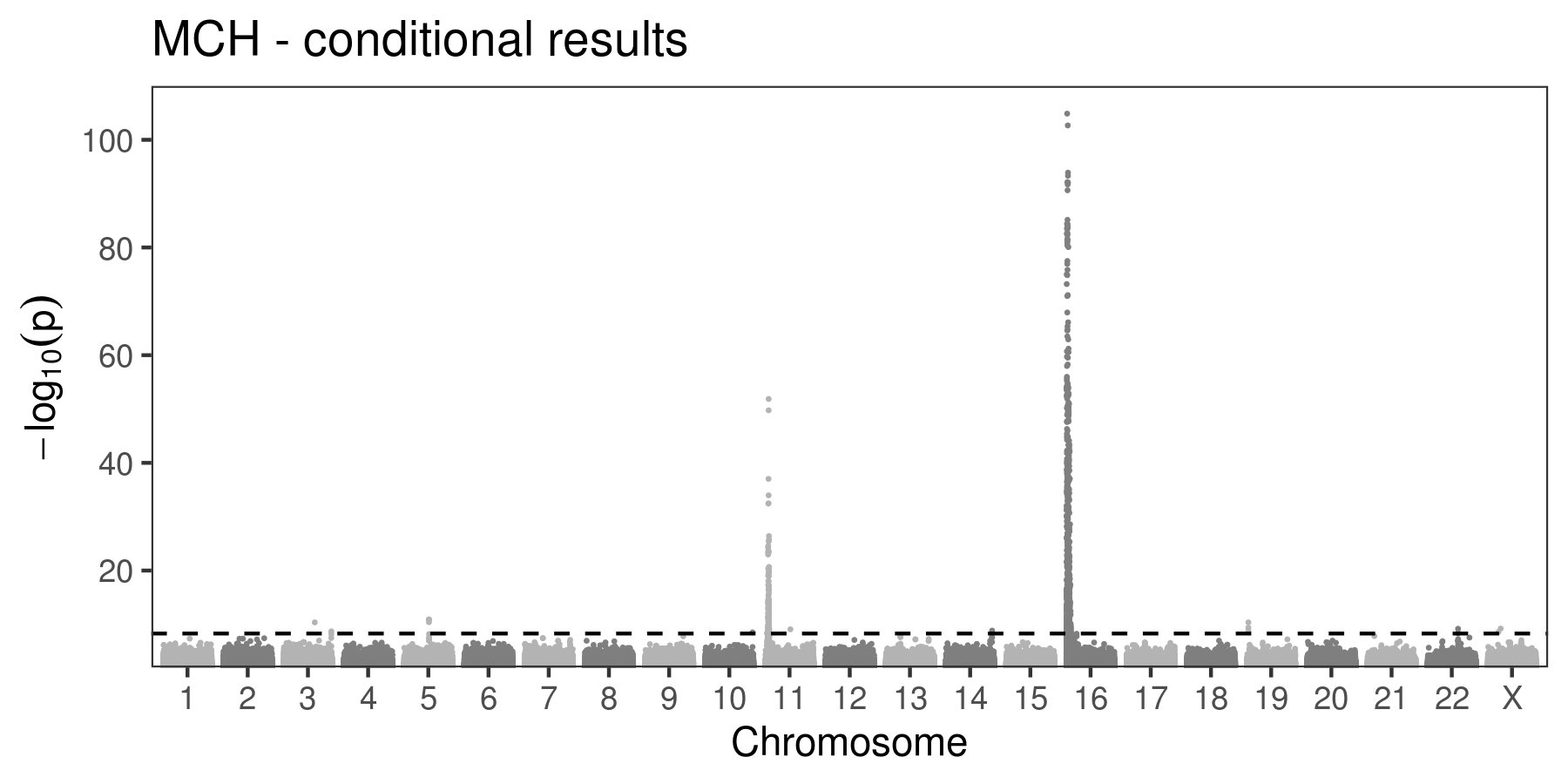

(D)

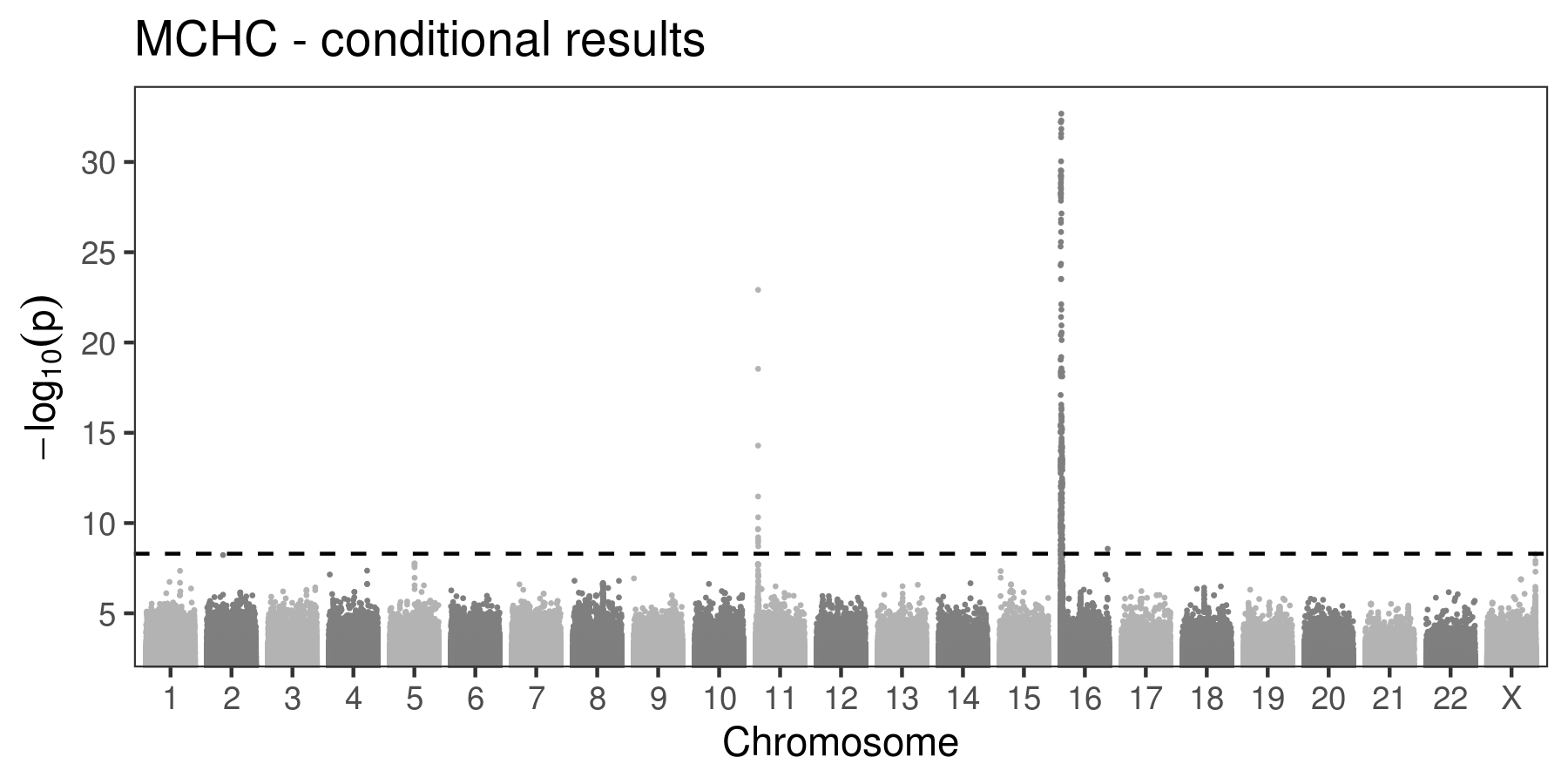

(E)

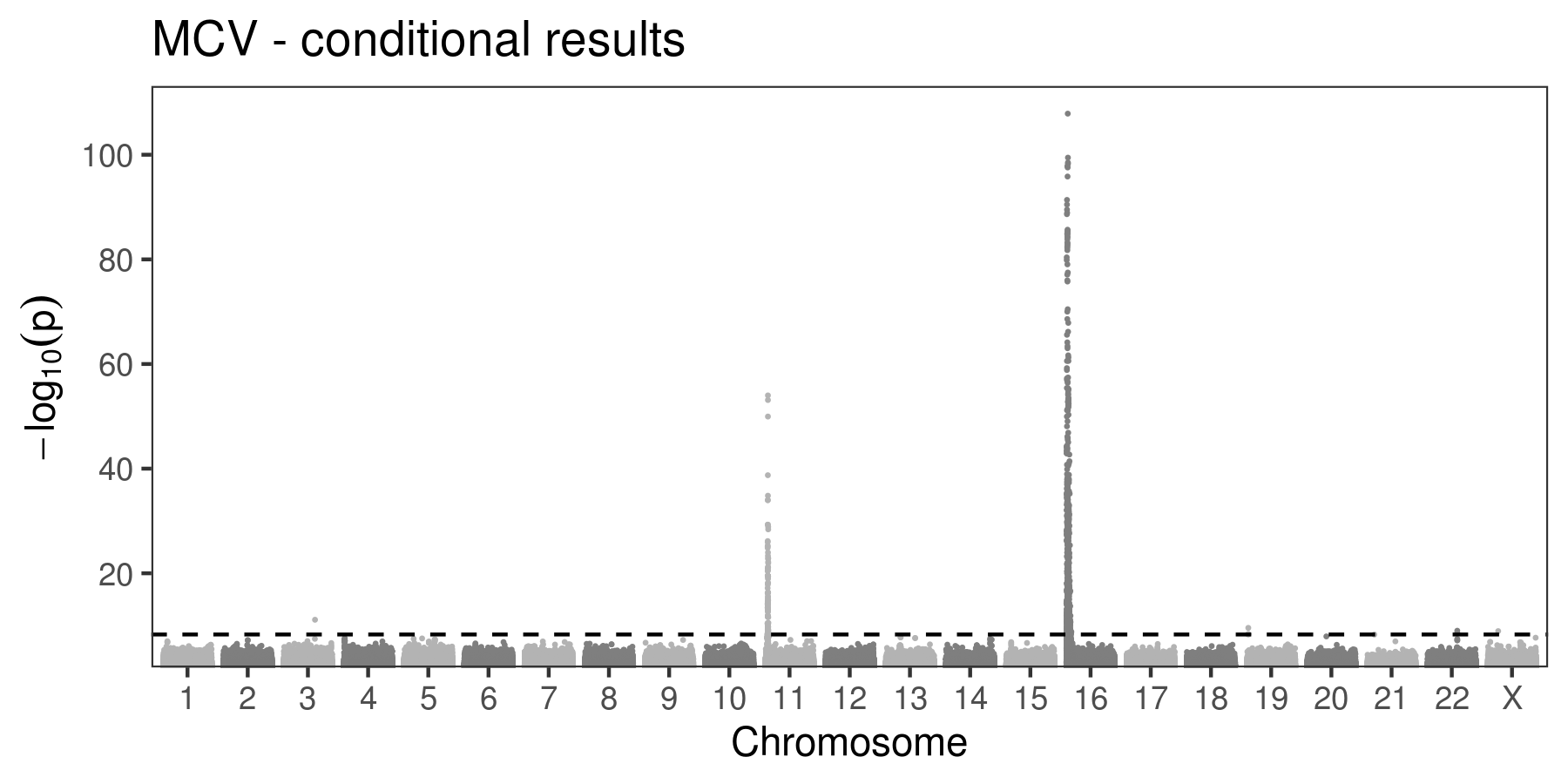

(F)

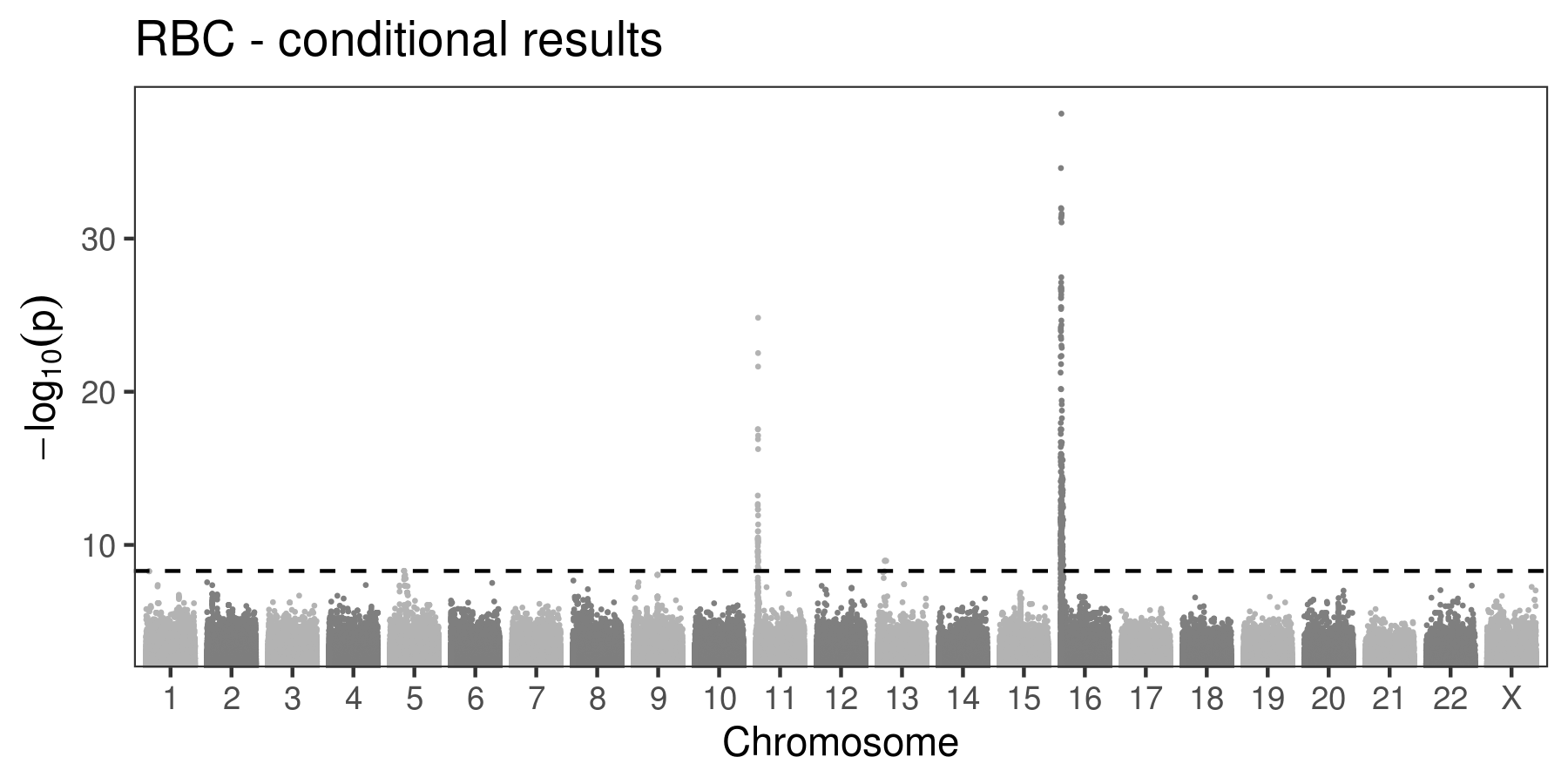

(G)

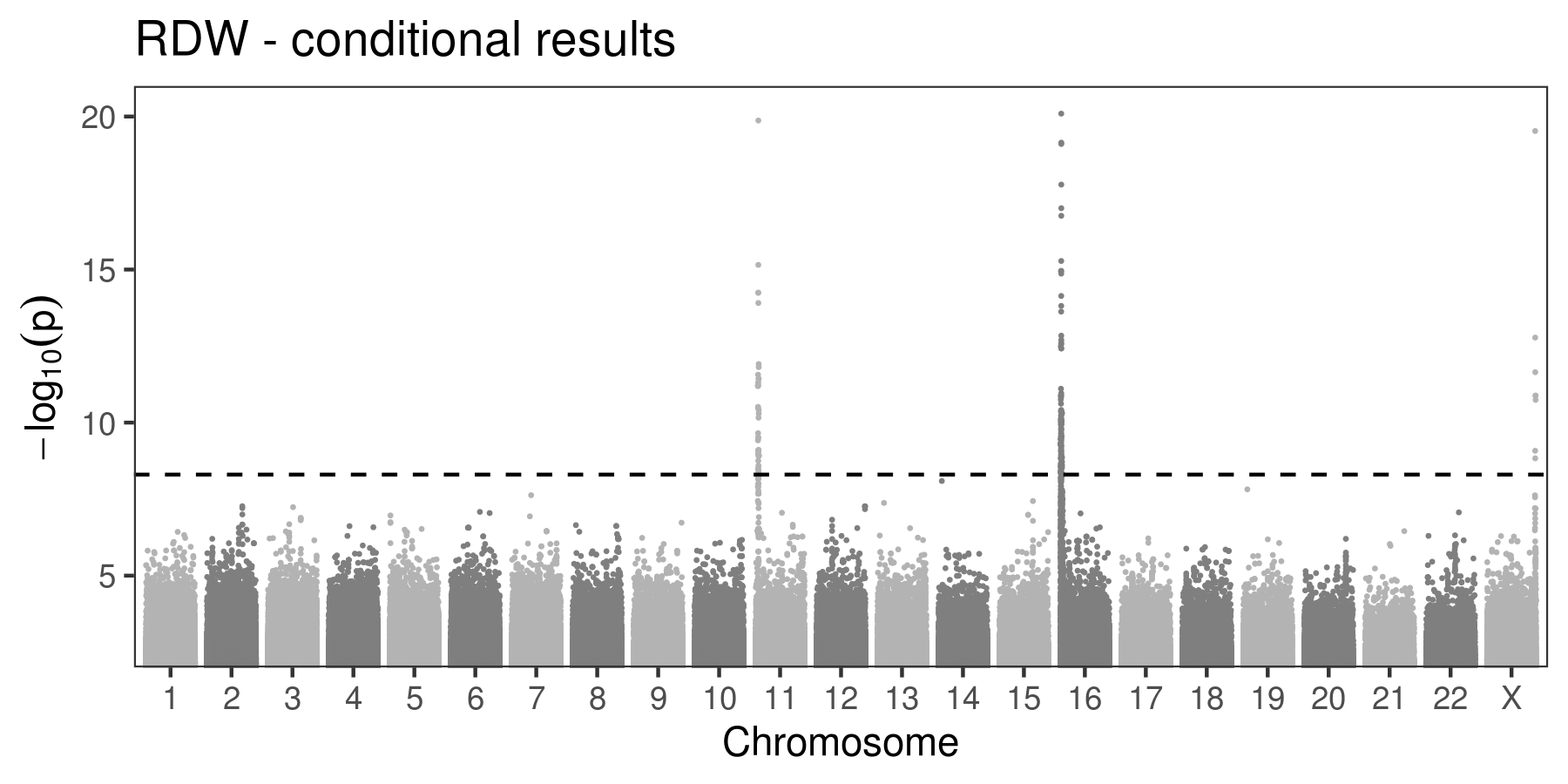

**Figure S4. Locuszoom plots of the 12 novel variants and conditionally independent variants identified in TOPMed. (A) HCT; (B) HGB; (C) MCH; (D) MCHC; (E) MCV; (F) RBC; (G) RDW**

(A)

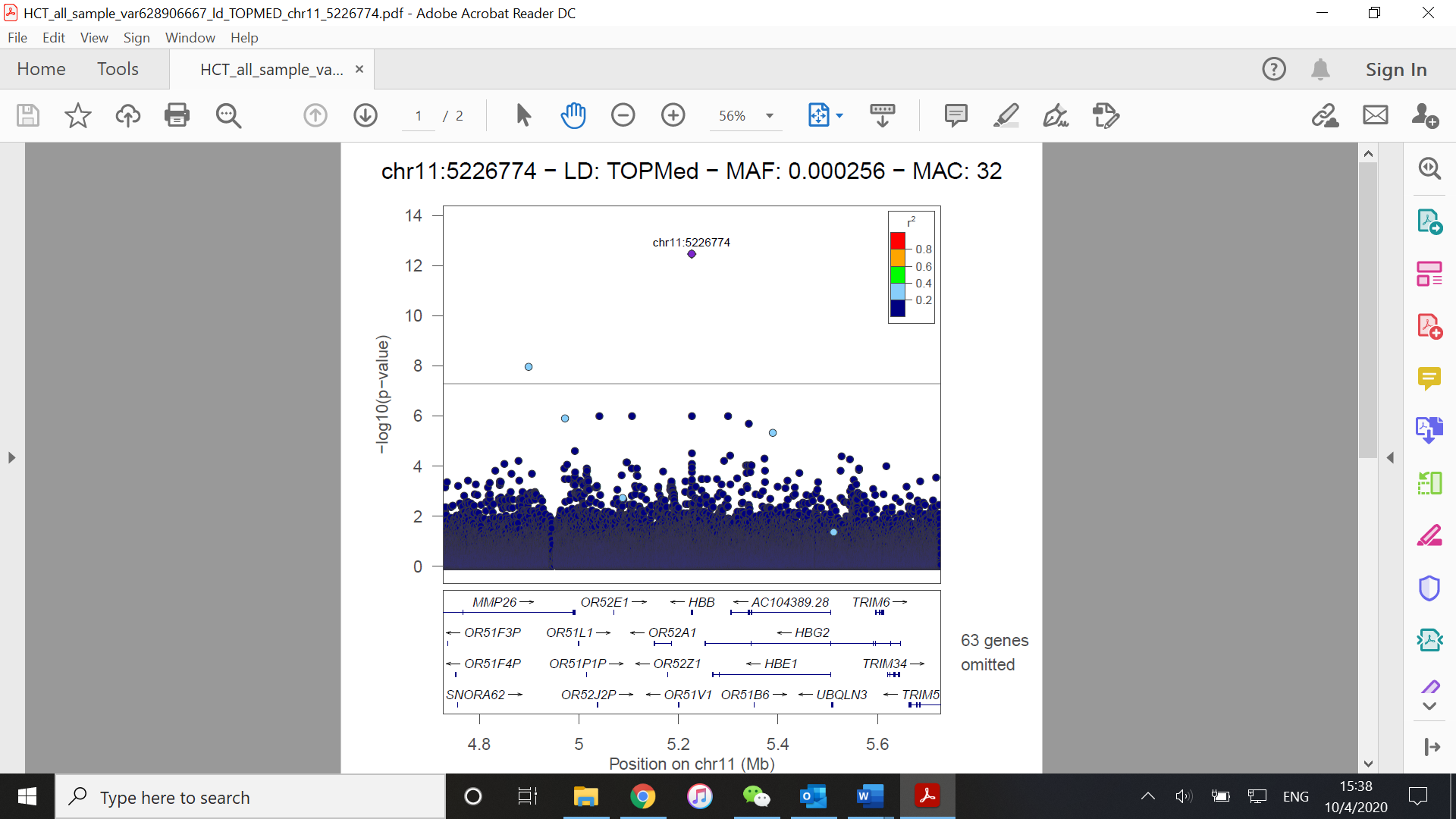

(B)

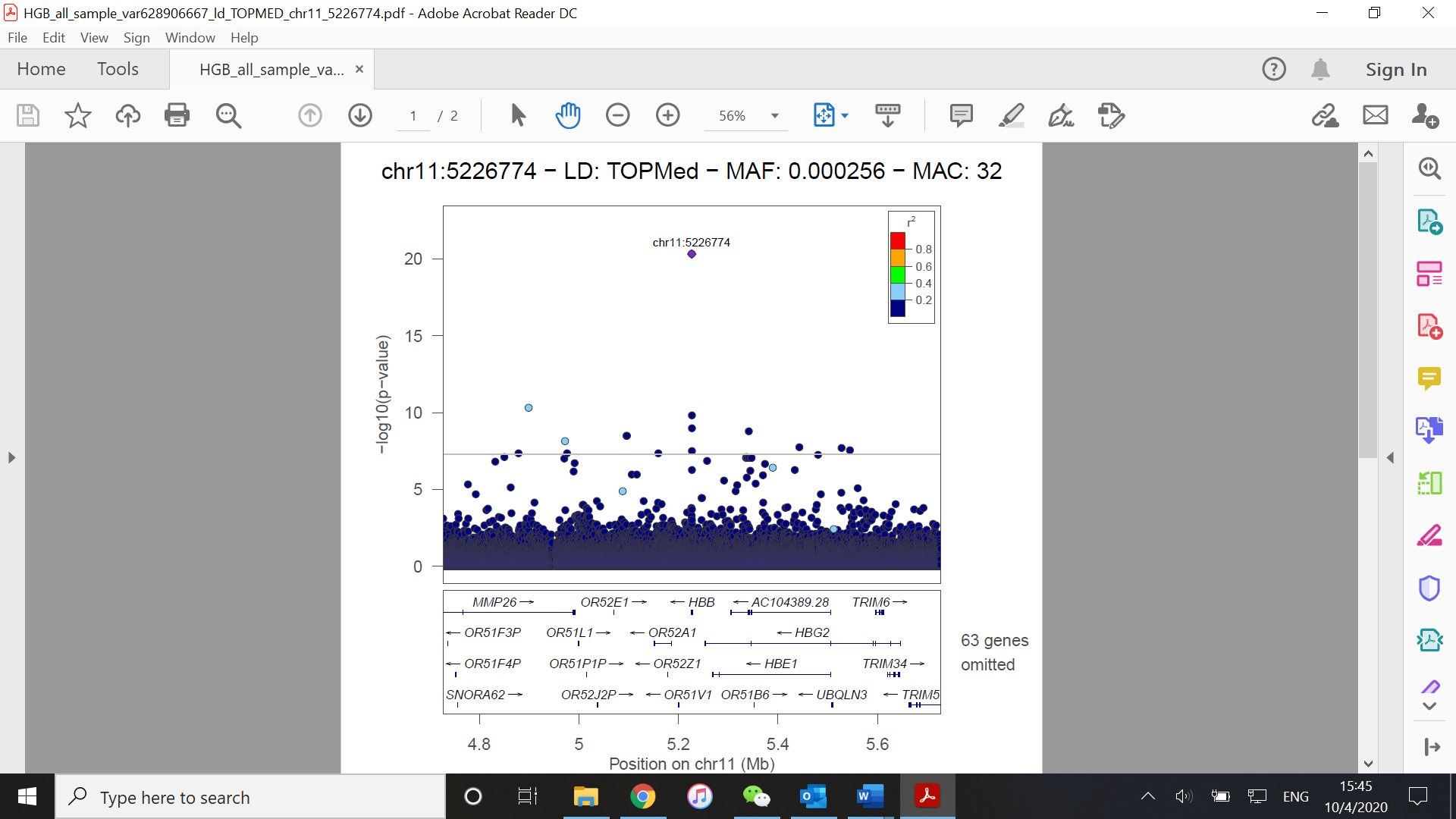

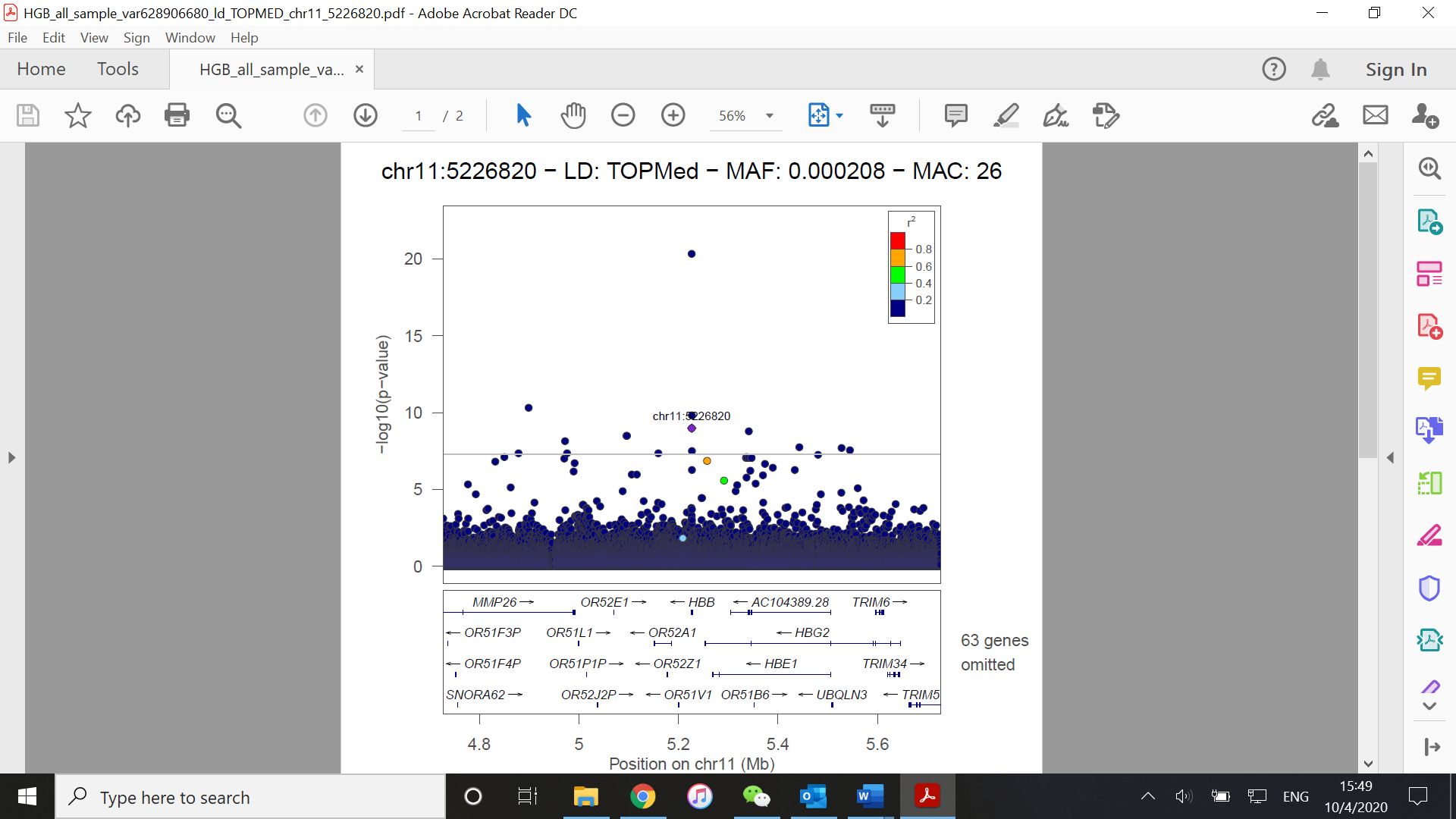

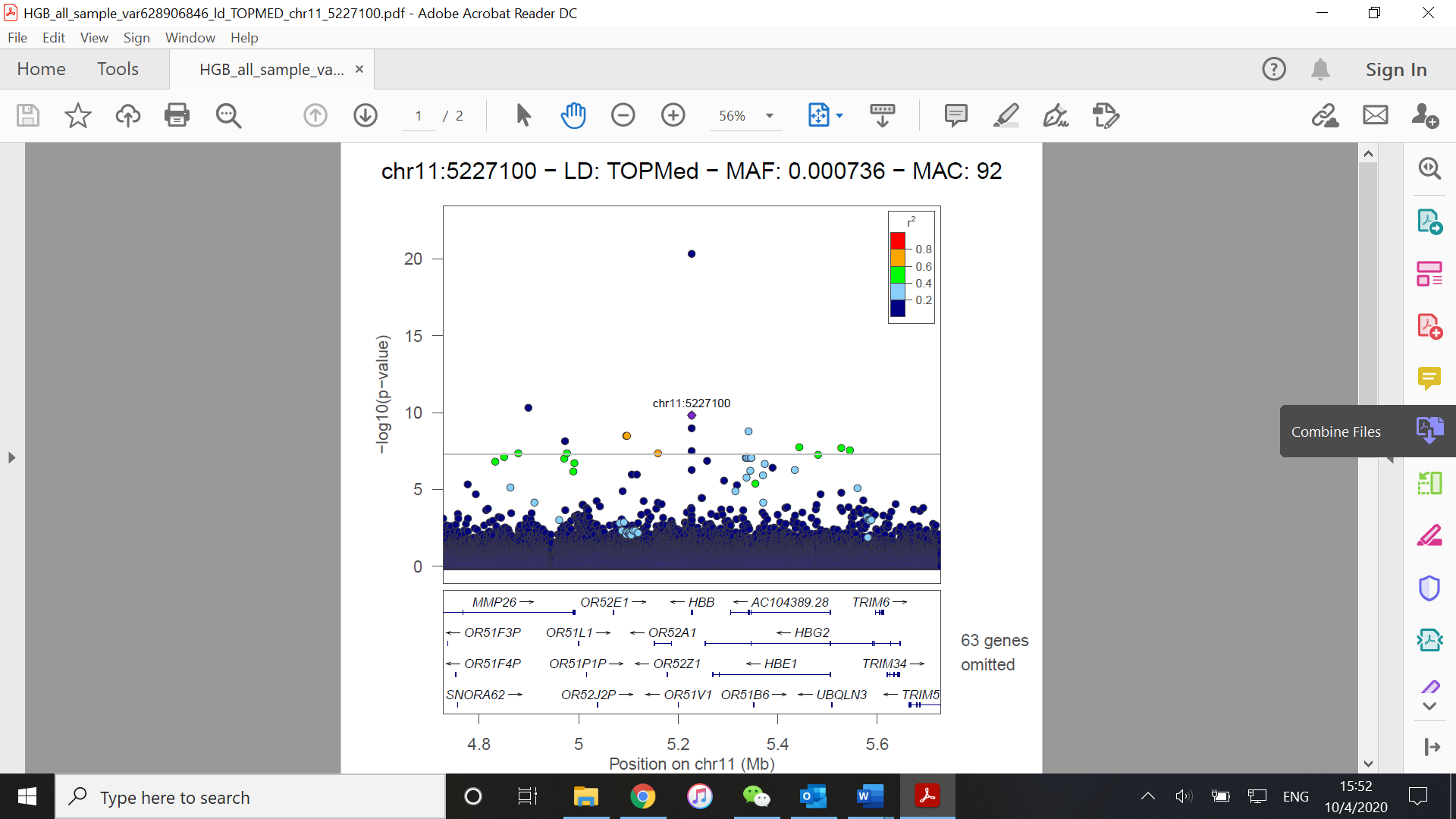

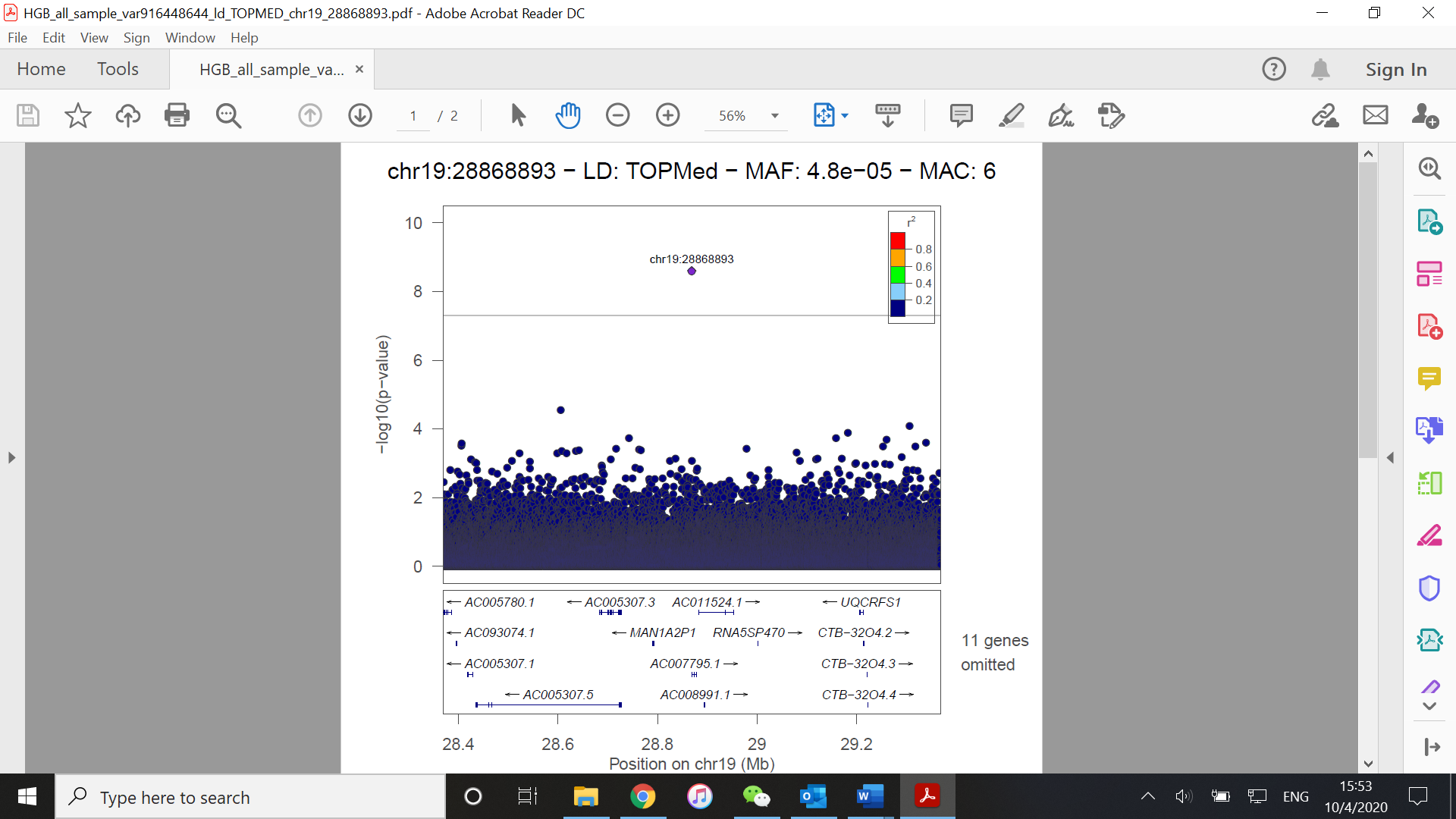

(C)

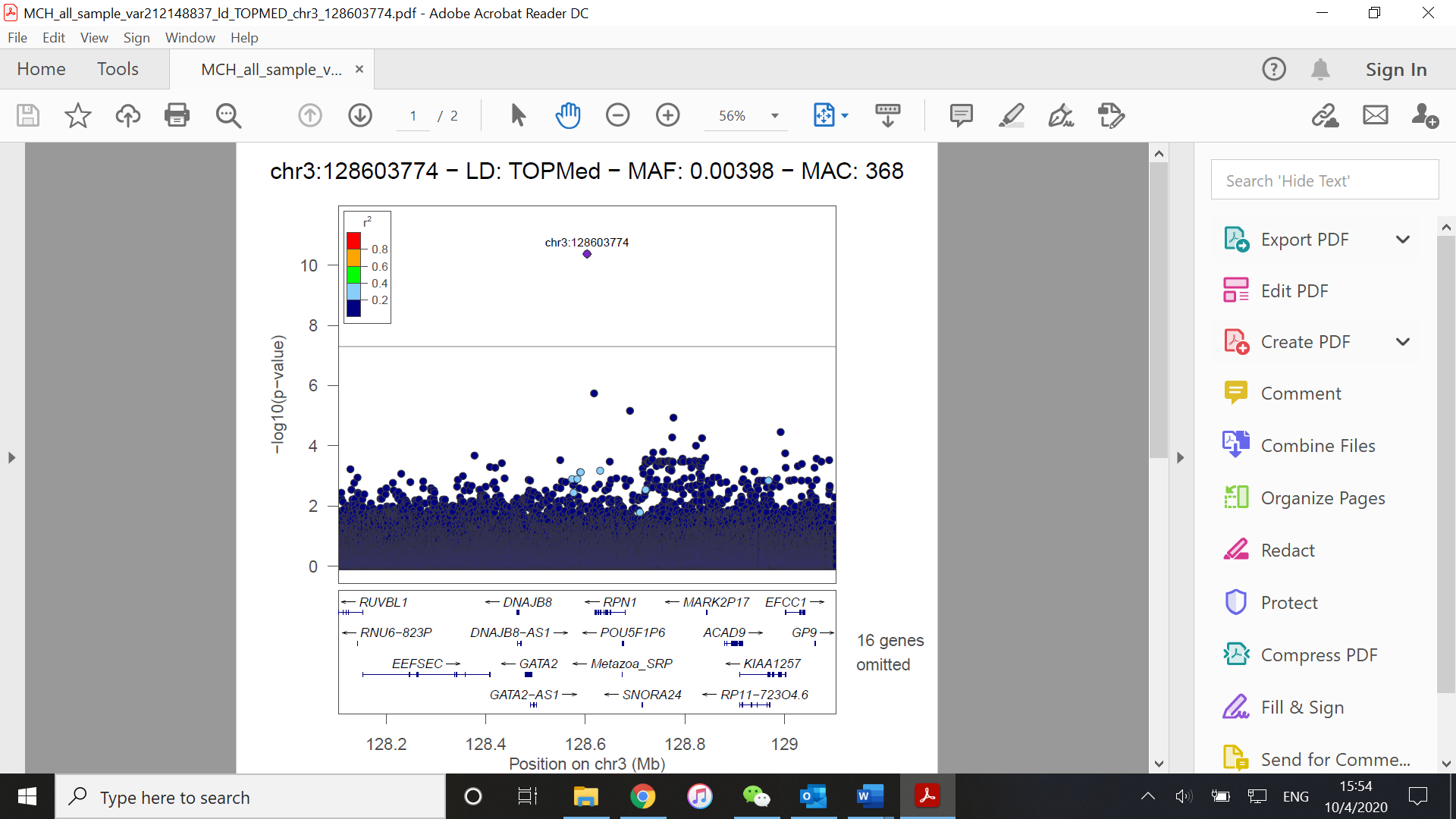

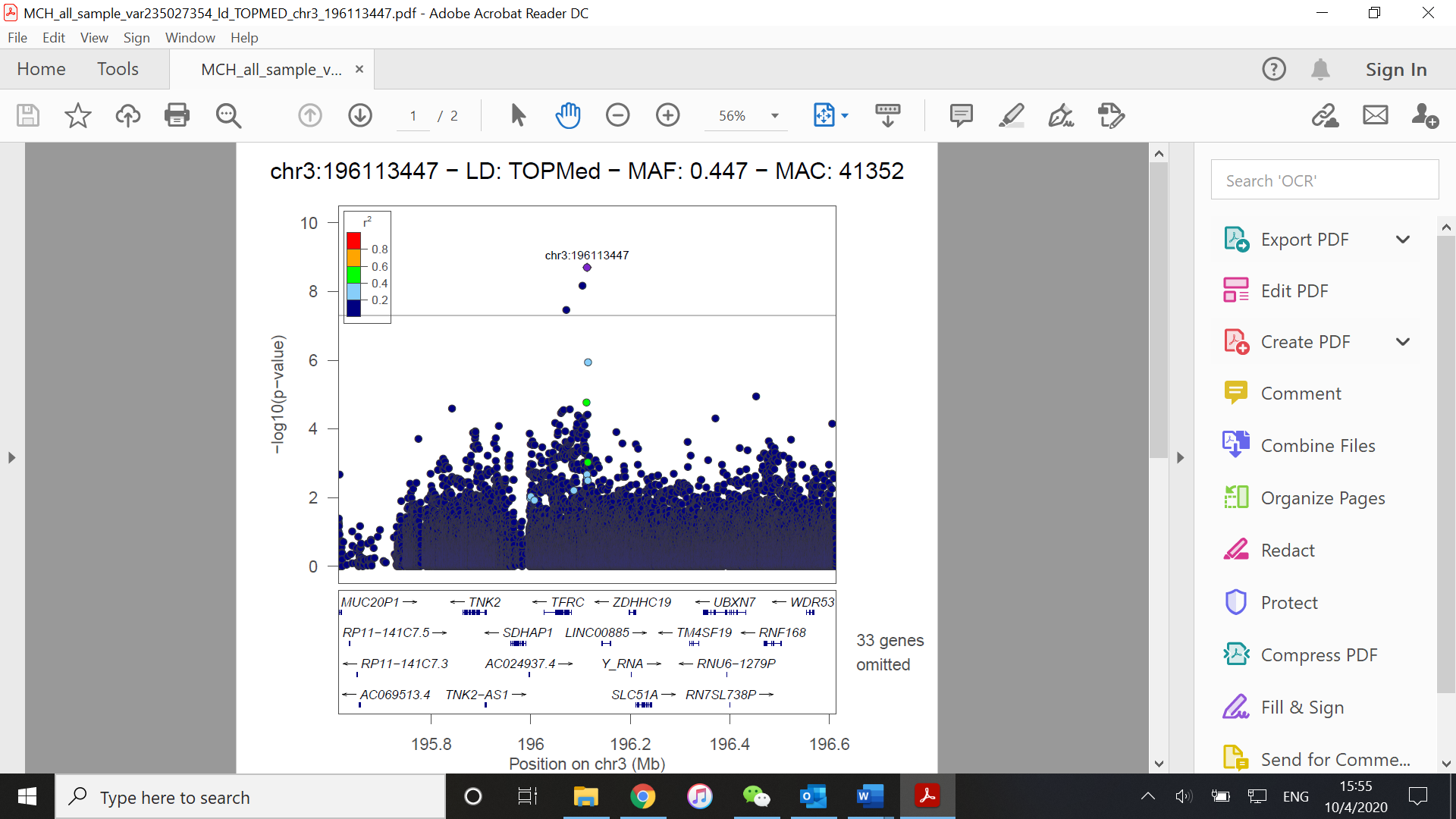

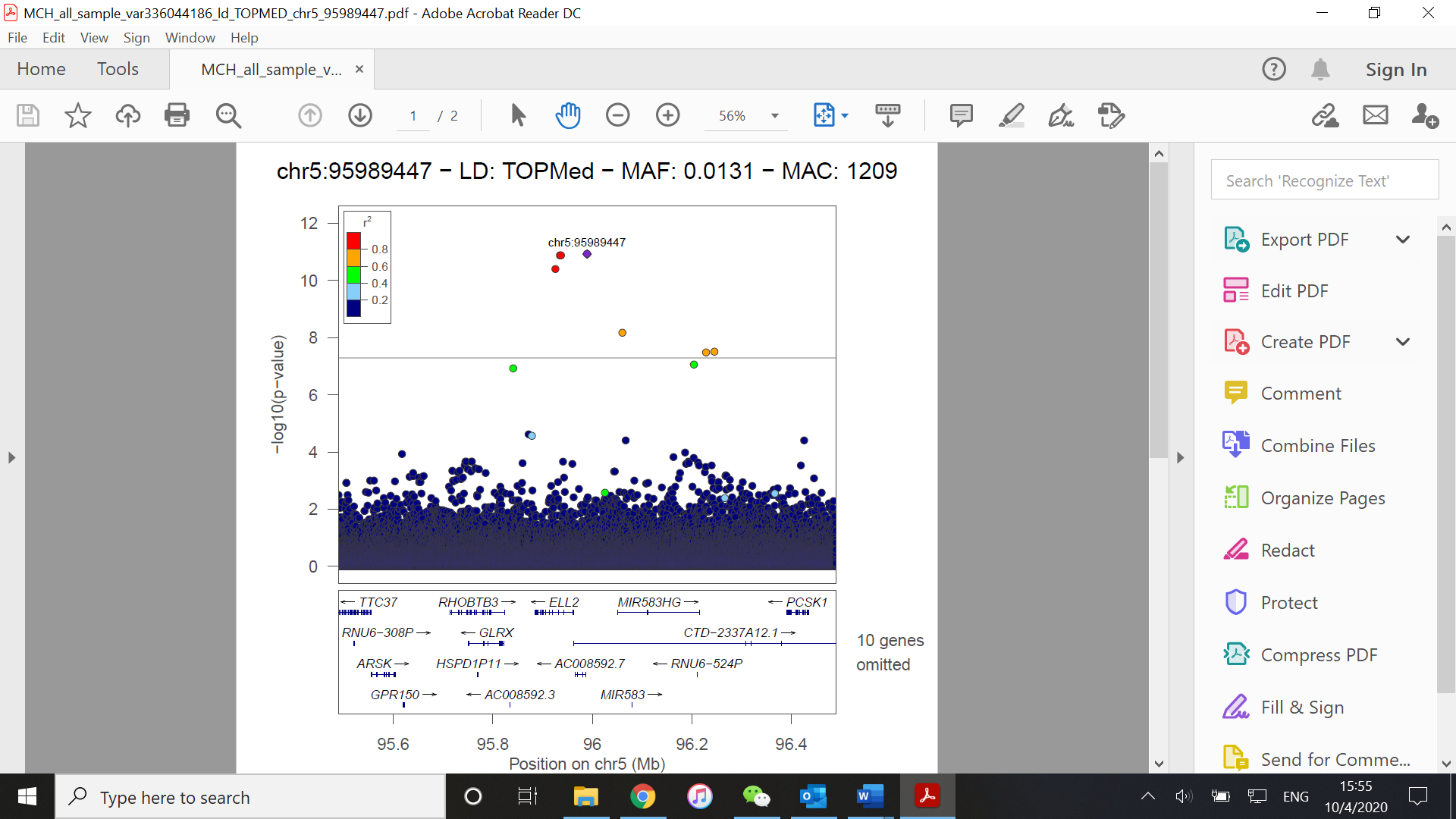

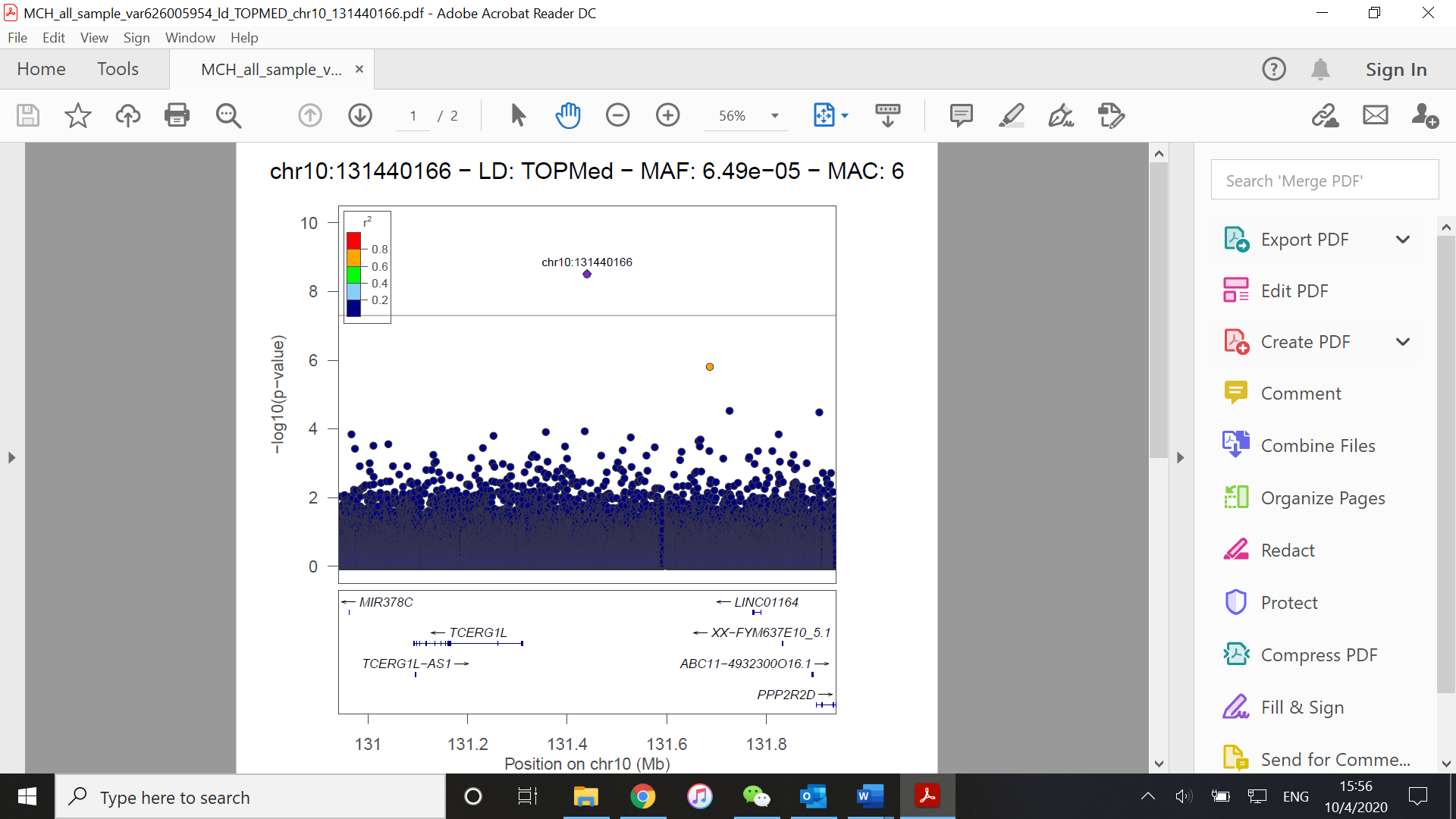

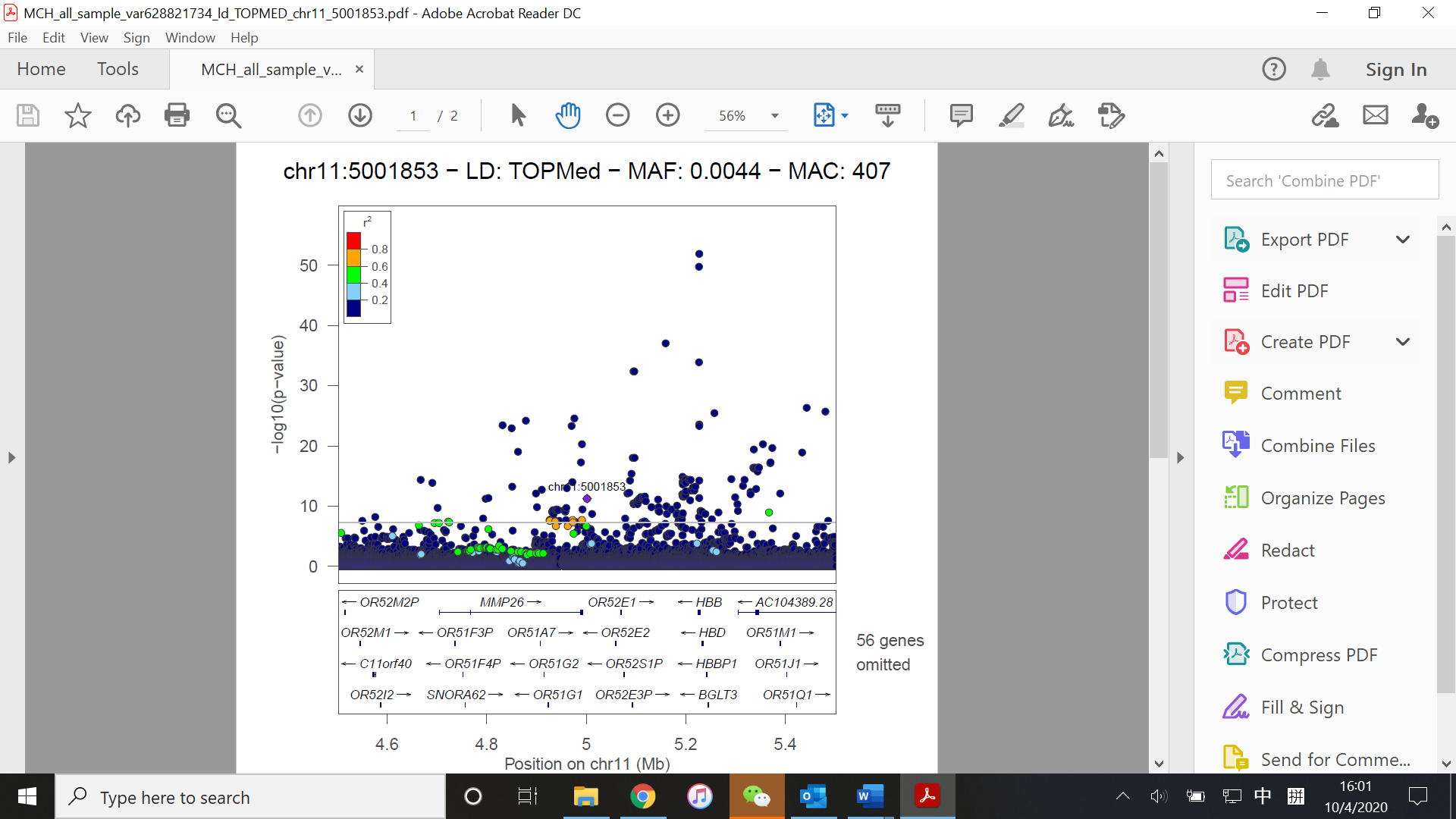

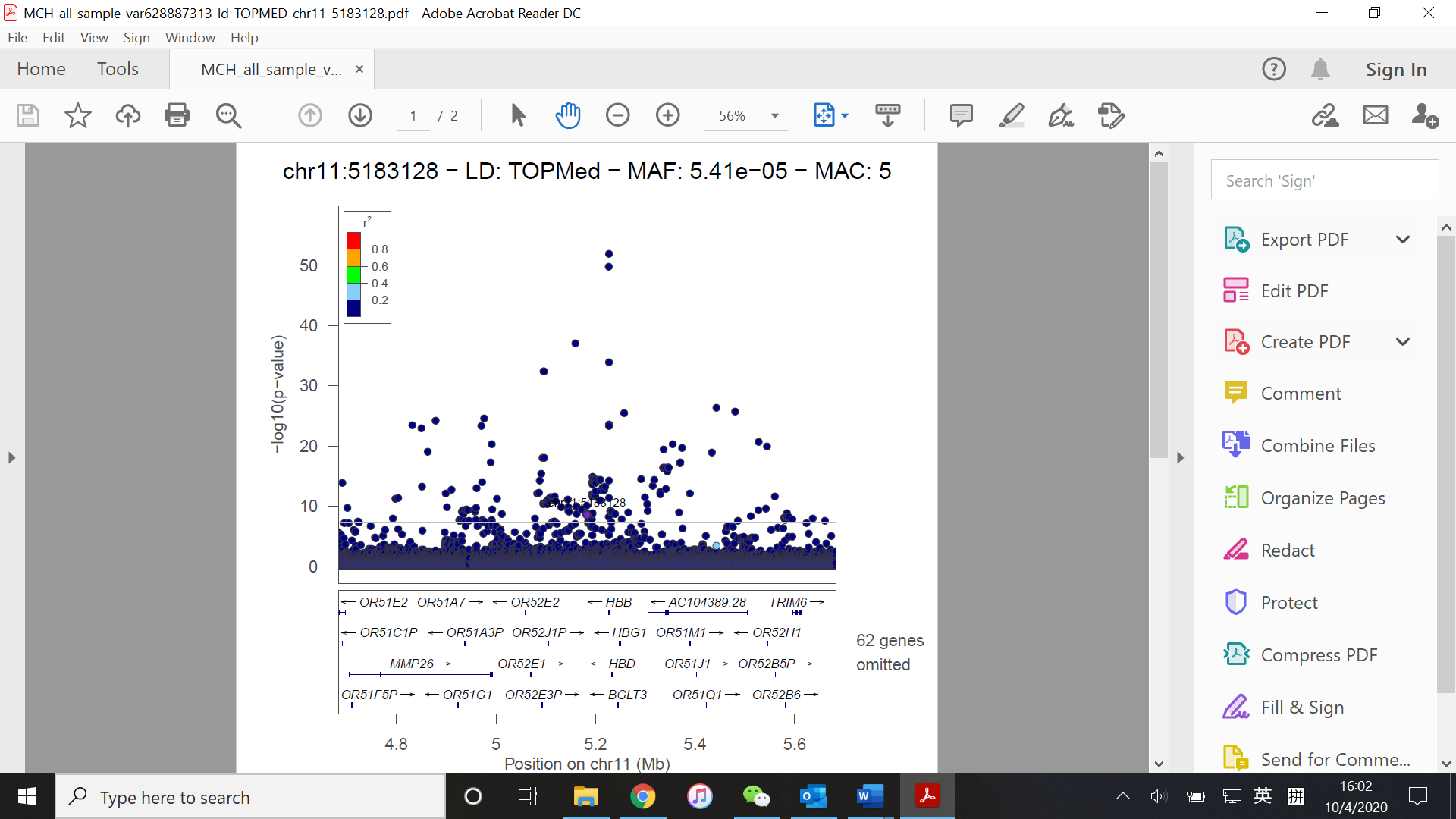

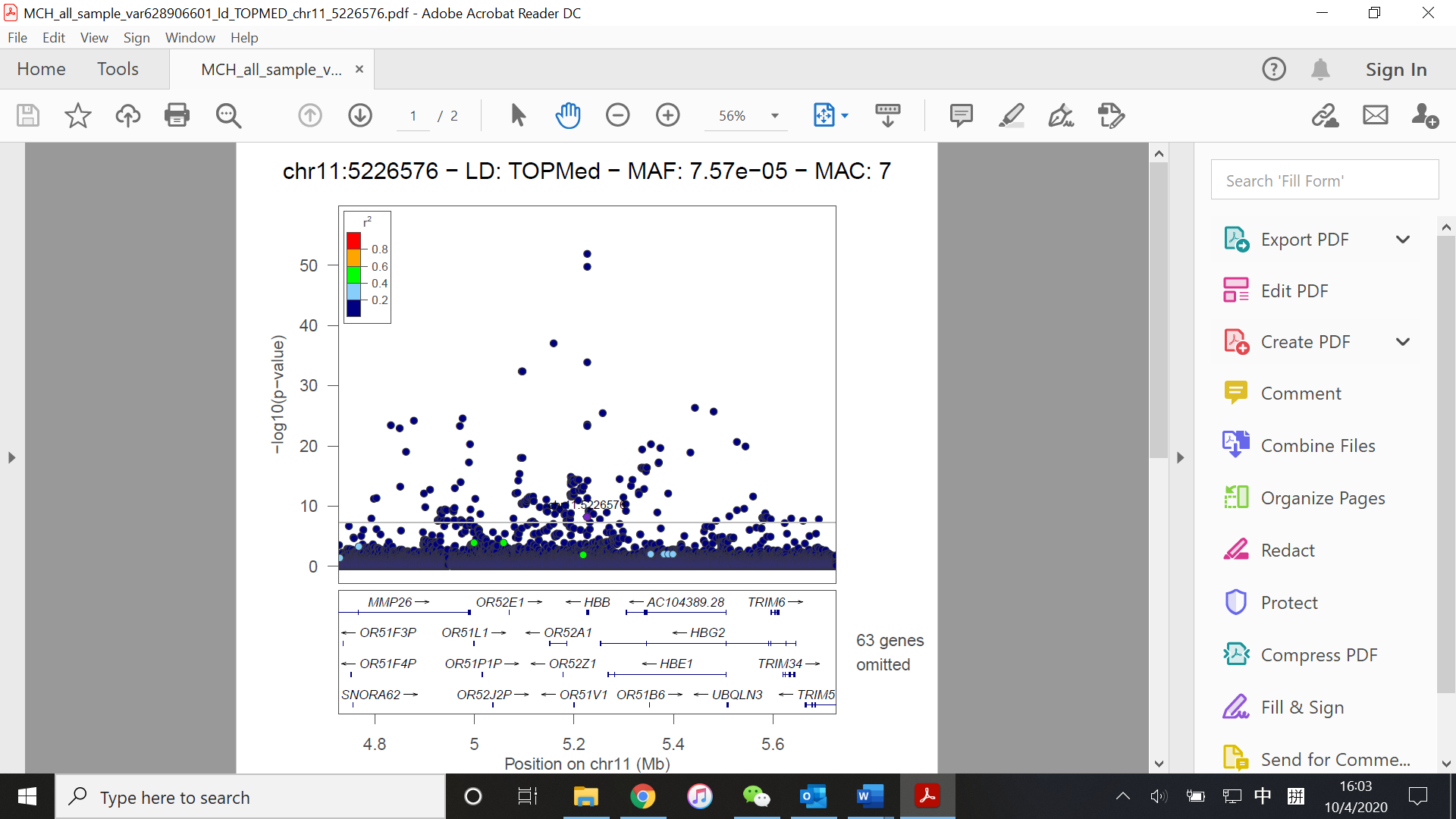

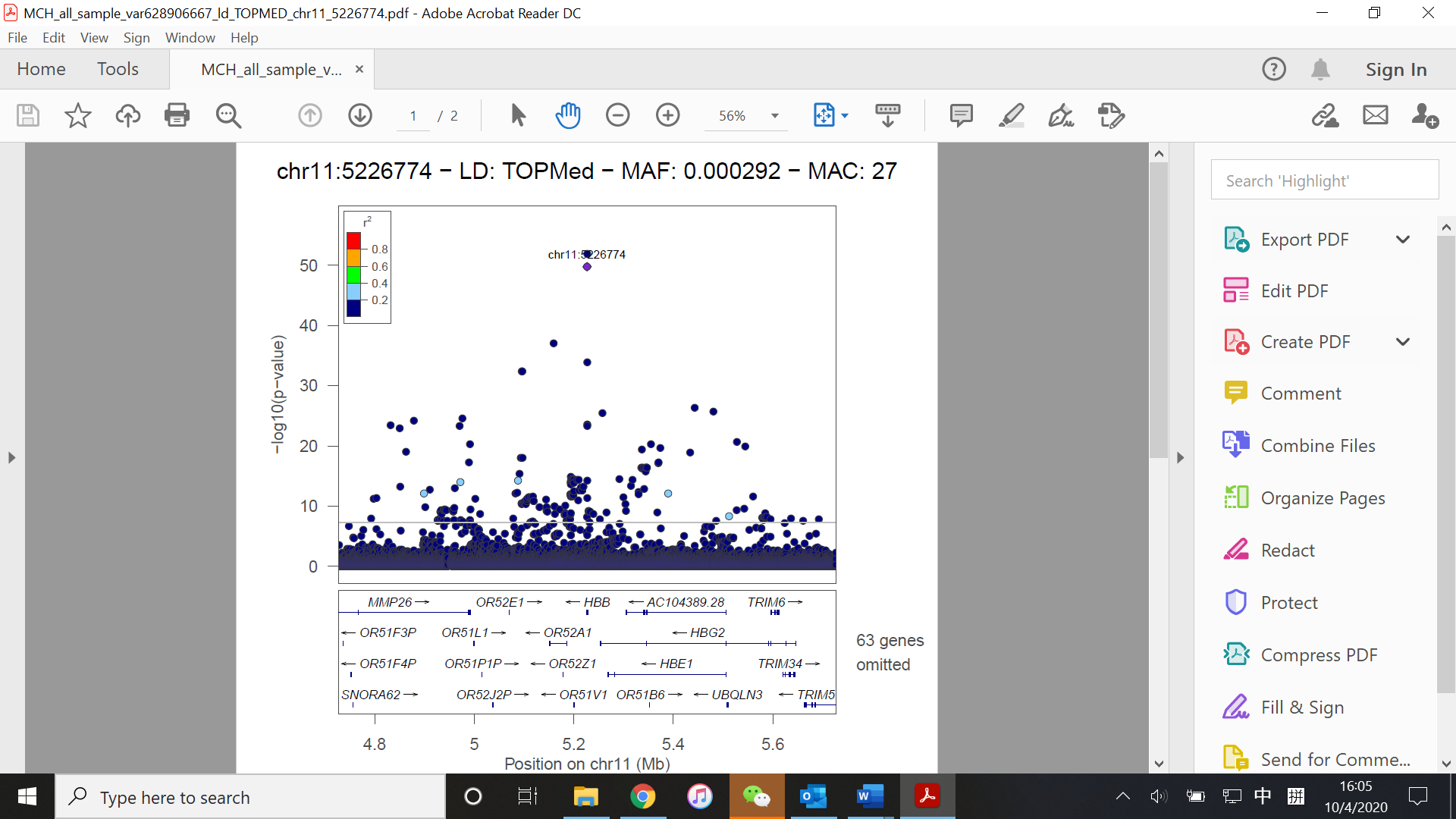

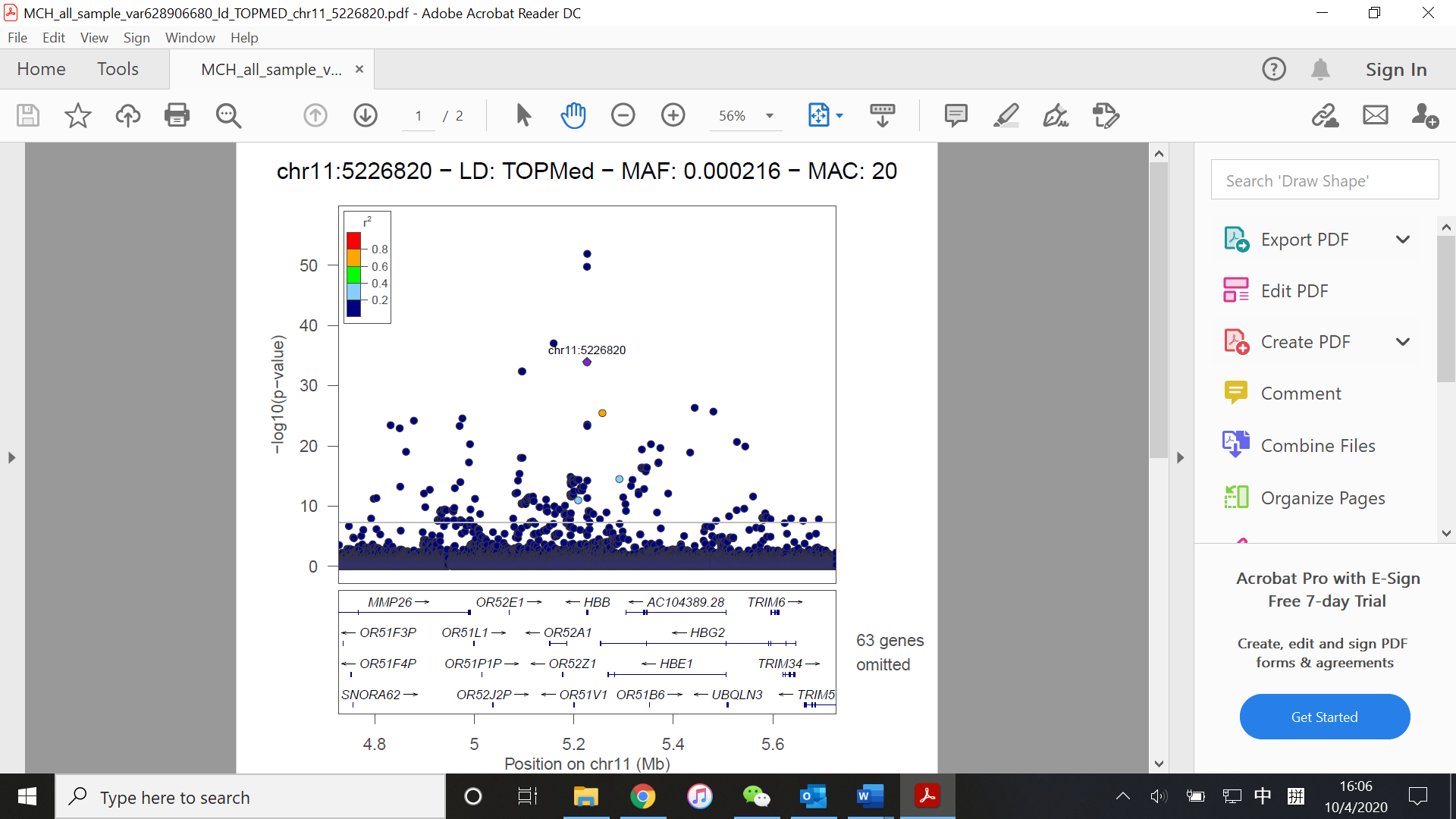

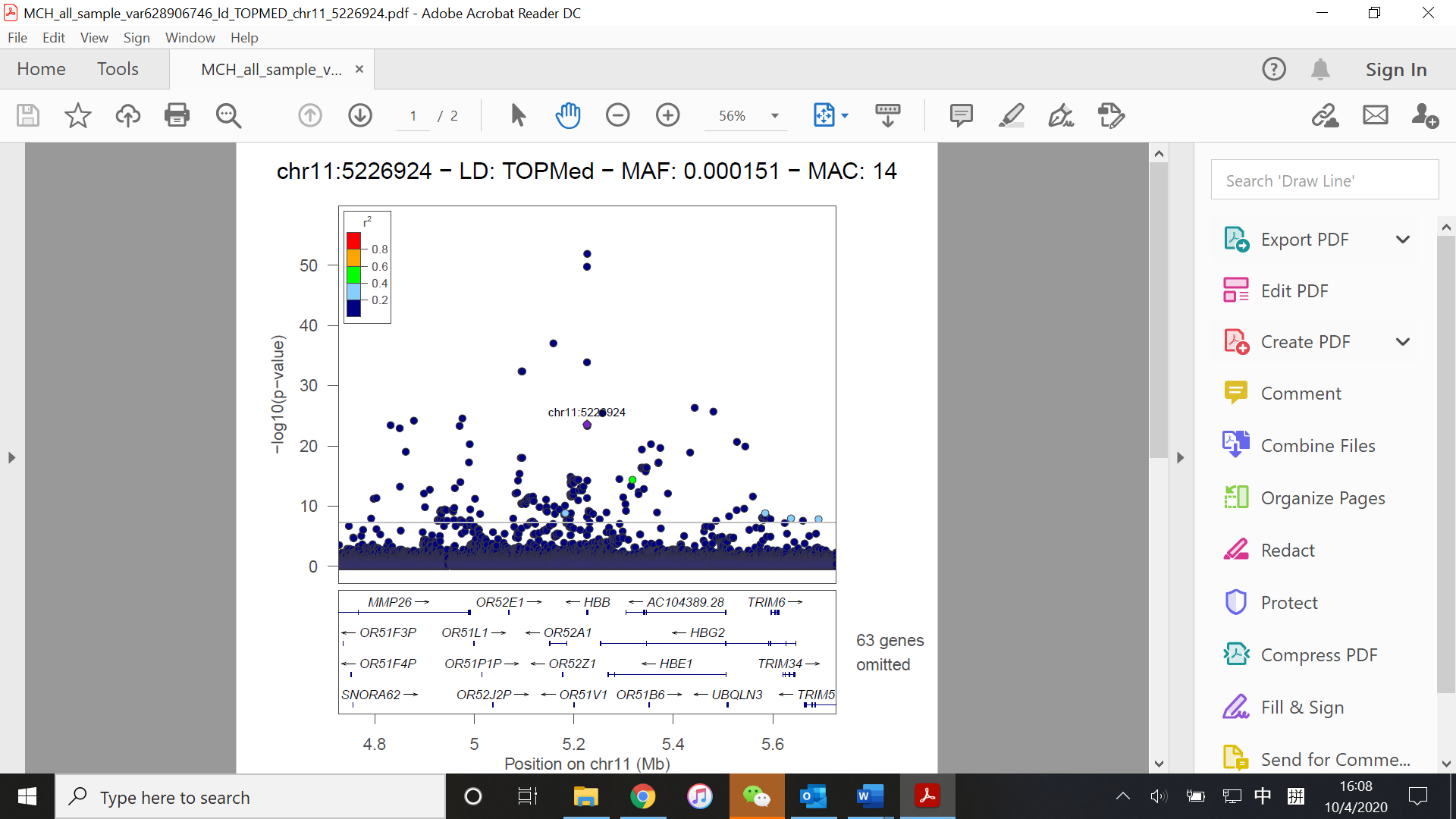

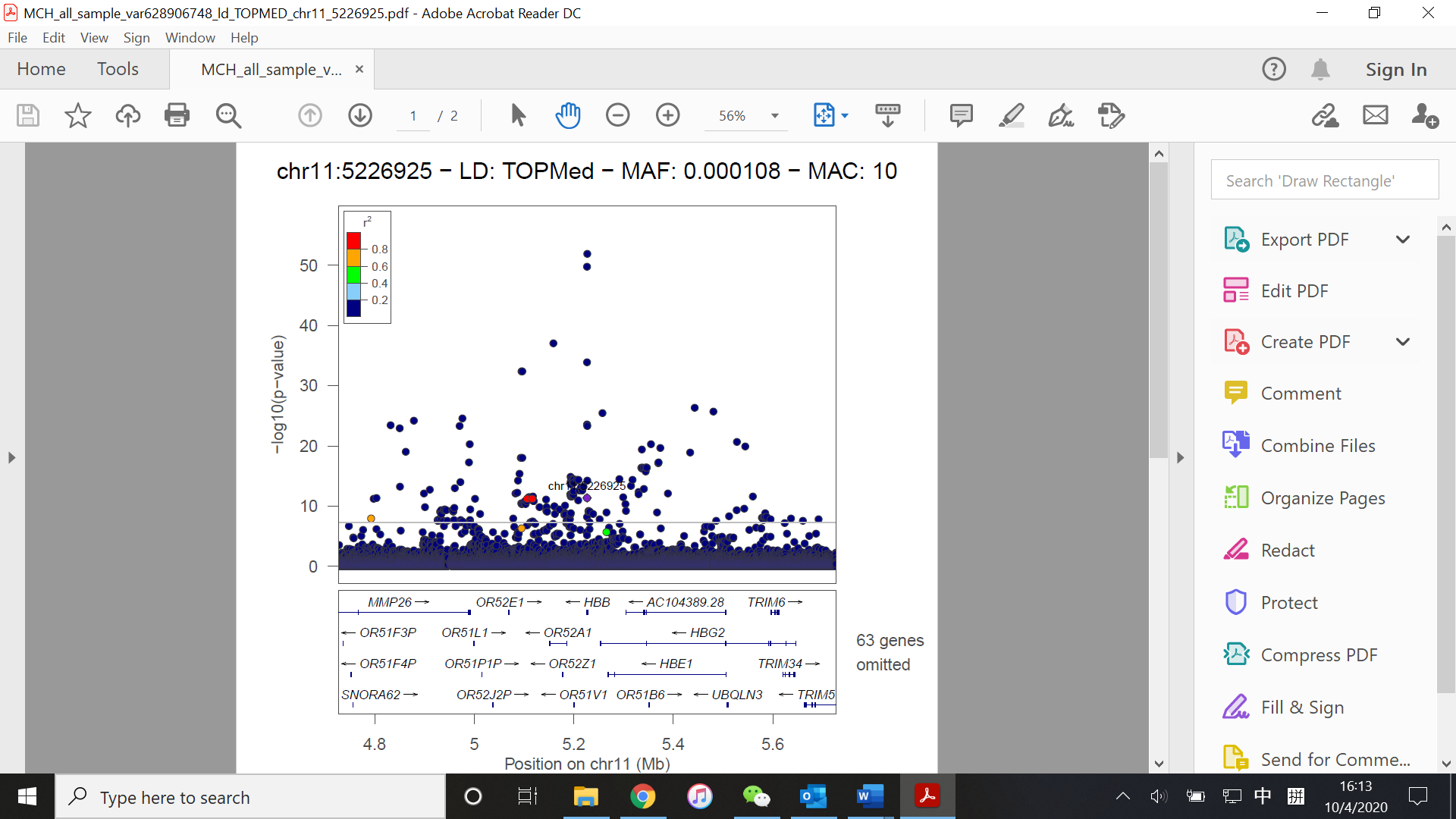

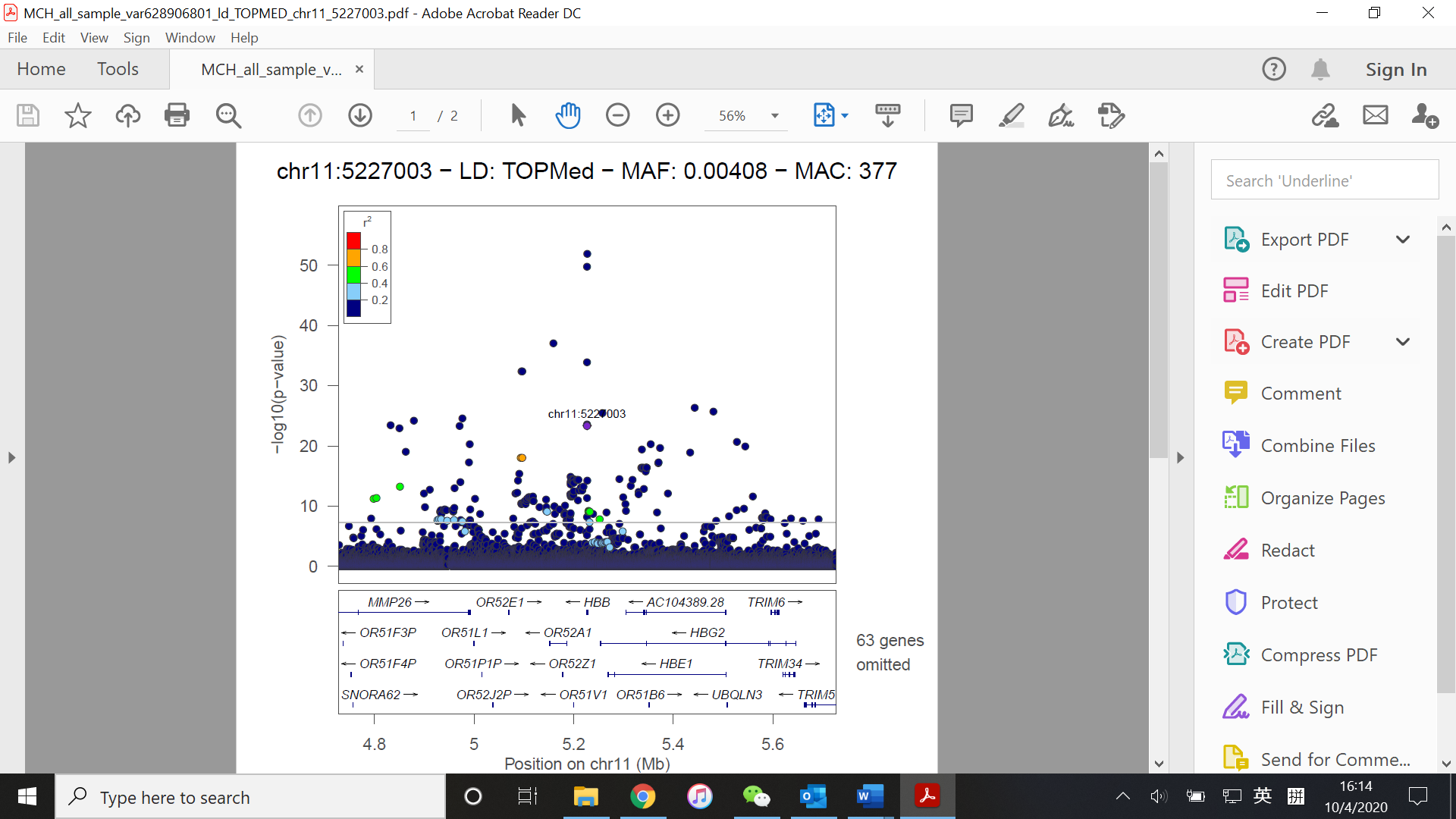

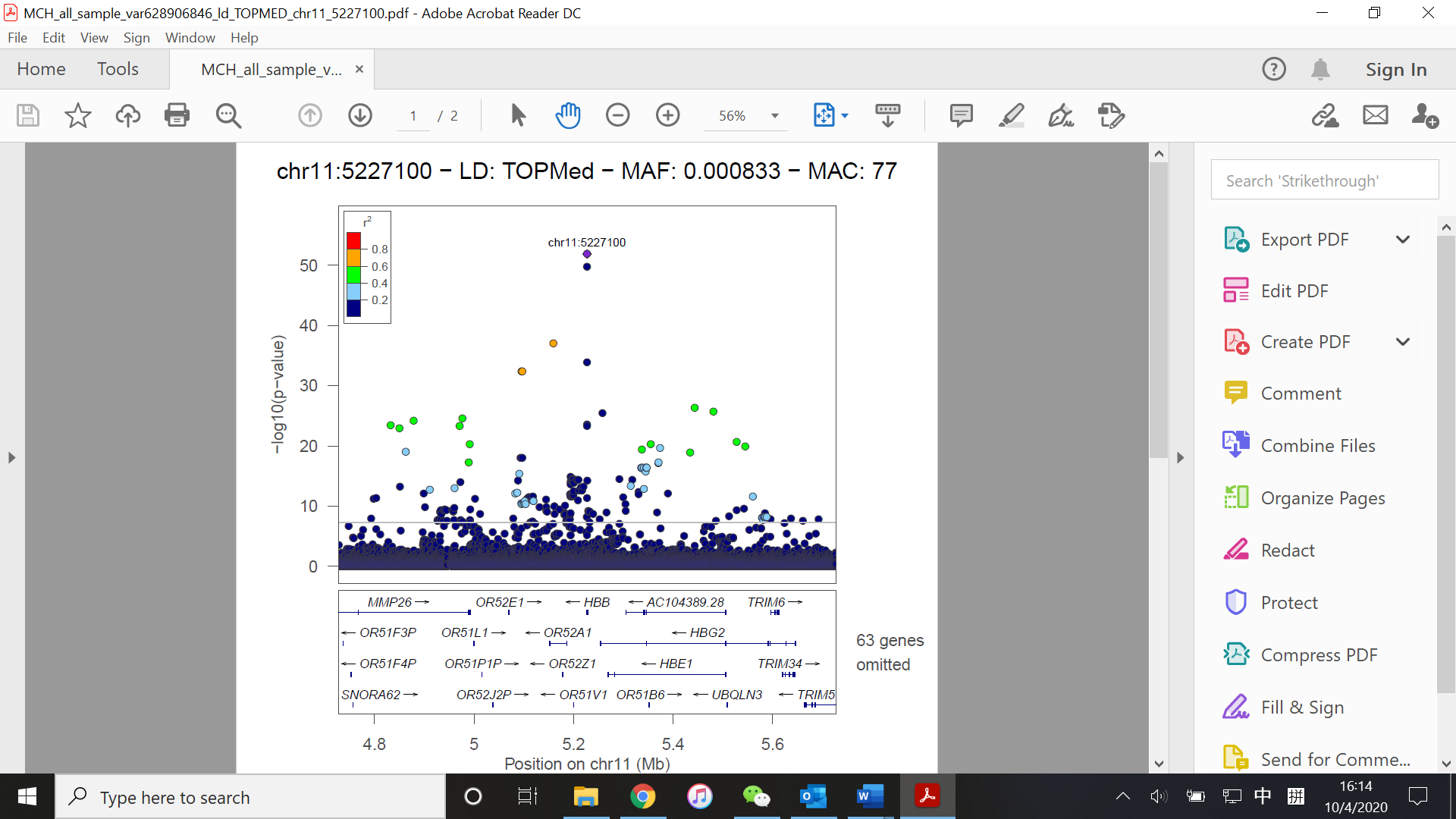

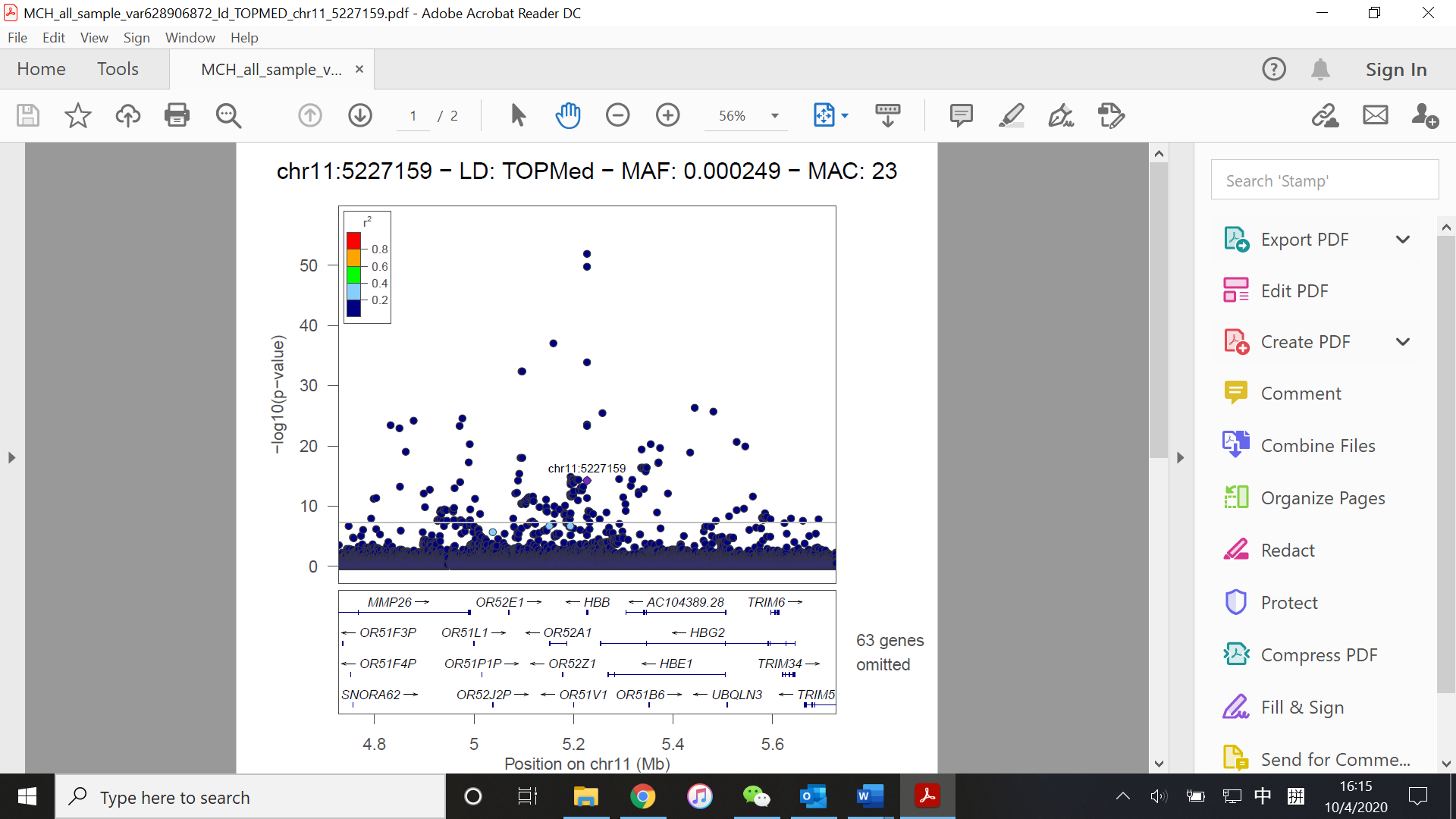

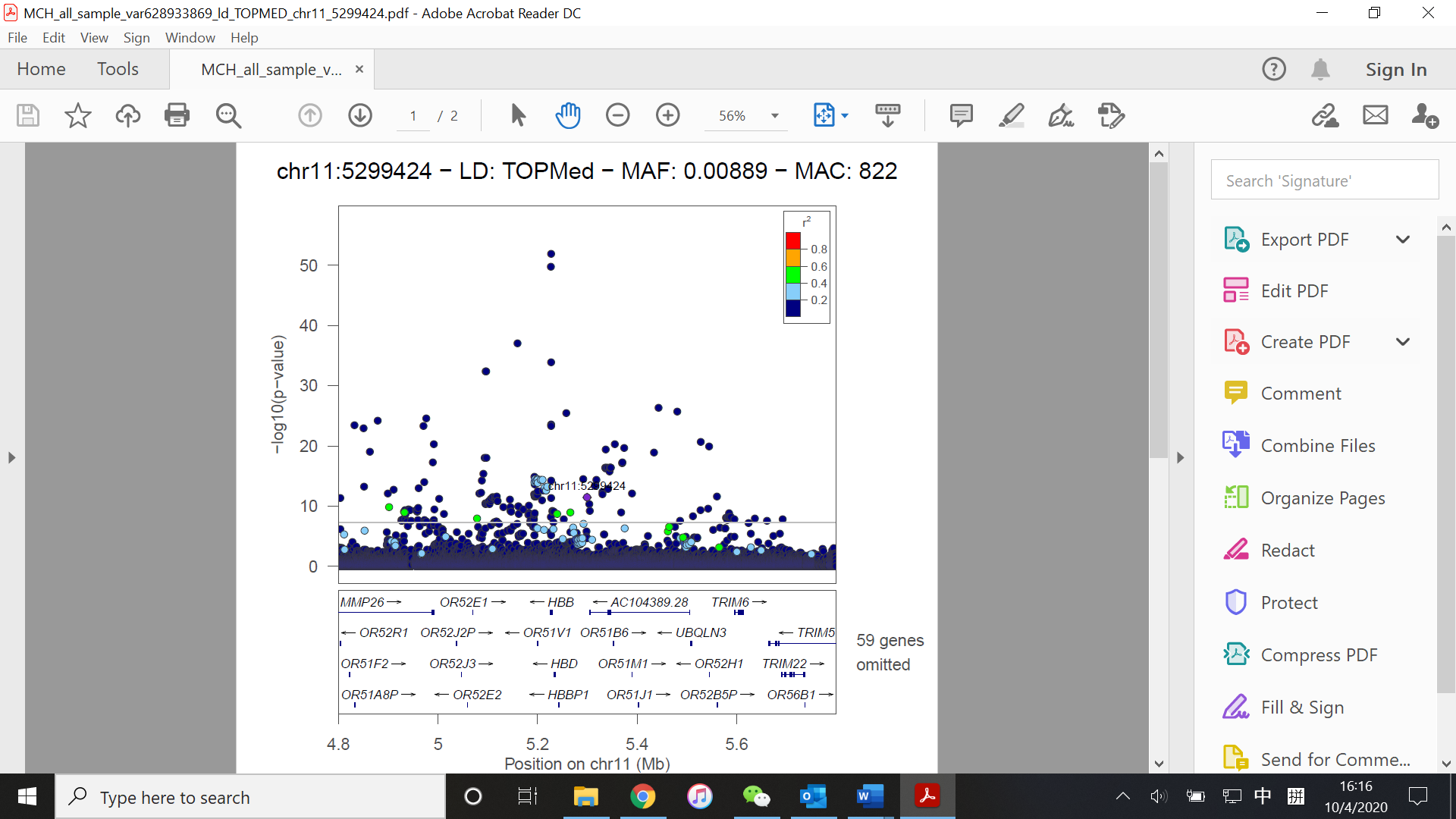

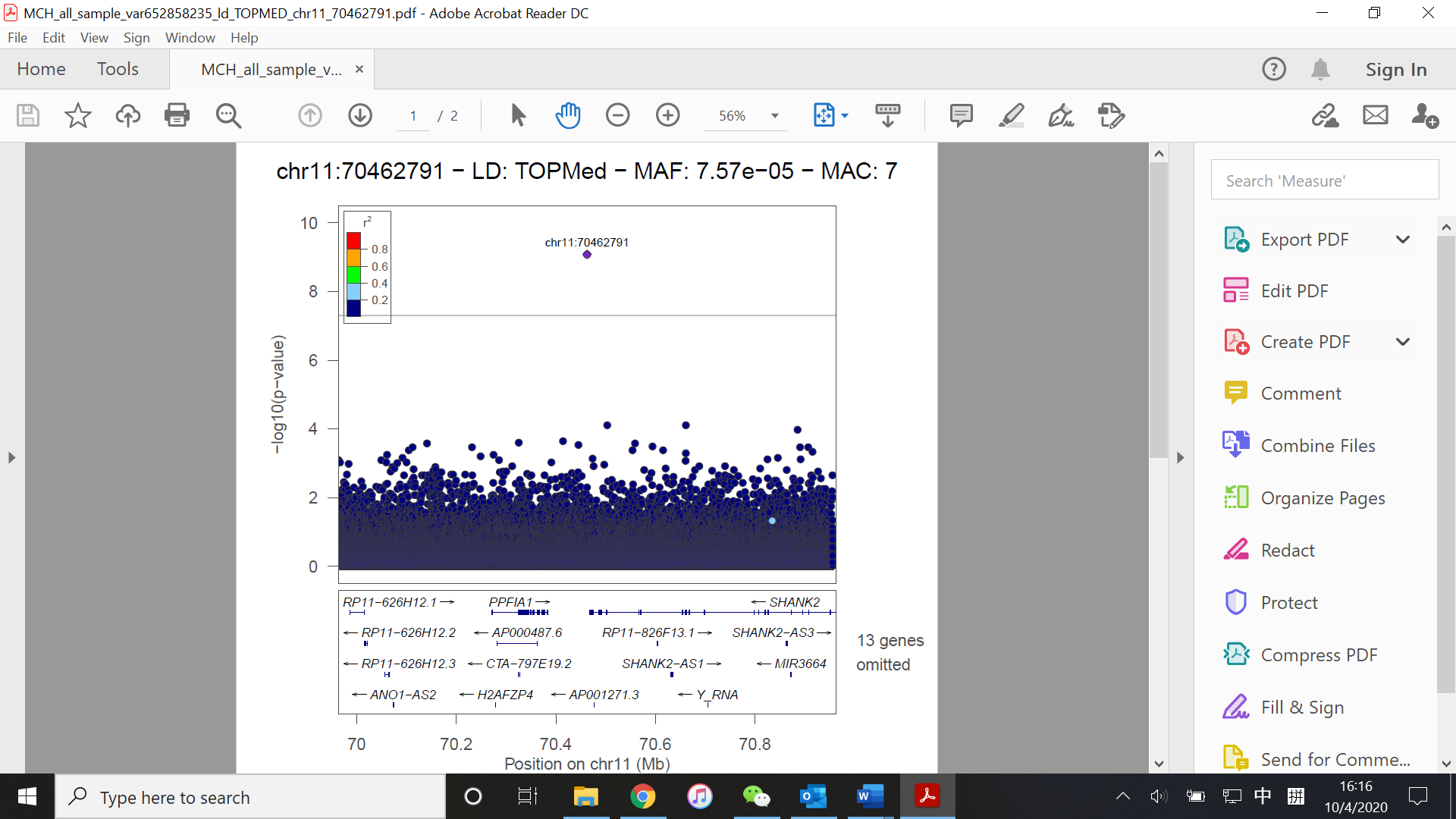

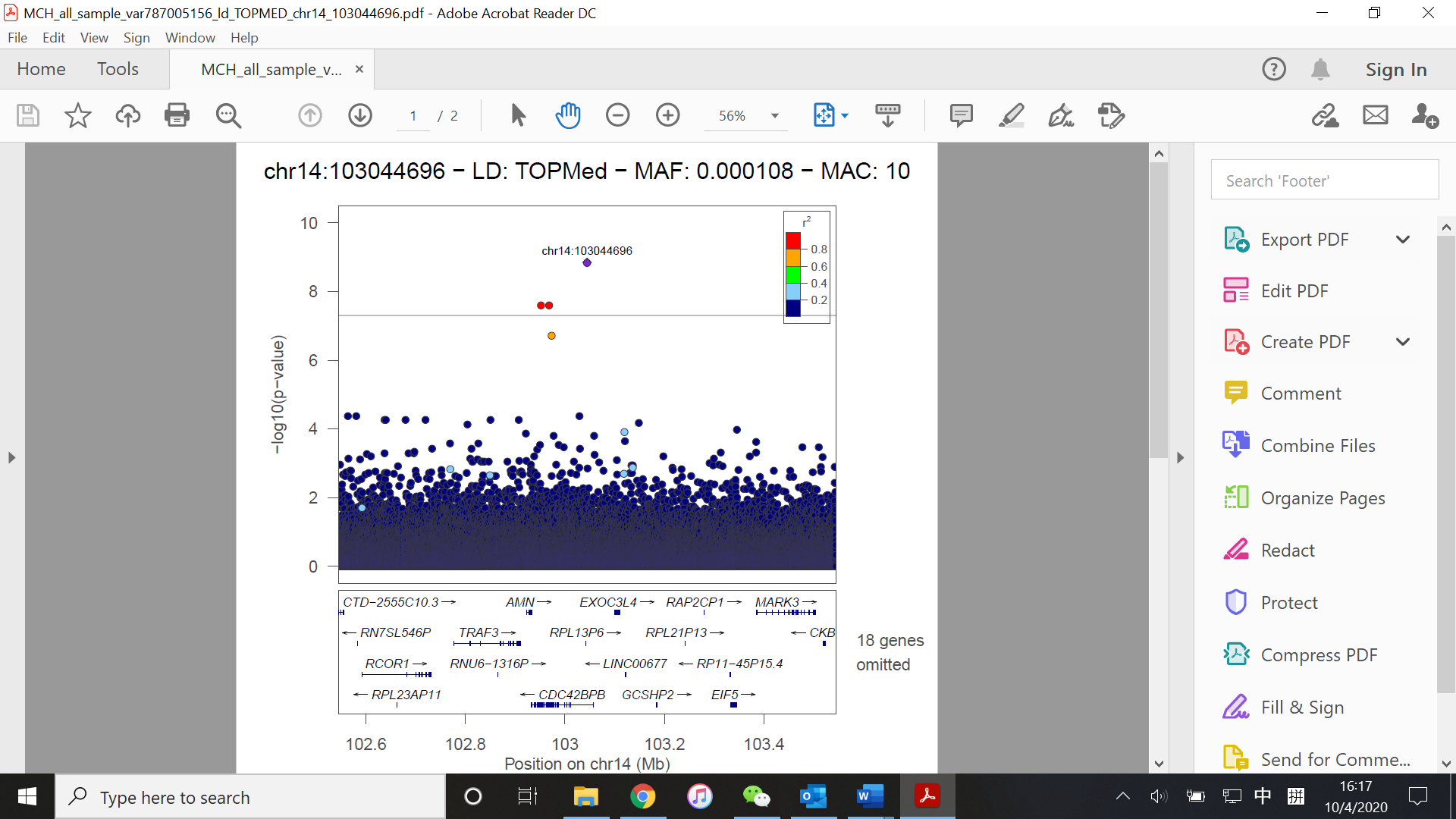

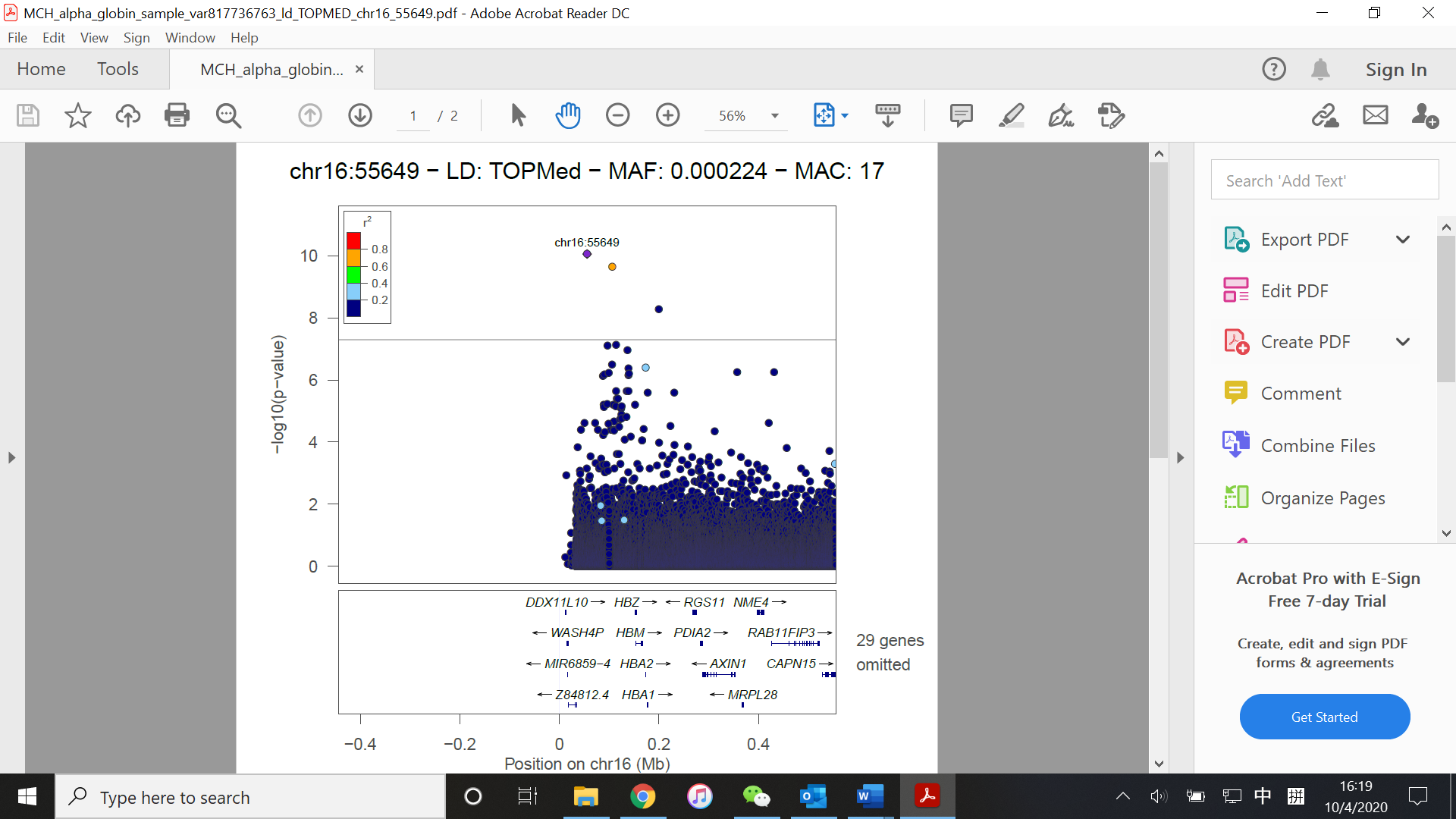

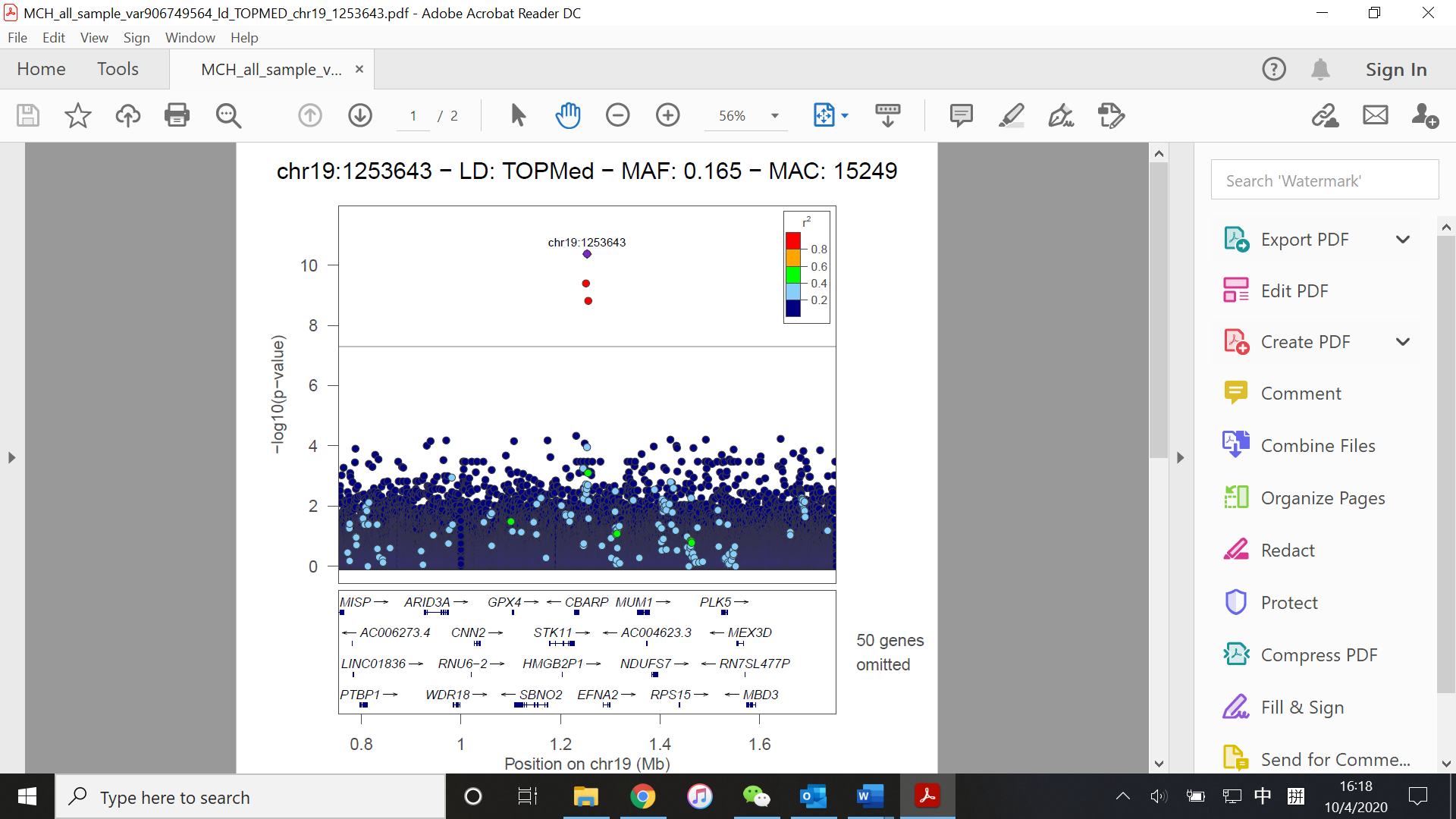

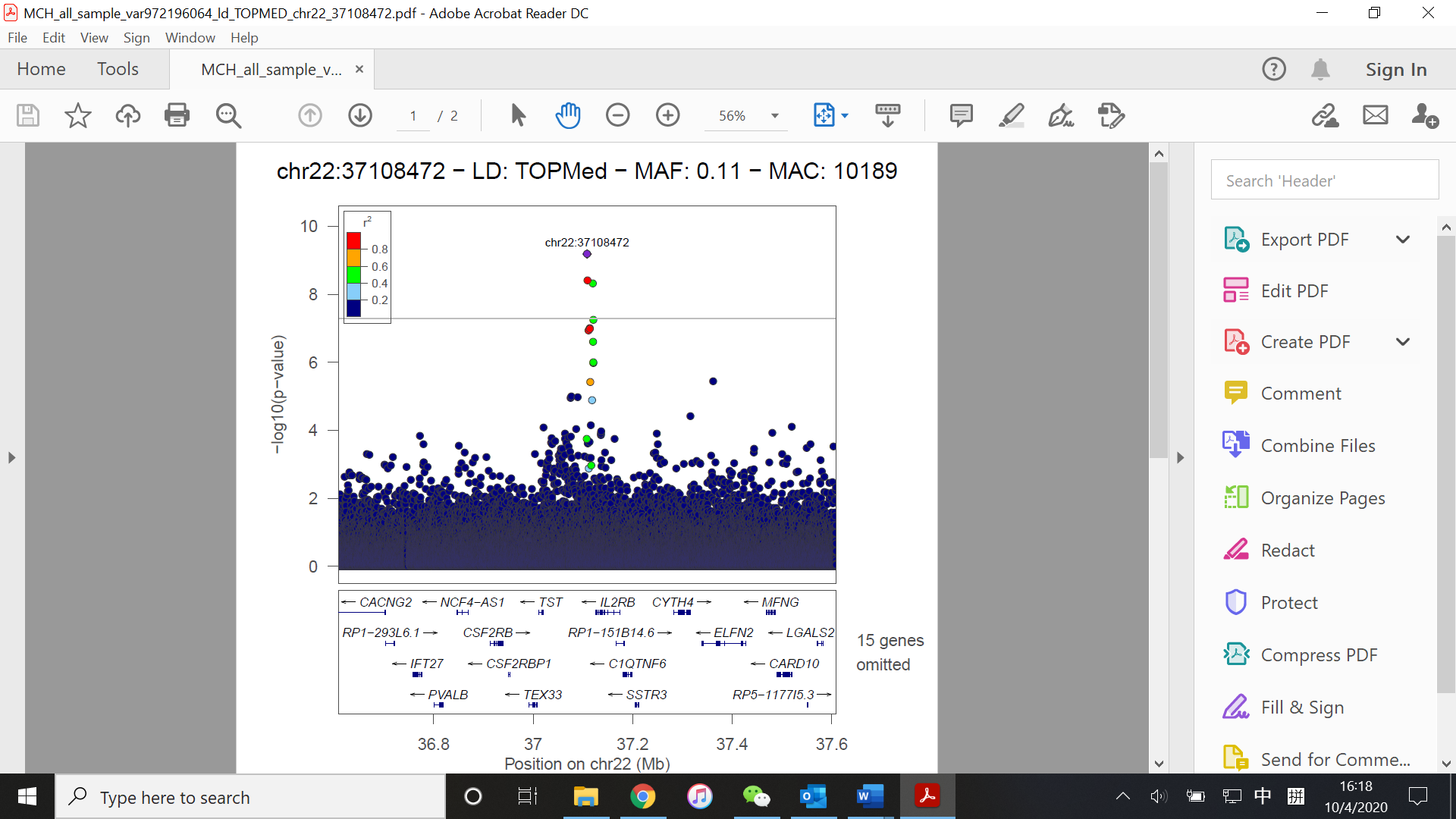

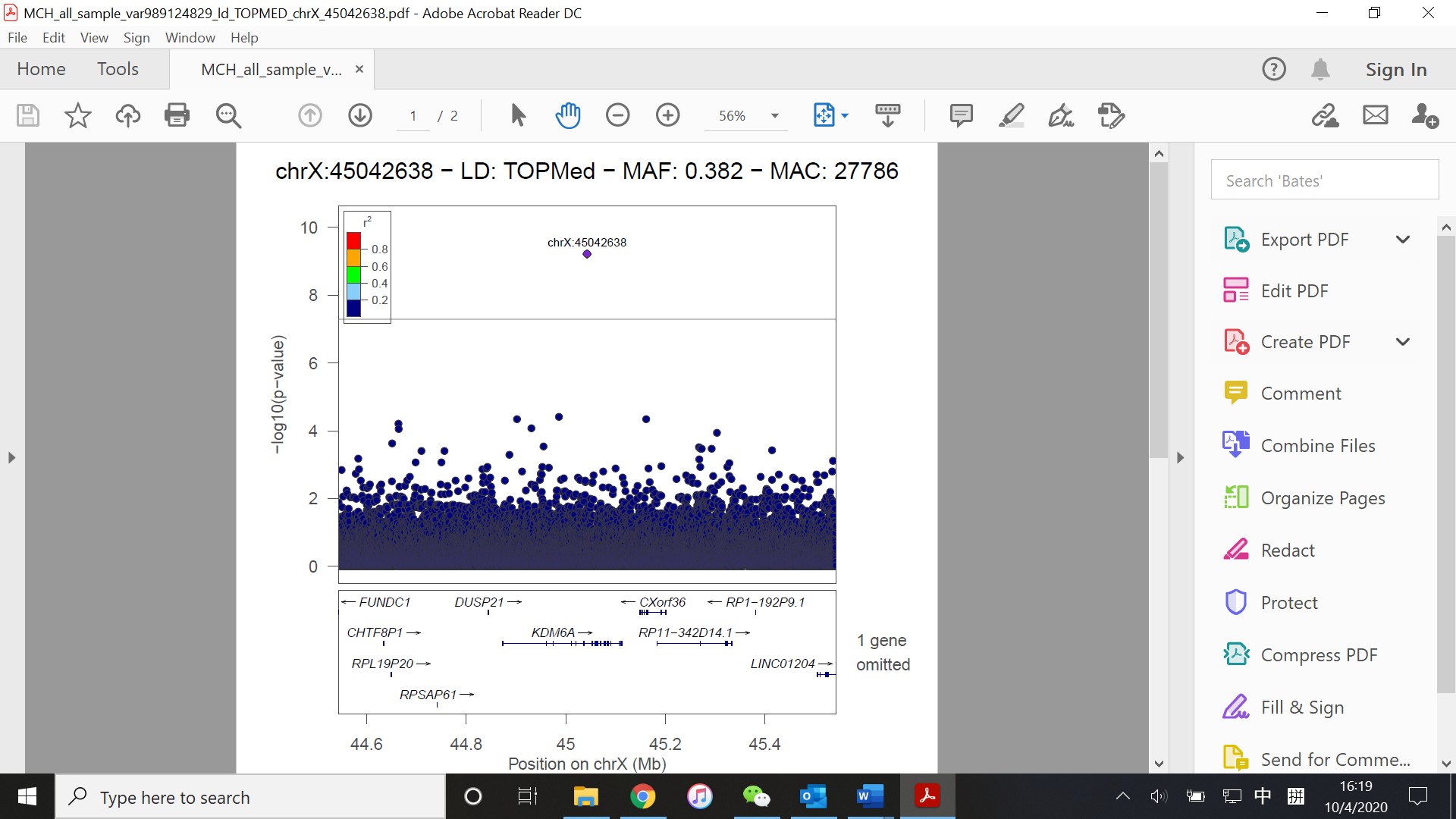

(D)

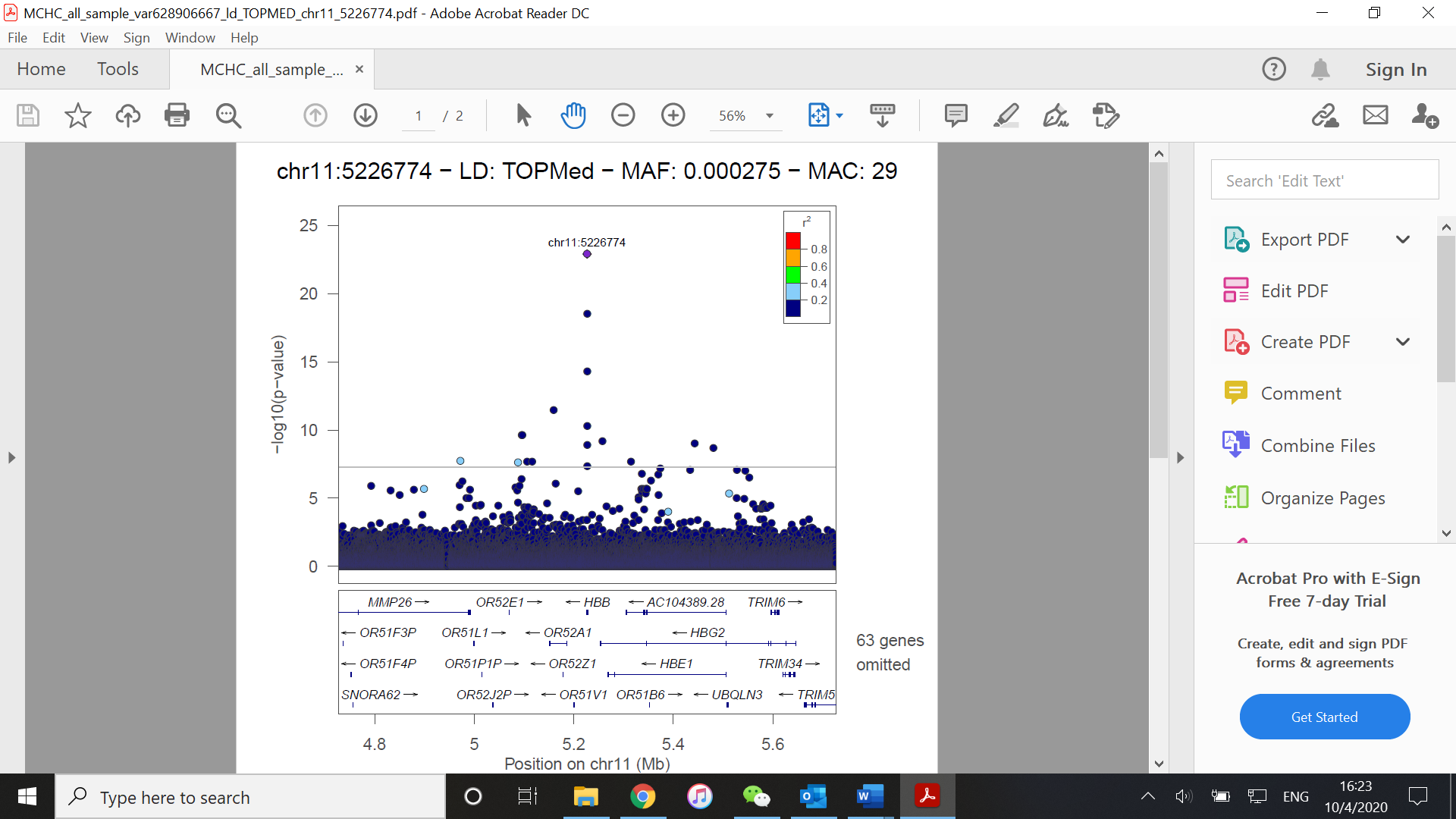

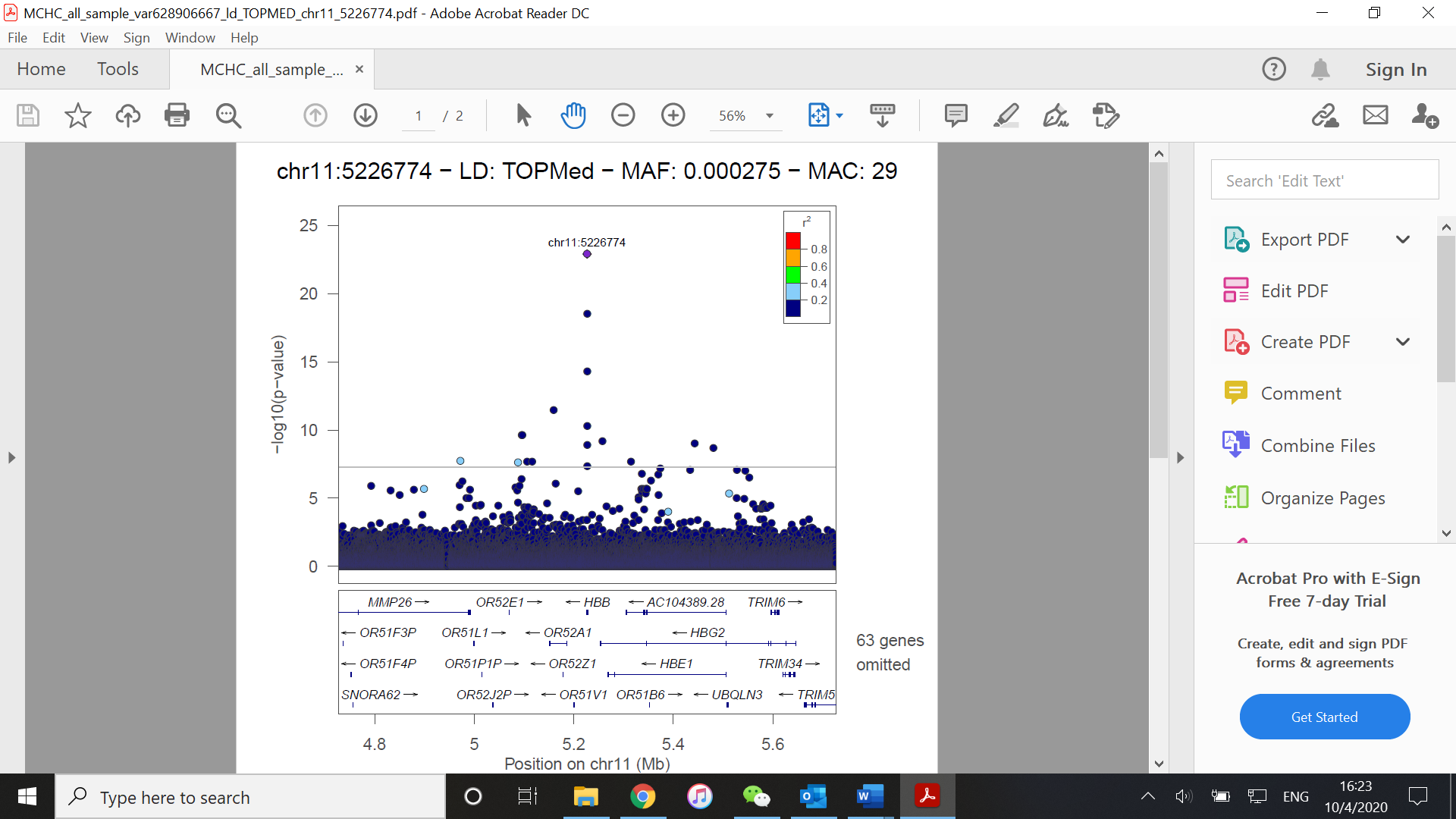

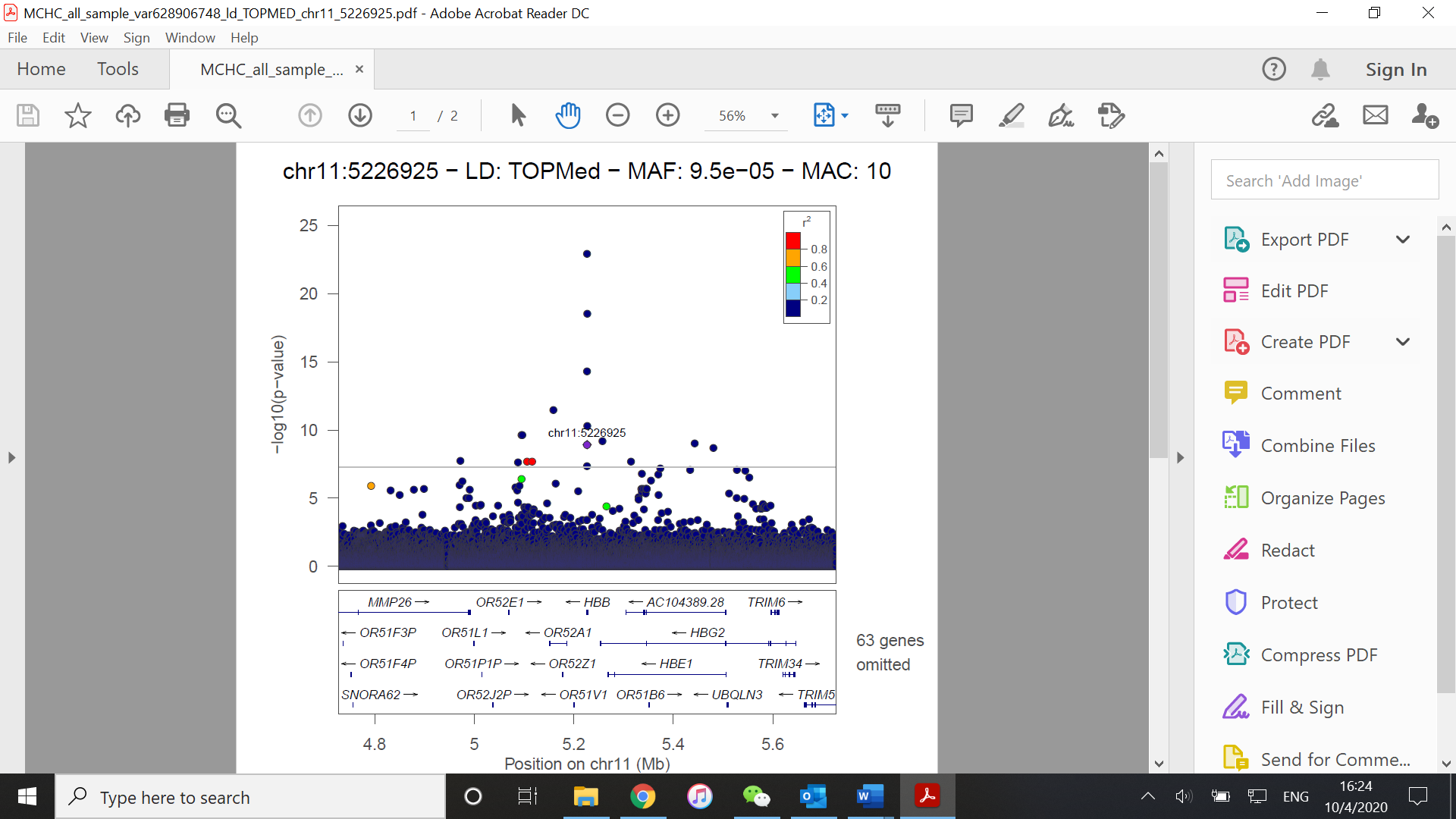

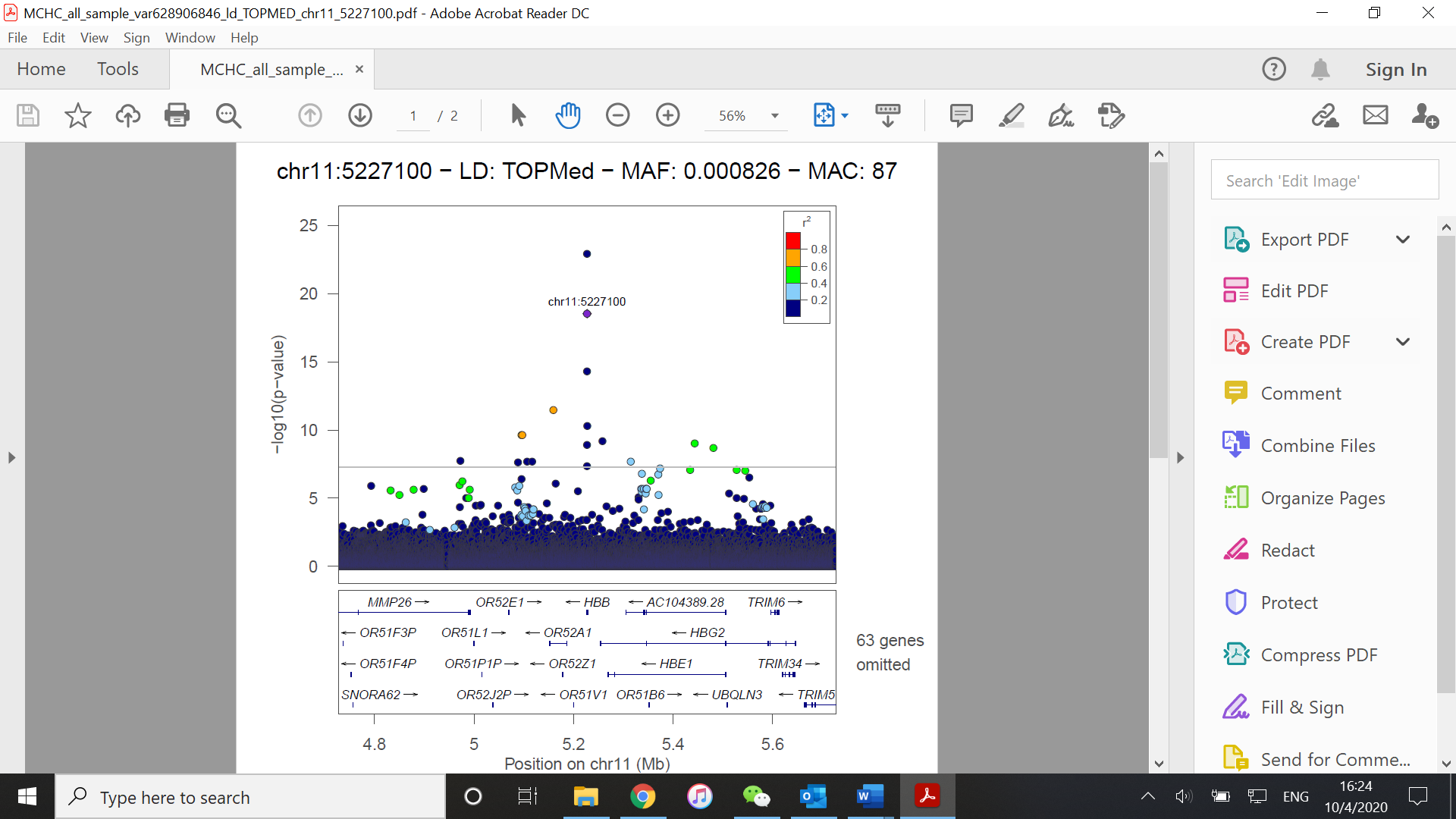

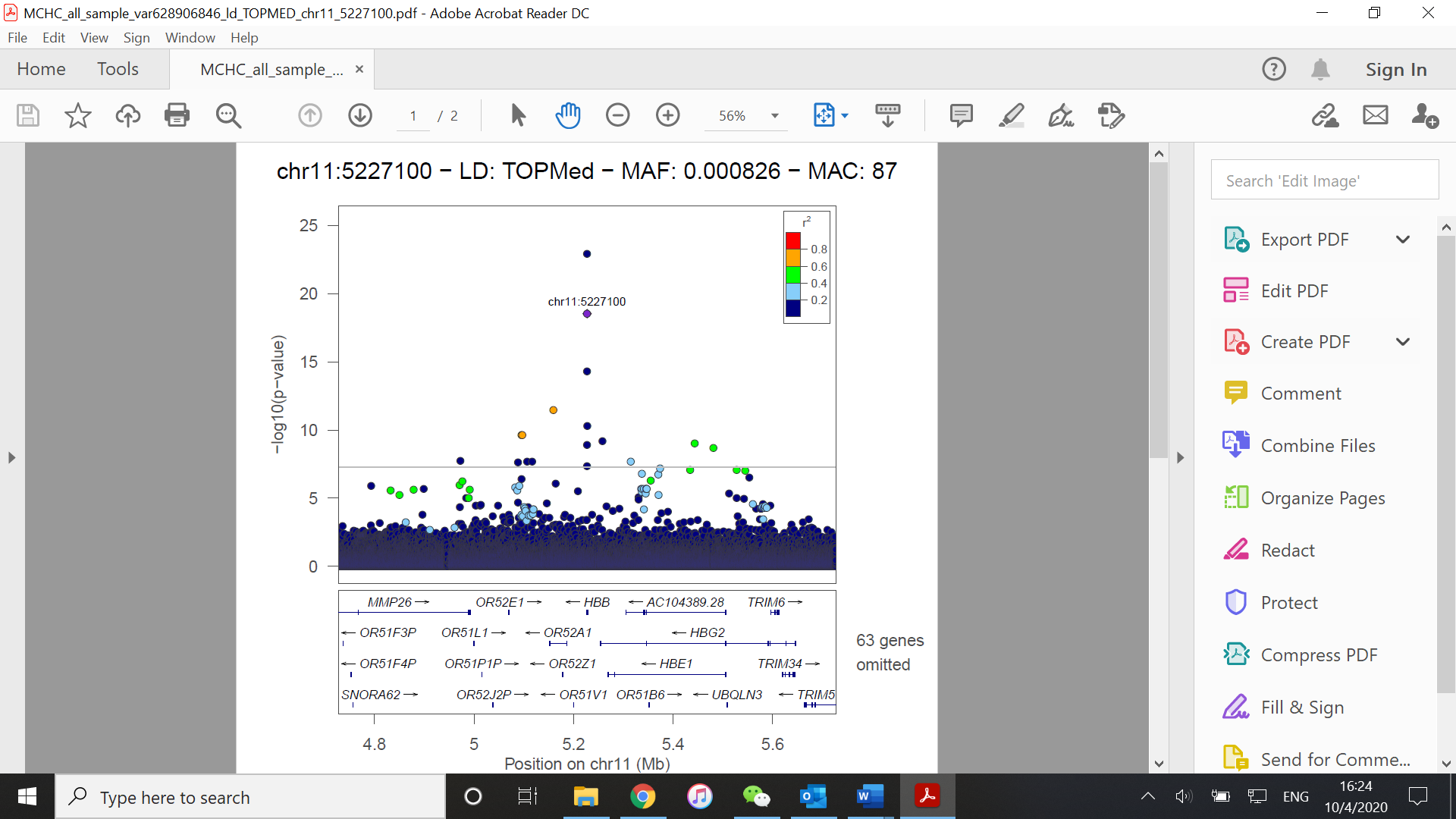

(E)

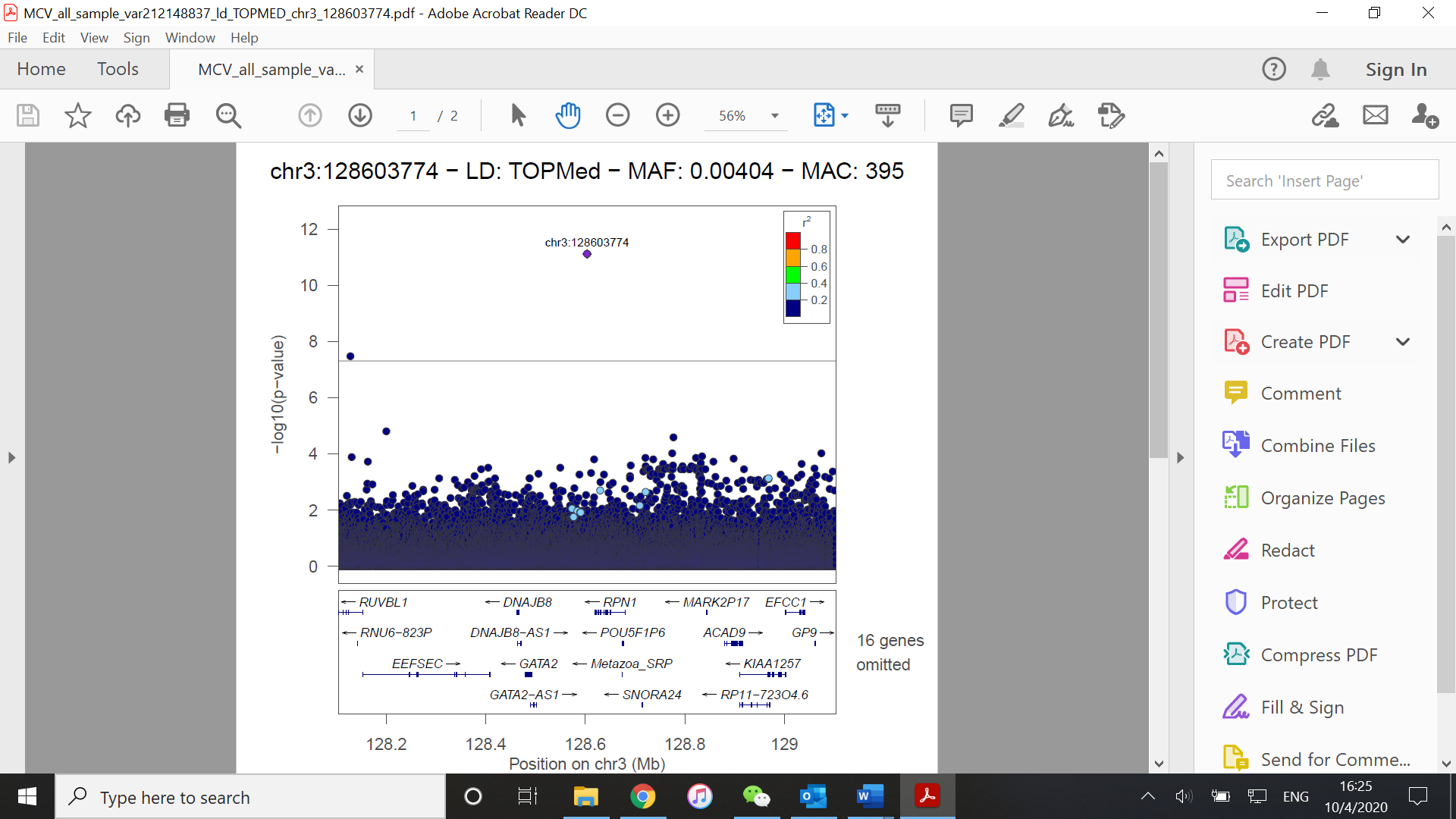

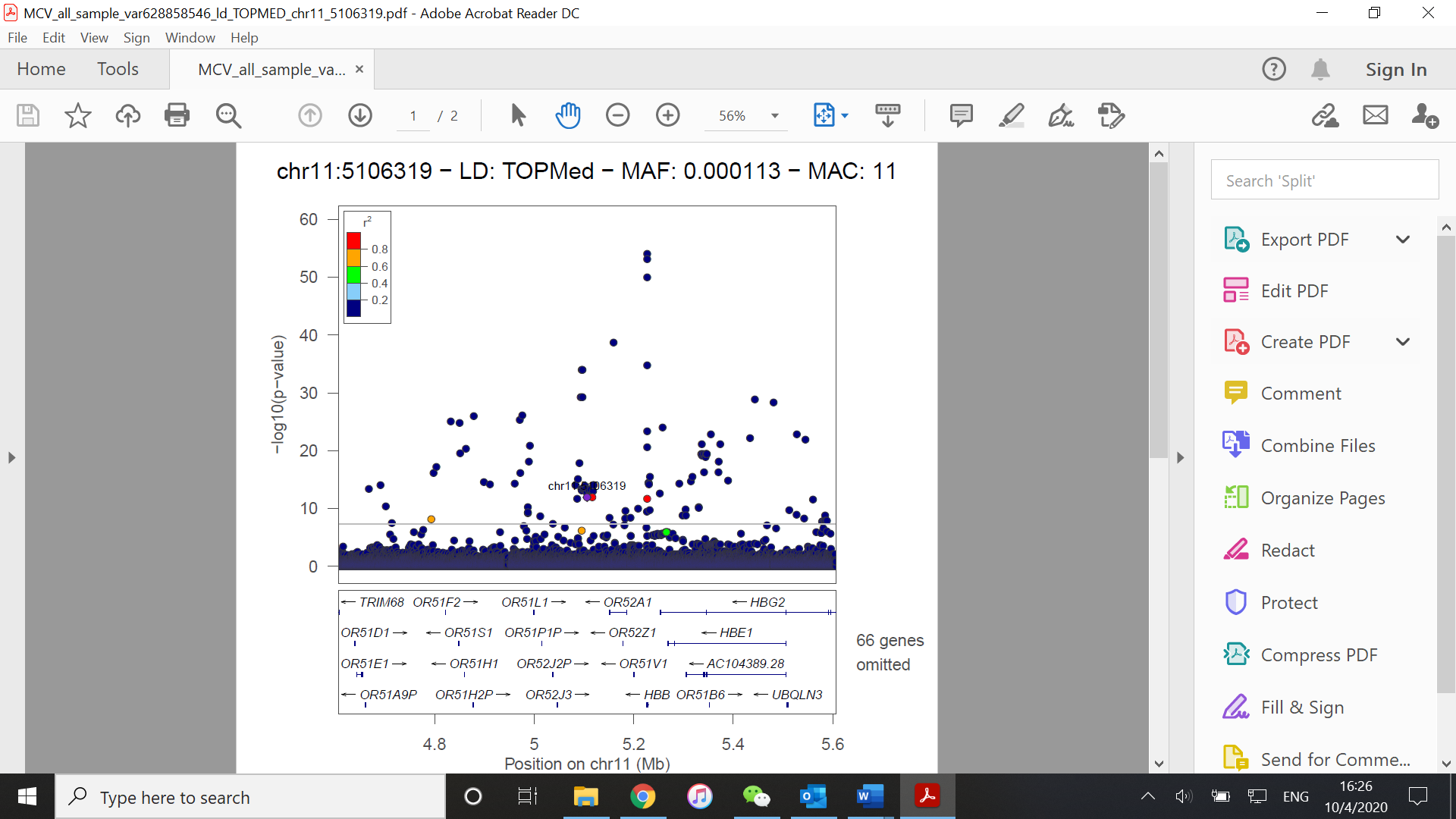

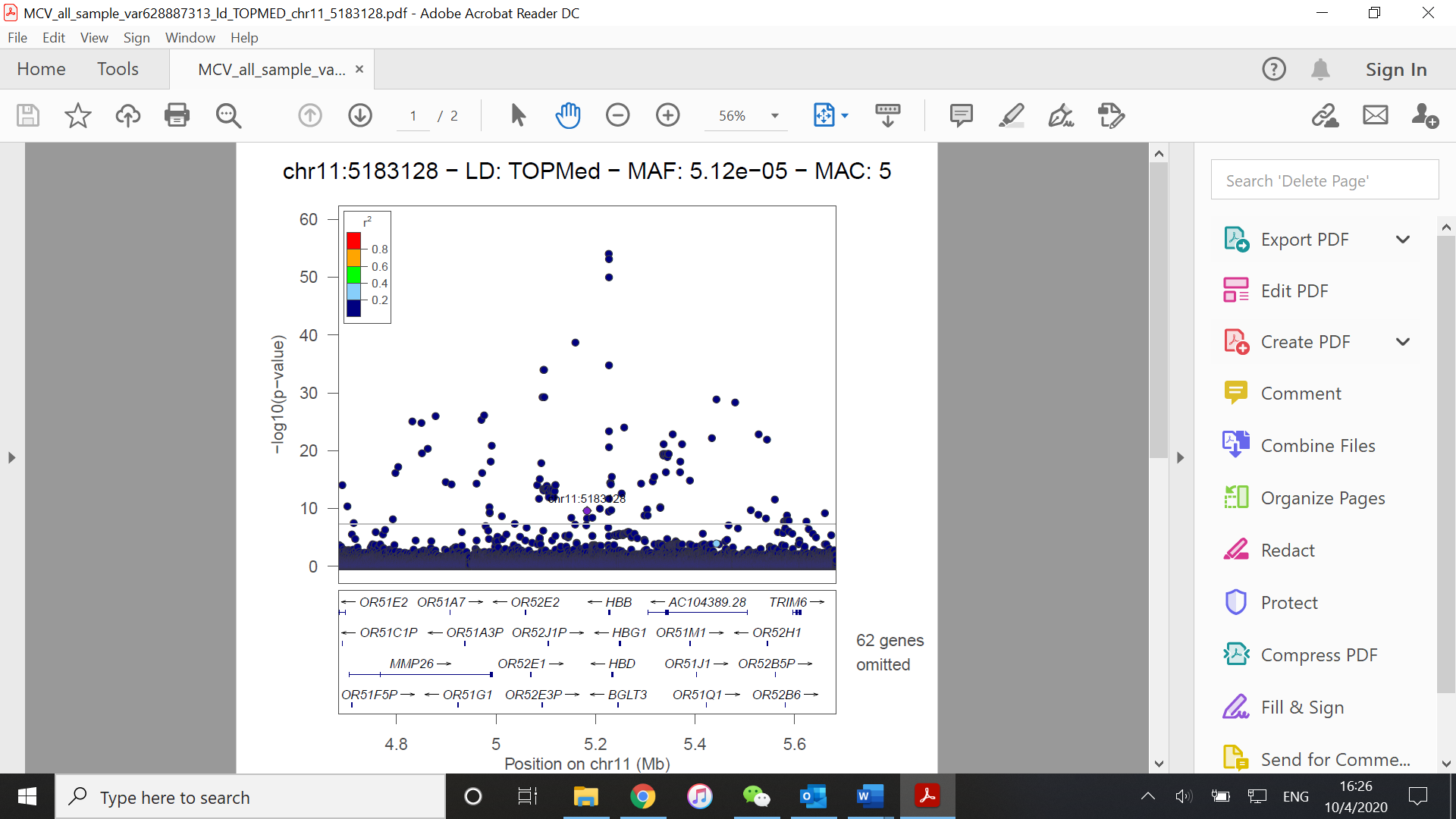

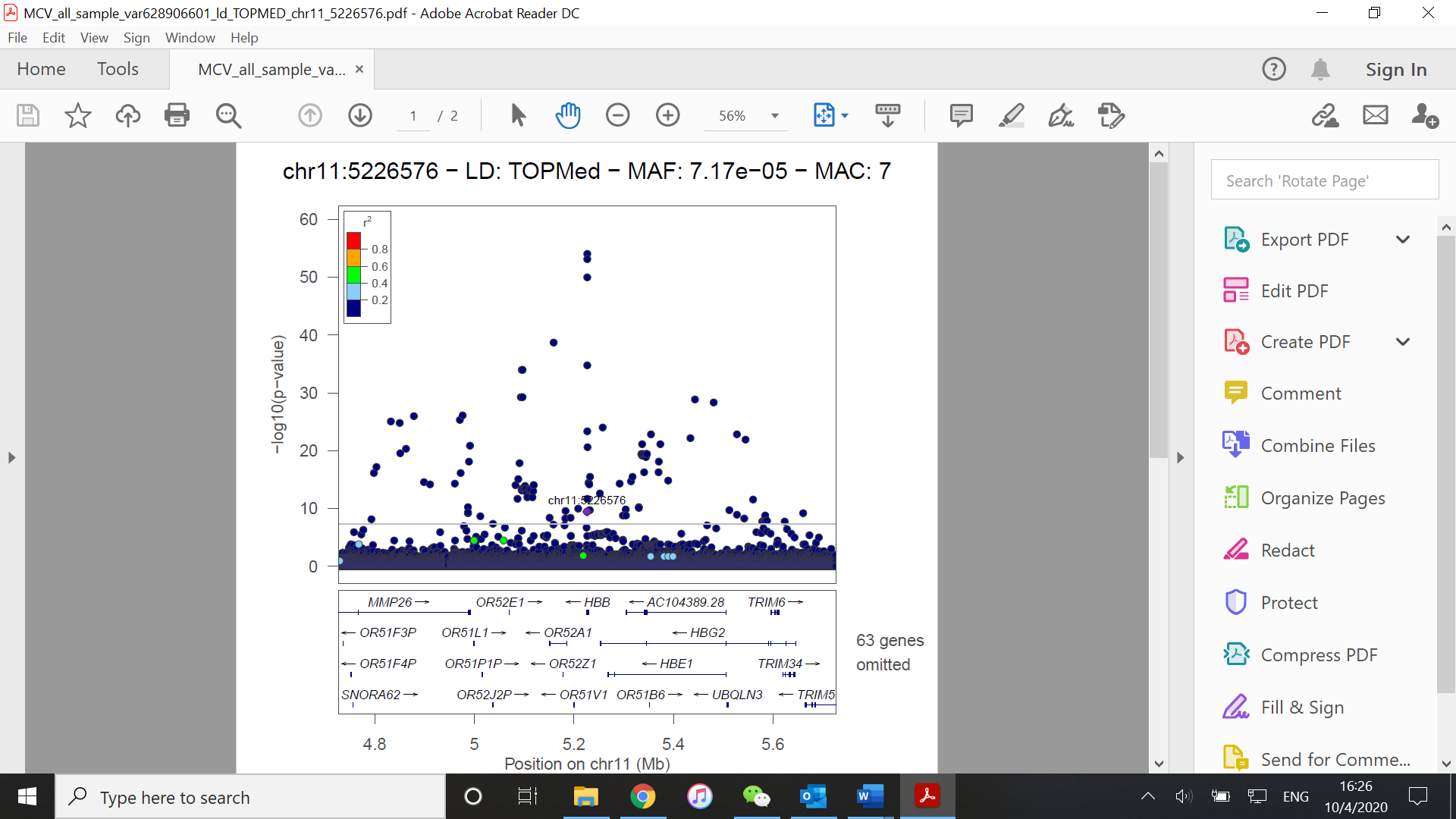

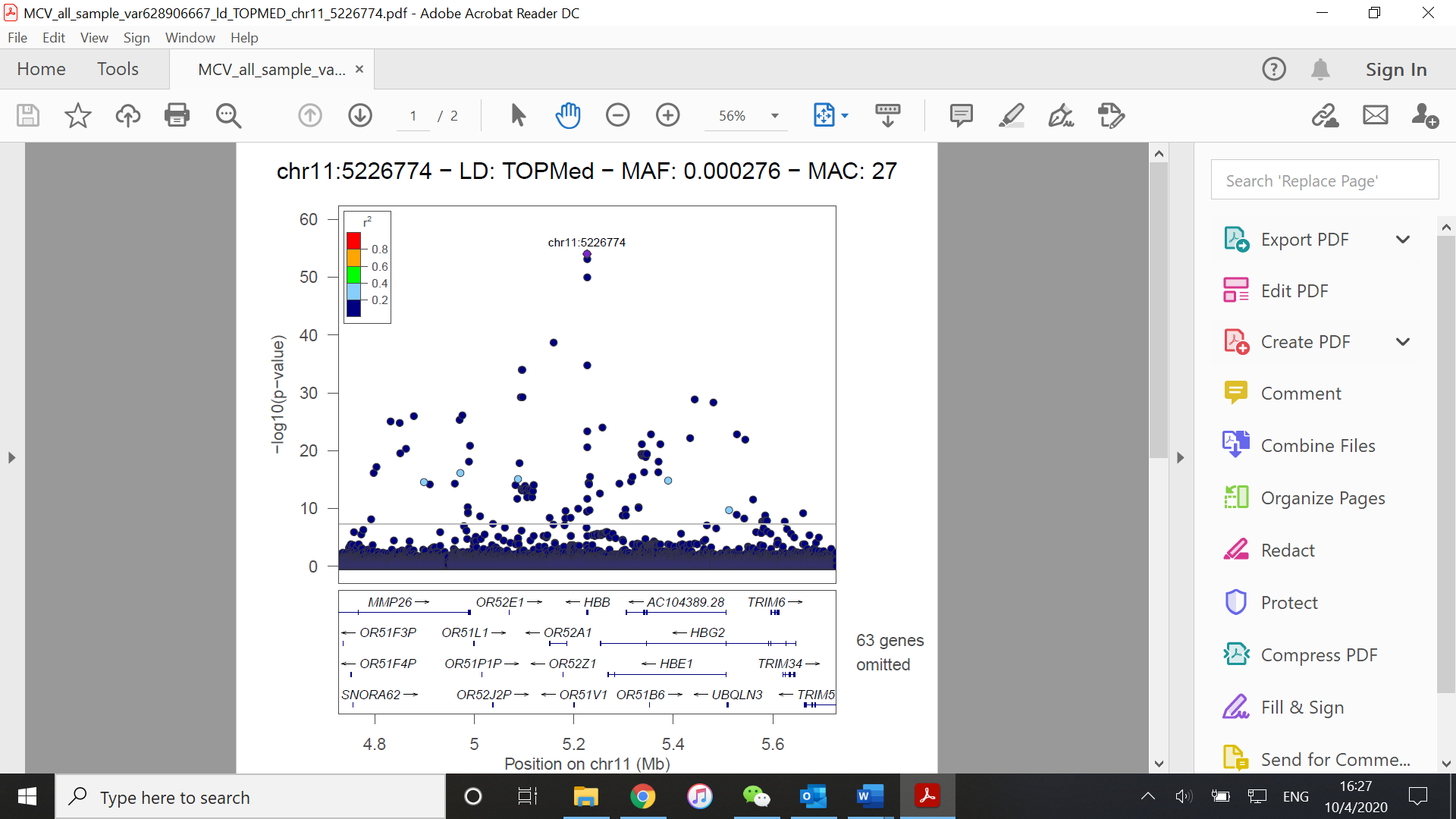

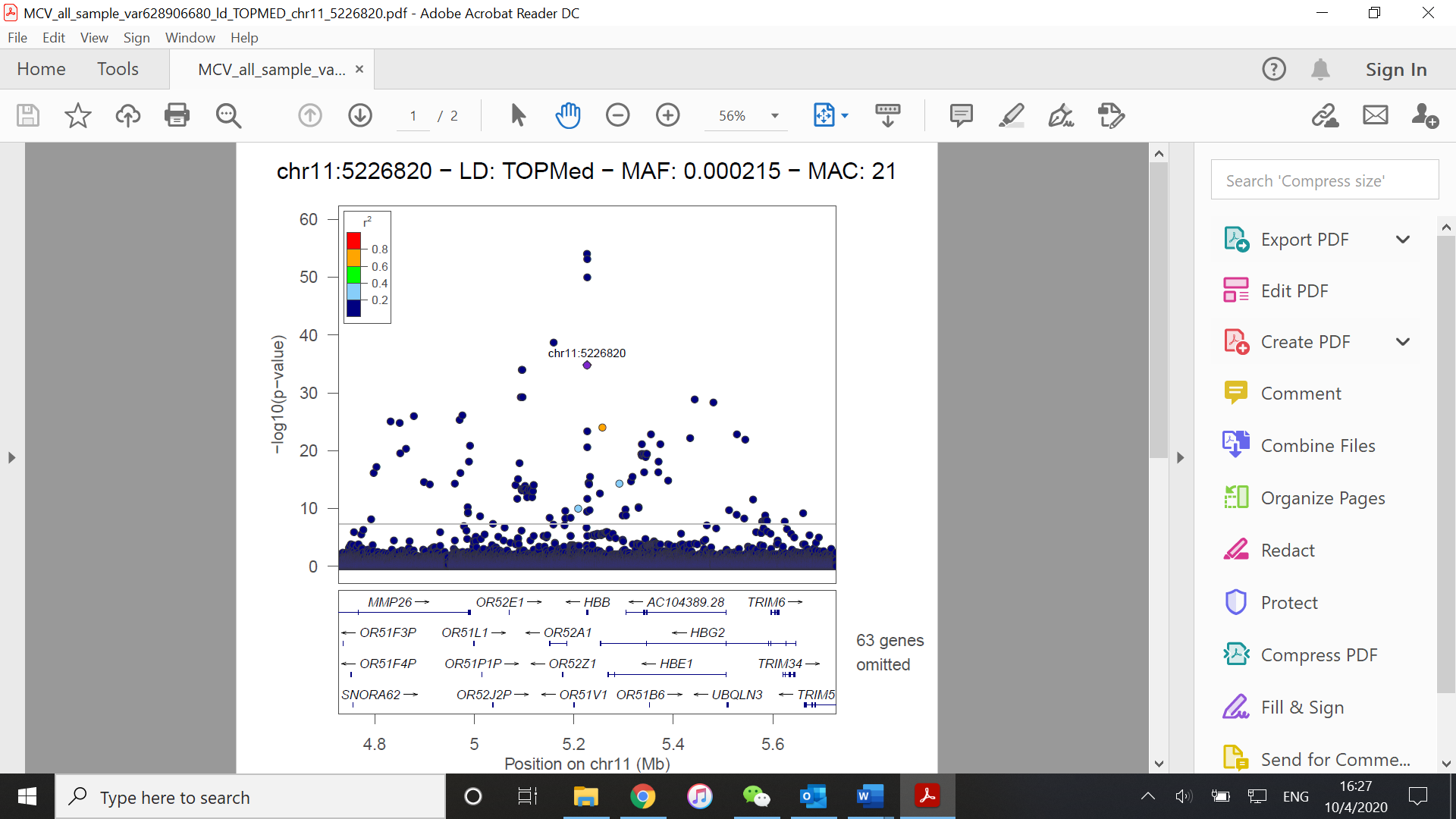

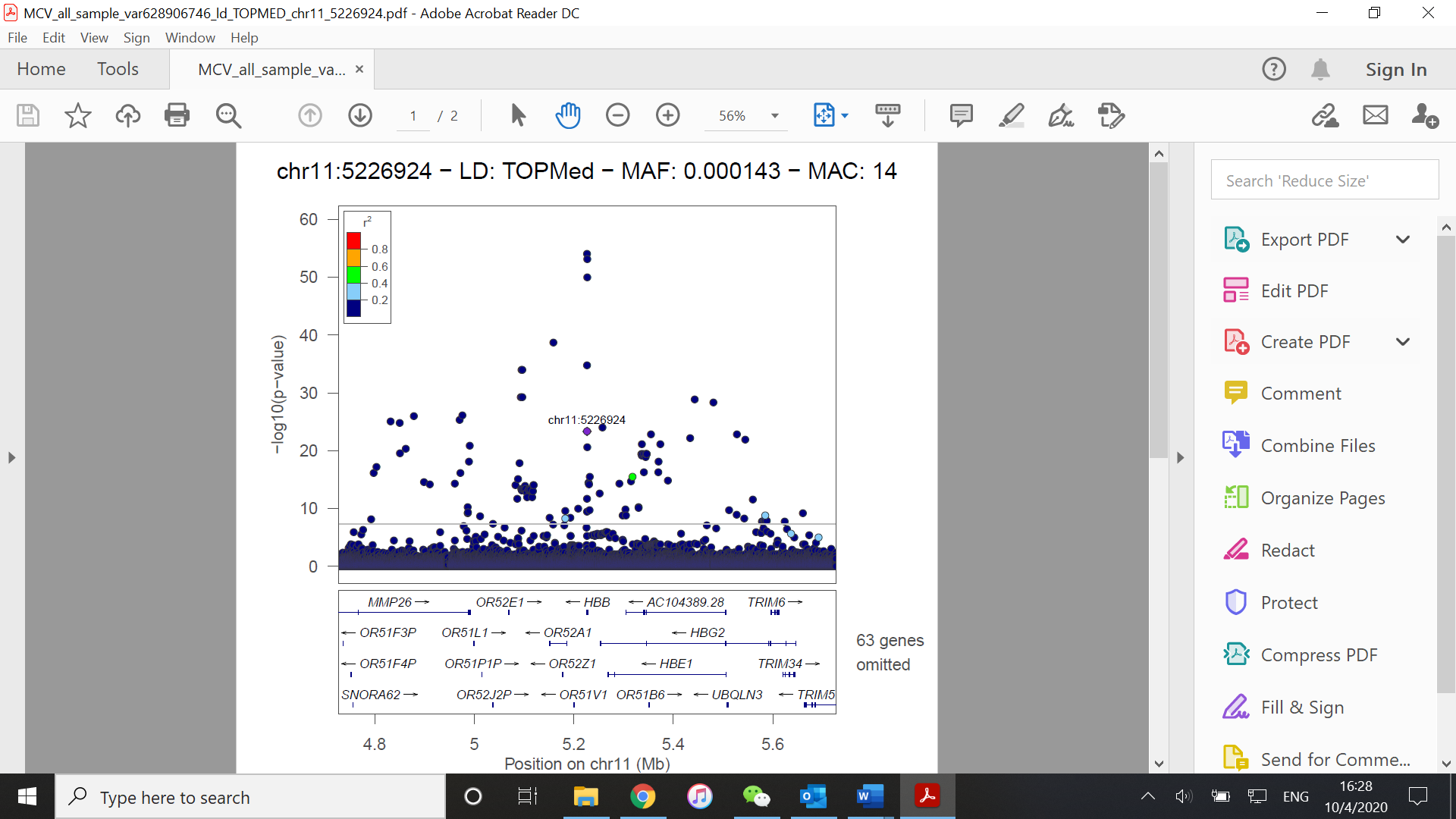

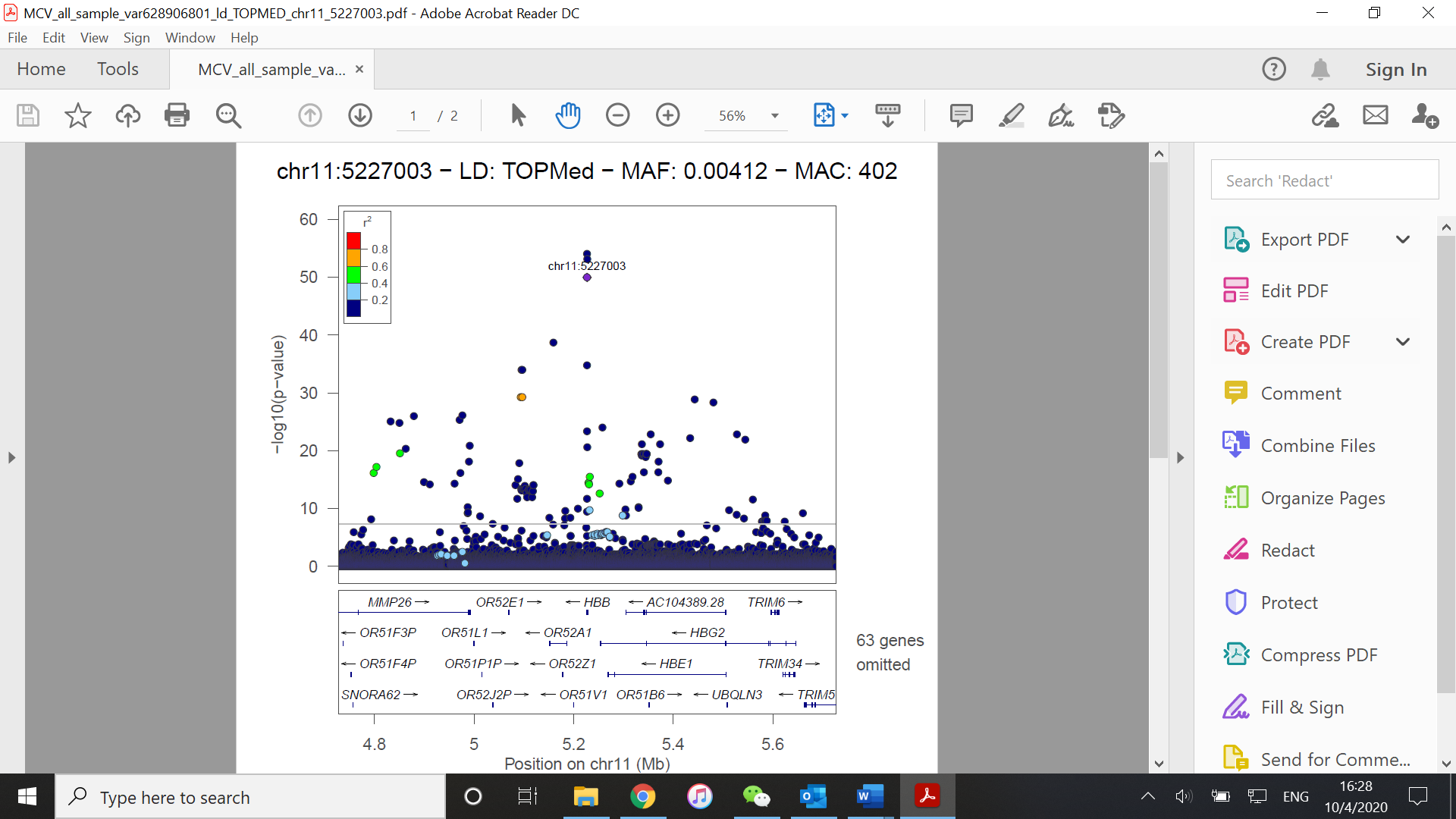

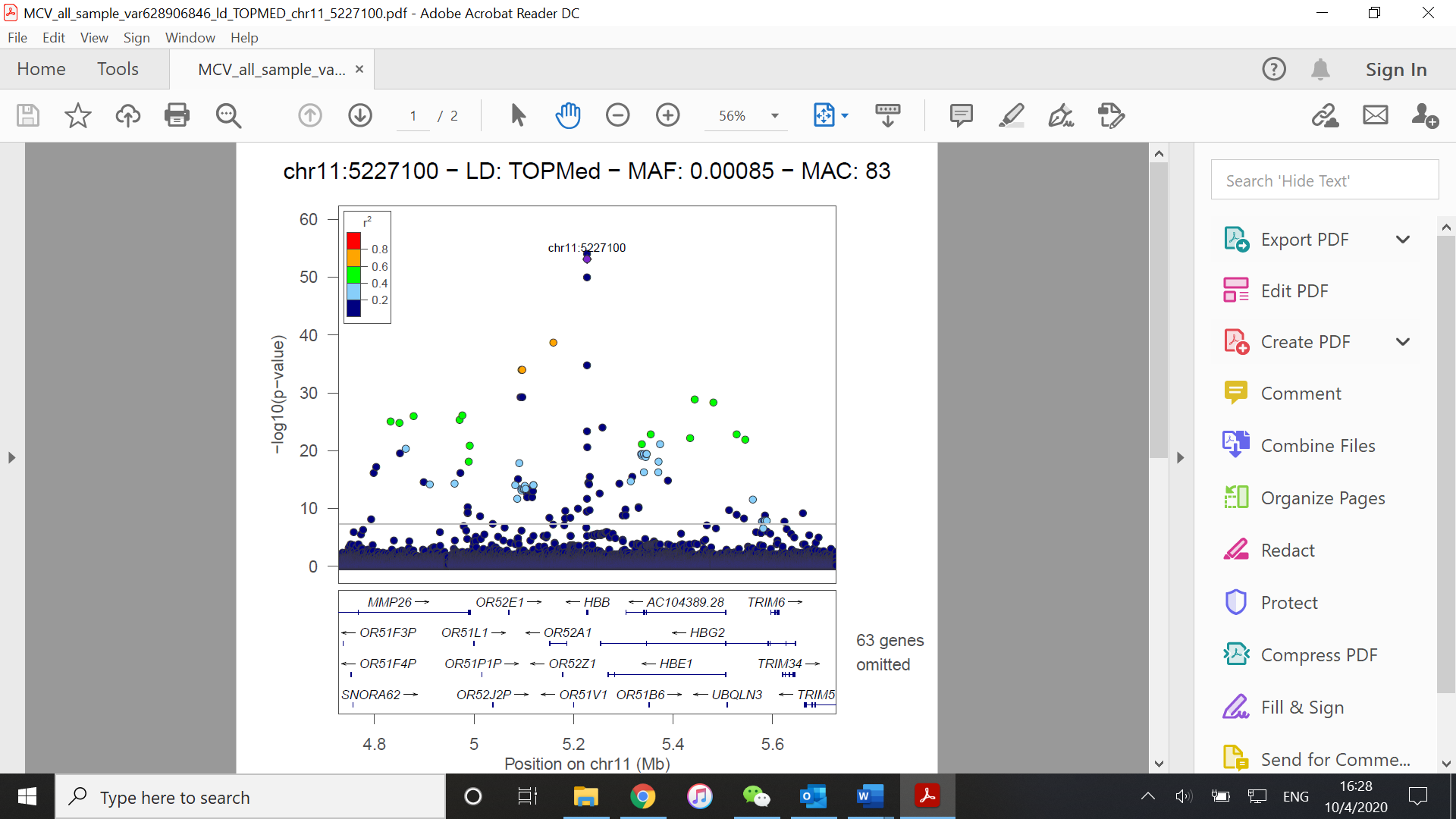

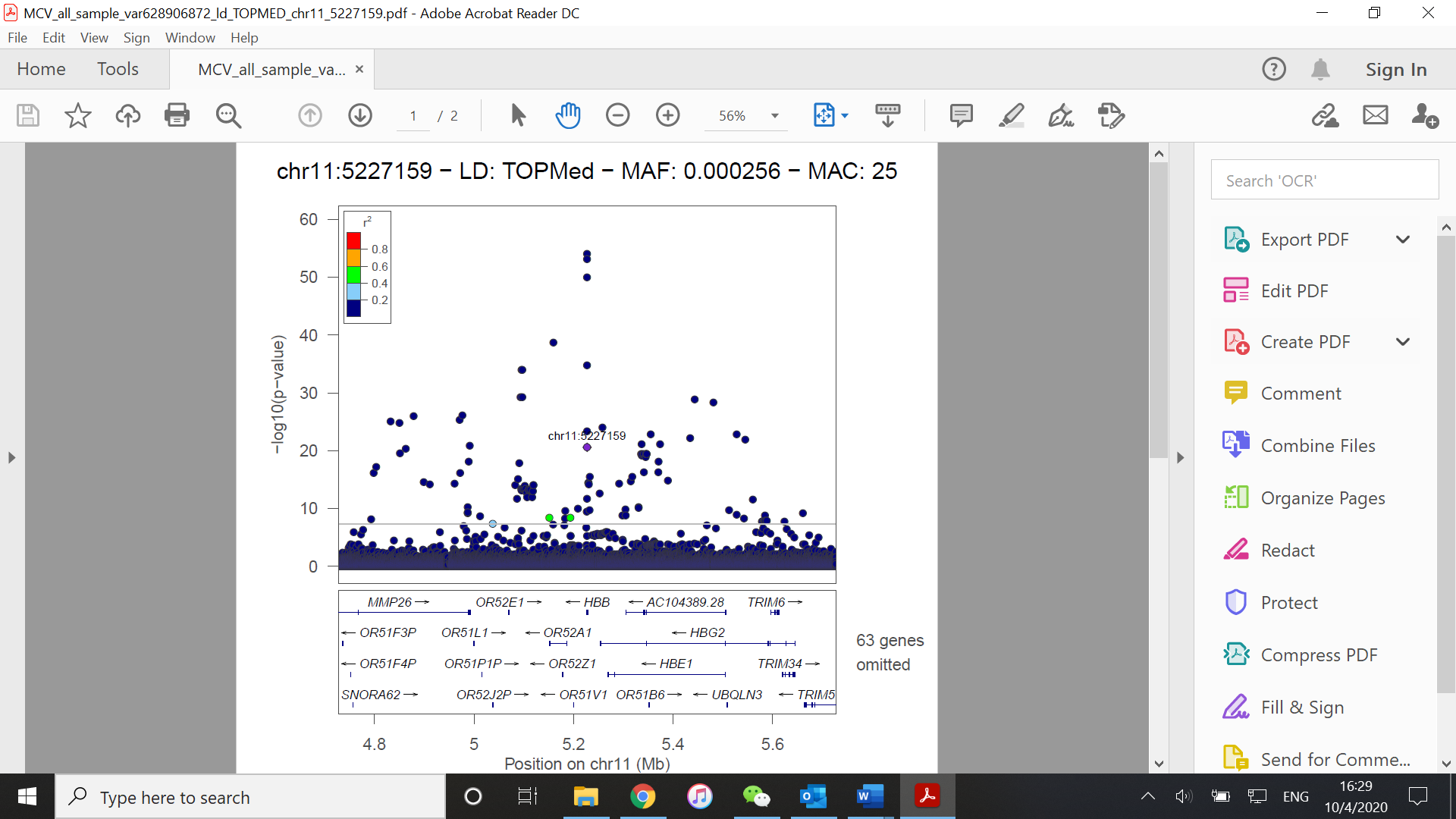

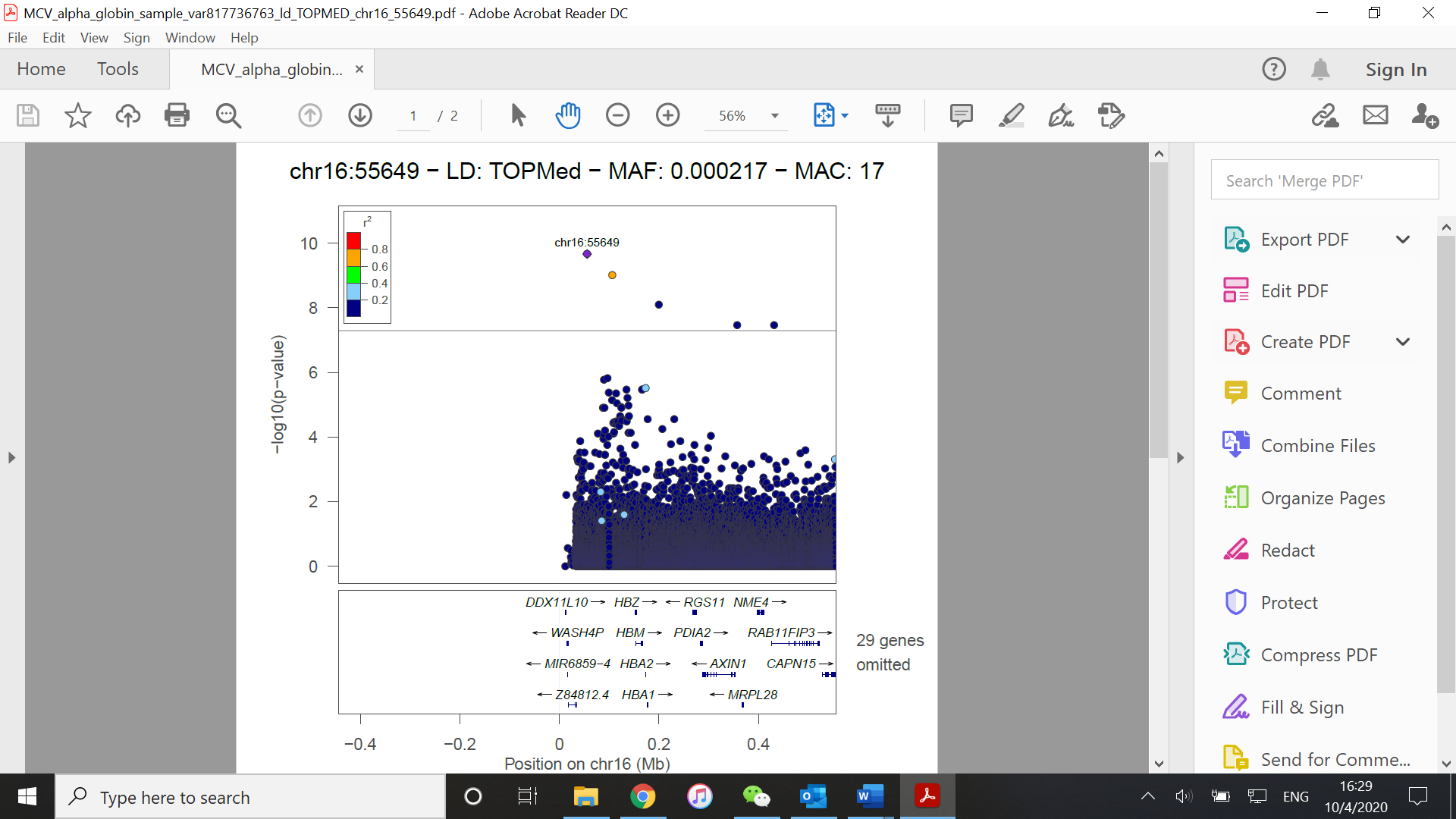

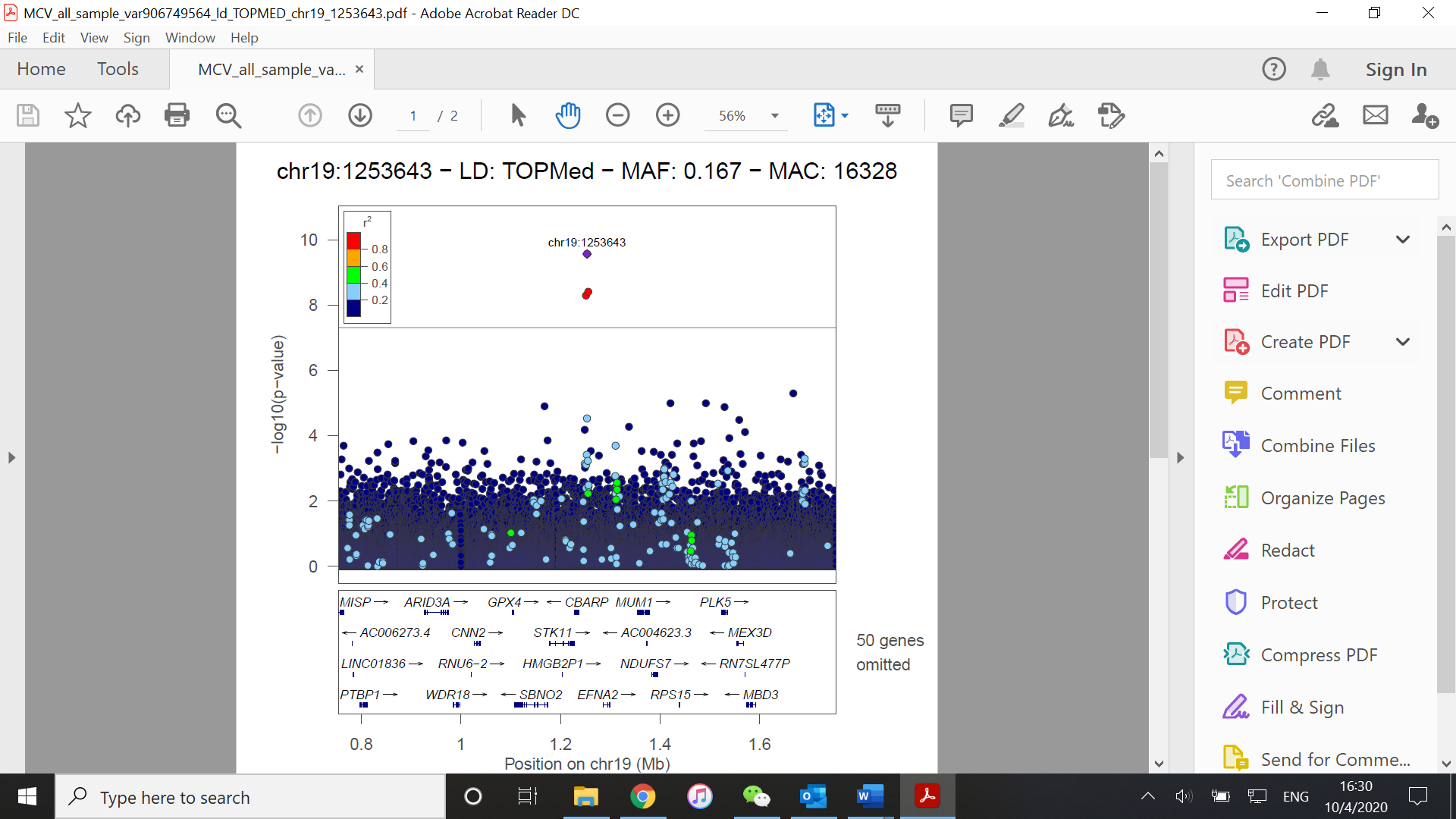

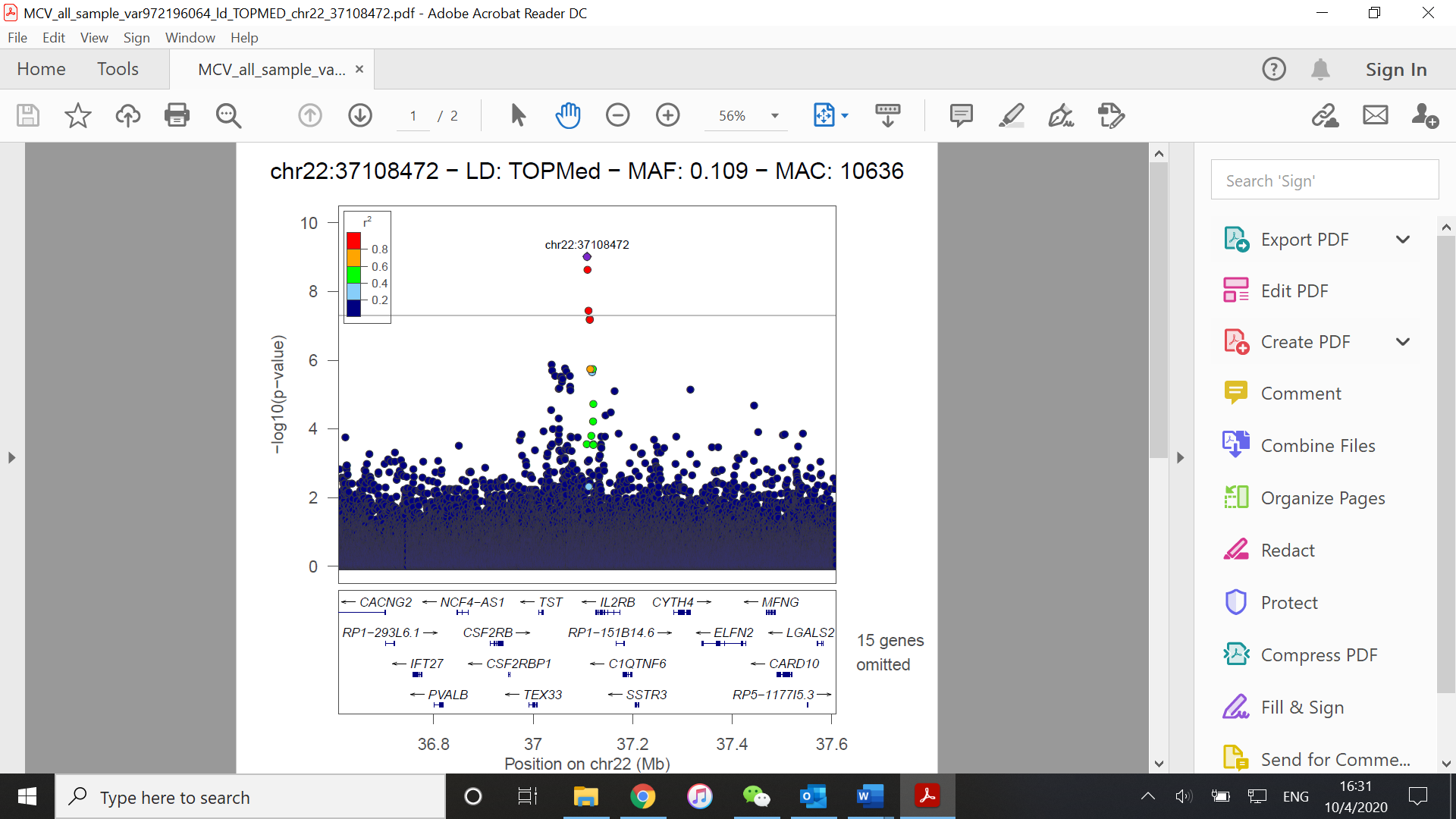

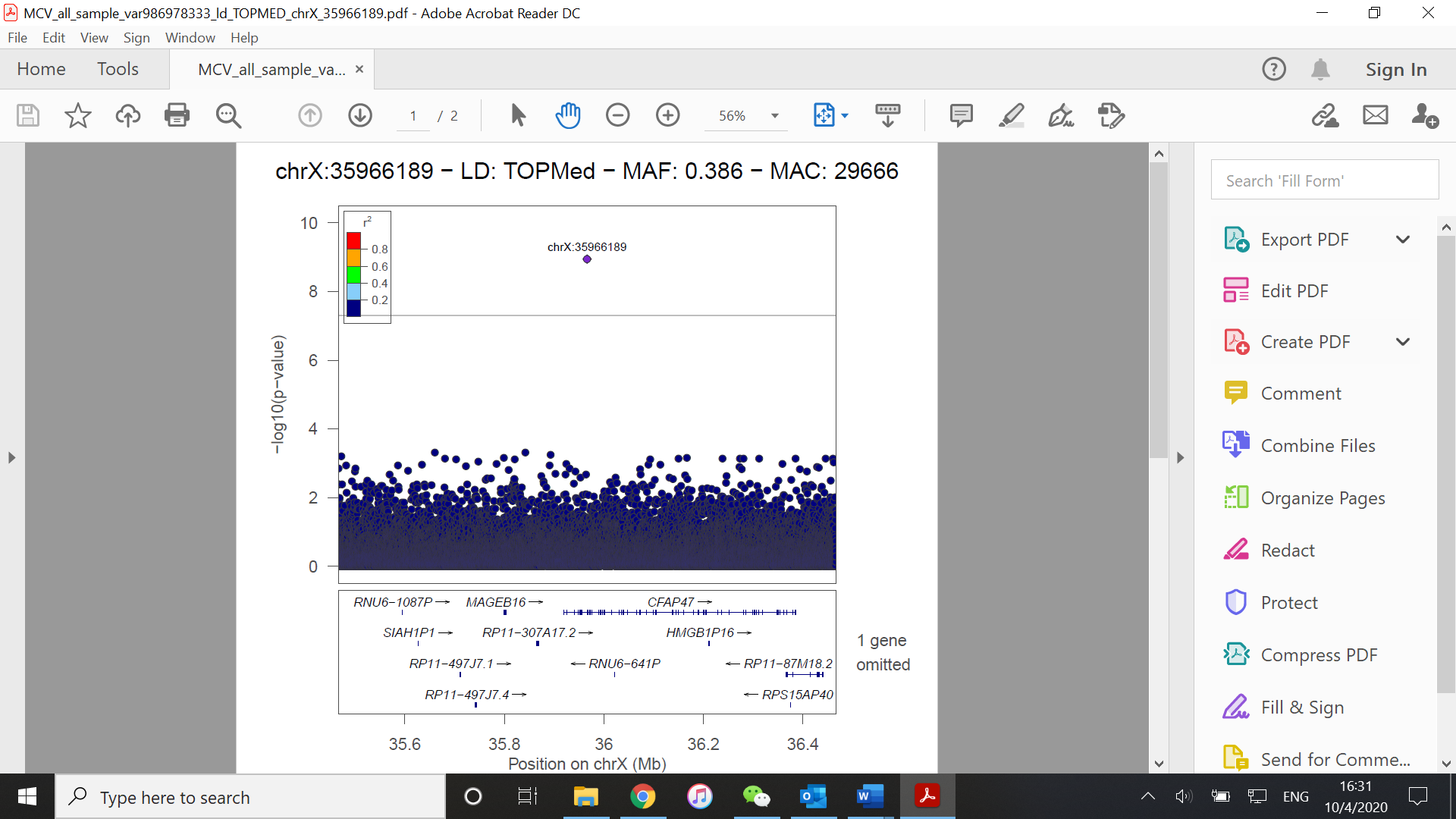

(F)

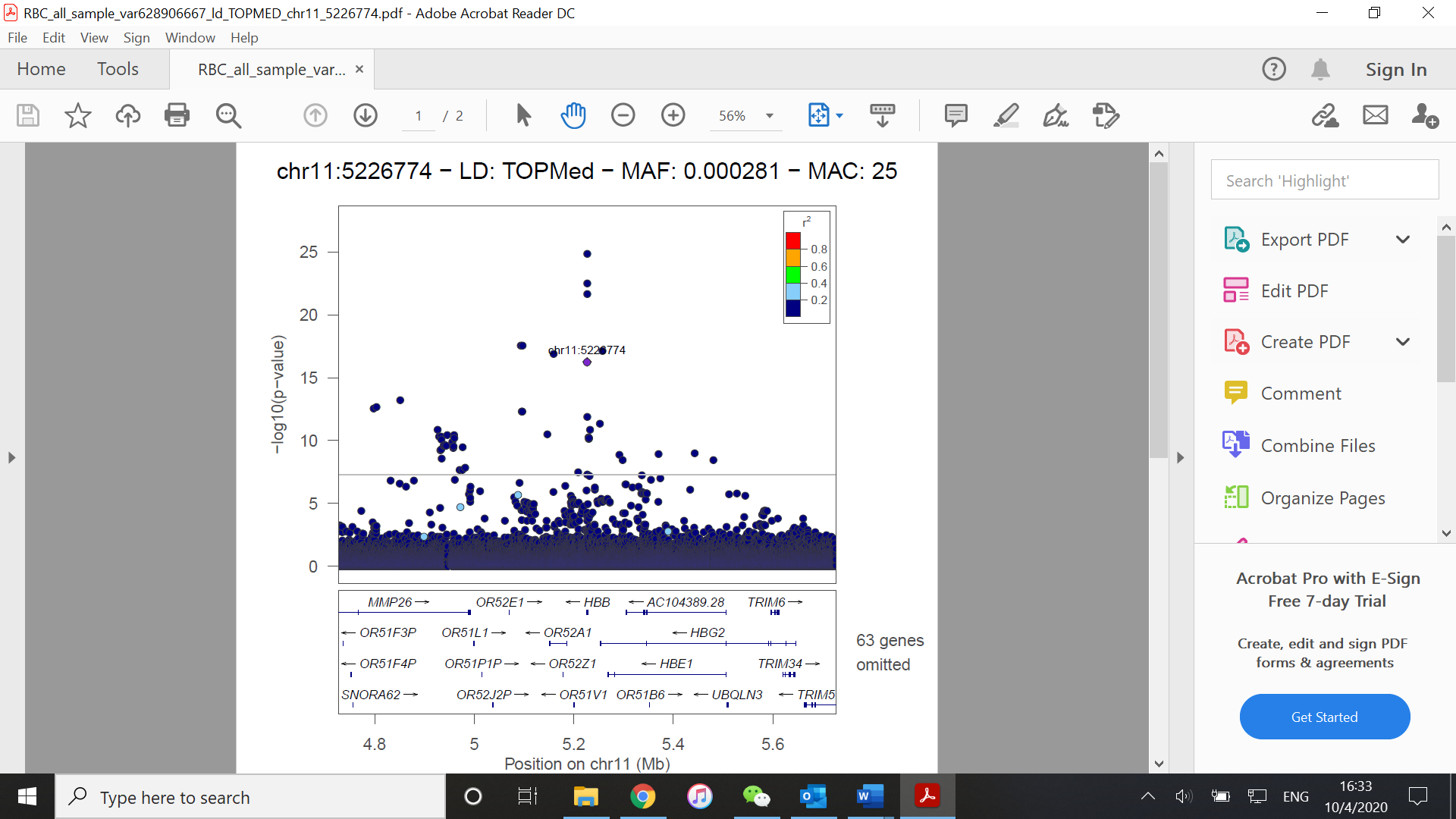

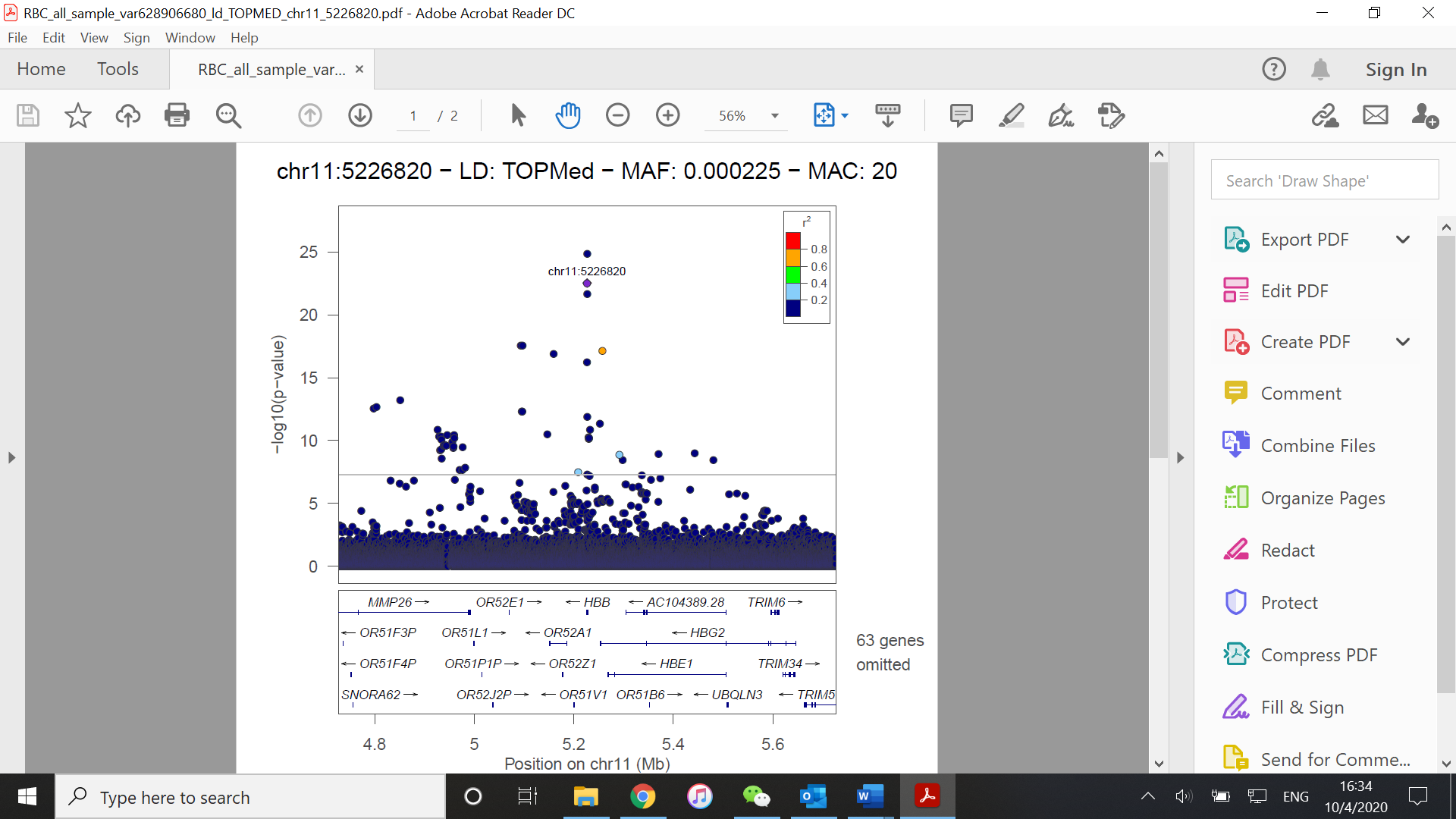

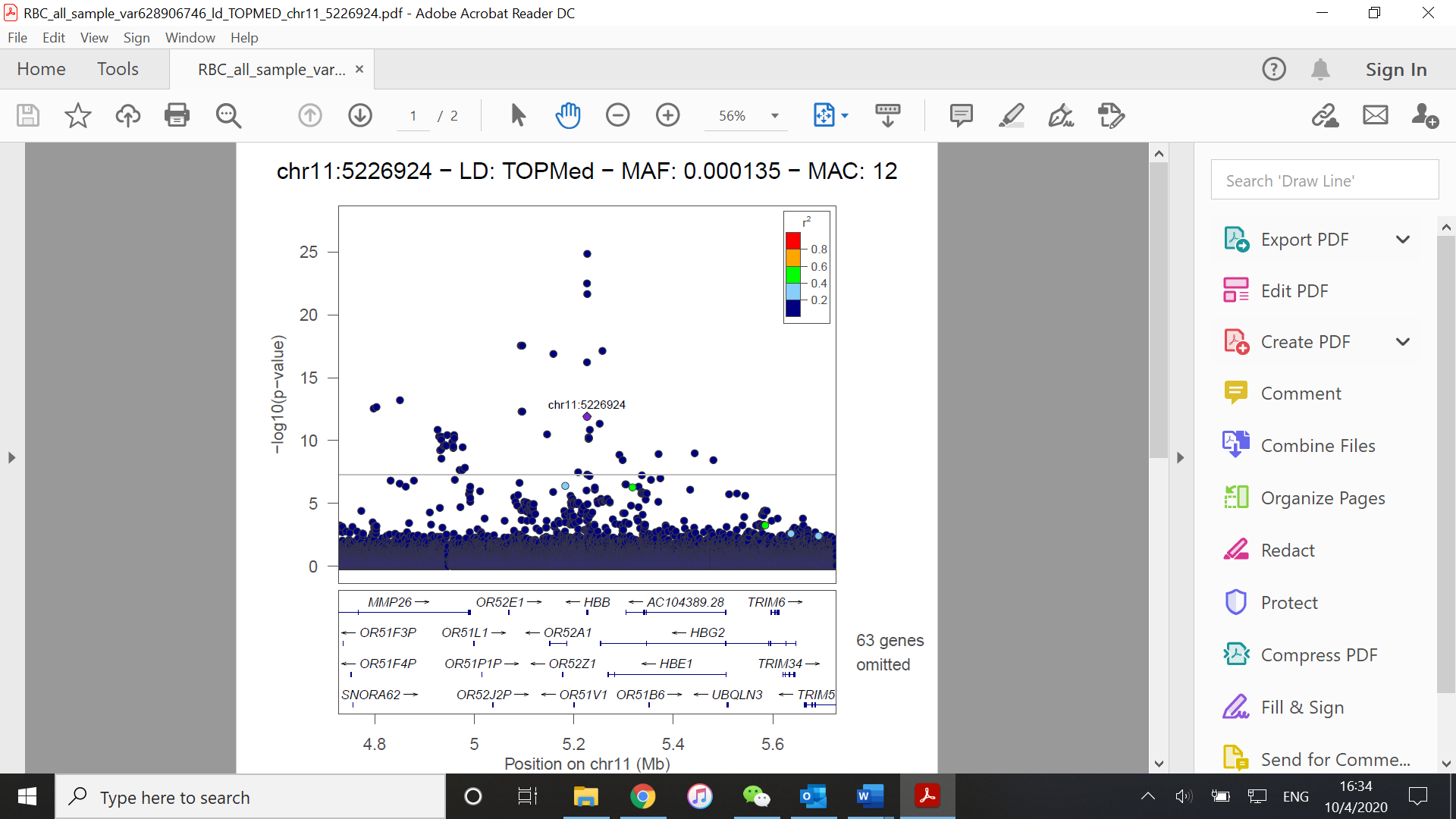

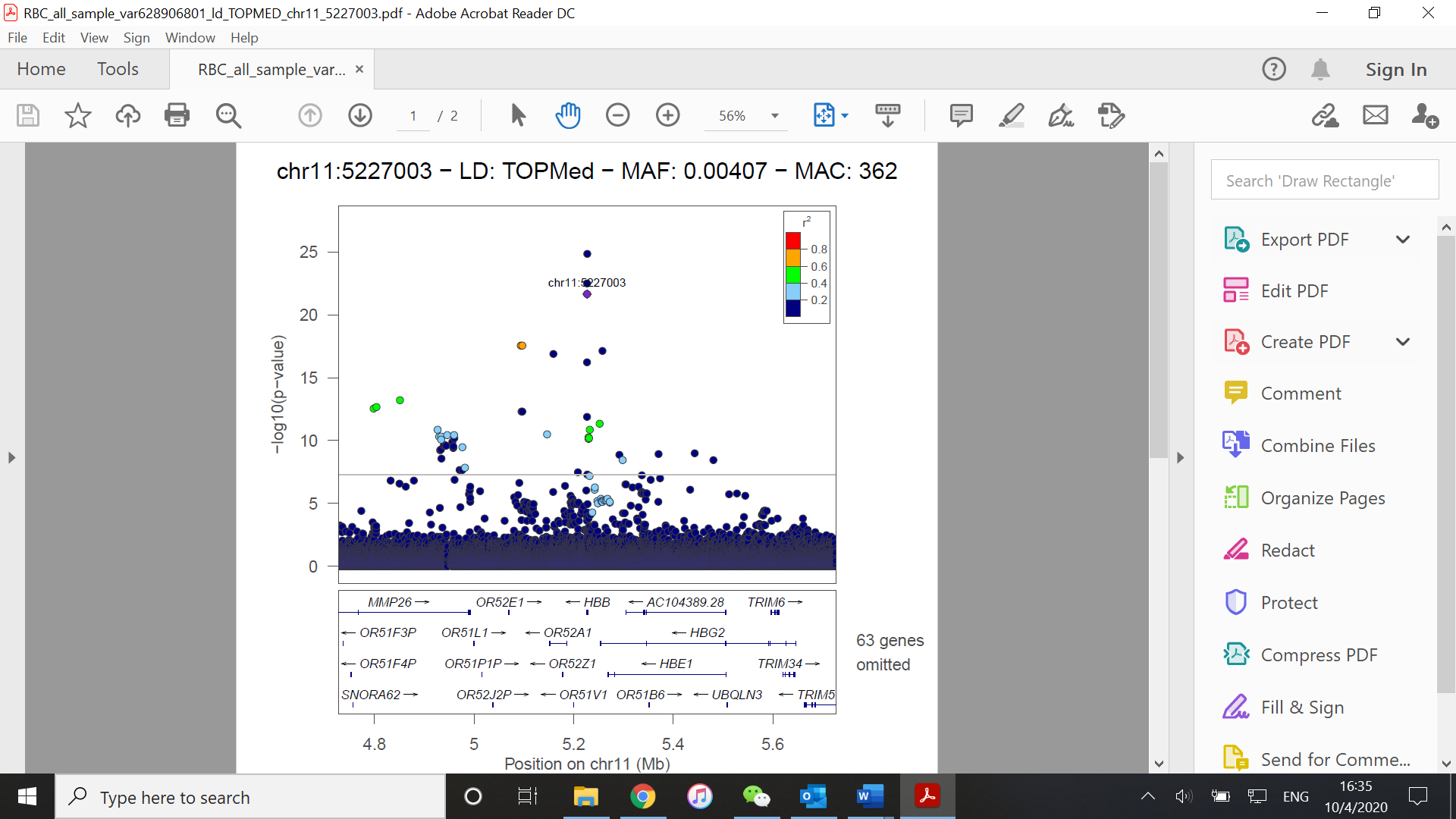

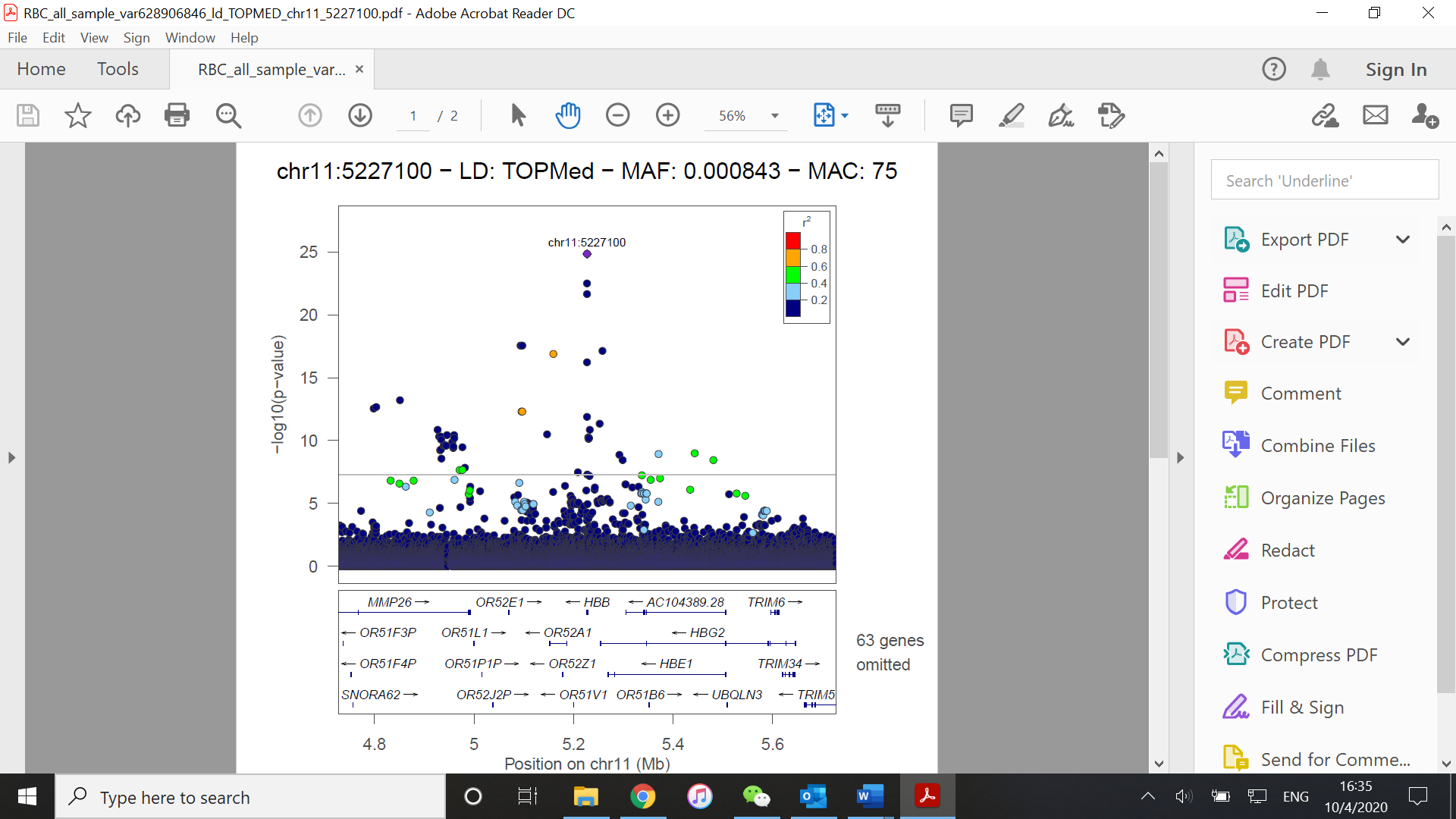

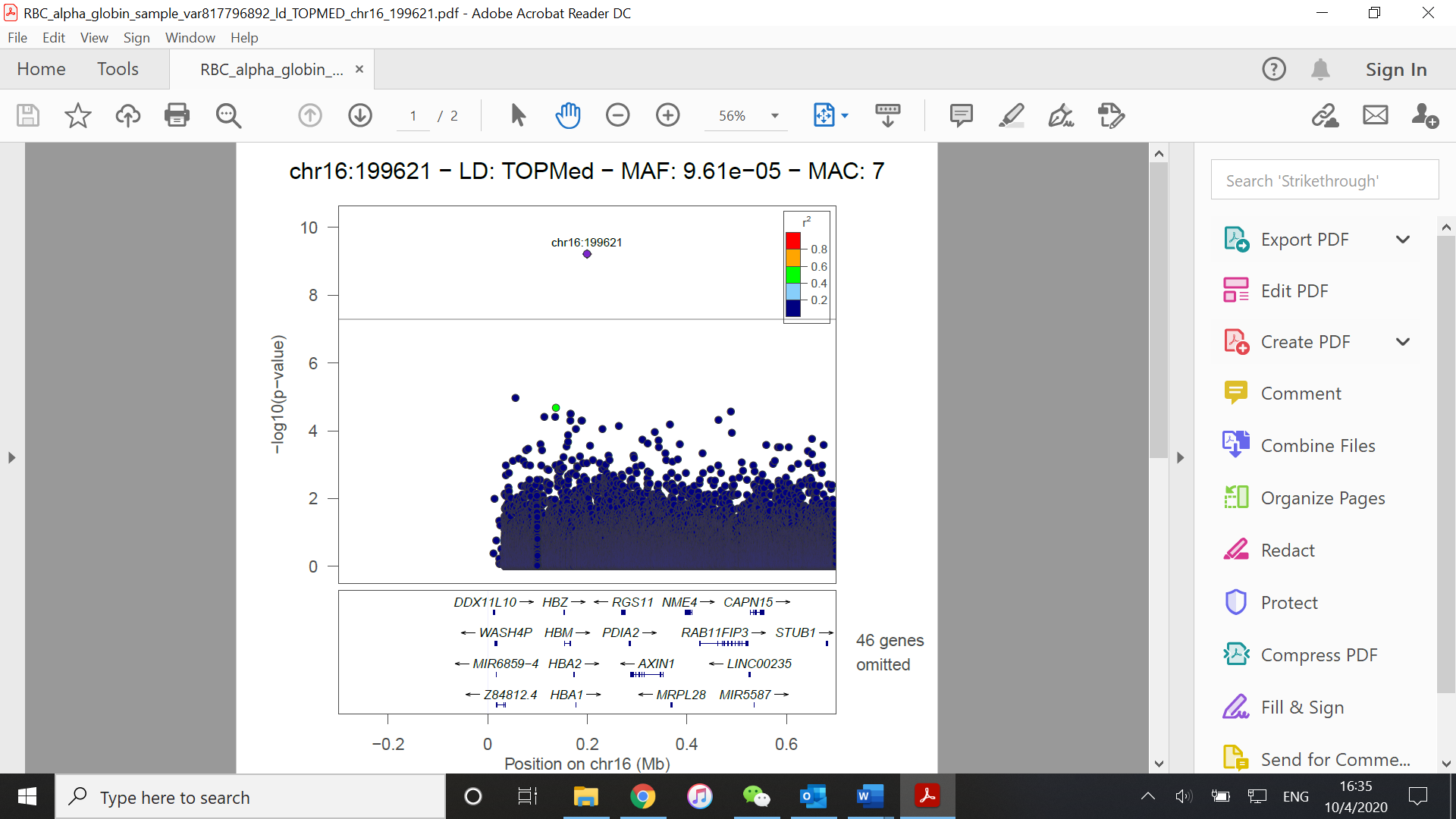

(G)

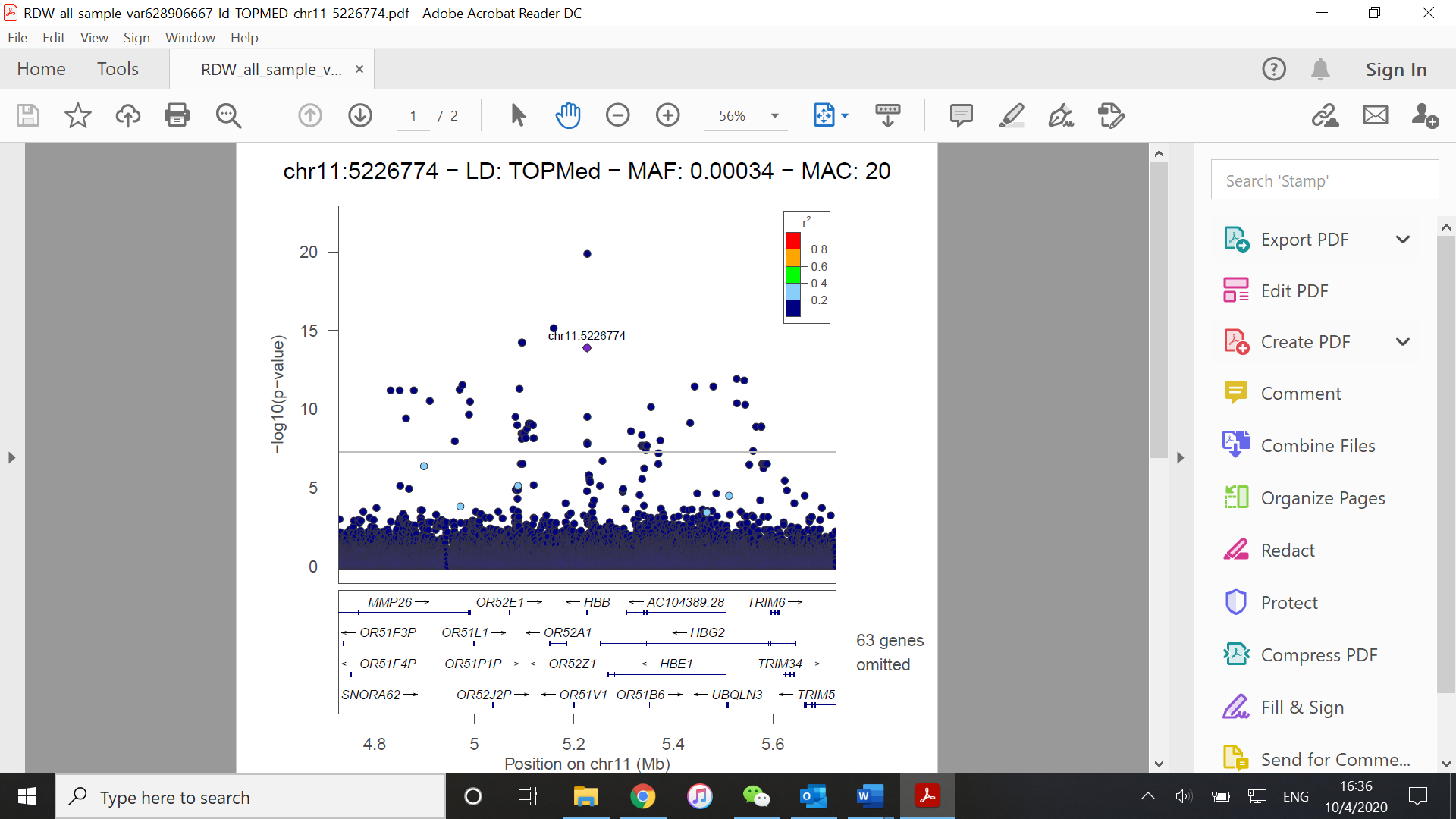

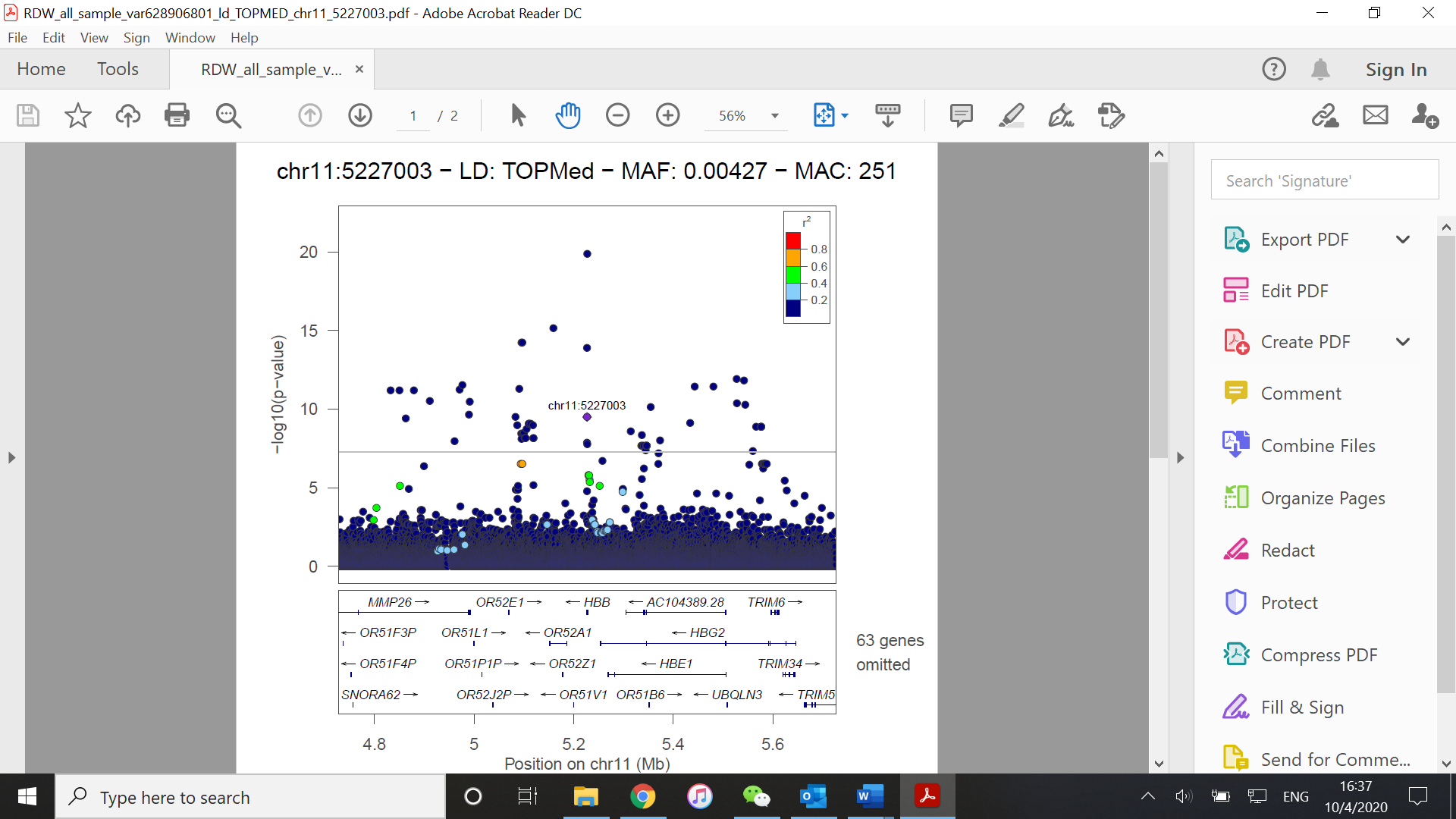

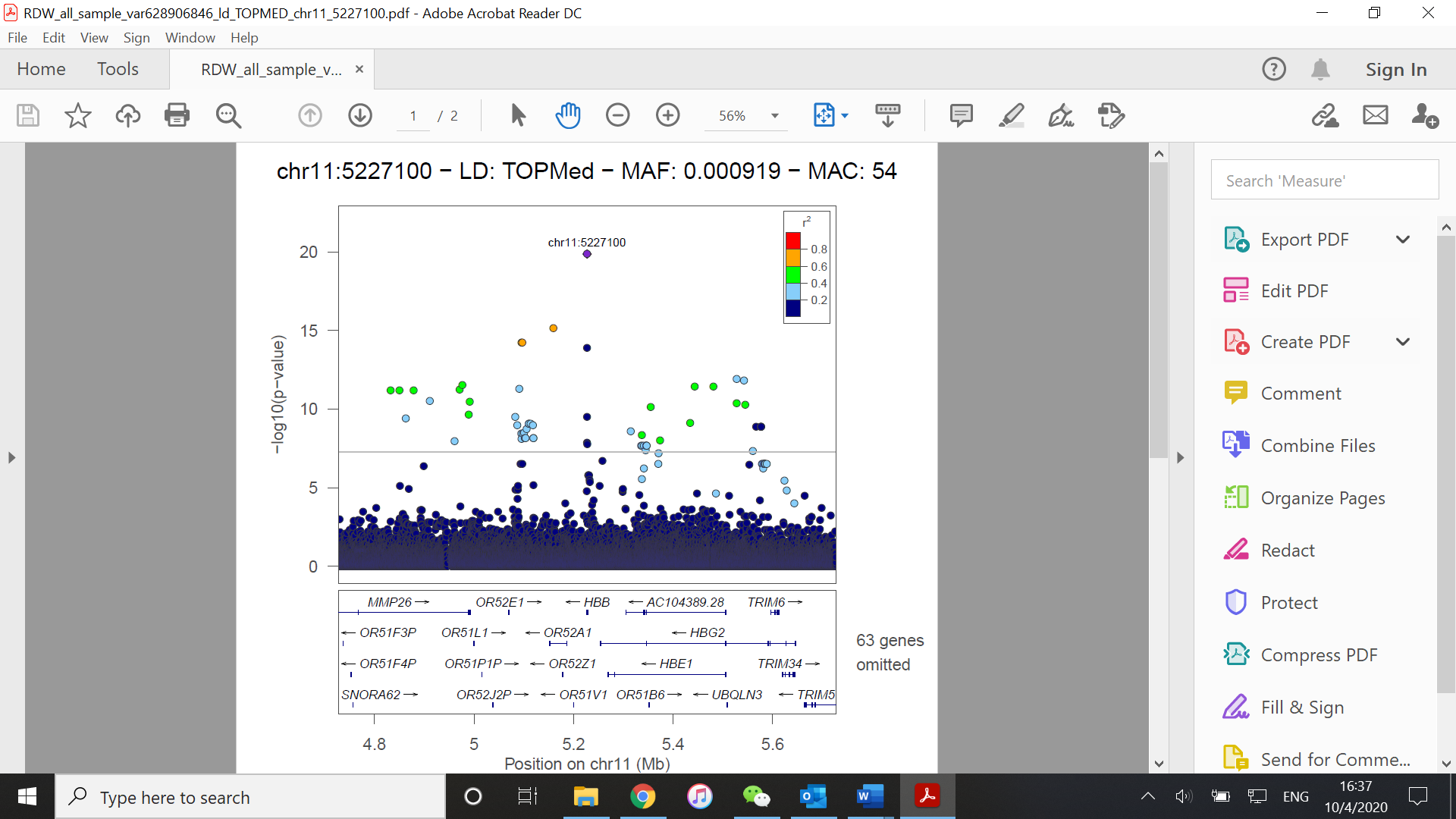

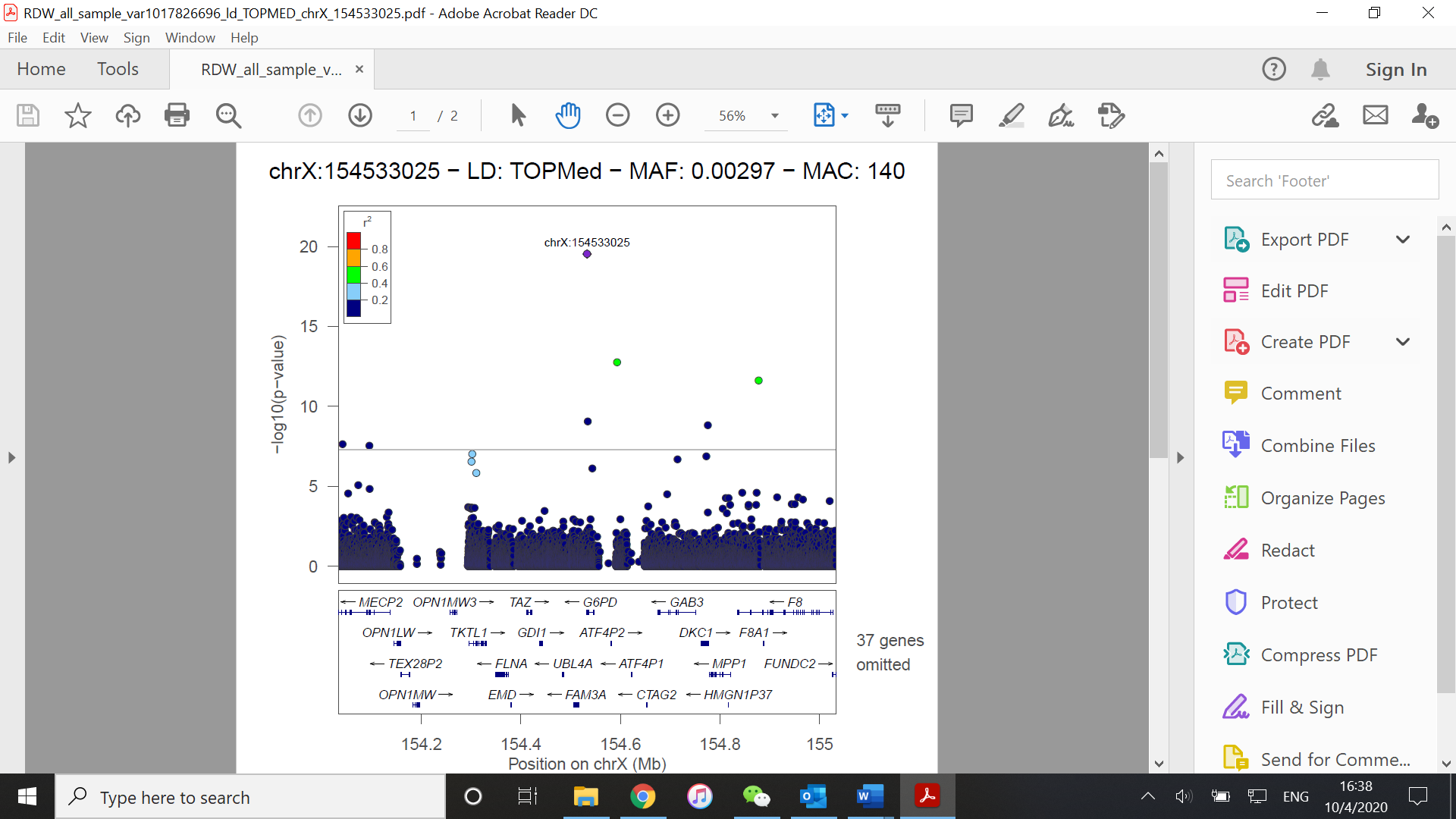

**Figure S5. Rare variants identified in the aggregated analysis in TOPMed. (A) HCT; (B) HGB; (C) MCH; (D) MCHC; (E) MCV; (F) RBC; (G) RDW.**

(A)

(A1)

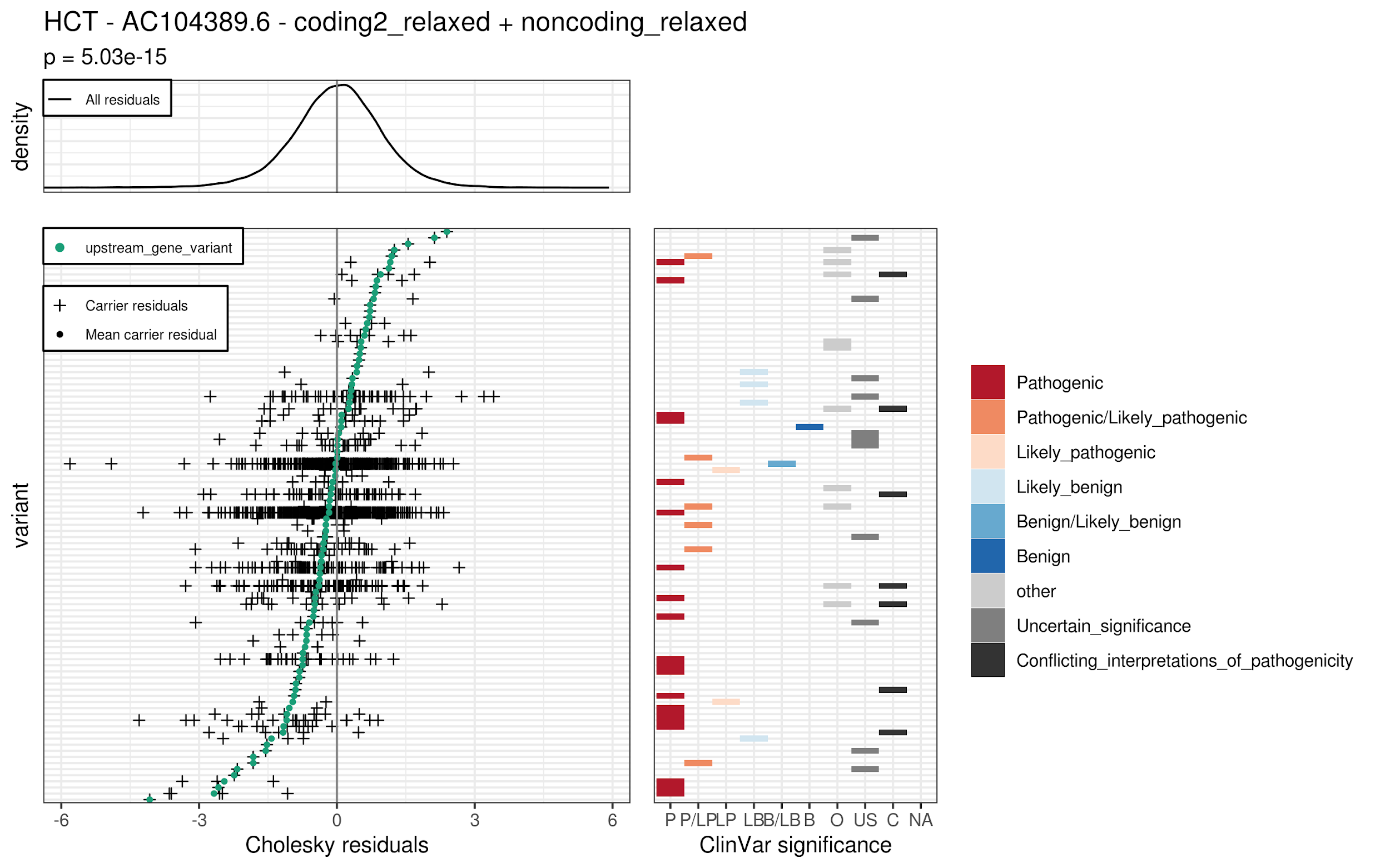

(A2)

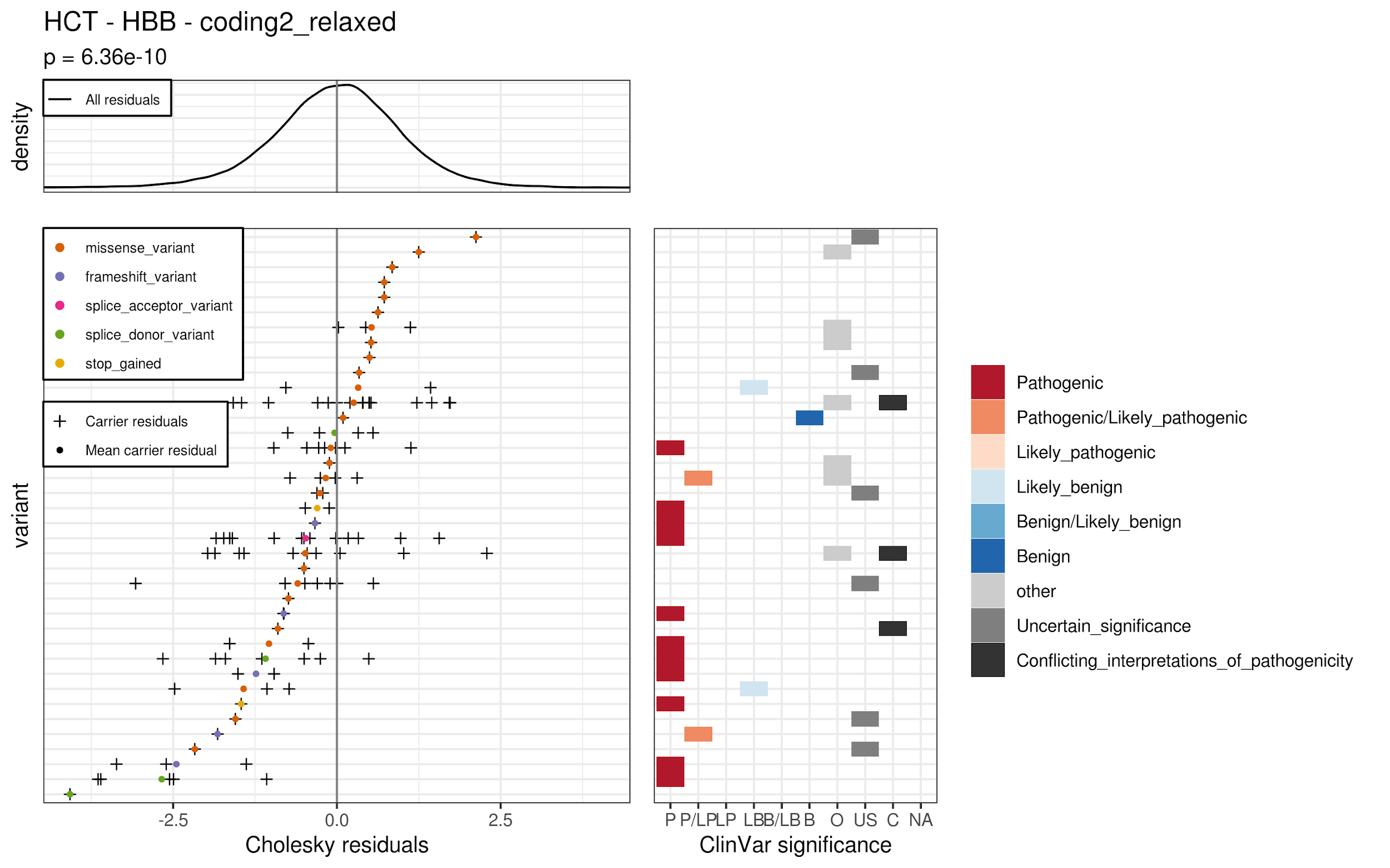

(A3)

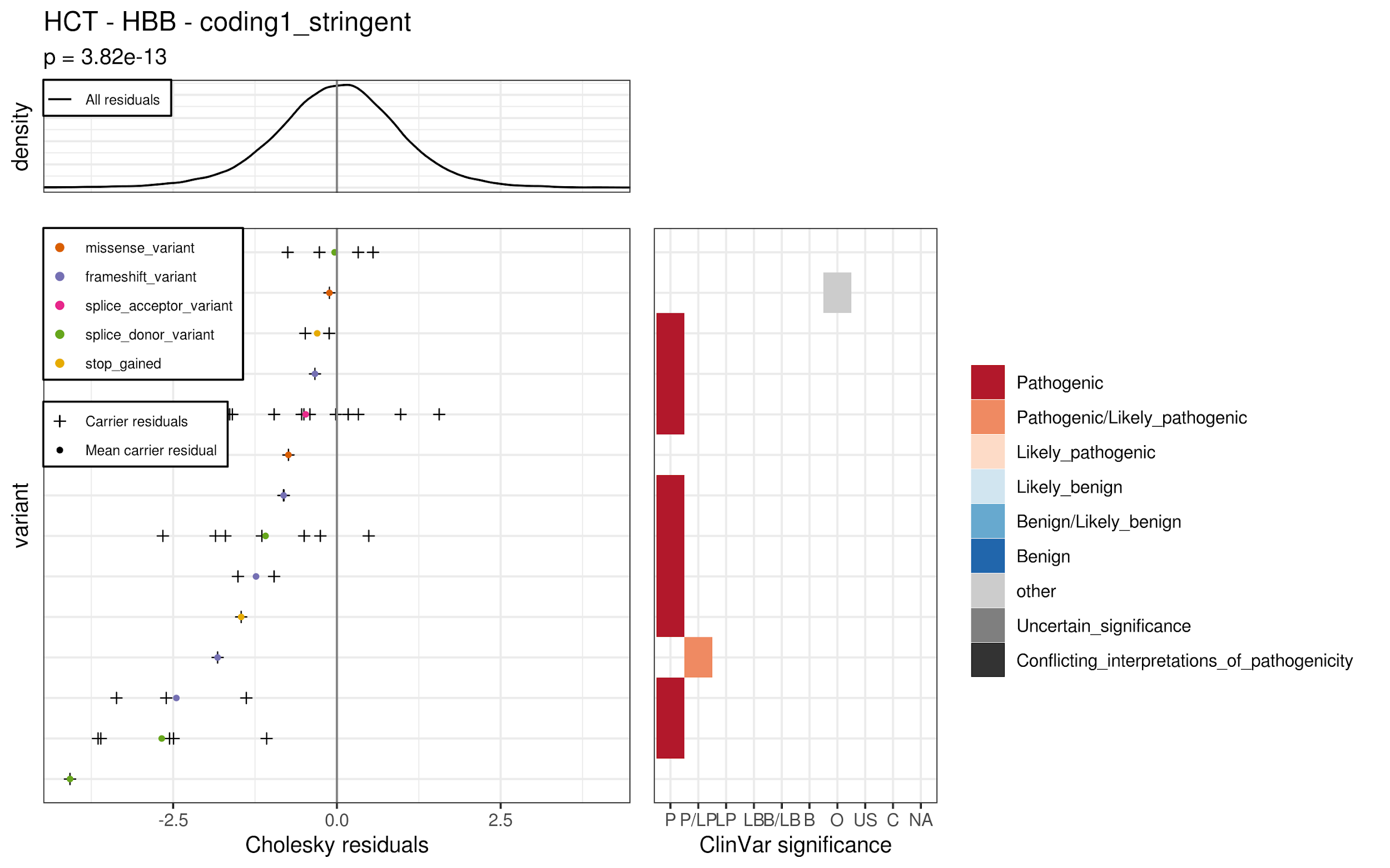

(A4)

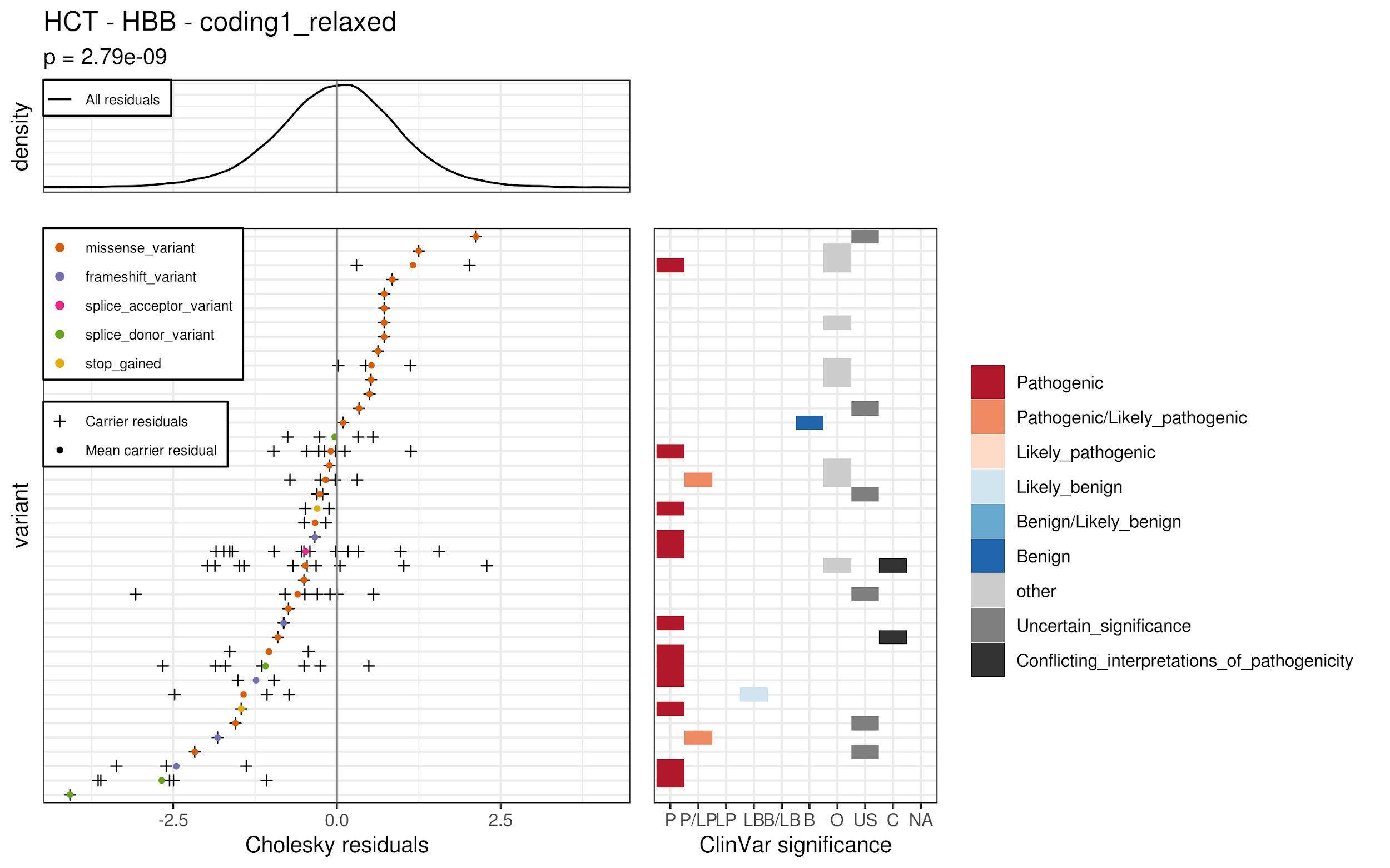

(A5)

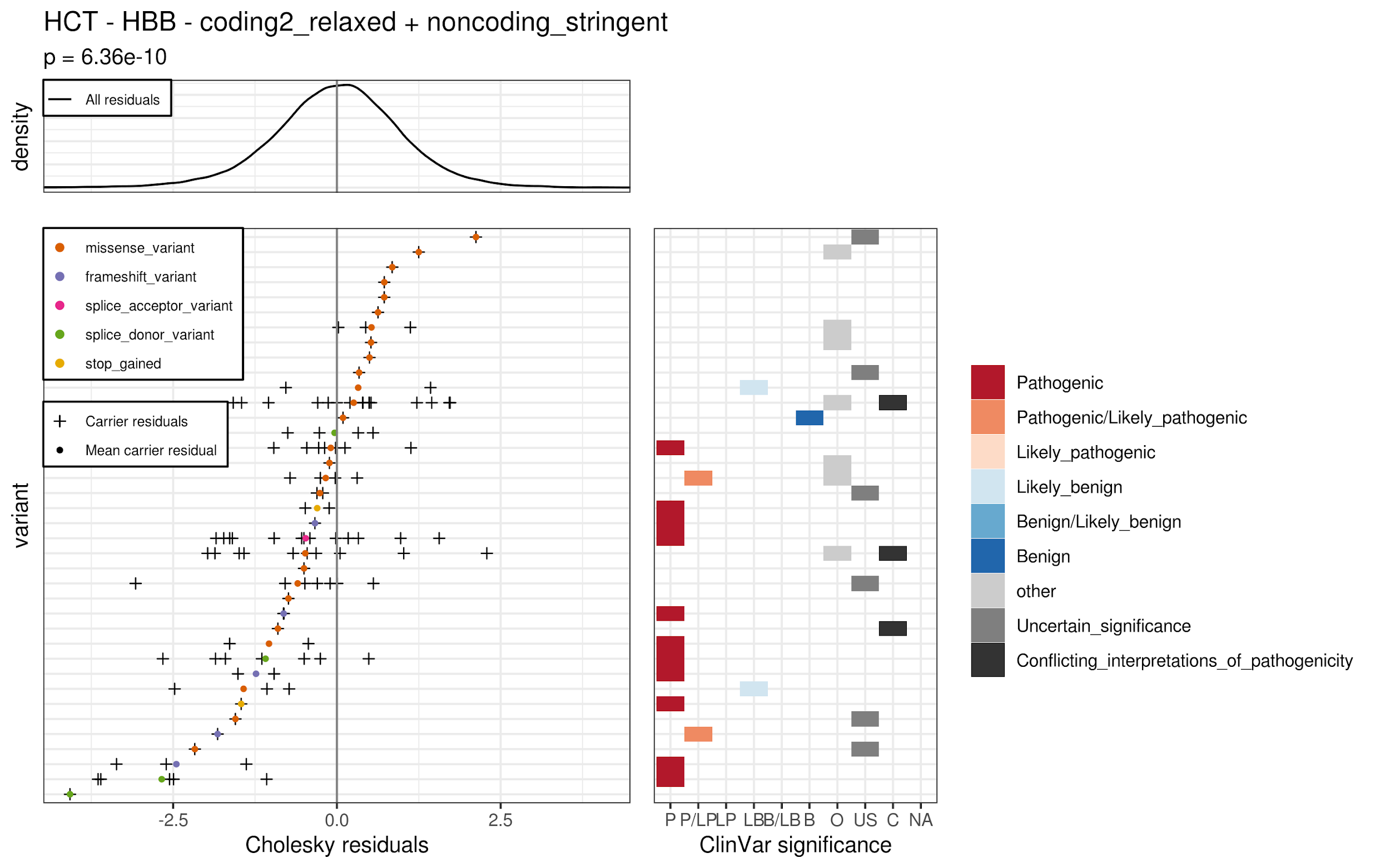

(B)

(B1)

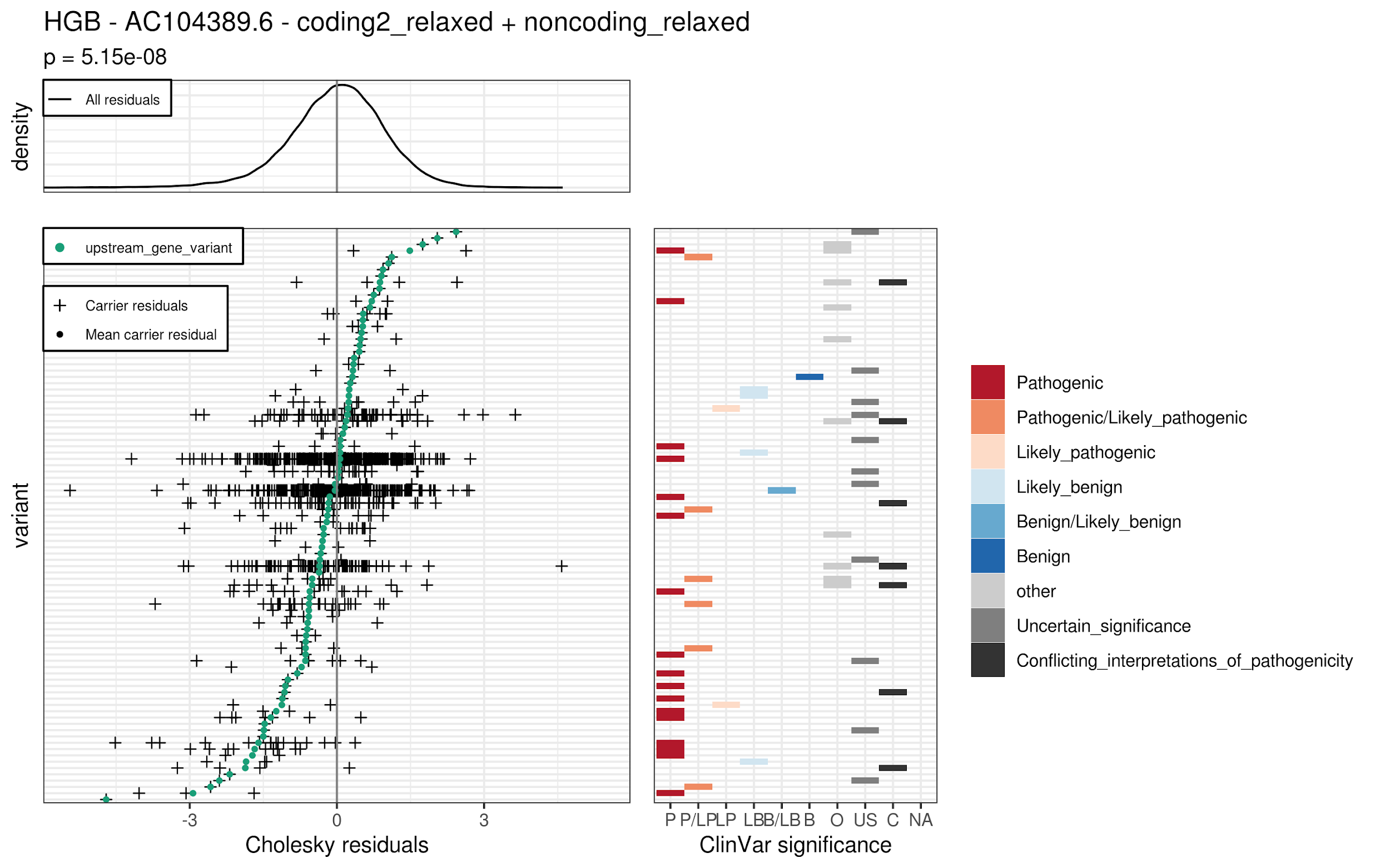

(B2)

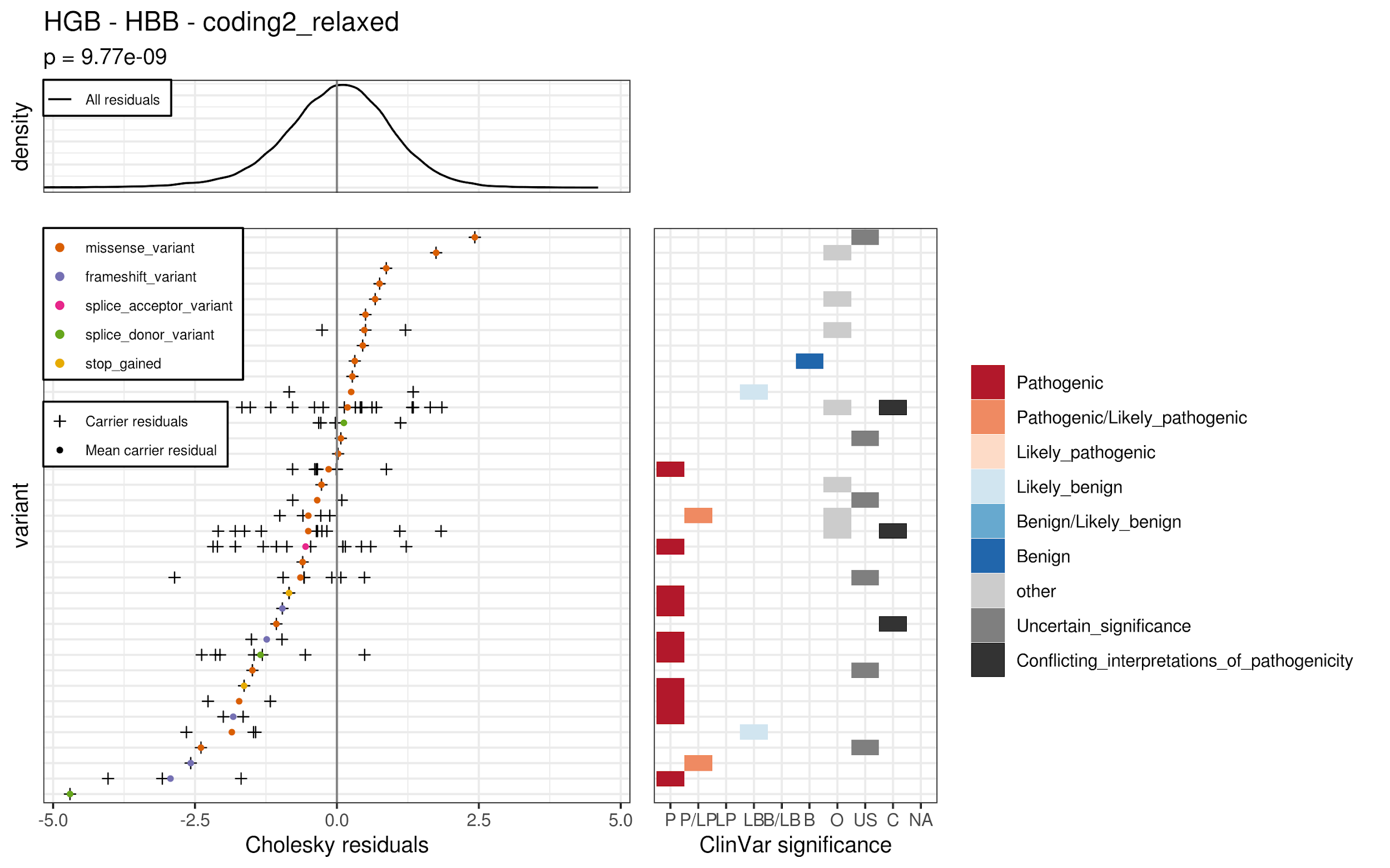

(B3)

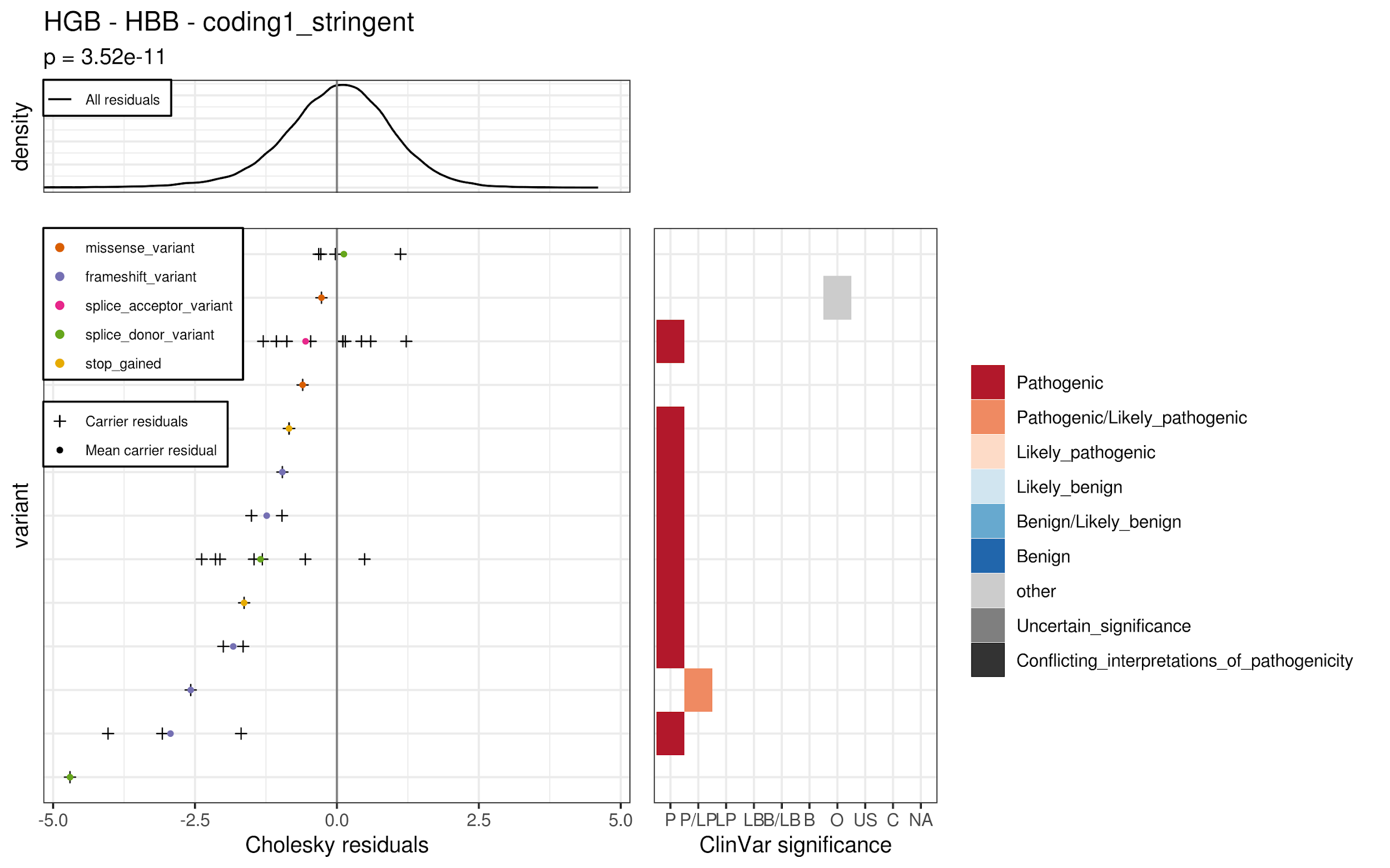

(B4)

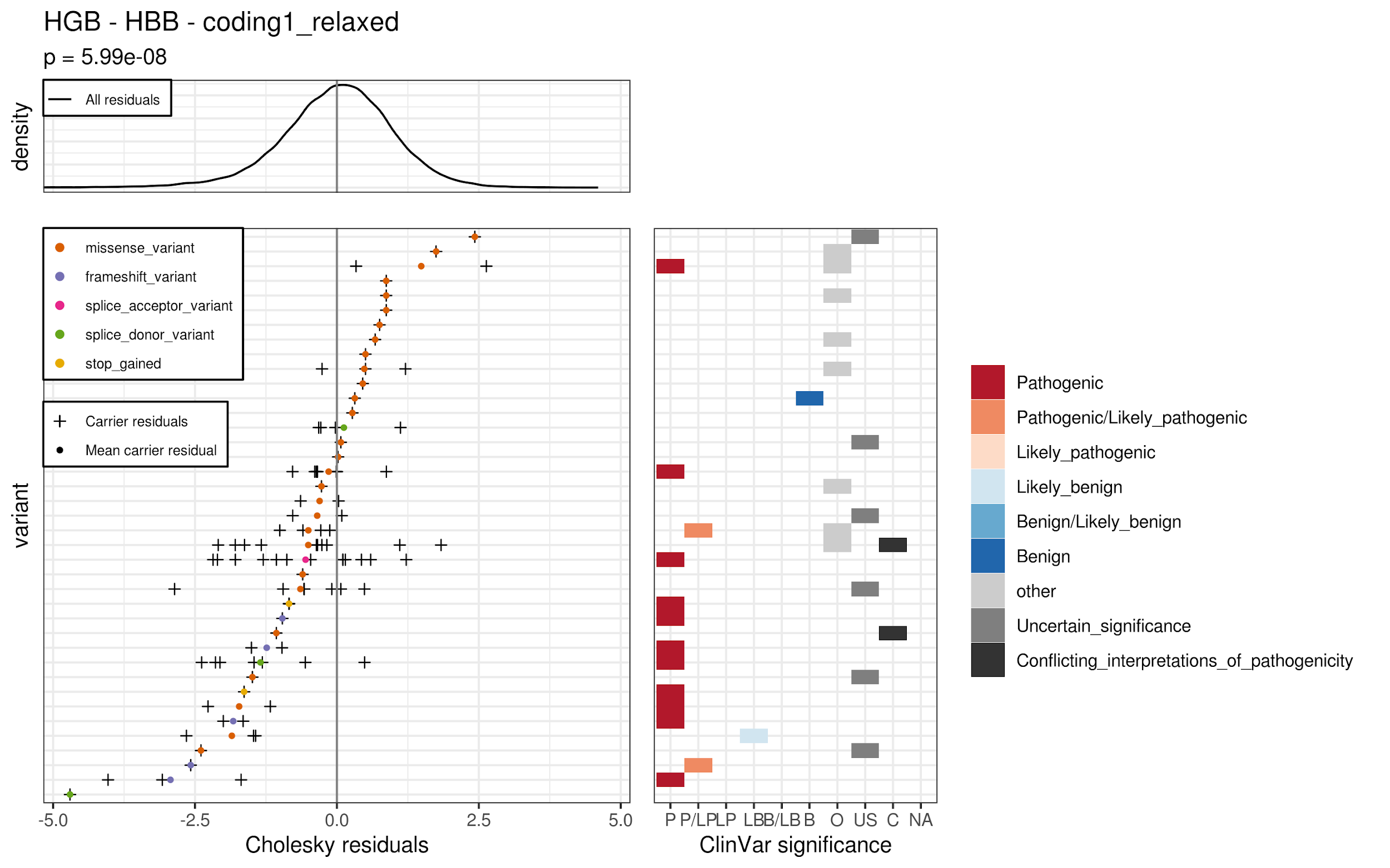

(B5)

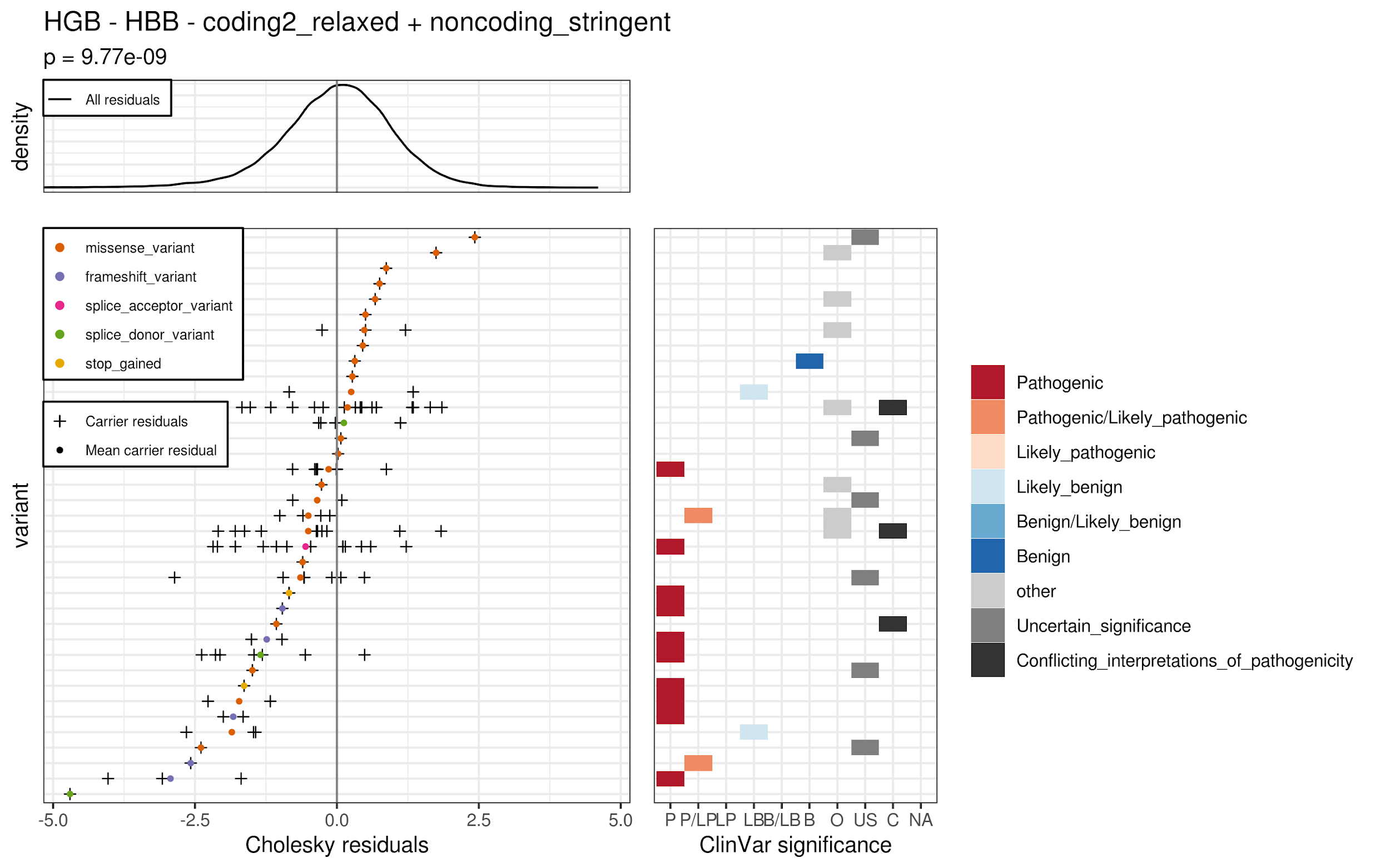

(C)

(C1)

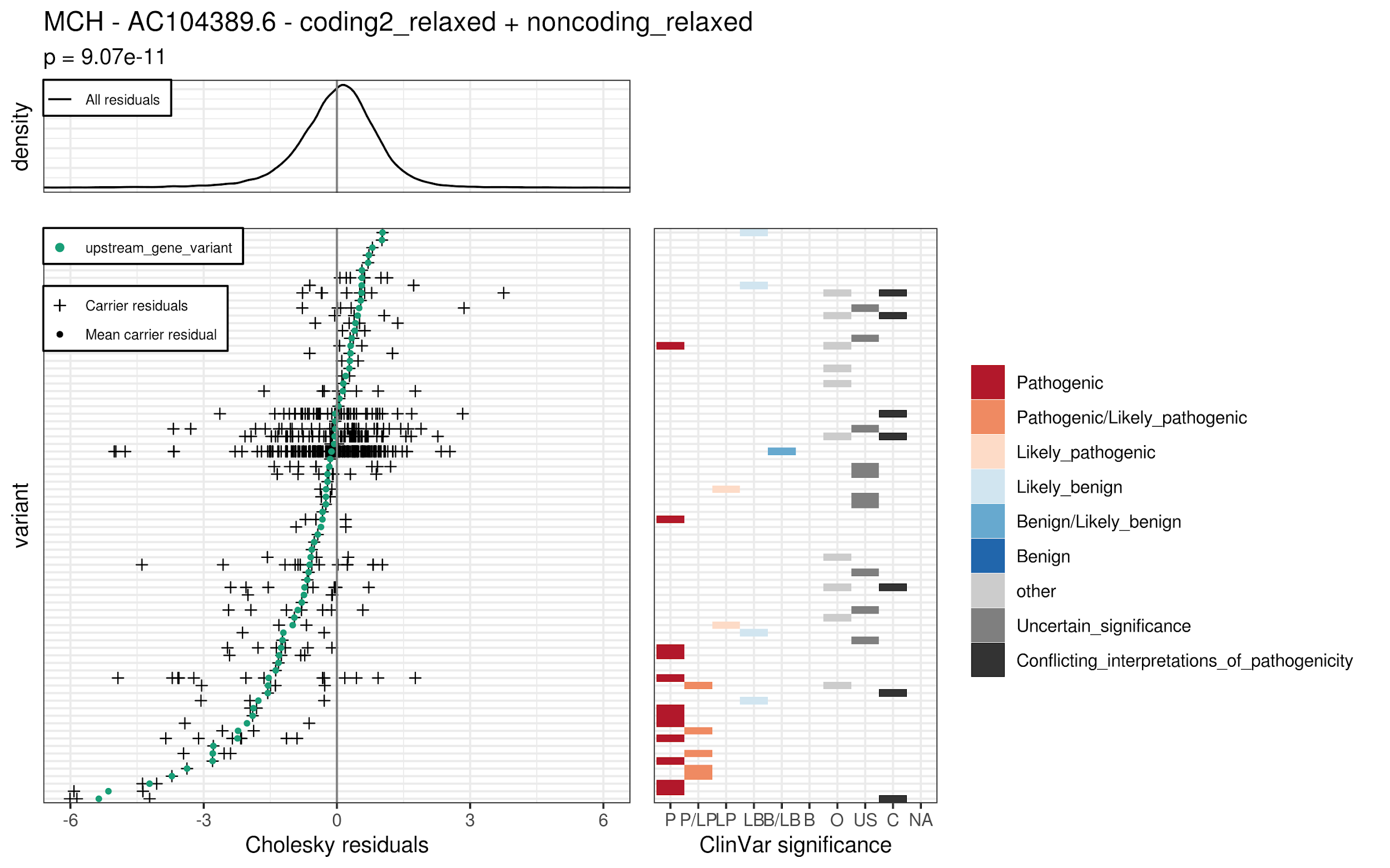

(C2)

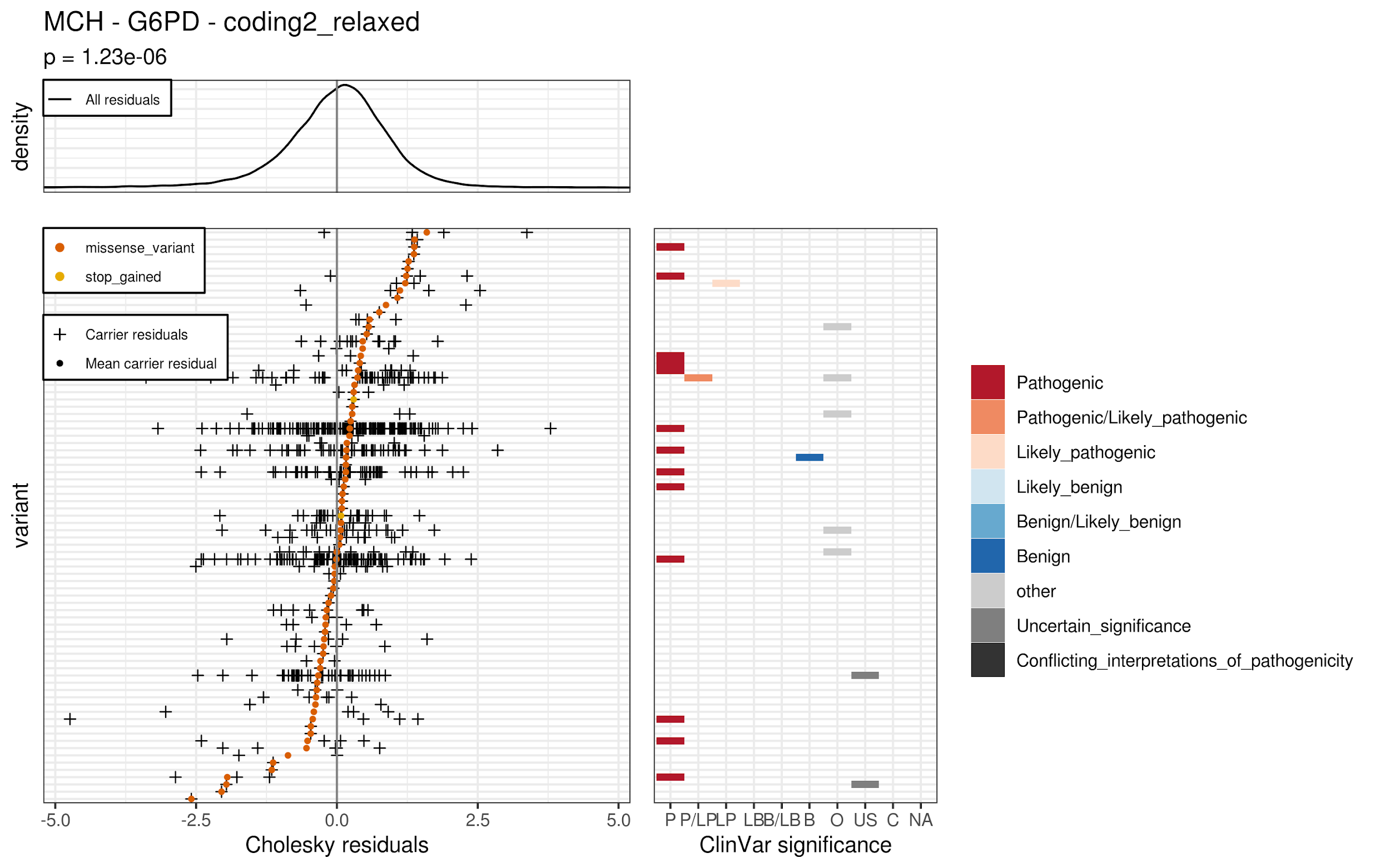

(C3)

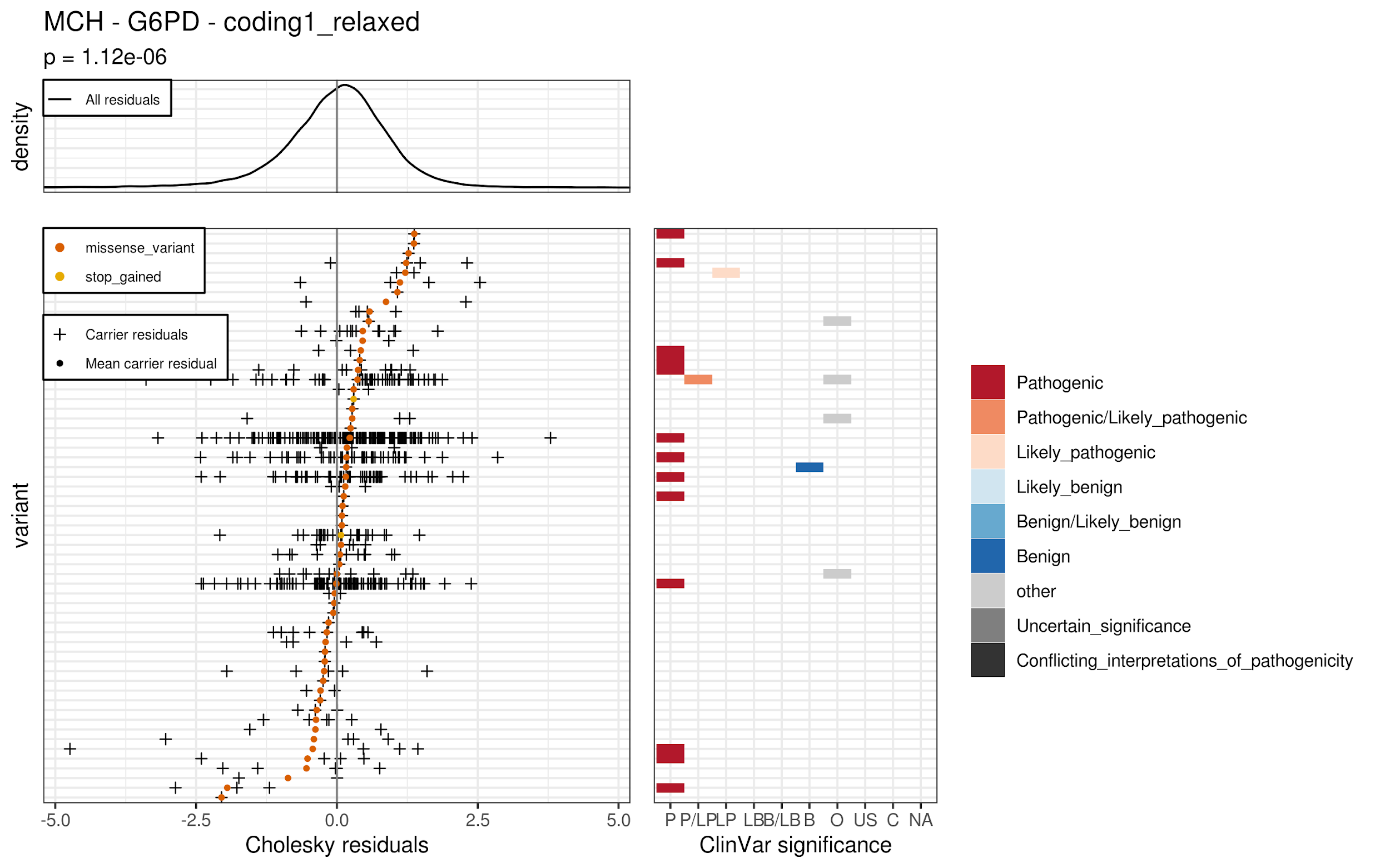

(C4)

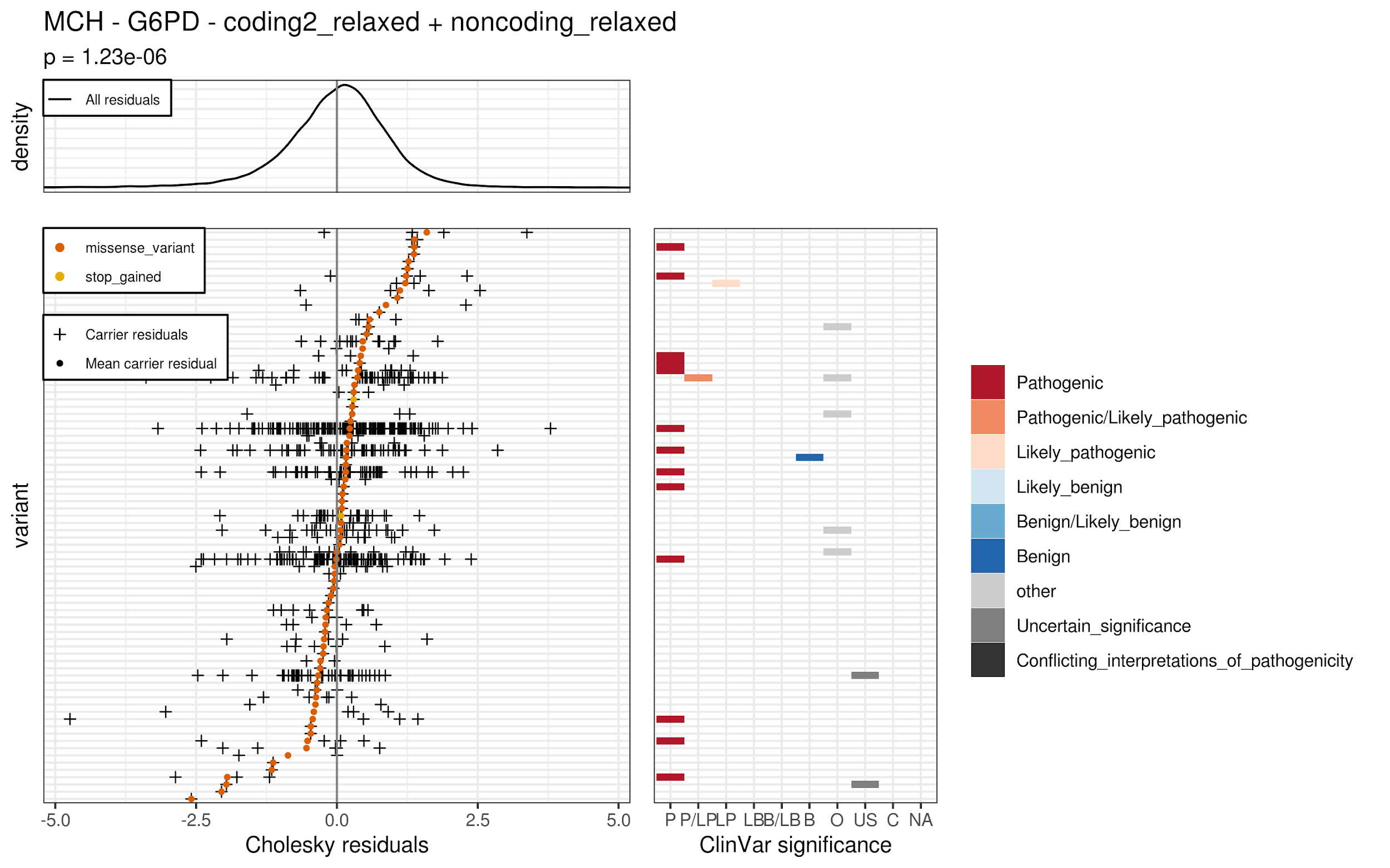

(C5)

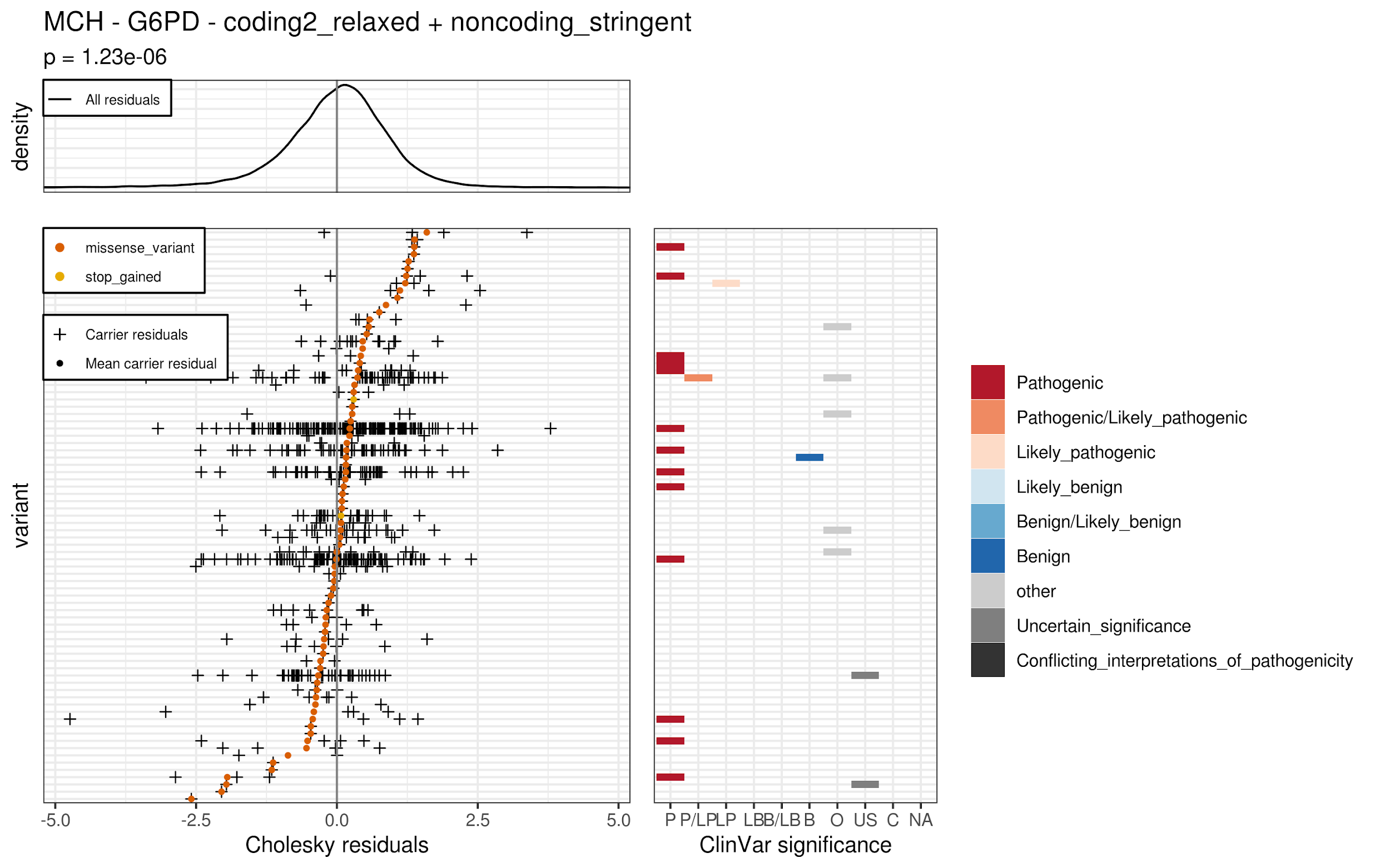

(C6)

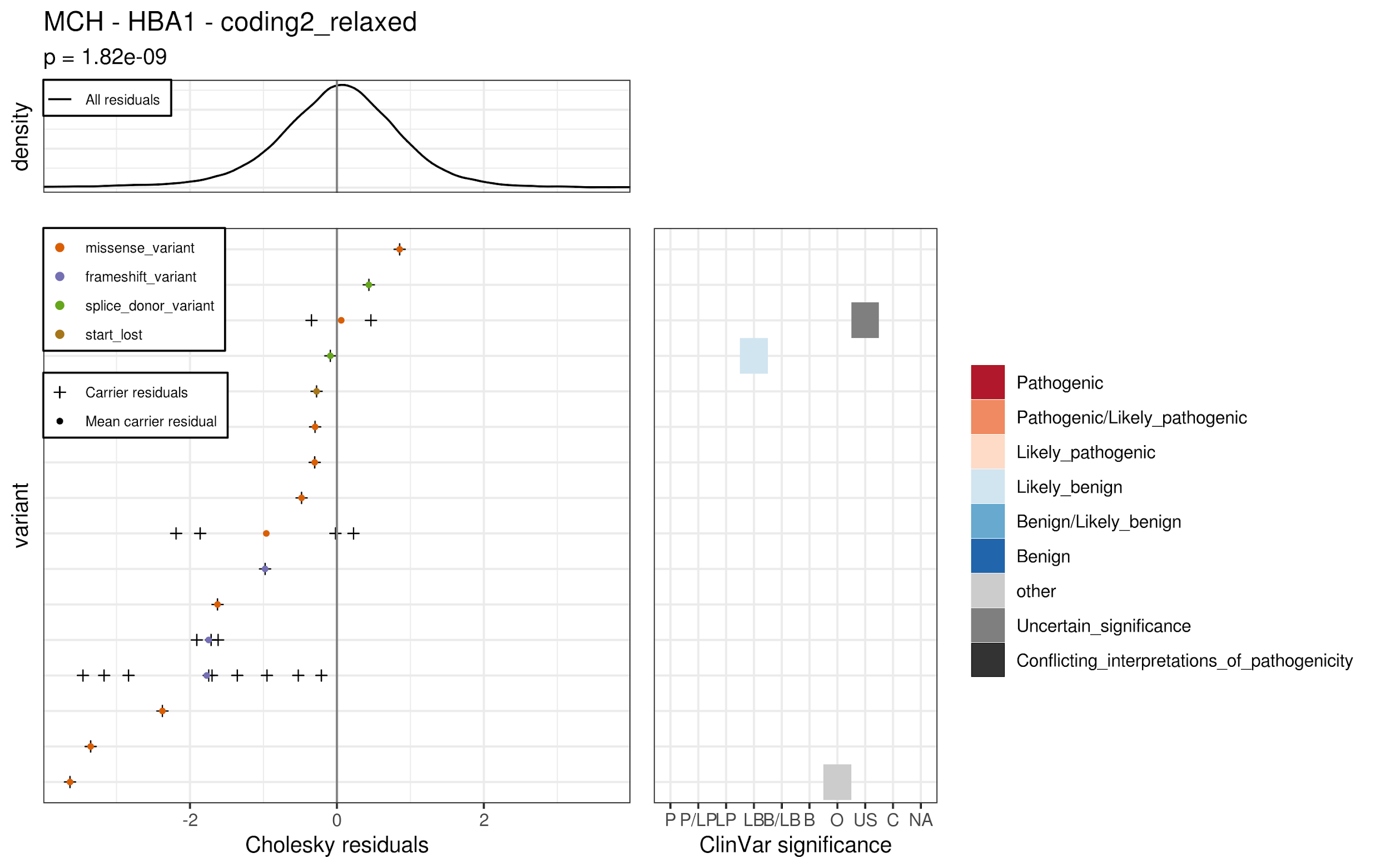
(C7)
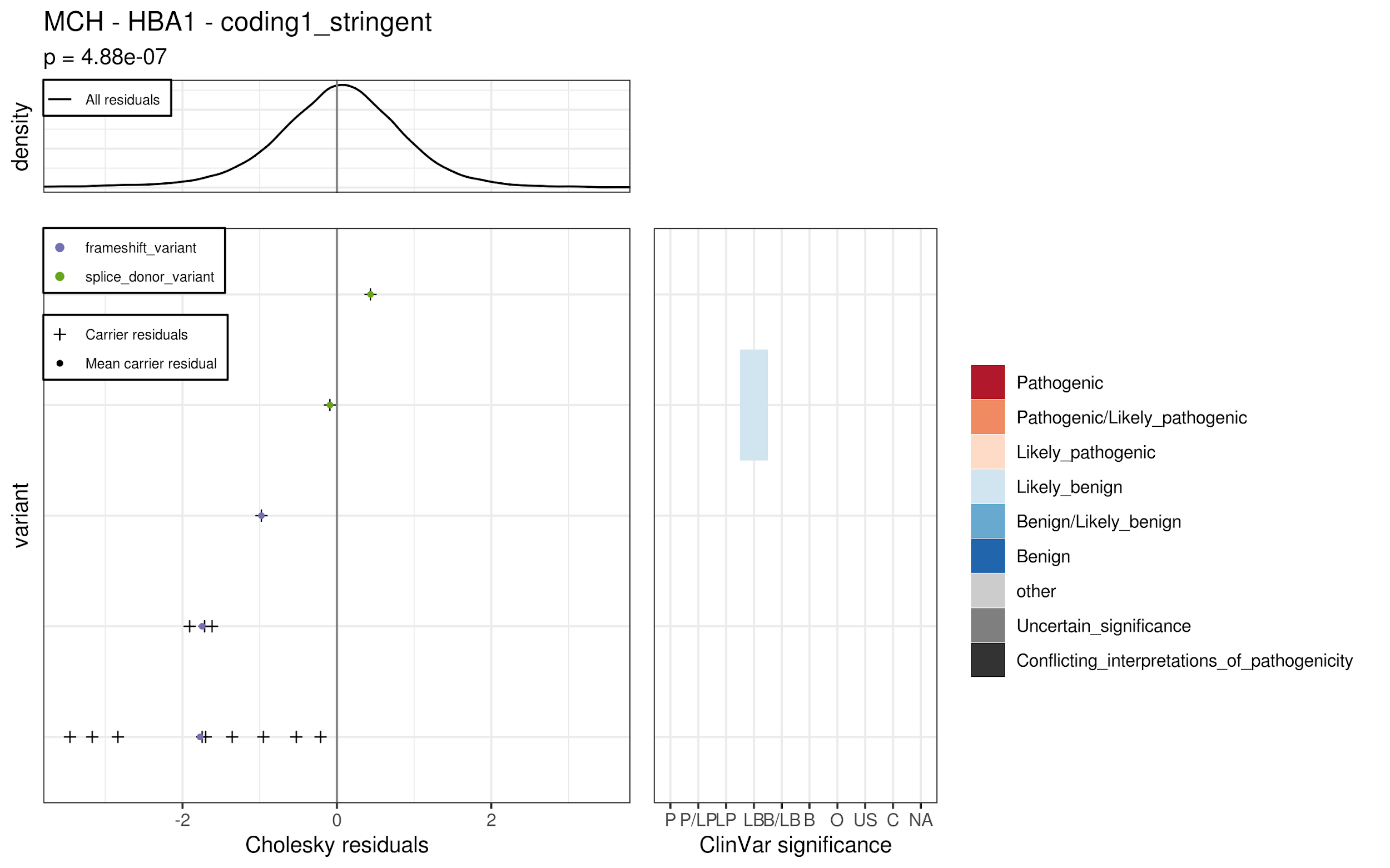
(C8)
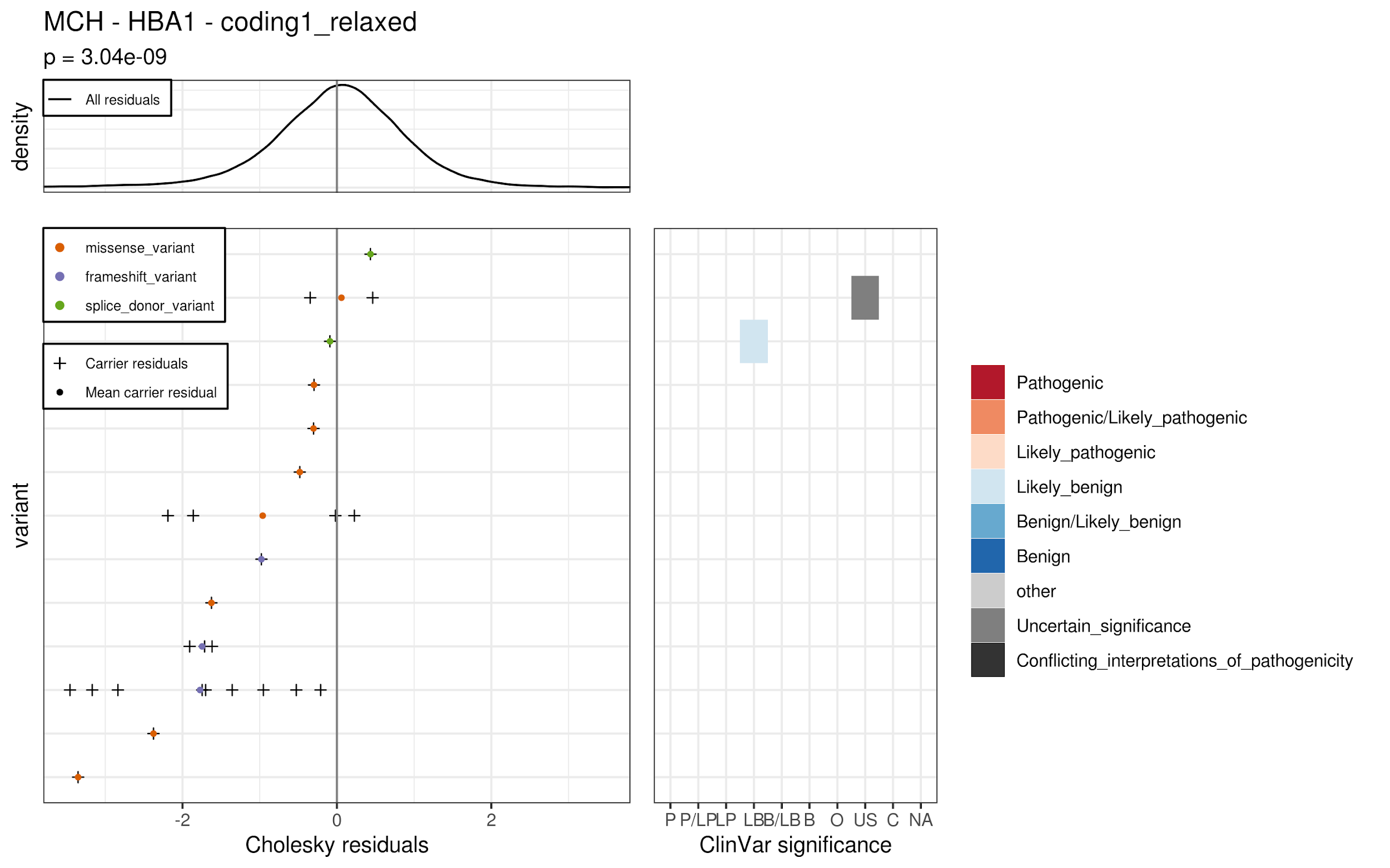
(C9)
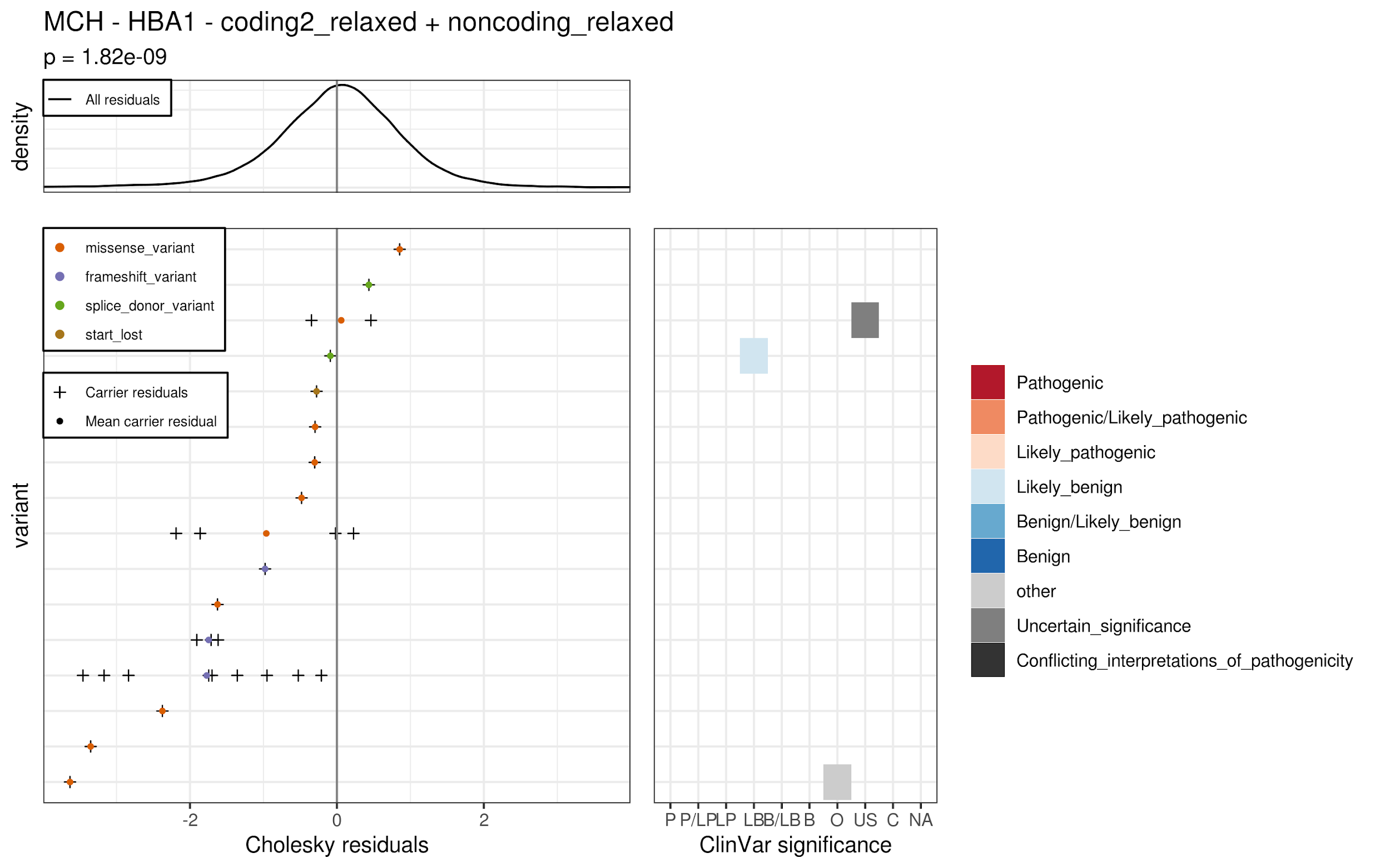
(C10)
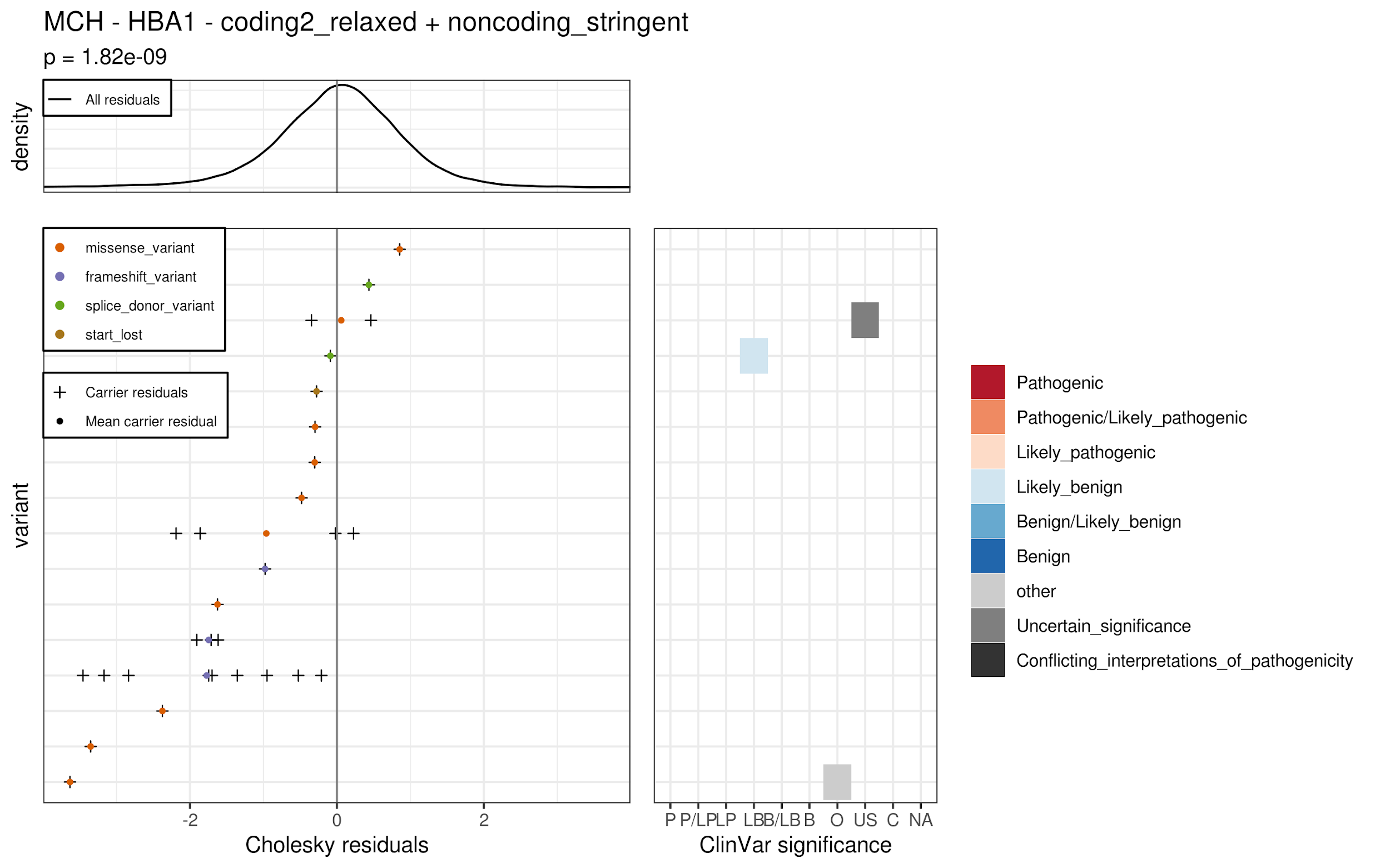
(C11)
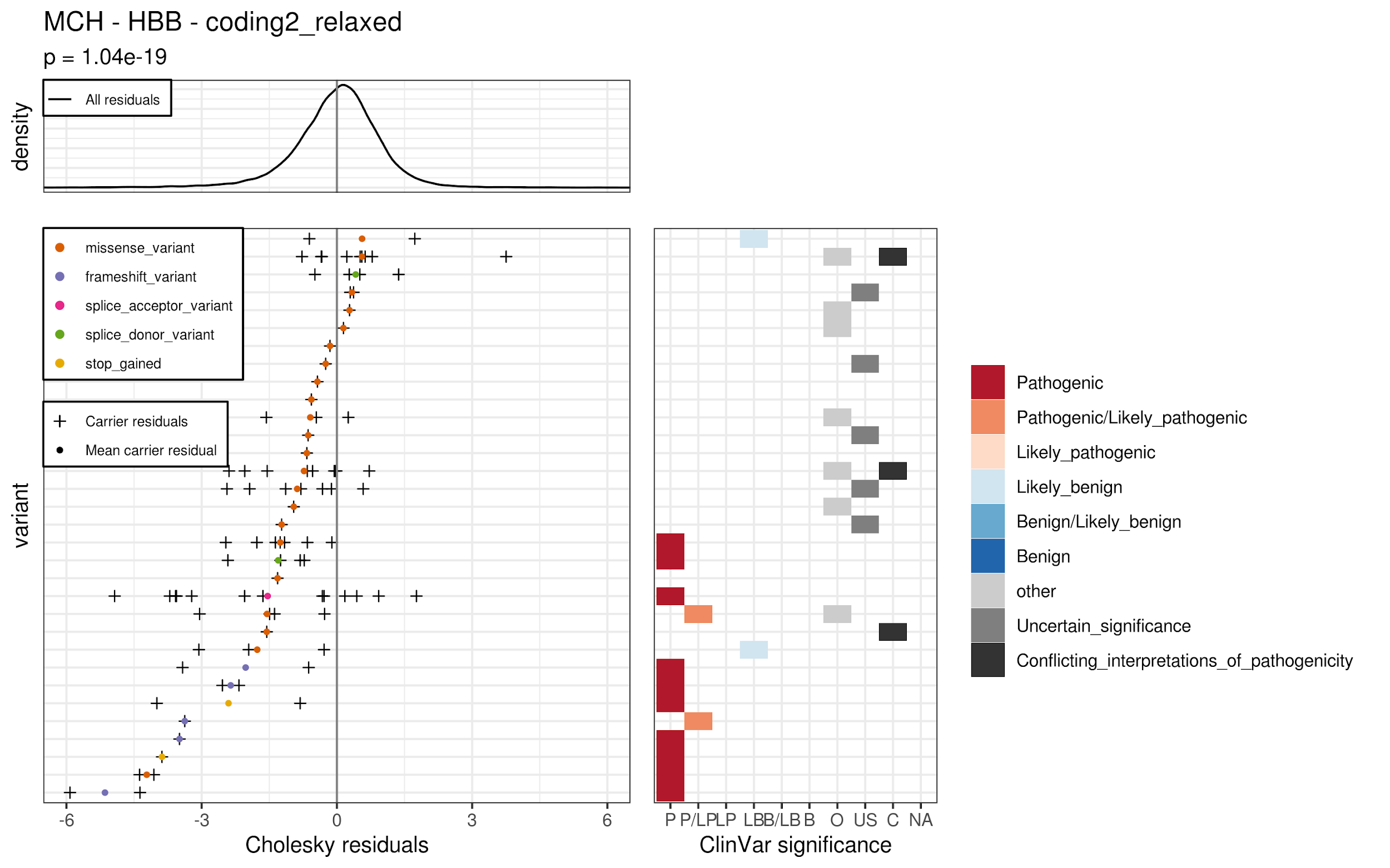
(C12)
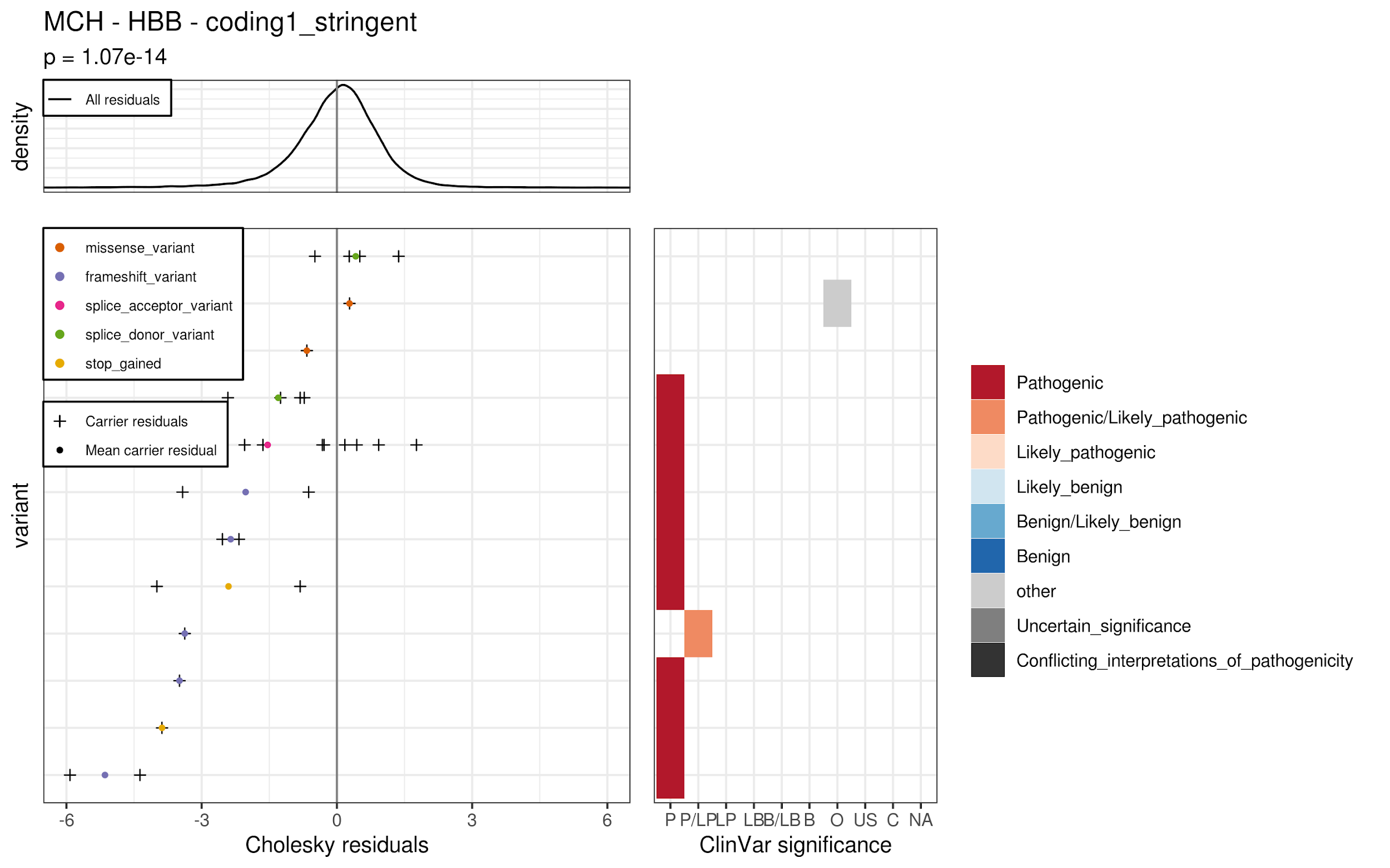
(C13)
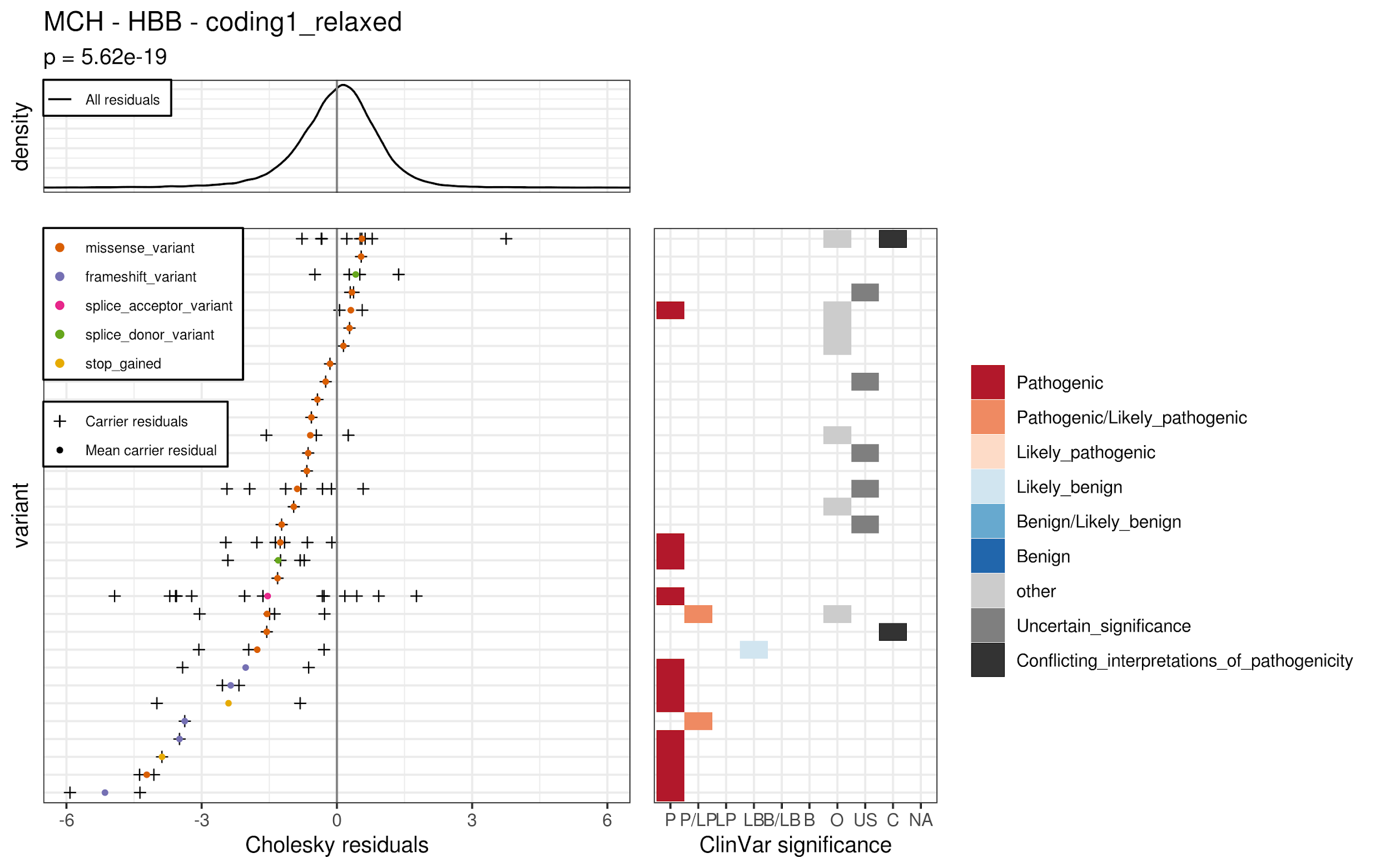
(C14)
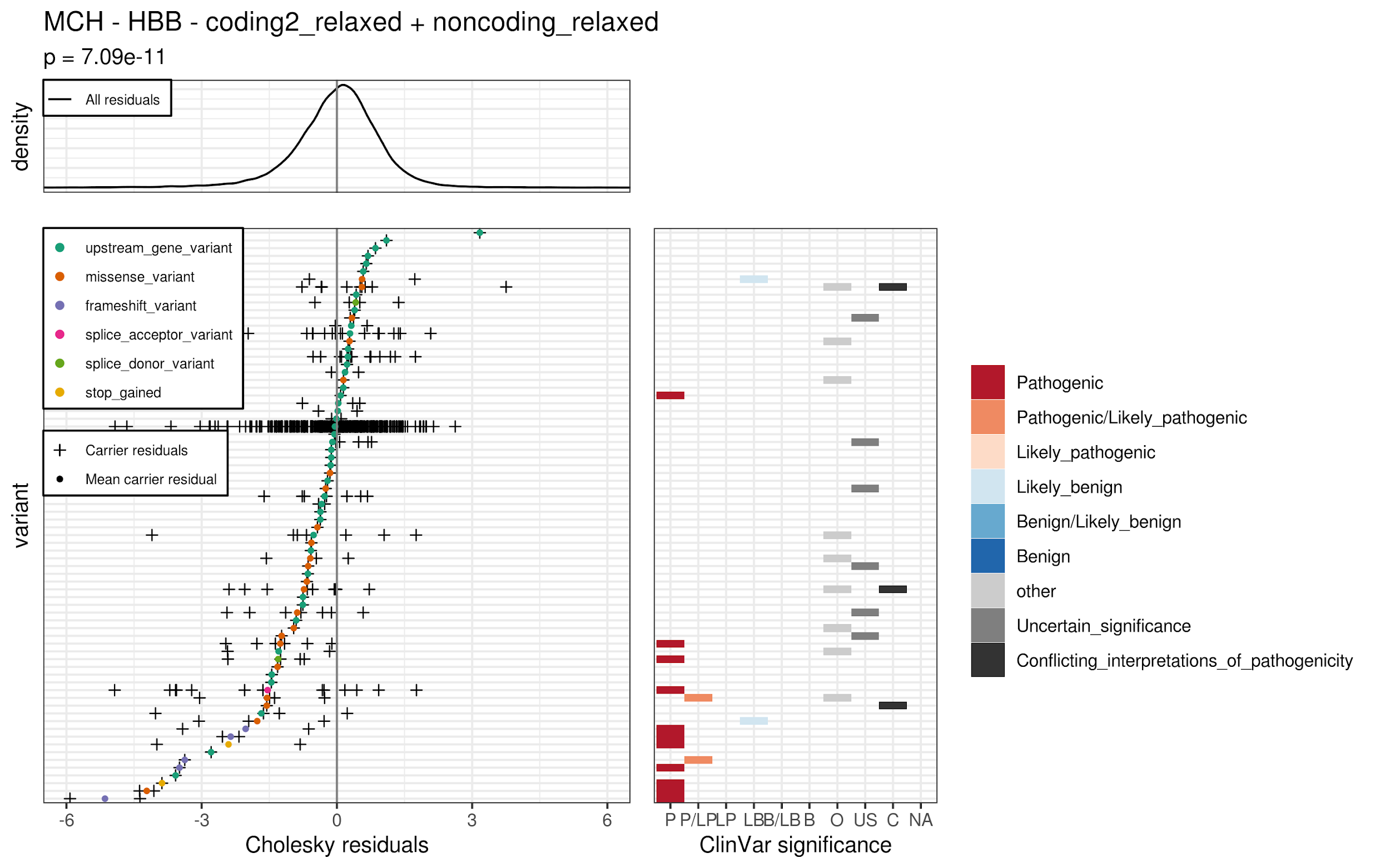
(C15)
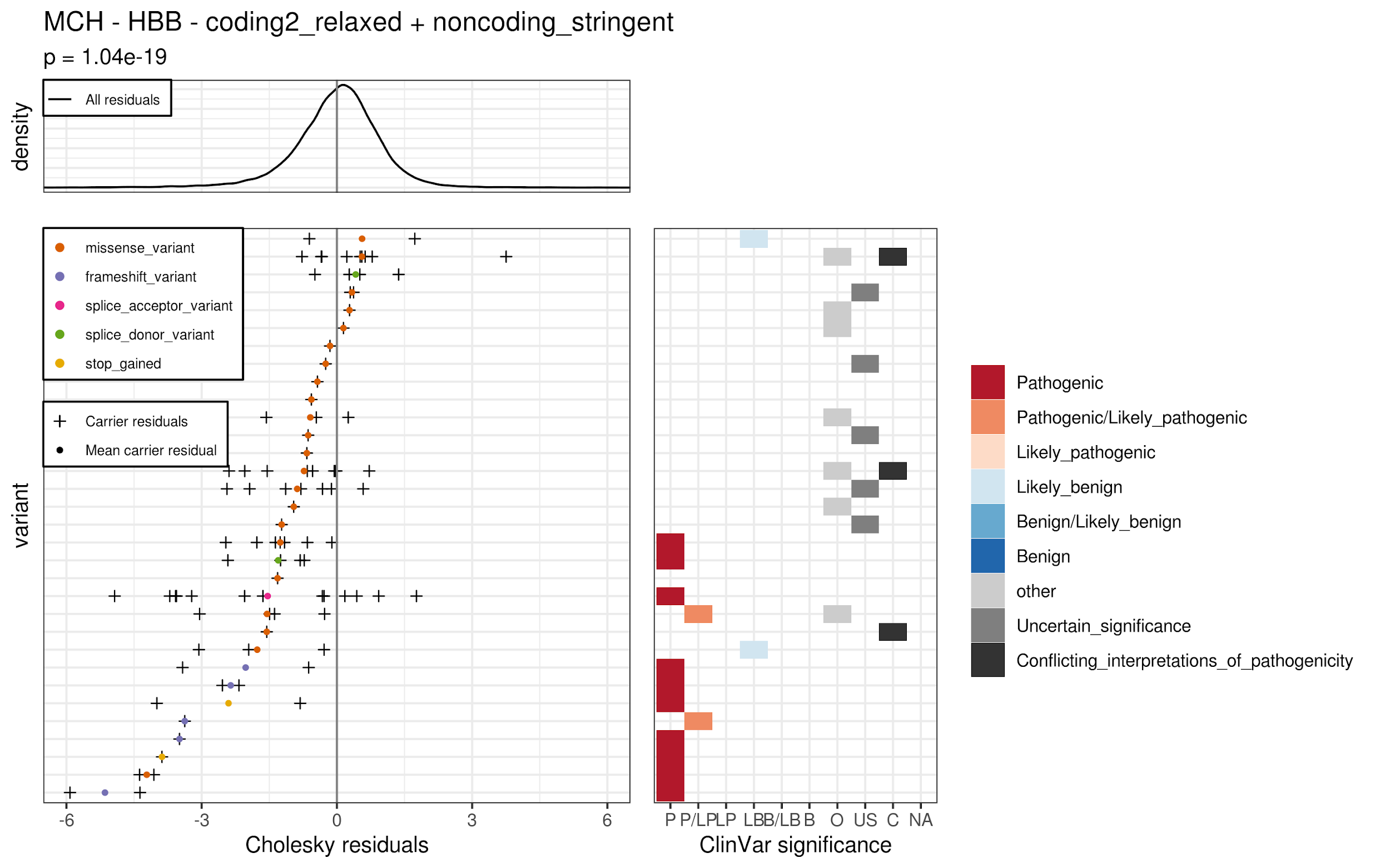
(C16)
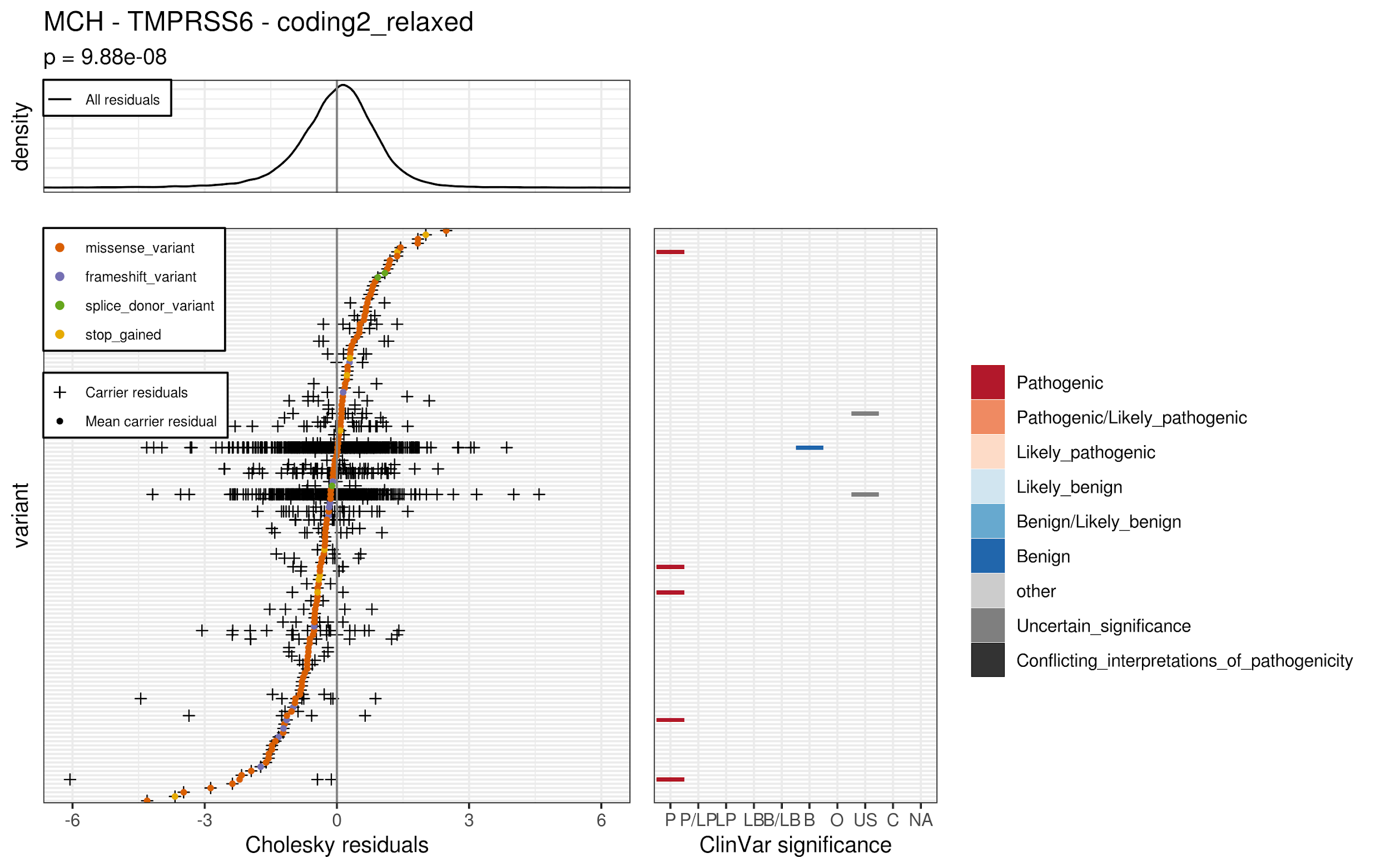
(C17)
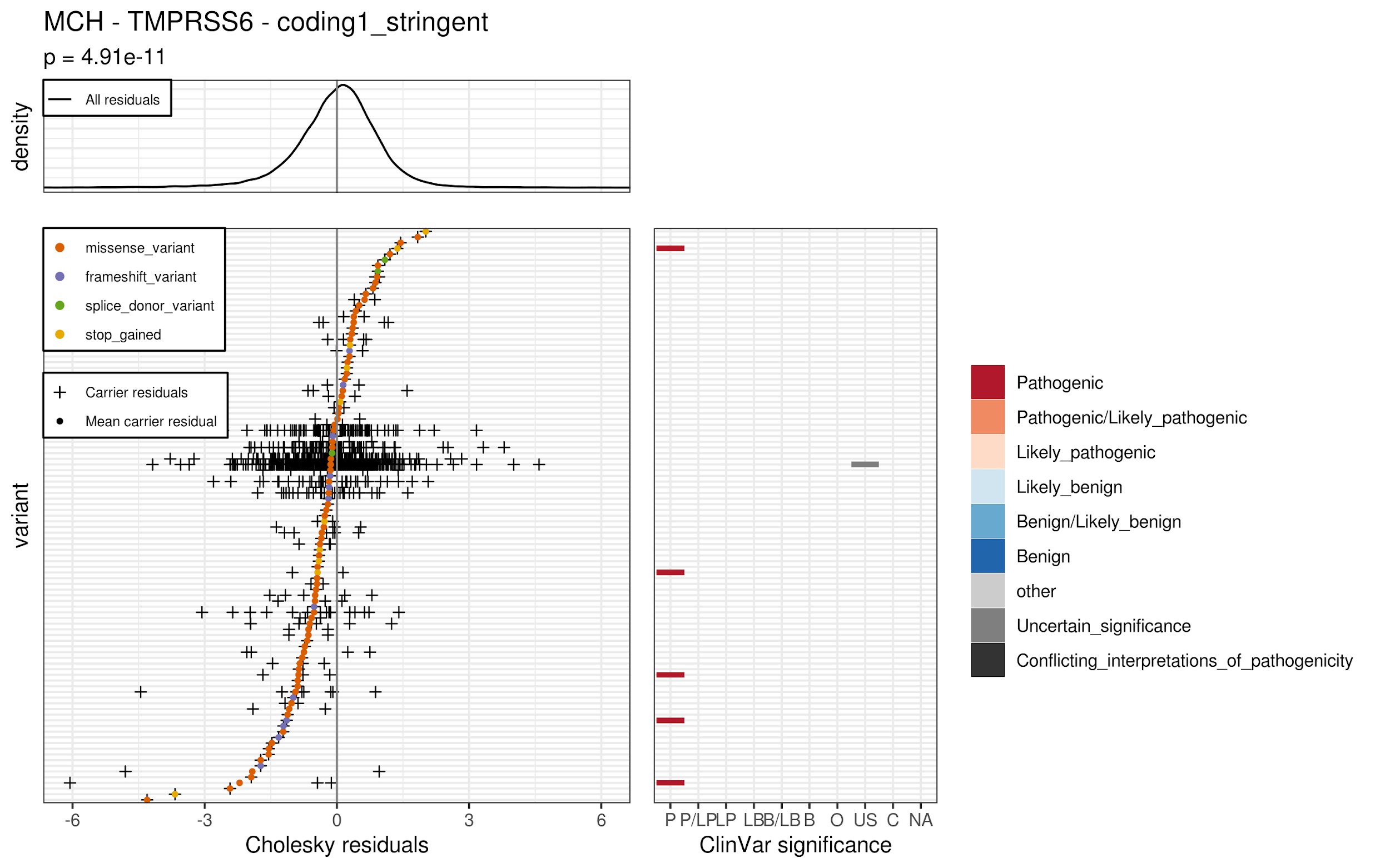
(C18)
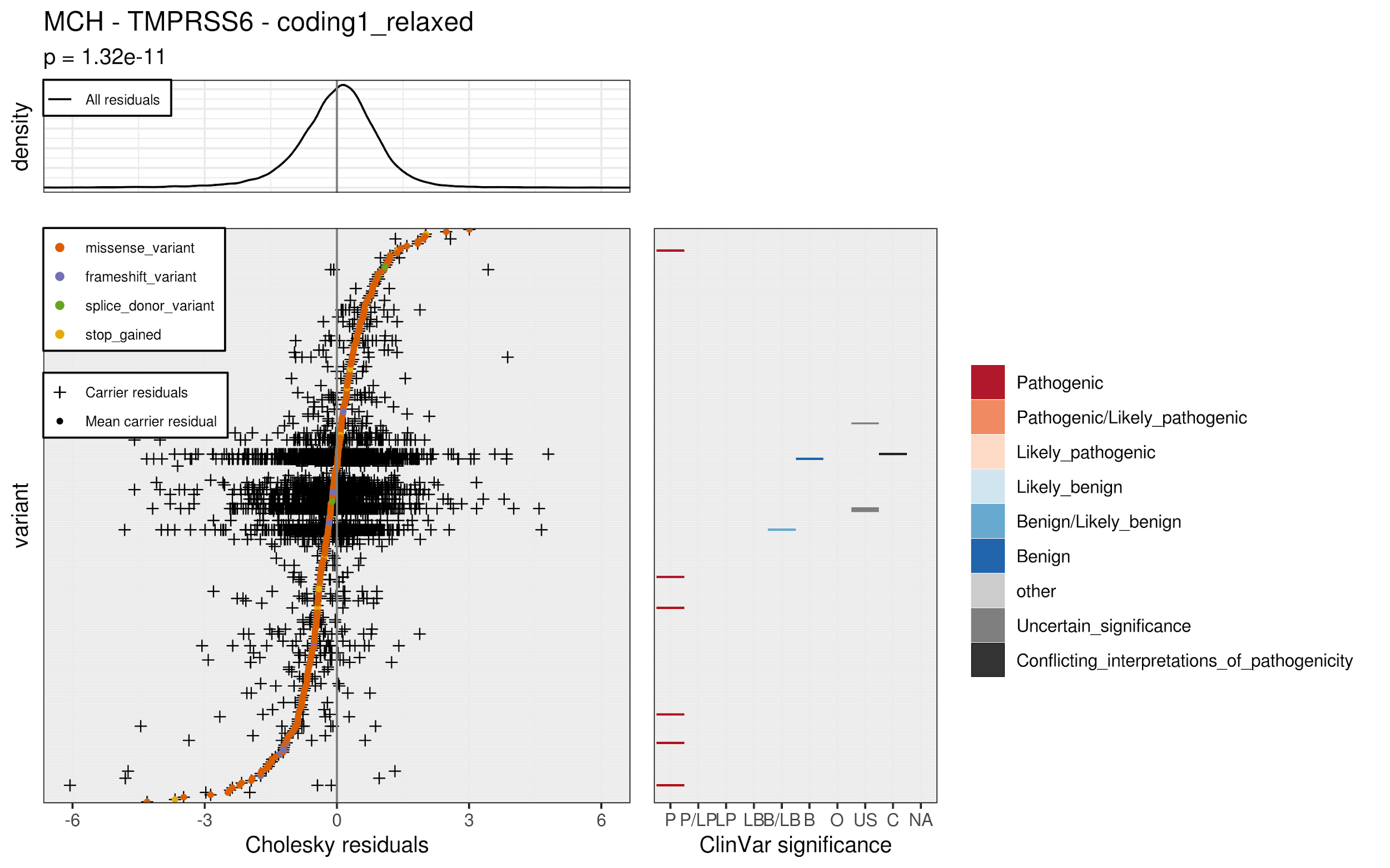
(C19)
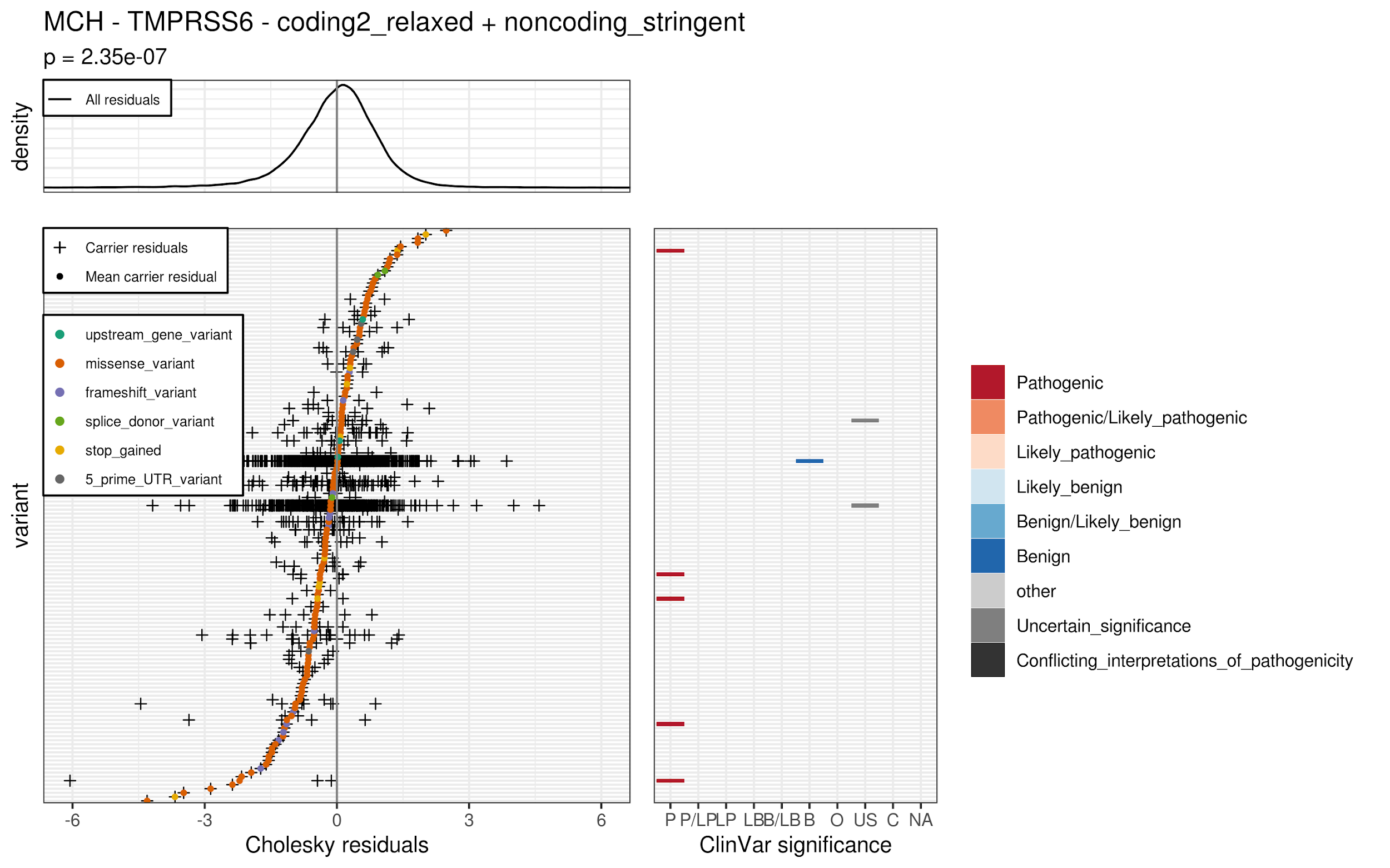

(D)

(D1)

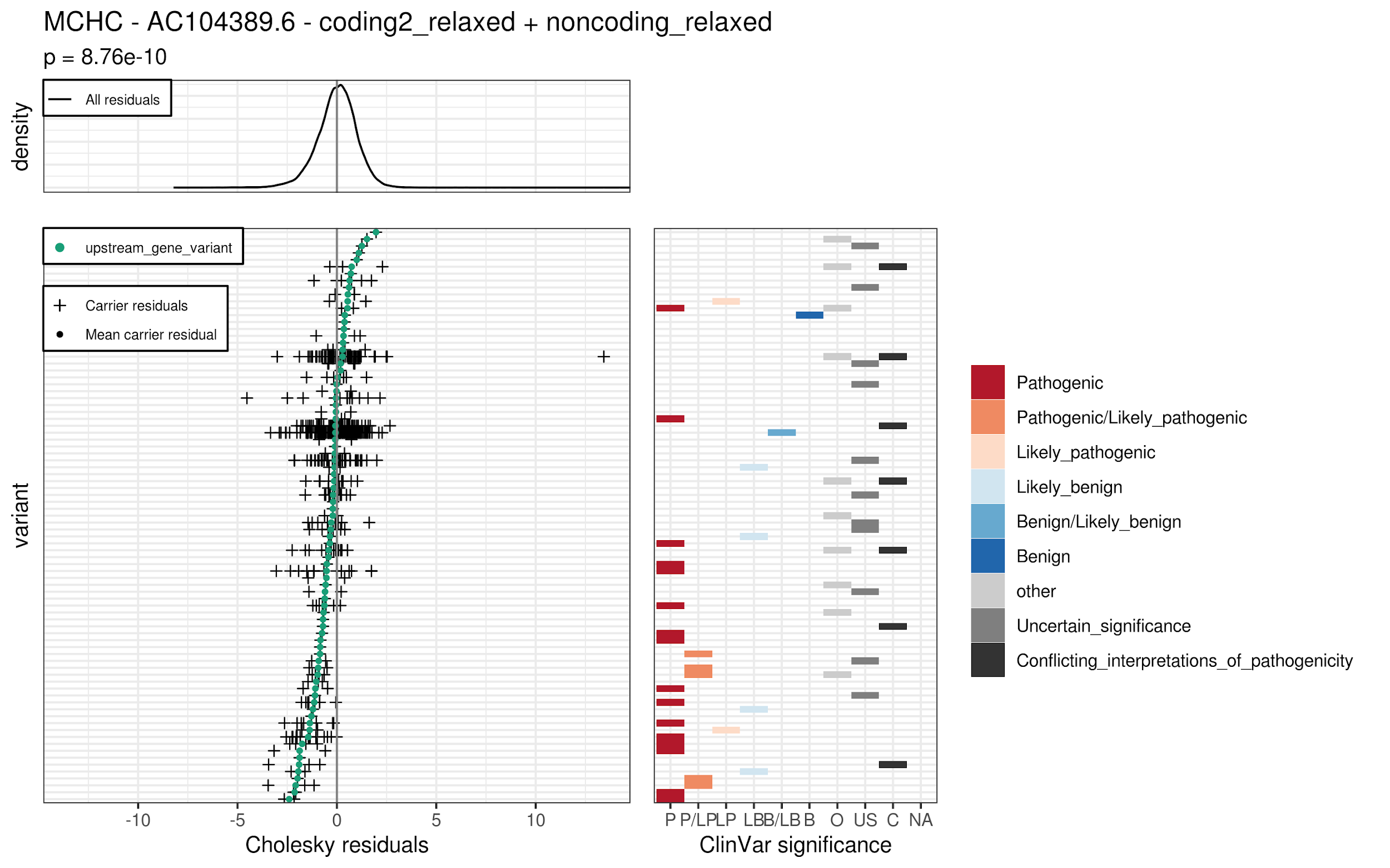
(D2)(D3)(D4)(D5)

(E)

(E1)

(E2)(E3)(E4)(E5)(E6)(E7)(E8)(E9)(E10)(E11)(E12)(E13)(E14)

(F)

(F1)

(F2)(F3)(F4)(F5)(F6)(F7)(F8)(F9)(G)

(G1)

(G2)(G3)(G4)(G5)(G6)(G7)(G8)(G9)(G10)(G11)(G12)(G13)(G14)
