## Supplemental Methods for "Whole genome sequencing association analysis of quantitative red blood cell phenotypes: the NHLBI TOPMed program"

**Participating studies**

*Amish*

The Amish Complex Disease Research Program includes a set of large community-based studies focused largely on cardiometabolic health carried out in the Old Order Amish (OOA) community of Lancaster, Pennsylvania (<http://medschool.umaryland.edu/endocrinology/amish/research-program.asp>). The OOA population of Lancaster County, PA immigrated to the Colonies from Western Europe in the early 1700’s. There are now over 30,000 OOA individuals in the Lancaster area, nearly all of whom can trace their ancestry back 12-14 generations to approximately 700 founders. Investigators at the University of Maryland School of Medicine have been studying the genetic determinants of cardiometabolic health in this population since 1993. To date, over 7,000 Amish adults have participated in one or more of our studies.

Due to their ancestral history, the OOA are enriched for rare exonic variants that arose in the population from a single founder (or small number of founders) and propagated through genetic drift. Many of these variants have large effect sizes and identifying them can lead to new biological insights about health and disease. The parent study for this WGS project provides one (of multiple) examples. In our parent study, we identified through a genome-wide association analysis a haplotype that was highly enriched in the OOA that is associated with very high LDL-cholesterol levels. At the present time, the identity of the causative SNP – and even the implicated gene – is not known because the associated haplotype contains numerous genes, none of which are obvious lipid candidate genes. A major goal of the WGS that will be obtained through the NHLBI TOPMed Consortium will be to identify functional variants that underlie some of the large effect associations observed in this unique population.

*ARIC*

The ARIC study is a population-based cohort study consisting of 15,792 men and women that were drawn from four U.S. communities (Suburban Minneapolis, Minnesota; Washington County, Maryland; Forsyth County, North Carolina, and Jackson, Mississippi) [^1^](http://sciwheel.com/work/citation?ids=5836966&pre=&suf=&sa=0). It was designed to investigate the causes of atherosclerosis and its clinical outcomes, and variation in cardiovascular risk factors, medical care, and disease by race, sex, location, and date. Participants were between age 45 and 64 years at their baseline examination in 1987-1989 when blood was drawn for DNA extraction and participants consented to genetic testing.

*BioMe*

The Charles Bronfman Institute for Personalized Medicine at Mount Sinai Medical Center (MSMC), BioMe Biobank, founded in September 2007, is an ongoing, broadly-consented electronic health record-linked clinical care biobank that enrolls participants non-selectively from the Mount Sinai Medical Center patient population. The MSMC serves diverse local communities of upper Manhattan, including Central Harlem (86% African American), East Harlem (88% Hispanic/Latino), and Upper East Side (88% Caucasian/White) with broad health disparities.

*CARDIA*

The Coronary Artery Risk Development in Young Adults (CARDIA) Study is a study examining the development and determinants of clinical and subclinical cardiovascular disease and their risk factors. It began in 1985-1986 with a group of 5,115 black and white men and women aged 18-30 years. The participants were selected so that there would be approximately the same number of people in subgroups of race, gender, education (high school or less and more than high school) and age (18-24 and 25-30) in each of 4 centers: Birmingham, AL; Chicago, IL; Minneapolis, MN; and Oakland, CA.

*CHS*

The Cardiovascular Health Study (CHS) is a population-based cohort study of risk factors for coronary heart disease and stroke in adults 65 years and older conducted across four field centers [^2^](http://sciwheel.com/work/citation?ids=3957807&pre=&suf=&sa=0). The original predominantly European ancestry cohort of 5,201 persons was recruited in 1989-1990 from random samples of people on Medicare eligibility lists from four US communities. Subsequently, an additional predominantly African-American cohort of 687 persons was enrolled for a total sample of 5,888. Institutional review committees at each field center approved the CHS, and participants gave informed consent. Blood samples were drawn from all participants at their baseline examination, and DNA was subsequently extracted from available samples. These analyses were limited to participants with available DNA who also consented to genetic studies. Participants were examined annually from enrollment to 1999 and continued to be under surveillance for stroke following 1999.

*COPDGene*

COPDGene (also known as the Genetic Epidemiology of COPD Study) is an NIH-funded, multicenter study. A study population of more than 10,000 smokers (1/3 African American and 2/3 non-Hispanic White) has been characterized with a study protocol including pulmonary function tests, chest CT scans, six minute walk testing, and multiple questionnaires. Five years after this initial visit, all available study participants are being brought back for a follow-up visit with a similar study protocol. This study has been used for epidemiologic and genetic studies. Previous genetic analysis in this study has been based on genome-wide SNP genotyping data. Approximately 1,900 subjects underwent whole genome sequencing in this NHLBI WGS project, including severe COPD subjects and resistant smoking controls. The COPDGene Study web site is: <http://www.copdgene.org/>.

*FHS*

FHS is a three-generation, single-site, community-based, ongoing cohort study that was initiated in 1948 to investigate prospectively the risk factors for CVD including stroke. It now comprises 3 generations of participants: the Original cohort followed since 1948 [^3^](http://sciwheel.com/work/citation?ids=9571293&pre=&suf=&sa=0); their Offspring and spouses of the Offspring, followed since 1971 [^4^](http://sciwheel.com/work/citation?ids=5959010&pre=&suf=&sa=0); and children from the largest Offspring families enrolled in 2002 (Gen 3) [^5^](http://sciwheel.com/work/citation?ids=5959012&pre=&suf=&sa=0). The Original cohort enrolled 5,209 men and women who comprised two-thirds of the adult population then residing in Framingham, MA. Survivors continue to receive biennial examinations. The Offspring cohort comprises 5,124 persons (including 3,514 biological offspring) who have been examined approximately once every 4 years. The Gen 3 cohort contains 4,095 participants.

*GeneSTAR*

In 1982 The Johns Hopkins Sibling and Family Heart Study was created to study patterns of coronary heart disease and related risk factors in families with early-onset coronary disease, identified from10 Baltimore area Hospitals. GeneSTAR continues to study mechanisms of coronary heart disease and stroke in families using novel models and exciting new methods. GeneSTAR is a family-based study in initially healthy brothers and sisters, and offspring of people with early-onset coronary disease, The goal is to discover and amplify mechanisms of stroke and coronary heart disease. Our African American and European American family cohort has undergone extensive screening, genetic testing, and follow-up for new cardiovascular disease, stroke, and other clinical events for 5 to 32 years.

*HCHS/SOL*

The Hispanic Community Health Study/Study of Latinos (HCHS/SOL) is a multi-center study of Hispanic/Latino populations with the goal of determining the role of acculturation in the prevalence and development of diseases, and to identify other traits that impact Hispanic/Latino health [^6^](http://sciwheel.com/work/citation?ids=3604289&pre=&suf=&sa=0) . The study is sponsored by the National Heart, Lung, and Blood Institute (NHLBI) and other institutes, centers, and offices of the National Institutes of Health (NIH). Recruitment began in 2006 with a target population of 16,000 persons of Cuban, Puerto Rican, Dominican, Mexican or Central/South American origin. Participants were recruited through four sites affiliated with San Diego State University, Northwestern University in Chicago, Albert Einstein College of Medicine in Bronx, New York, and the University of Miami. Recruitment was implemented through a two-stage area household probability design ^6^. The study enrolled 16,415 participants who were self-identified Hispanic/Latino and aged 18-74 years and the extensive psycho-social and clinical assessments were conducted during 2008-2011. Annual telephone follow-up interviews are ongoing since study inception. During the 2014-2017 second visit, the participants were re-examined again of various health outcomes of interest.

*JHS*

The Jackson Heart Study (JHS, <https://www.jacksonheartstudy.org/jhsinfo/>) is a large, community-based, observational study whose participants were recruited from urban and rural areas of the three counties (Hinds, Madison and Rankin) that make up the Jackson, MS metropolitan statistical area (MSA). Participants were enrolled from each of 4 recruitment pools: random, 17%; volunteer, 30%; currently enrolled in the Atherosclerosis Risk in Communities (ARIC) Study, 31% and secondary family members, 22%. Recruitment was limited to non-institutionalized adult African Americans 35-84 years old, except in a nested family cohort where those 21 to 34 years of age were also eligible. The final cohort of 5,301 participants included 6.59% of all African American Jackson MSA residents aged 35-84 during the baseline exam (N-76,426, US Census 2000). Among these, approximately 3,700 gave consent that allows genetic research and deposition of data into dbGaP. Major components of three clinic examinations (Exam 1 – 2000-2004; Exam 2 – 2005-2008; Exam 3 – 2009-2013) include medical history, physical examination, blood/urine analytes and interview questions on areas such as: physical activity; stress, coping and spirituality; racism and discrimination; socioeconomic position; and access to health care. Extensive clinical phenotyping includes anthropometrics, electrocardiography, carotid ultrasound, ankle-brachial blood pressure index, echocardiography, CT chest and abdomen for coronary and aortic calcification, liver fat, and subcutaneous and visceral fat measurement, and cardiac MRI. At 12-month intervals after the baseline clinic visit (Exam 1), participants have been contacted by telephone to: update information; confirm vital statistics; document interim medical events, hospitalizations, and functional status; and obtain additional sociocultural information. Questions about medical events, symptoms of cardiovascular disease and functional status are repeated annually. Ongoing cohort surveillance includes abstraction of medical records and death certificates for relevant International Classification of Diseases (ICD) codes and adjudication of nonfatal events and deaths. CMS data are currently being incorporated into the dataset.

*MESA*

The MESA study is a study of the characteristics of subclinical cardiovascular disease (disease detected non-invasively before it has produced clinical signs and symptoms) and the risk factors that predict progression to clinically overt cardiovascular disease or progression of the subclinical disease [^7^](http://sciwheel.com/work/citation?ids=2385970&pre=&suf=&sa=0). MESA researchers study a diverse, population-based sample of 6,814 asymptomatic men and women aged 45-84. Thirty-eight percent of the recruited participants are white, 28 percent African-American, 22 percent Hispanic, and 12 percent Asian, predominantly of Chinese descent. Participants were recruited from six field centers across the United States: Wake Forest University, Columbia University, Johns Hopkins University, University of Minnesota, Northwestern University and the University of California - Los Angeles.

*SAFS*

The San Antonio Family Study (SAFS) is a complex pedigree-based mixed longitudinal study designed to identify low frequency or rare variants influencing susceptibility to cardiovascular disease, using WGS information from 2,590 individuals in large Mexican American pedigrees from San Antonio, Texas. The major objectives of this study are to identify low frequency or rare variants in and around known common variant signals for CVD, as well as to find novel low frequency or rare variants influencing susceptibility to CVD.

*WHI*

The Women’s Health Initiative (WHI) is a long-term, prospective, multi-center cohort study that investigates post-menopausal women’s health [^8^](http://sciwheel.com/work/citation?ids=1209240&pre=&suf=&sa=0). WHI was funded by the National Institutes of Health and the National Heart, Lung, and Blood Institute to study strategies to prevent heart disease, breast cancer, colon cancer, and osteoporotic fractures in women 50-79 years of age. WHI involves 161,808 women recruited between 1993 and 1998 at 40 centers across the US. The study consists of two parts: the WHI Clinical Trial which was a randomized clinical trial of hormone therapy, dietary modification, and calcium/Vitamin D supplementation, and the WHI Observational Study, which focused on many of the inequities in women’s health research and provided practical information about the incidence, risk factors, and interventions related to heart disease, cancer, and osteoporotic fractures.

**Phenotype harmonization**

Because multiple studies contributed to the analysis, RBC phenotypes were harmonized across studies such that they could be analyzed together (https://www.biorxiv.org/content/10.1101/2020.06.18.146423v1). For most studies, variables were obtained from dbGaP. Data for the BioME study, the COPDGene study, and some participants in the WHI study were transferred directly to investigators. The study variables were QC’d to identify recording errors or other problematic data. After QC, variables were converted to a consistent measurement unit across studies. When possible, harmonized variables were calculated using study-derived variables. If QC uncovered data quality issues with the study-derived variable or if the study did not provide the specific variable of interest, the harmonized variable was instead calculated from the components (e.g., MCH = 10 * hemoglobin / red cell count). Finally, QC of the harmonized variable was performed to check that no large differences between studies remained.

Both FHS and ARIC measured phenotypes at multiple times in a given participant. For these studies, a single measurement for each participant was selected for each trait. To maximize the sample size and minimize batch effects within studies, harmonized data for different RBC traits for a single subject may have been measured at different visits. Within FHS, only the Offspring cohort had measurements at multiple exams. For these participants, the measurement at the most recent exam was chosen. For the ARIC study, the visit with the most non-missing phenotype values across all participants was chosen first. For subjects without measurements at this visit, the visit with the next most non-missing values was chosen, and so forth. As a consequence, values for the same participant for different RBC phenotypes were sometimes measured at different visits."

Trait-specific QC procedures were also performed. We excluded participants with HCT values >80% and those with HCT <5% (n=14). Similarly for HGB, we excluded participants with HGB measurements >30 g/dL and <5 g/dL (n=31). For participants from WHI, for both the HCT and HGB analyses, we excluded those with an HCT/HGB ratio >7 (n=11). Participants with MCH values >75 pg were excluded from analysis (n=2). In MCHC, we excluded participants with measurements $\geq$60 g/dL (n=1). For the MCV analysis, outliers with values >150 fL were excluded (n=2). In the RBC and RDW analyses, no participants were excluded based upon measured trait values.

**Statistical analyses**

*Genetic Ancestry and Relatedness*

Principal components (PCs) of genetic ancestry and pairwise relatedness measures were estimated for all 140,062 samples included in the TOPMed ‘freeze 8’ genotype release. Autosomal genetic variants passing the quality filter with a MAF > 0.01 and missing call rate < 0.01 were LD-pruned with an r^2^ threshold of 0.1 to obtain a set of 638,486 effectively independent variants for genetic ancestry and relatedness estimation. PC-AiR [^9^](http://sciwheel.com/work/citation?ids=3828309&pre=&suf=&sa=0) was used to obtain ancestry informative PCs robust to familial relatedness; the first 11 PCs showed evidence of population structure. PC-Relate [^10^](http://sciwheel.com/work/citation?ids=3828306&pre=&suf=&sa=0) was then used to estimate pairwise kinship coefficients (KCs) for all pairs of samples, conditional on the genetic ancestry captured by PC-AiR PCs 1-11; these KC estimates reflect only recent genetic relatedness, e.g. due to pedigree structure. The PC-Relate KC estimates were used to construct a 4th degree sparse, block-diagonal, empirical kinship matrix (KM) for association testing, using the procedure recommended in Gogarten et al [^11^](http://sciwheel.com/work/citation?ids=7863263&pre=&suf=&sa=0).: any pair of samples with estimated KC > 2^(-11/2)^ ~ 0.022 were clustered in the same block; all KC estimates within a block of samples were kept, regardless of value; and all KC estimates between blocks were set to 0. By using a sparse block-diagonal KM, the association tests are more computationally efficient yet recent genetic relatedness is still accounted for. We subset the freeze-wide PCs and sparse KM to the appropriate set of participants for each analysis.

*HARE for Imputation of Race/Population Membership using Genetic Ancestry*

Ancestry groups were based on a combination of participants reported race/ethnicity and genetic ancestry represented by PCs from PC-AiR [^9^](http://sciwheel.com/work/citation?ids=3828309&pre=&suf=&sa=0). To infer race/population group membership for participants with missing values, we used the HARE method [^12^](http://sciwheel.com/work/citation?ids=7633580&pre=&suf=&sa=0) . HARE is a machine learning algorithm that uses a support vector machine (SVM) to determine stratum assignment, taking as input genetically estimated PC values and reported race/ethnicity for each participant. Strata are defined by the unique reported race/ethnicity values provided, then the HARE SVM uses the input (training) data to learn the probability of stratum membership across the entire PC space. The output of HARE consists of multinomial probability vectors of stratum membership for each participant. HARE was run on a subset of samples included in the TOPMed freeze 8 genotype release; specifically, samples for participants from non-US populations (e.g. XXX) and the Amish participants (because they were very distinct in PC space) were excluded from the HARE analysis. HARE was run using the first 9 PC-AiR PCs generated on this subset of samples to represent genetic ancestry with the following reported race/population groups: Asian, Black, Central American, Cuban, Dominican, Mexican, Puerto Rican, South American, and White. The genetic data from the 31,918 participants with either unreported or non-specific (e.g. ‘Multiple’ or ‘Other’) race and population membership was included in the HARE analysis, but they were not used to train the SVM. These participants were assigned to a population stratum based on their highest HARE output probability of membership. All other participants remained in the population stratum corresponding to their reported race/population group. Amish participants were assigned to their own stratum.

*Conditional analyses*

We performed three types of conditional analysis in the discovery stage. The first conditional analyses adjusted each trait for variants that passed the QC filter and were previously reported to be associated with the particular RBC trait. These variants were pruned to a set with linkage disequilibrium (LD) r2 < 0.8 such that a variant with a more significant p-value were preferentially retained over those with lower p-values. Known variants failing the QC filter were included if any variants within 1 MB of the known variant remained significant after adjusting for the passing variants only. We refer to this analysis as the “RBC trait-specific conditional analysis”. In the second conditional analysis, we included all previously reported variants for *any* of the seven RBC traits as well as any failed variants included in any of the trait-specific conditional analyses. Variants were again pruned to LD < 0.8 with preferential selection based on p-value. We refer to the second conditional analysis as the “RBC trait-agnostic conditional analysis”. Finally, we performed iterative conditional analysis by chromosome for each trait. For this third conditional analysis, we started with the trait-specific conditional analysis, added in the most significant variant per chromosome, and re-ran association tests on that chromosome (“iterative conditional analysis”). This process was repeated until no variants remained significant.

*Aggregation Strategies*

For aggregate association testing, five distinct methods were used to aggregate rare variants into gene-based groups using GENCODE v29 gene model. Three strategies only included coding variants, and two strategies additionally included non-coding variants. Variants were further filtered using one or more deleterious prediction scores to enrich for likely causal variants. The detail method used for each strategy is provided below

1. **Coding filter 1 - Stringent (C1-S)**: This strategy includes high confidence predicted LoF variants inferred using LOFTEE (https://github.com/konradjk/loftee), missense variants predicted deleterious by all of SIFT4G[26633127]<=0.05, Polyphen2_HDIV>0.5[20354512], Polyphen2_HVAR>0.5[20354512], and variants predicted as “Deleterious” by LRT [19602639] and inframe indels or synonymous variants with Fathmm-XF score[28968714] > 0.5
2. **Coding filter 1 - Relaxed (C1-R)**: This strategy is same as C1-S but the missense filter was relaxed to retain variants predicted deleterious by any of SIFT4G, Polyphen2_HDIV, Polyphen2_HVAR scores
3. **Coding filter 2 - Relaxed (C2-R)**: This strategy is the same as C1-S but missense variants were filtered using MetaSVM score and a relatively relaxed set of missense variants was retained by applying MetaSVM score [25552646] > 0 filter
4. **Coding filter 2 - Relaxed & Non-coding filter-Relaxed (C2-R+NC-R)**: This strategy includes variants included in C2-R and additional regulatory variants. Regulatory variants were included if they overlapped with enhancer(s) or promoters linked to a gene using GeneHancer[28605766], or 5 Kb upstream of the Transcription start site. Within these regions only those variants were retained which had Fathmm-XF score > 0.5 **or** overlap with regions labelled as either“CTCF binding sites,” “Transcription factor binding sites” as annotated by the Ensembl regulatory build annotation[25887522]
5. **Coding filter 2 - Relaxed & Non-coding filter-Stringent (C2-R +NC-S)** :This strategy includes variants included in C2-R and additional regulatory variants using a stringent filtering criteria. Even in this method regulatory variants in Genehancer[28605766] linked regulatory regions and 5 Kb upstream of the Transcription start site of gene were included. However within these regions only those variants were retained which had Fathmm-XF score > 0.5 **and** which overlapped with regions labelled as “Promoters,” “Promoter flanking regions,” “Enhancers,” “CTCF binding sites,” “Transcription factor binding sites” or “Open chromatin regions” as annotated by the Ensembl regulatory build annotation[25887522].

The annotation based variant filtering and gene based aggregation was performed using TOPMed freeze 8 WGSA Google BigQuery annotation database on the BiodataCatalyst powered by Seven Bridges platform (http://doi.org/10.5281/zenodo.3822858). The annotation database was built using variant annotations generated by Whole genome Sequence annotator version v0.8 [26395054] and formatted by WGSAParsr version 6.3.8 (<https://github.com/UW-GAC/wgsaparsr>). The GENCODE v29 gene model based varint consequences were obtained from Ensembl Variant effect predictor (VEP)[26683364] incorporated within WGSA. When using a deleteriousness prediction score, respective author recommended cut points were used to retain likely deleterious variants.
